# Risk factors and preventive interventions for post Covid-19 condition: systematic reviews

**DOI:** 10.1101/2022.03.25.22272949

**Authors:** Jennifer Pillay, Sholeh Rahman, Samantha Guitard, Aireen Wingert, Lisa Hartling

**Affiliations:** Alberta Research Centre for Health Evidence (ARCHE), Department of Pediatrics, University of Alberta Faculty of Medicine and Dentistry, Edmonton, Alberta, Canada

## Abstract

**Background:** The Covid-19 outbreak has presented many challenges to governments and healthcare systems, including observations of symptoms that persist beyond acute infection labelled as post Covid-19 condition.

**Objectives:** To systematically identify and synthesize evidence around pre-existing and clinical risk factors for post Covid-19 condition (occurring ≥12 weeks after positive test/symptom onset) (KQ1), and interventions during the acute and post-acute phases of the illness that could potentially prevent post Covid-19 condition (KQ2).

**Methods:** We searched Medline and Embase (Jan 2021-Aug 12 2021 [KQ1], and Jan 2020-Jul 28, 2021 [KQ2]), Clinicaltrials.gov, organizational websites, and reference lists of included studies and relevant systematic reviews. Two investigators independently reviewed abstracts and full-text articles against a priori inclusion criteria, and disagreements were resolved through discussion or by consulting a third reviewer. One investigator abstracted data and assessed risk of bias using design-specific criteria, and a second investigator checked data abstraction and assessments for completeness and accuracy. Meta-analysis was performed when there was sufficient clinical and methodological similarity in an exposure-outcome comparison, based on prespecified variables. We assessed the certainty of evidence using the Grading of Recommendations, Assessment, Development and Evaluation approach (GRADE). A relative effect/association of 0.75-1.49 was considered as “little-to-no”, whereas 0.50-0.74/1.5-1.99 was “small-to-moderate” and <0.50/ ≥2.00 was “large” for fewer/benefit or more/harm, respectively

**Results:** From 4,672 (KQ1) and 3,781 (KQ2) citations we included 17 and 18 studies, though 4 studies were included for both KQs. We found small-to-moderate associations between female sex and higher non-recovery, fatigue, and dyspnea (moderate certainty). Severe or critical acute-phase Covid-19 severity (versus not) has probably (moderate certainty) a large association with increased cognitive impairment, a small-to-moderate association with more non-recovery, and a little-to-no association with dyspnea. There may be (low certainty) large associations between hospitalization during the acute illness and increased non-recovery, increased dyspnea, and reduced return to work. There may be small-to-moderate associations between several other risk factors and post Covid-19 condition outcomes, including age ≥60 versus <60 (functional incapacity), non-White people (lower return to work), children age >6 versus <2 years (non-recovery), having ≥1 versus no comorbidities (non-recovery), chronic pulmonary disease (fatigue), rheumatologic disorder (depression/anxiety), and chronic obstructive pulmonary disease or hypertension (cognitive impairment). Several other risk factors had low certainty for little-to-no association with one or more outcomes (e.g. diabetes, cardiovascular disease) or very low certainty. Interventions to prevent post Covid-19 condition included medications (standard and traditional/ayurvedic), stem cell therapy, rehabilitation or similar therapies, and screening/referrals at either acute phase (symptom onset to 4 weeks) or early post-acute phase (4-8 week), with short (12-16 weeks) or longer (>16 weeks) follow-up for outcomes. We are very uncertain about the effects of preventive interventions, mainly due to risk of bias, inconsistency/lack of consistency (single studies), and in some cases imprecision.

**Conclusions:** Guidelines in relation to surveillance, screening services, and other services such as access to sickness and disability benefits, might need to focus on females and those with previously severe Covid-19 illness. Interventions targeting fatigue, dyspnea, and cognitive impairment (especially in those who had severe Covid-19) may be good to prioritize for development and evaluation to provide evidence on their effects. Inputs from patients and primary care providers should be taken into account when developing new care pathways and some tailoring to individual needs will likely be paramount. Continuous assessment of the rapidly emerging evidence is important to better shape our understanding as the body of evidence grows. Sufficiently powered prospective trials of preventive interventions are warranted.

**PROSPERO registration:** CRD42021270354

## BACKGROUND

The ongoing global pandemic caused by coronavirus disease 2019 (Covid-19) has affected nearly 270 million people so far and has resulted in more than 5 million deaths worldwide (1). While most people infected with Covid-19 typically recover within a few weeks (2), some may experience persistent symptoms lasting for several weeks or even months after the initial infection (3, 4). Several terminologies and definitions have been proposed to describe prolonged Covid-19 illness (5-8). The term “post Covid-19 condition” was established by the WHO as of October 6, 2021, to refer to new or ongoing symptoms in individuals with a history of probable or confirmed SARS-CoV-2 infection, occurring usually 3 months from the onset of the infection, lasting for at least 2 months following initial recovery that cannot be explained by an alternative diagnosis (9). The exact frequency and nature of post Covid-19 condition remains largely unknown, partly due to the lack of a consistent definition and ascertainment criteria prior to October 2021 (10). Emerging evidence indicate varying estimates, ranging from 10% to more than 80% of infected individuals suffering from on-going symptoms for weeks or months after the initial infection, with most commonly reported symptoms being fatigue, weakness, and breathlessness among others (11-14).

Our understanding of factors predisposing to post Covid-19 condition is limited (10, 13). Evidence indicates that occurrence and intensity of post-Covid symptoms may be influenced by several factors, including age, gender, pre-existing conditions, and the level of care received during initial stages of the disease (4, 15-19). There are also uncertainties around whether the type of the treatment received during the acute phase of Covid-19 influences longer outcomes (20).

Many local governments and healthcare systems have already been pushed beyond their limits to cope with the rapid spread of acute Covid-19 infection and its serious implications on healthcare utilization and management of resources. Given the timeline of the pandemic and large number of people infected with Covid-19, it is anticipated that post Covid-19 condition will become the new public health challenge to tackle (10). There is a clear need for better understanding this emerging threat and indeed, it has been urged to prioritize research in this area (20-22). Studying factors that predispose an individual to post Covid-19 condition will help to identify high-risk groups, target potential interventions to those groups, and establish effective patient-care pathways. This, in turn, would ensure evidence-based allocation of resources and a better preparedness of health systems to overcome the challenge. Hence, the objective of these systematic reviews was to identify and synthesize evidence around risk factors of post Covid-19 condition and interventions provided to patients during the acute and post-acute phases of the disease that could potentially prevent post Covid-19 condition.

## METHODS

### Review approach and key questions (KQs)

We undertook two systematic reviews following a pre-defined, registered protocol (CRD42021270354)(23). The reviews are reported following Preferred Reporting Items for Systematic Reviews and Meta-analyses (24, 25). During protocol development, a working group comprised of members of our research team, representatives from the Public Health Agency of Canada (PHAC) and the Canadian Agency for Drugs and Technologies in Health (CADTH), and clinical experts was formed to refine the review questions and PICOTS components (population, intervention(s) or exposure(s), comparator(s), outcome(s), timing, setting, and study design). The following KQs were determined to be addressed:

#### KQ1

Among people who have had Covid-19, what are the associations between pre-existing and clinical risk factors and development of post Covid-19 condition?

#### KQ2

Among people in the acute (symptom onset to 4 weeks) or early post-acute phase (4-8 weeks) of Covid-19 what are the effects of interventions to prevent post Covid-19 condition?

### Eligibility criteria

**Tables S1** and **S2** in the Supplement details our eligibility criteria for each KQ. For the purpose of this review, we defined post Covid-19 condition as symptoms persisting ≥12 weeks after a positive Covid-19 test or symptom onset.

The population of interest for KQ1 included people of any age in the general population (with/without previous Covid-19) or those with Covid-19. For KQ2, we included people of any age in the acute (0-4 weeks since a positive test/symptom onset) or the post-acute (4-8 weeks) phase of Covid-19; studies could have ≤20% of participants at 9-12 weeks post-Covid. After the protocol development but before data extraction, it was decided by the working group to exclude studies where a majority of the participants had been admitted to an intensive care unit (ICU) during the acute phase, because of the presumed similarity and overlap with post-ICU syndrome for which there are existing treatment pathways and guidelines.

For KQ1, we were interested in pre-existing risk factors (e.g., demographic variables, BMI, specific chronic conditions affecting a relatively large population and found to have an effect on Covid-19 severity (26), Covid-19 vaccination status, etc.) and clinical risk factors arising during acute phase of Covid-19 (e.g., presence of dyspnea, number of symptoms/symptom severity, need for hospitalization or ICU admission, etc.)(27). We defined the comparator/control as people without the exposure of interest (e.g., male versus female, people without diabetes, etc.) or with different levels of the exposure (e.g., different age group or BMI category). For KQ2, we included any potentially preventive intervention that started before 8 weeks after a positive Covid-19 test/symptom onset (in ≥80% of study participants). The comparator was usual medical care (e.g., supportive care for acute Covid-19); we included studies with no comparator if there were no other studies with a comparator for the intervention.

The outcomes of interest were selected by a rating approach (28). Groups of clinical experts, policy makers, and patients were asked to rate the importance of each proposed outcome in terms of how important the effect of a risk factor or intervention on the outcome would be for patients with long-term symptoms, and for the government to offer new types of healthcare services to improve the outcome. The rating was on a 9-point scale, with scores 0-3 indicating “not very important”, 4-6 indicating “important but not critical”, and 7-9 indicating “critical” outcomes. Based on this rating, we included outcomes rated as “critical” for both KQs, which included: non-recovery/persistent symptoms; major cardiovascular event or organ impairment; moderate/severe or persistent (≥3 weeks) fatigue, breathlessness/dyspnea, impairment in functional capacity, cognitive impairment, sleep disturbances, pain including headaches and chest pain; important impact on quality of life; clinical/pathological levels of psychopathology (e.g., anxiety, depression, post-traumatic stress disorder); and unable to return to full-time work/school/education or caring role. For KQ2, due to the expectation of scarce evidence we also included outcomes rated as important, including: all-cause hospital admission, emergency department visits, requiring urgent care outside of hospital, requiring referral for specialist care for physical and/or mental health (may include new onset of disease such as diabetes), requiring pulmonary rehabilitation and/or long-term oxygen therapy, and any or serious adverse effects of the intervention/treatment. For KQ1, we included outcomes assessed at least 12 weeks after Covid-19 diagnosis or symptom onset (including studies where mean follow-up duration ± 1 standard deviation was ≥12 weeks). For KQ2, the post-baseline follow-up had to be at least 3 weeks and outcomes measured ≥12 weeks’ post-Covid. If outcomes were assessed during follow-up from an intervention and included <12 week data (e.g., hospitalization during follow-up), we looked for data specific to event timing (e.g., in figures or text) or contacted authors for data on events occurring ≥12 weeks post-Covid.

We included peer-reviewed articles, results in trial registrations (if no report published yet), and pre-prints of primary studies, including (for KQ1) prospective/retrospective observational studies with ≥ 300 participants with Covid-19, and (for KQ2) randomized and quasi-randomized or experimental studies (e.g., controlled before-after, interrupted time series, uncontrolled before-after implementation studies), prospective or retrospective cohort studies with control groups, case-control studies, and case series/uncontrolled cohorts. For uncontrolled studies reporting continuous outcomes, baseline and follow-up scores needed to be reported.

For all non-randomized studies, we prioritized multivariable adjusted data (or other similar adjustment methods, i.e., matching/stratification) where at least age, sex (when applicable), some measure of Covid-19 illness severity (e.g., hospitalization, ICU admission, etc.), and comorbidities were taken into account. For KQ1, we included studies with adjustment for at least two of the four variables, whereas for KQ2 we included all studies regardless of adjustment.

### Literature search and study selection

The search strategies for each KQ (Supplement) were developed by a research librarian and peer-reviewed by a second librarian using the PRESS 2015 checklist (29). Concepts related to post Covid-19 were combined with concepts related to risk factors (KQ1) and interventions (KQ2). Search vocabulary and syntax were adjusted across databases. In order to facilitate search updates, the searches were conducted in Ovid using a multifile search for Medline® including Epub Ahead of Print, In-Process & Other Non-Indexed Citations and Embase. We limited the search to studies in English or French from any country/setting, published from January 2021 (KQ1) and June 2020 (KQ2) onwards. The date limit was applied given the timeline of Covid-19 emergence and our focus on long-term outcomes, and (for KQ1) because we had access to other reviews from which to locate studies published earlier (12, 13, 30).

Moreover, our earlier review on risk factors associated with Covid-19 severity (including long-term outcomes) with literature search updated in April 2021, did not identify any studies prior to Fall 2020 (26). These searches were run on August 12, 2021 (KQ1) and July 28, 2021 (KQ2). Additionally, we searched Clinicaltrials.gov and several organizational websites based on previous input on those most relevant to Canada, including Government of Canada’s First Nations and Inuit Health Branch, PHAC, United States Centers for Disease Control and Prevention, Public Health England, European Centre for Disease Prevention and Control, CADTH Covid-19 Evidence Portal, and the reference lists of the included studies and relevant systematic reviews.

Search results were uploaded to an EndNote library (v. X9, Clarivate Analytics, Philadelphia, PA), duplicates were removed, and then were exported to DistillerSR (https://www.evidencepartners.com/). Following the pre-defined eligibility criteria, a two-step study selection was done in DistillerSR, first by title-abstract (screening) and then by full-text (selection). Before each step, all reviewers involved in study selection piloted a random sample of records to resolve any ambiguousness. During screening, we applied the liberal accelerated method whereby each title-abstract requires one reviewer to include but two to exclude. Any potentially relevant record was retained for full-text review. Study selection was done in duplicate (i.e., two independent reviewers) with arbitration by a third reviewer in case of a disagreement. If additional information was required to make a final decision on a study, we contacted the corresponding author twice via e-mail over two weeks. We excluded the study if there was no response after the two attempts. We documented the screening process in a PRISMA flow-diagram and recorded the reasons for all full-text exclusions.

## Data extraction and management

We developed standardized data extraction forms in Excel. After piloting the form, one reviewer extracted data from the included studies independently and a second reviewer verified all data for accuracy and completeness. Disagreements were resolved by discussion or by consulting a third reviewer.

From each study, we extracted information related to: study characteristics, population characteristics, setting and type of care during acute phase, risk factors (KQ1) or intervention characteristics (KQ2) and comparators, follow-up length, analysis details, outcome definitions and ascertainment, and results data. For all outcomes we prioritized including dichotomous (e.g., proportion with event or important degree of dysfunction) over continuous (e.g., mean score) data, and data attributed to or changed since Covid-19 (e.g., change in outcome pre versus post-Covid). We also prioritized outcomes assessed using a valid measurement tool/scale that best represented the outcome domain (23) and did not include those based on single question about experiencing a symptom unless it indicated the symptoms were persistent/moderate/severe. In cases of missing or unclear information, we contacted the study authors for clarification, and ceased contact after two attempts if we received no response. In case of multiple analyses reported for an outcome in observational studies, we extracted the most adjusted data. If data for outcomes at multiple time points were reported we selected the data closest to 12 and 24 months, with a preference for longer versus shorter time. We converted continuous to binary data where possible, to enable a pooled estimate across several studies (23).

### Risk of bias assessment

We assessed risk of bias of included studies using the JBI critical appraisal checklist for cohort studies (31) (KQ1), Cochrane RoB 2.0 for randomized studies (32) and JBI critical appraisal checklist (quasi-experimental) for quasi-randomized and other experimental studies, such as controlled before-after studies (31) (KQ2). For all study designs, we added a question about Covid-19 ascertainment (domain considered as low risk if ≥90% were lab-confirmed). For KQ1, we collected information about missing outcome data (i.e., measured as per methods but no results reported), and for KQ2 observational studies, we added three additional questions about selective reporting, missing outcome data, and whether a sufficient amount of eligible patients (>50%) were enrolled. The overall risk of bias for an outcome was considered “low” if all domains were at low risk of bias, of “some concern” if fewer than two domains were at high risk of bias and we did not feel the study conclusions would be impacted by that domain (e.g., ascertainment of exposure), and “high” if ≥2 domains were assessed as being high risk.

After piloting each tool on a sample of studies, one reviewer assessed the quality of each study independently, followed by verification by a second reviewer. Disagreements were resolved by discussion or arbitration by a third reviewer, if needed. To inform our assessment, we used information from all publications associated with any study as well as trial registrations and protocols for KQ2 studies. We did not exclude studies due to high risk of bias, however, risk of bias was considered when assessing the certainty of evidence using the Grading of Recommendations, Assessment, Development and Evaluation approach (GRADE).

### Data synthesis

For both KQs, we charted the study data in terms of the main variables of intervention/exposure, Covid-19 illness severity (hospitalized, mixed, non-hospitalized), timing of outcome measurement, and outcome. As pre-specified, we considered a meta-analysis if there was enough clinical and methodological similarity in an exposure-outcome comparison and data for that comparison was available in at least 75% of the studies. For the risk factors related to acute-phase illness severity (e.g., critical/severe illness vs. not), with the exception of need for hospitalization which was only analyzed in mixed populations, we analyzed data separately for hospitalized, mixed severity and non-hospitalized populations to avoid confounding. Although acute Covid-19 severity was defined using different standards across the studies, we pooled estimates based on similarities in description of each level of severity (e.g., requiring mechanical/non-mechanical ventilation, high flow oxygenation, etc.). Analysis was performed in Review Manager (RevMan, v.5.3, Copenhagen: The Nordic Cochrane Centre, the Cochrane Collaboration, 2014). For KQ1 and studies with a control group in KQ2, we planned to use a pairwise random-effects meta-analysis. We employed a generic inverse variance method for KQ1 because of the preference for using relative measures from adjusted analysis. For all syntheses whether or not from meta-analysis, we categorized findings by magnitude: a relative effect/association of 0.75 to 1.49 was considered as “little-to-no”, whereas 0.50 to 0.74 and 1.5 to 1.99 were “small-to-moderate” and <0.50 or ≥2.00 were “large” effects/associations for fewer/benefit or more/harm, respectively. In absence of a meta-analysis, consensus was made on a best estimate (within one of these categories) of effect/association across studies while considering their relative weight by sample size. As pre-defined, we considered several variables for either grouping studies or conducting subgroup analyses if substantial heterogeneity existed in magnitude (e.g., different categories of conclusions) or direction of association/effects, including hospitalized versus non-hospitalized/mixed population, lab-confirmed Covid-19 versus otherwise, timing of outcome assessment (e.g., 12-22 weeks versus ≥22 weeks), symptom severity for both KQs as well as components of the intervention for KQ2 (e.g., enrollment in acute versus post-acute phase, online versus in-person intervention, follow-up timing). We also conducted sensitivity analysis by removing studies having high risk of bias, particularly those in KQ1 that only sufficiently reported univariate analyses despite conducting multivariate analysis. We would have tested for small study effects using funnel plots and Egger’s regression test if an analysis included at least 10 studies.

### Certainty of evidence

The certainty of evidence for each exposure-outcome association across the studies was assessed by at least two reviewers using GRADE (33, 34). We assessed our certainty in the categorical conclusions about the magnitude of association/effect as described above, though if an association had higher certainty at a smaller magnitude (small-to-moderate vs. large association) we chose to report the higher certainty magnitude. For observational studies, we started at high certainty for KQ1 (35) and low certainty for KQ2 (28). For randomized and non-randomized trials in KQ2, we started at high certainty. We rated down the certainty for concerns related to risk of bias, indirectness (mainly in terms of whether reported outcome measures were a good conceptual match to our outcomes of interest), inconsistency (in direction and/or magnitude of effect) across the studies or lack of consistency (single studies), imprecision (95% confidence intervals indicating the effect/association may allow for more than one conclusion e.g. little-to-no and a small-to-moderate association), and/or reporting bias domains by one or two levels depending on how serious the concerns were, that is how much overall conclusions appear to be impacted by the domain. The final certainty of evidence (i.e., high, moderate, low, very low) and the reasoning for each are presented in summary tables.

## RESULTS

### KQ1: Risk factors

We identified 4,612 records from searching databases and 150 records from other sources; 17 unique records met the eligibility criteria and were included (**Figure 1;** see **Supplement** for the lists of excluded studies for KQs 1 and 2) (4, 14-19, 36-45). Study characteristics are included in **Table 1** and **Table S3**. The studies originated from China (n=5), Italy (n=2), Norway (n=2), Russia (n=2), Switzerland (n=2), and one each from the UK, USA, Sweden, and Turkey. Fourteen studies were classified as prospective and three as retrospective cohort studies. The studies included a median of 540 participants (range: 304-11,955), with lab-confirmed Covid-19 in majority of studies (n=13), and varying baseline Covid-19 severity (hospitalized n=9, non-hospitalized n=3, mixed severity n=5). Of the included studies, only one was exclusively in children (40). The median age in other studies was 53.0 years (range: 42.7-69.0) and the median male proportion across the studies was 49%. Most of the studies (n=12) were assessed as having some concern for risk of bias, mainly due to issues arising from incomplete follow-up data, insufficient statistical adjustment (e.g., results not adjusted for comorbidities or illness severity), or outcome/exposure assessment method (i.e., self-reported outcomes or not measured with valid tools) (**Table S4**). Only one study (37) was assessed as having low risk of bias, which included elderly people previously hospitalized with Covid-19.

**Figure 1.**
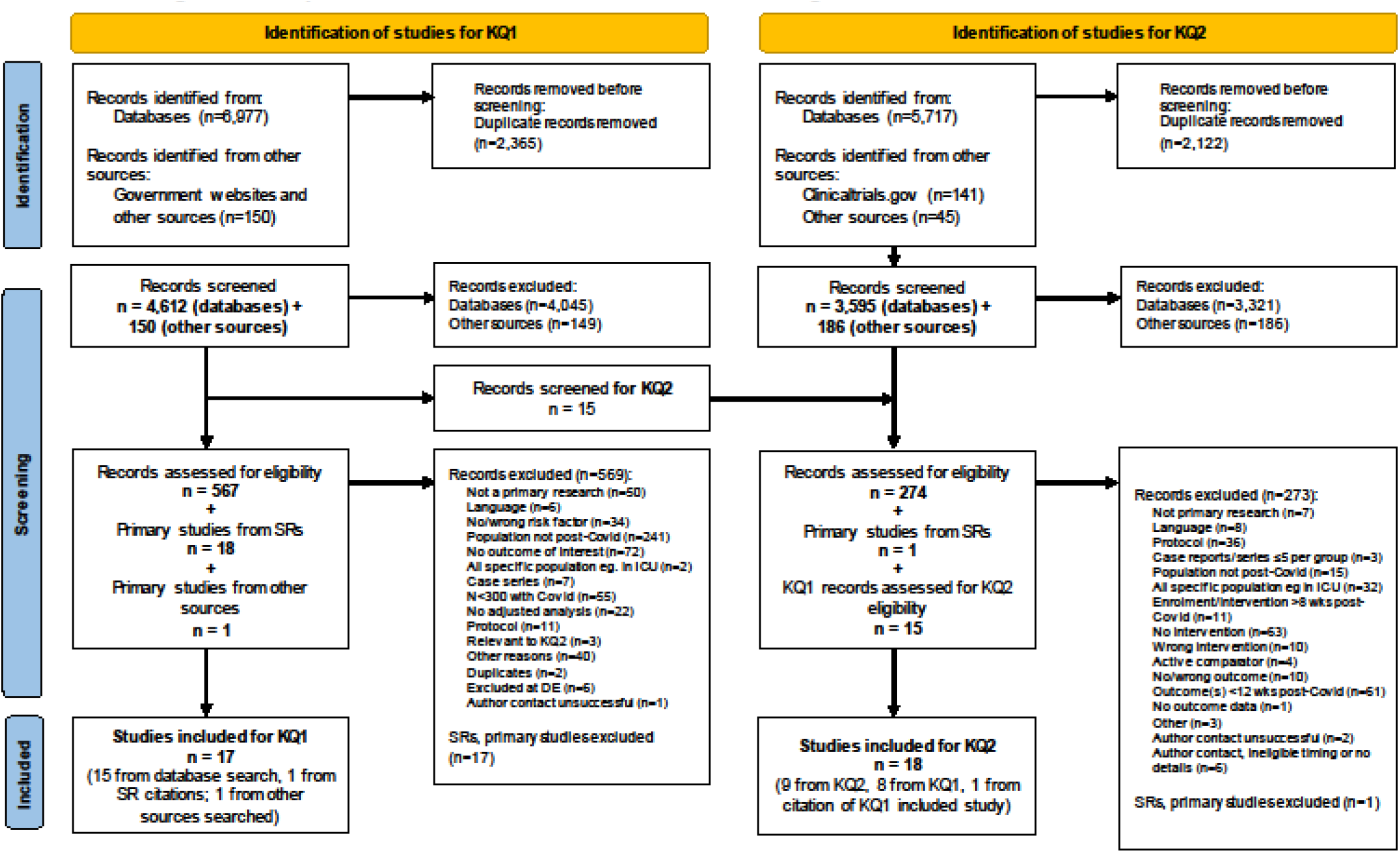
Flow of Literature.

**Table 1:**
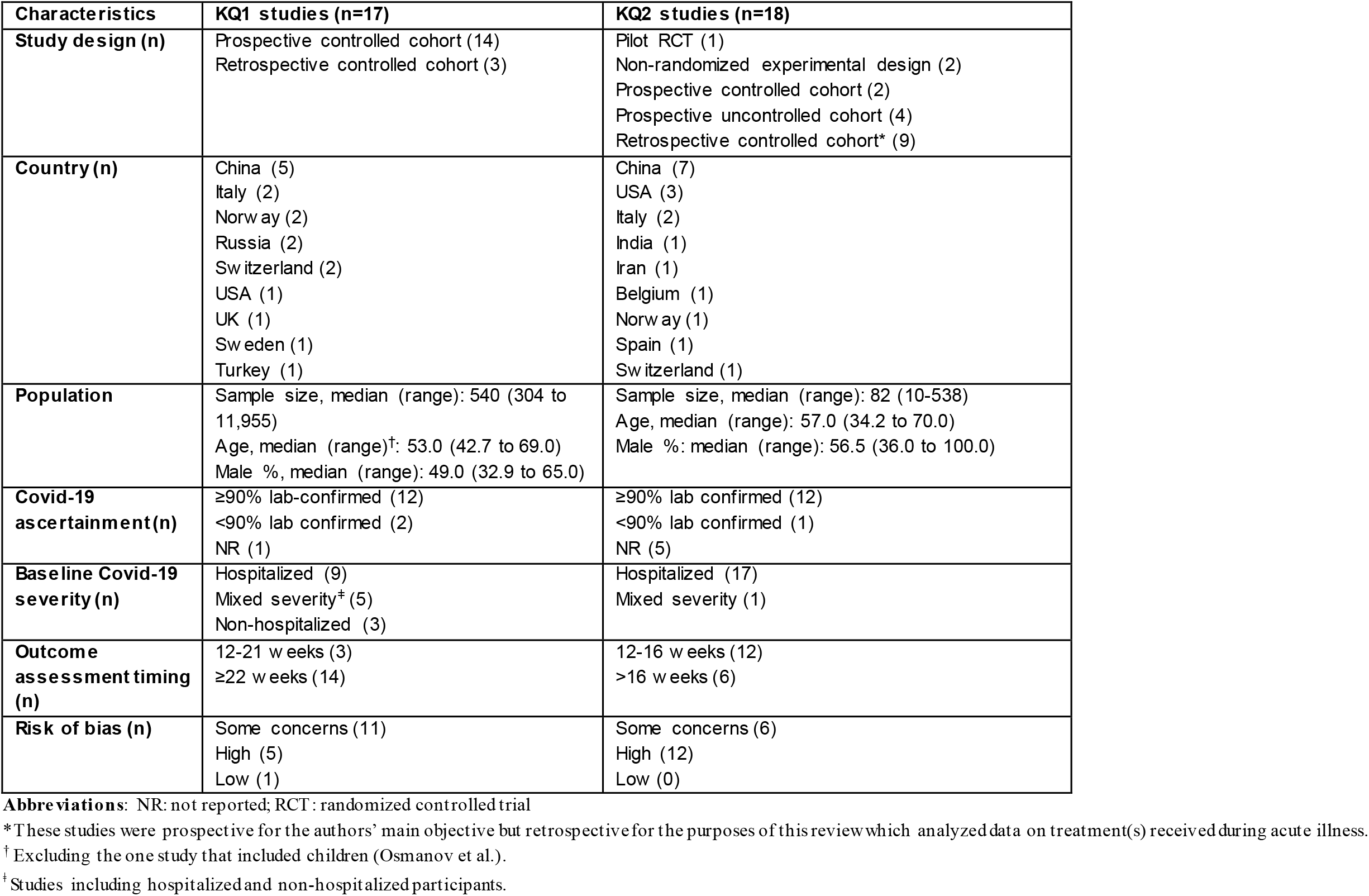
Summary of characteristics of studies included in key questions 1 and 2.

The certainty of evidence was downgraded for all risk factor-outcome associations, mostly due to concerns related to risk of bias, inconsistency (i.e., single study or inconsistent findings across studies), and/or indirectness (e.g., reported outcomes not aligning with review question). **Table 2** summarizes the findings where the evidence was of low or greater certainty; **Tables S5-S8** contain more detail including individual study and meta-analysis results, including risk factors for which there was very low certainty.

**Table 2:**
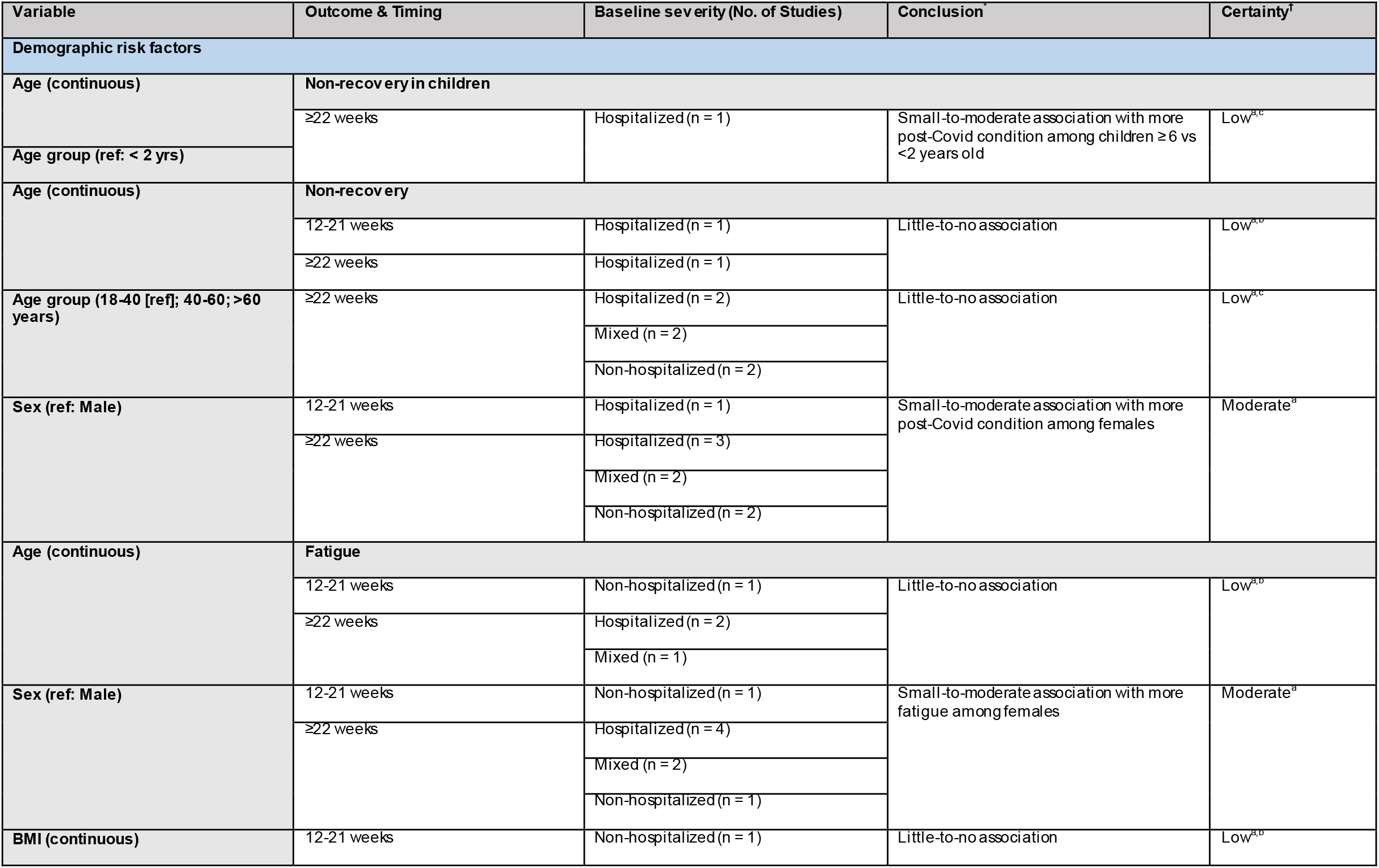
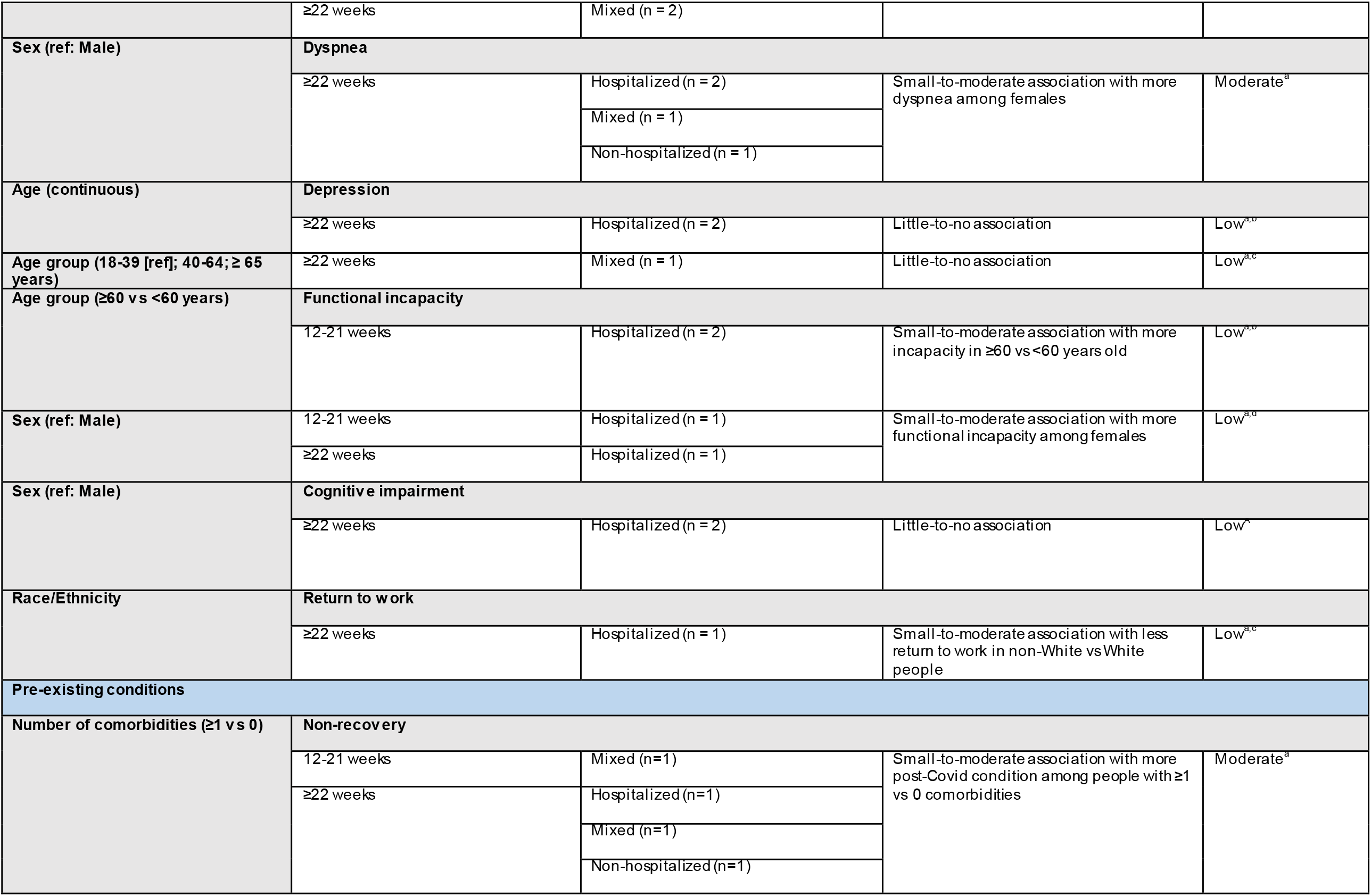
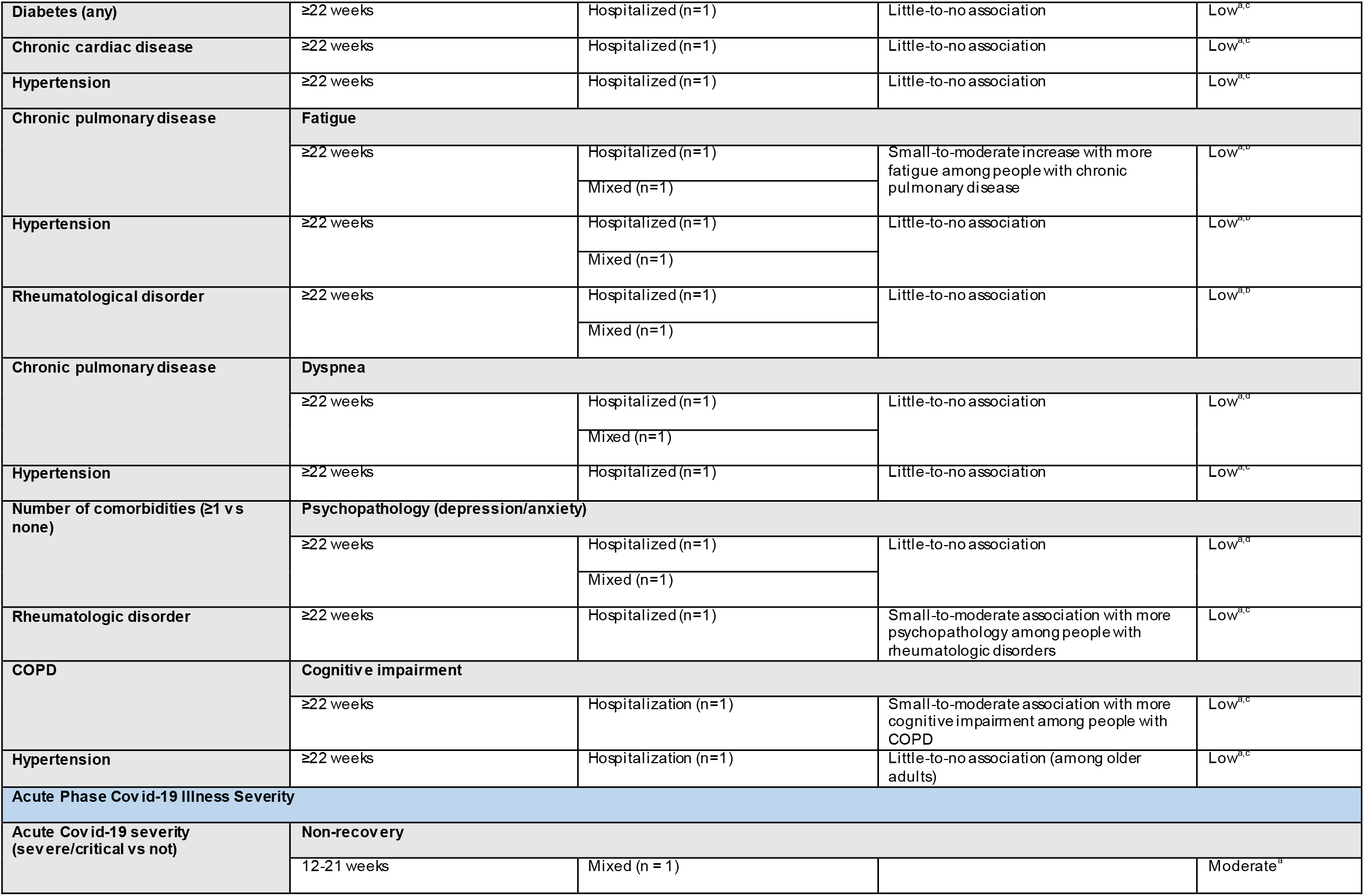
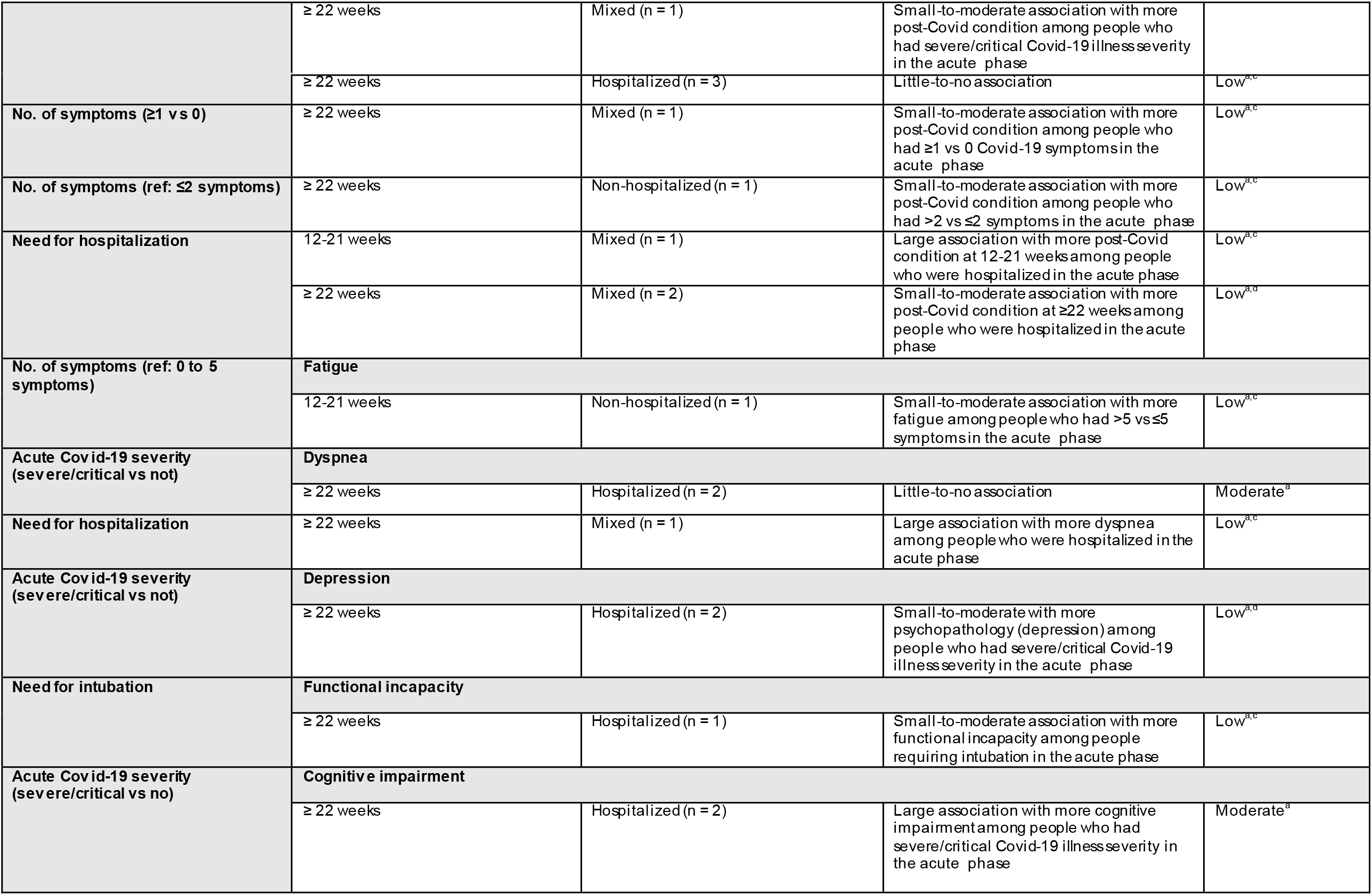
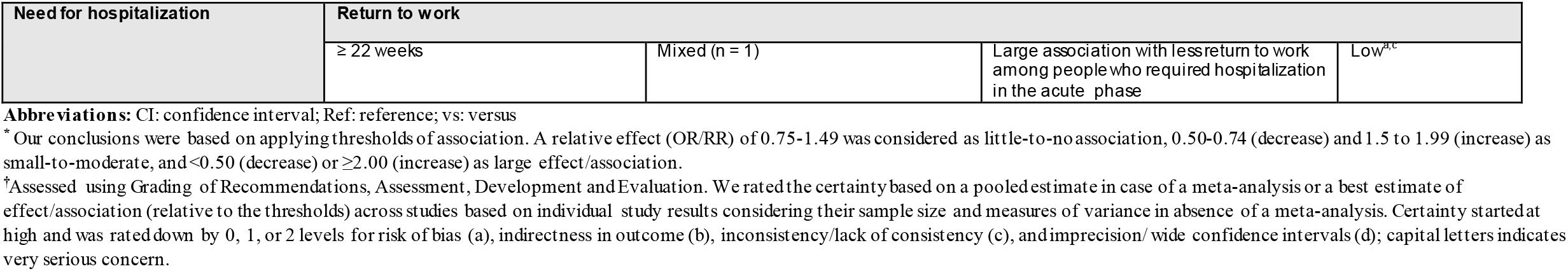
Summary of evidence for associations between risk factors and post Covid-19 condition with moderate or low level of certainty.

#### Demographic risk factors

The certainty of evidence was moderate for a small-to-moderate association between female sex and higher: non-recovery (8 studies, n=6,613) (15-18, 38, 39, 41, 43), fatigue (8 studies, n=7,116) (14, 15, 17, 36, 38, 39, 43, 44), and dyspnea (4 studies, n=3,817) (15, 17, 38, 39). The certainty of evidence was low for a small-to-moderate association with higher: non-recovery among children aged ≥6 versus <2 years (1 study, n=518) (40), functional incapacity in hospitalized adults aged ≥60 versus <60 years old (2 studies, n=867) (15, 42), functional incapacity in females (2 studies, n=867) (15, 42), and lower return to work (at a median of 6.7 months from symptom onset) in non-White people (1 study, n=382) (19). Several findings had low certainty for little-to-no association, and several risk factors had very low certainty evidence (**Table S5**).

#### Pre-existing conditions

The certainty of evidence was low for a small-to-moderate association between: number of comorbidities (i.e., ≥1 versus 0) and non-recovery (4 studies, n=2,069) (4, 15-17), chronic pulmonary disease with fatigue (2 studies, n= 2,961) (36, 38), rheumatologic disorder with depression/anxiety (1 study, n=2,649) (38), and chronic obstructive pulmonary disease (COPD) or hypertension with cognitive impairment (1 study, n=1,539) (37). Post Covid-19 condition was found to have little-to-no association (low certainty) with a few other pre-existing conditions, including diabetes and cardiovascular diseases, and there was very low certainty for several other pre-existing conditions (**Table S6**).

#### Acute-phase Covid-19 illness severity

Severe or critical acute Covid-19 illness severity (versus not) showed a moderate certainty of evidence for a large association with cognitive impairment (hospitalized populations; 2 studies, n=2,335) (37, 43), small-to-moderate association with non-recovery (mixed-severity populations; 2 studies, n=1,438) (4, 17), and little-to-no association with dyspnea (hospitalized populations; 2 studies, n=2,976) (15, 38). There was a low certainty of evidence for a large association between hospitalization during the acute phase with: non-recovery (1 study, n=1,007) (4), more dyspnea (1 study, n=431) (17), and reduced return to work (1 study, n=11,955) (45).

The certainty of evidence was low for a small-to-moderate association between ≥1 versus 0 acute Covid-19 symptoms (1 study, n=599)(18) and ≥3 versus ≤2 symptoms (1 study, n=304) (16) with non-recovery. A small-to-moderate association with low certainty was also found between: number of acute Covid-19 symptoms and fatigue (1 study, n=458) (44), severe or critical acute Covid-19 severity (versus not) and depression (2 studies, n=4,382) (14, 38), and need for intubation and functional incapacity (1 study, n=382) (19). **Table S7** includes detailed findings by study and synthesis.

### KQ2: Preventive interventions

We identified 3,595 records from searching databases and 186 records from other sources; 18 unique records, including 8 studies identified through the KQ1 search, met the eligibility criteria and were included (**Figure 1; Tables 1** and **S9**) (14, 19, 36, 41, 46-59). Most of the studies originated from China (n=7); others were from USA (n=3), Italy (n=2), and one each from India, Iran, Belgium, Norway, Spain and Switzerland. Nine studies were classified as retrospective controlled cohorts; others were prospective (controlled/uncontrolled) cohorts (n=6), non-randomized experimental (n=2) and a pilot randomized controlled trial (56). The studies included a median of 82 participants (range: 10-538) with 100% lab-conformed Covid-19 in majority of studies (n=14). Only one study involved hospitalized and home-isolated participants (36), while the other 17 studies were among hospitalized individuals with mixed severity (ranging from being asymptomatic to severe/critical). We assessed the risk of bias for objective (i.e., measured) and subjective (i.e., self-reported) outcomes separately; none of the studies were rated as having low risk of bias across all outcomes (**Tables S10-S12**). The only randomized study identified was a pilot trial with 10 participants (56) that had concerns in several domains related to randomization, blinding, outcome measurement, and reporting the findings. The most common sources of potential bias in other non-randomized studies (experimental and cohort) were related to unreliable outcome measurement, incomplete follow-up, and not accounting for confounding factors.

Interventions identified as potentially preventing post Covid-19 condition included standard medication (14, 19, 36, 41, 47, 50, 56-59), traditional/ayurvedic medication (52, 53), stem cell therapy (49), rehabilitation or similar therapies (46, 51, 54), and screening and referrals (48, 55). We grouped interventions by timing, as those implemented during acute (symptom onset to 4 weeks) phase of Covid-19 with outcomes assessed at shorter (12-16 weeks) or longer (>16 weeks) follow-up duration post Covid-19; and interventions implemented during early post-acute (4-8 weeks) phase of Covid-19 with aforementioned shorter and longer follow-up time (**Table 3**).

**Table 3:**
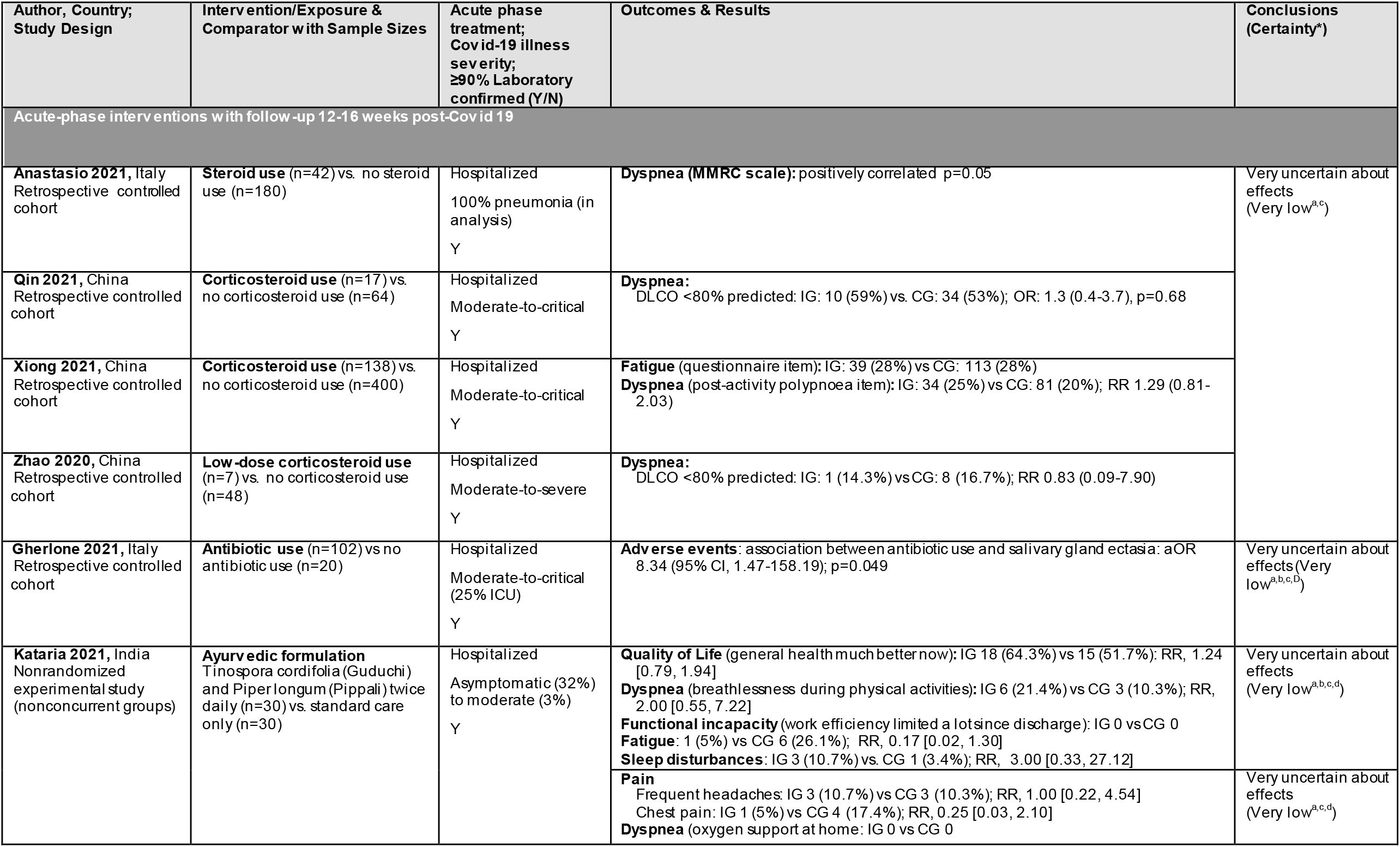
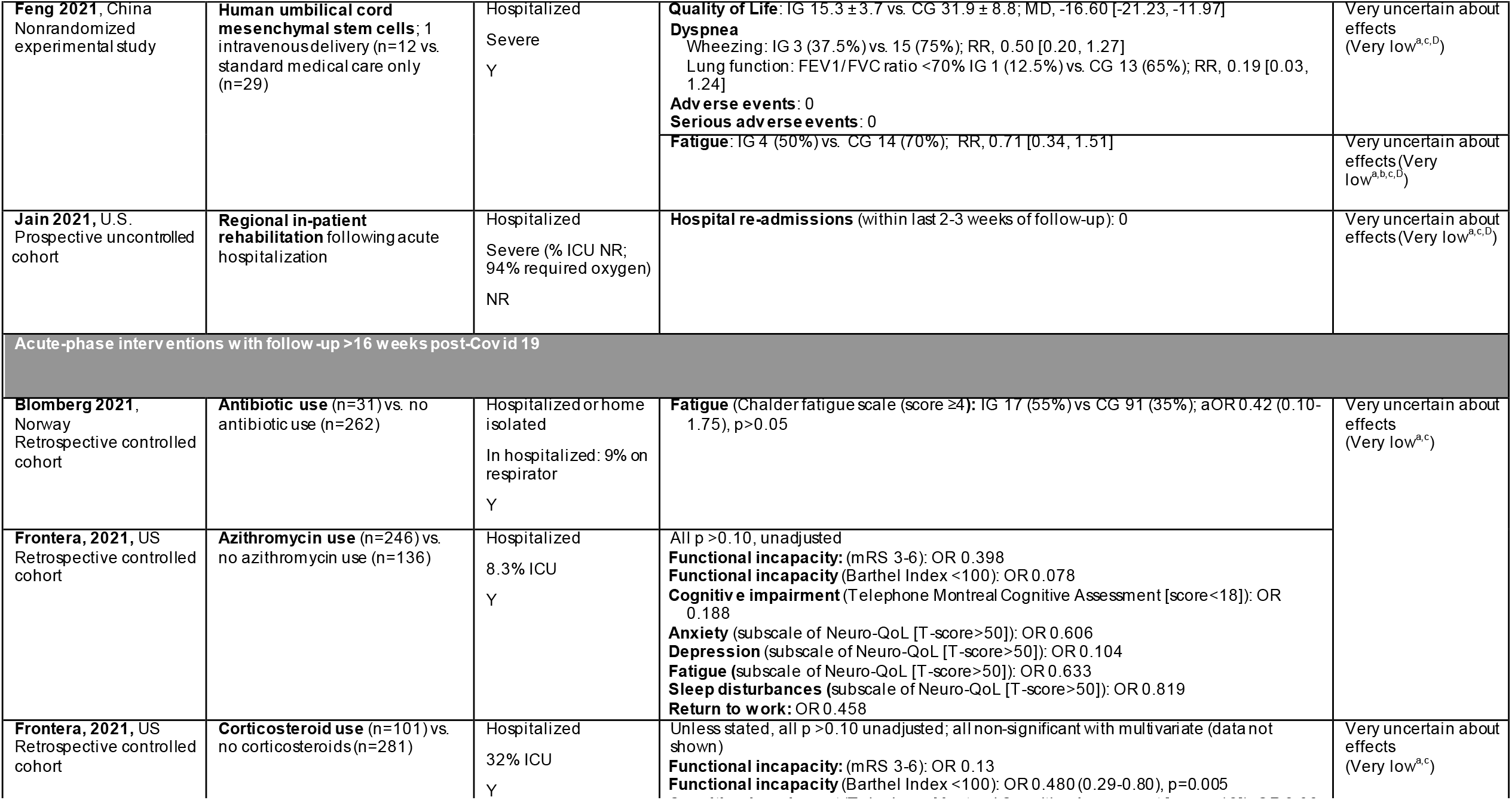
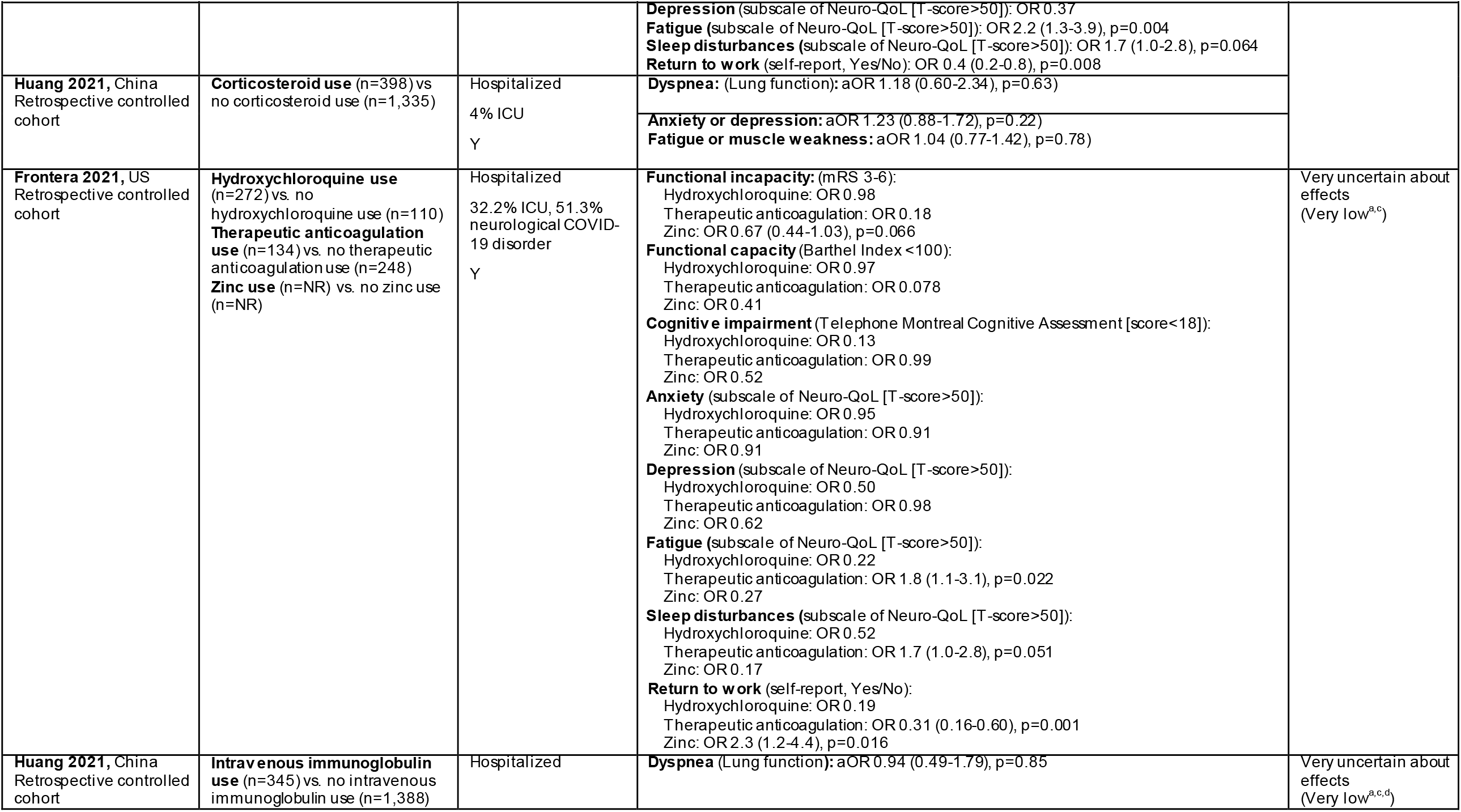
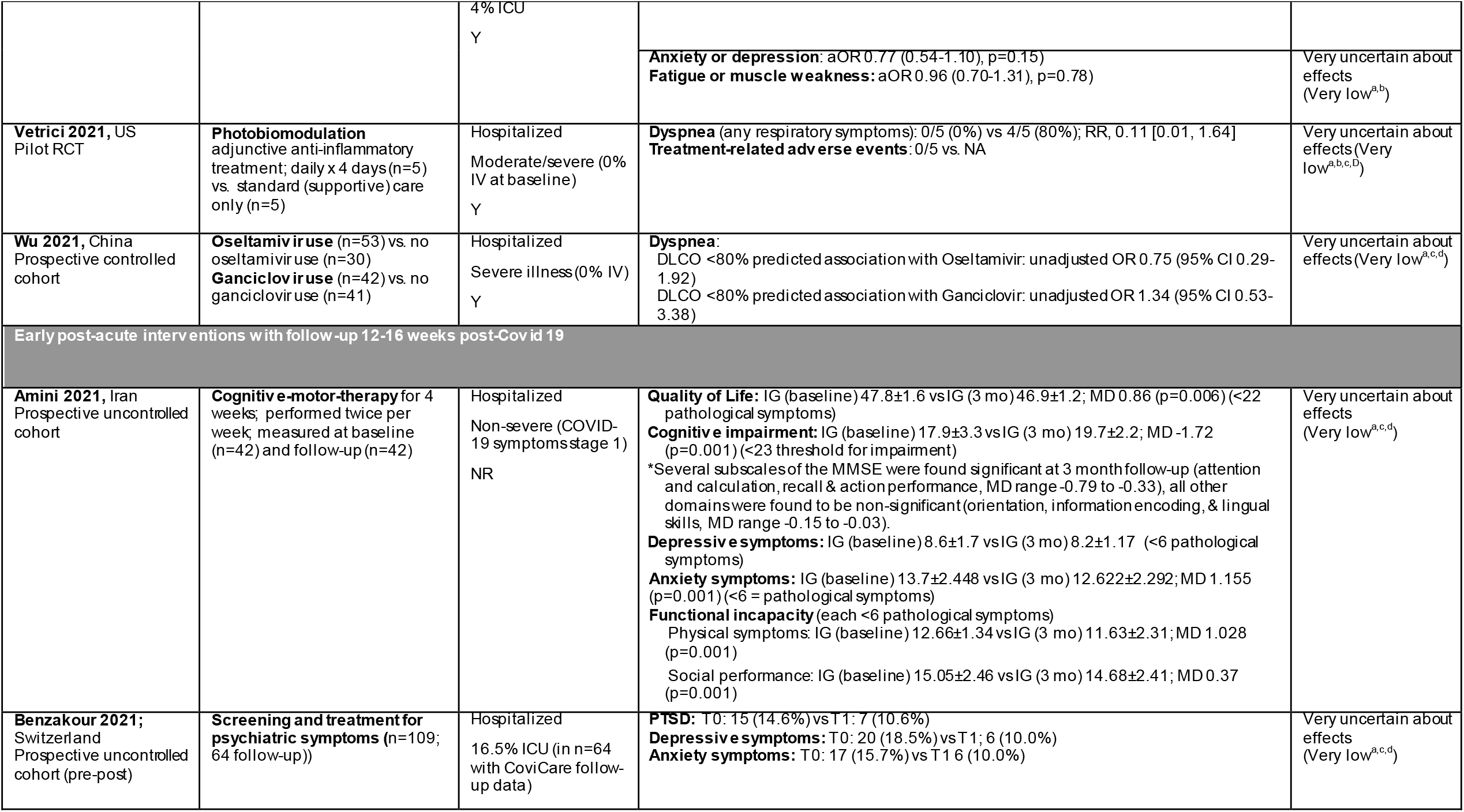
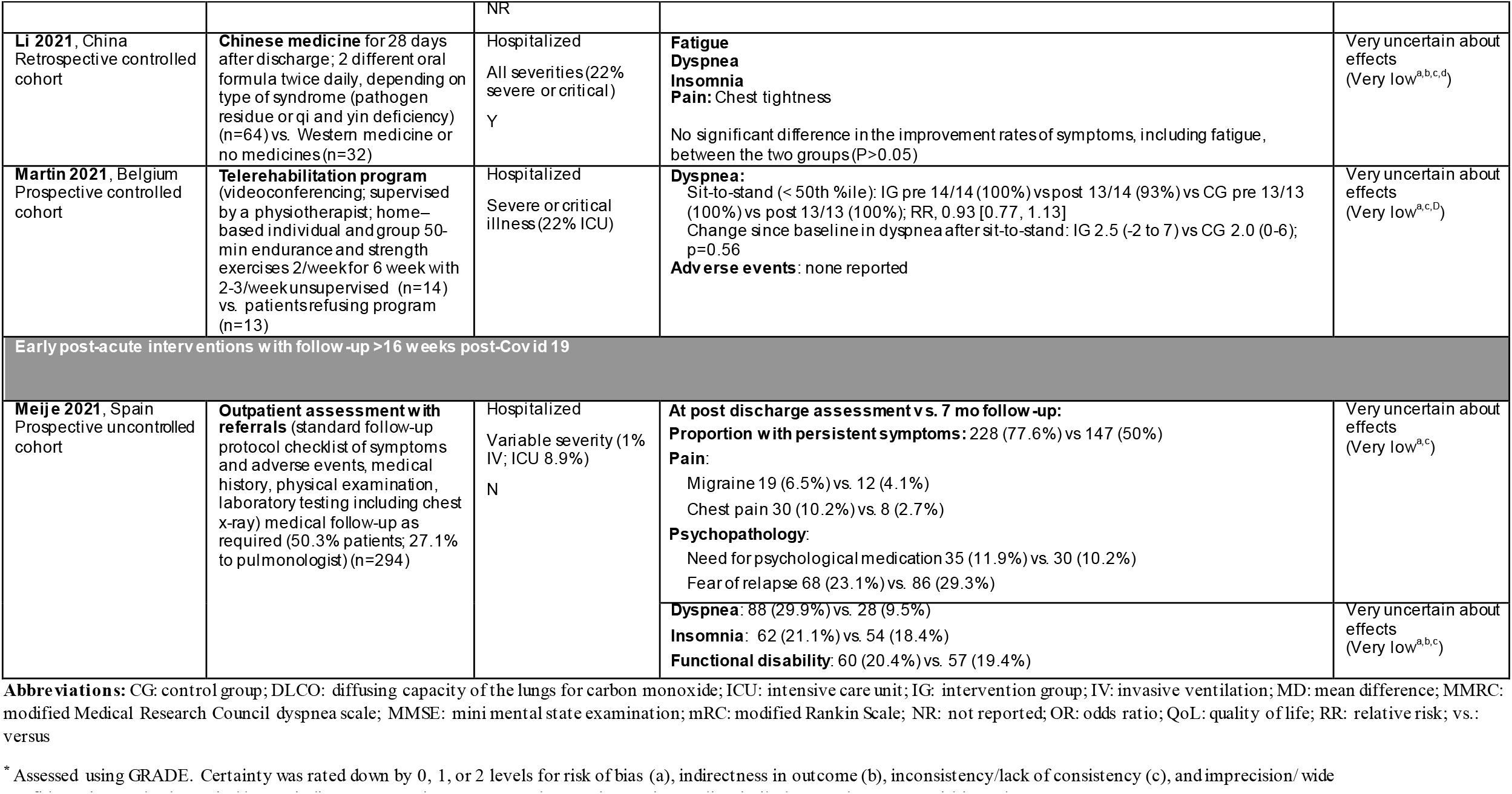
Summary of evidence for preventive interventions for post Covid-19 condition, by timing of implementation and follow-up duration.

With the exception of steroids and antibiotics, we identified single studies for all interventions. Across all intervention-outcome comparisons, the certainty of evidence was found to be very low, mainly due to risk of bias, inconsistency/lack of consistency (i.e., single study effect), and in some instances imprecision.

Four studies (n=896) (41, 47, 58, 59) reported on steroid use during the acute phase in hospitalized patients with dyspnea assessed at 12-16 weeks post Covid-19. Two studies (n=675) (19, 36) reported on use of antibiotics during the acute phase in hospitalized and home-isolated participants, with fatigue assessed at ≥6 months after Covid-19 diagnosis. The certainty of evidence for both comparisons was assessed as very low due to serious concerns about risk of bias and inconsistency.

## DISCUSSION

These systematic reviews were conducted in response to the growing recognition of an emerging threat and anticipated burden of post Covid-19 condition.

### Risk factors

Most of the findings had low or very low certainty evidence, often due to concerns related to risk for bias and inconsistent findings across the studies (in case of a pooled estimate) or a single study effect, although generally the reported outcomes aligned well with the review question (i.e., no indirectness).

The only risk factors found to have a moderate certainty in their association with more than one post Covid-19 condition outcome were female sex and Covid-19 illness severity during the acute phase. Based on this evidence, being a female is probably (moderate certainty) associated with a small-to-moderate increase in post Covid-19 non-recovery, fatigue and dyspnea, while severe/critical acute Covid-19 (versus not severe/critical) has probably a large association with cognitive impairment (among those who were hospitalized with Covid-19), small-to-moderate association with non-recovery (among mixed populations), and little-to-no association with dyspnea (among those who were hospitalized with Covid-19). Among other risk factors, hospitalization during the acute phase may (low certainty) be associated with a large increase in non-recovery, dyspnea, and reduced return to work. We did not identify a large association for any other risk factor-outcome comparisons.

These findings add considerably to the existing evidence. A recent scientific report by the Belgian Healthcare Knowledge center identified female sex and level of care received during acute Covid-19 as potential risk factors for long-term outcomes following Covid-19 (60). The report was limited to studies conducted in Europe and the USA with symptoms reported at ≥4 weeks after the disease onset. Our review additionally involves a formal data synthesis and assessment of evidence certainty which were not included in the Belgian report. Findings from a recent rapid review by National Institute for Health and Care Excellence (NICE) also indicate that female sex and severity of acute Covid-19 illness (i.e., hospitalization, ICU admission) increase the risk of developing persistent symptoms after initial Covid-19 infection (30). Our certainty for these findings was higher, possibly because we restricted inclusion to higher quality studies (having adjustment) and followed more rigorous methods for study selection and data synthesis. Our findings are also supported by a rapid review of risk factors associated with chronic Covid-19 symptoms, undertaken by Alberta Health Services (AHS) (61). In their review, among multiple risk factors identified as having a significant association with post Covid-19 condition were female sex in previously hospitalized Covid-19 patients, and hospitalization and ICU admission in non-hospitalized and hospitalized populations, respectively. The findings in the AHS rapid review were based on statistical significance, while in our review we relied on the magnitude of association and its certainty using various criteria since lack of a statistical significance should not be taken as a lack of association (62). Compared with the AHS review, we limited outcomes to 30 days or beyond the date of Covid-19 diagnosis and employed a strict population size criterion for eligibility. Despite methodological variation across these reviews and timing of outcome assessment, findings support that there is probably a link between these risk factors and post Covid-19 outcomes. While the association between acute Covid-19 severity and persistent symptoms is plausible, it is yet not clear why females are at a higher risk of post Covid-19 condition. It has been suggested that this could be due to the likely effect of initial exposure as females are more commonly involved in industries that may have a greater exposure to the Covid-19 virus (e.g., healthcare, teaching) (63). Nonetheless, more research needs to be conducted to further establish these associations.

The certainty of evidence for individual pre-existing conditions was low or very low, and most of the evidence was for little-to-no association. Although existing evidence indicates older age might increase the risk of post Covid-19 symptoms regardless of the follow-up length (30, 61, 64, 65), most of our evidence was low certainty for little-to-no association apart from more functional incapacity in ≥60 versus <60 year old hospitalized patients. Our findings may differ from other reviews due to our strict eligibility criteria as we only included studies reporting outcomes at least 12 weeks after disease onset, to minimize the likelihood of symptoms associated with acute Covid-19, with at least 300 participants, and controlling for a minimum set of confounders (i.e., 2 or more of age, sex, comorbidities, and illness severity). Also, we excluded studies where all participants were admitted to an ICU to avoid overlapping of post Covid-19 condition with post-intensive care syndrome, which is characterized by similar features to post Covid-19 (66).

#### Limitation of the evidence and future direction

Our findings are mostly applicable to longer-term consequences of Covid-19 occurring ≥22 weeks after diagnosis or illness onset as we identified only a few studies with shorter follow-up length. Also, despite the large volume of data emerging, many studies came from a hospitalized population. Of all the included studies, we identified only one that was exclusively in children who were previously hospitalized with Covid-19 (40). The study reported on several potential risk factors including age, sex, acute Covid-19 severity, obesity, and allergic diseases, however, none were identified as having a strong association with outcomes of our review. Evidence was also sparse in relation to pre-existing socioeconomic variables (e.g., race/ethnicity, income, education, employment) and marginalized groups including indigenous communities, institutionalized populations, and persons with disability, despite being listed as priority populations for Covid-19 policies by several jurisdictions. Similarly, we did not locate studies reporting on associations between Covid-19 vaccination status and long-term consequences of Covid-19. Moreover, current studies are mostly based on self-reported outcome and exposure data, which could be subject to recall bias and misclassification, and may limit generalizability of the evidence to other populations (30). There is also the possibility of over/under-estimation of the reported associations as people from certain populations may be under-represented in primary studies (67). Further, based on the current literature, it is still challenging to determine if persistent symptoms are actually attributed to initial Covid-19 or would have occurred independent of the infection (60). To overcome these limitations, more robust evidence of risk factors is required particularly in non-hospitalized populations and community settings. Use of administrative data and establishing universal outcome definitions and assessment methods could ensure robustness of the evidence.

### Interventions

To the best of our knowledge, this is the only systematic review to explore interventions that could potentially prevent post Covid-19 outcomes evaluated at least 12 weeks after the disease onset. Based on the current evidence, we are very uncertain about the effect of any intervention to prevent persistent symptoms associated with Covid-19. Most of the evidence came from single studies in hospitalized patients with wide variation in sample size and methodology. Interventions reported in multiple studies (steroids and antibiotics) had either inconsistent findings or did not report on similar outcomes relevant to our review question. With the exception of the one pilot trial (56), all identified studies were non-randomized and all had concerns about risk of bias due to multiple issues.

Currently, there is some evidence suggesting that rehabilitation might have a beneficial effect on recovery from Covid-19 (68-71), and that there may be a positive effect of medications on post-acute clinical outcomes of Covid-19 (72, 73). These studies were not included in our synthesis due to either short follow-up time or the outcomes not being prioritized by the working group and patient panel for inclusion. Regarding Covid-19 vaccination as a possible preventive intervention, findings from a recent (published since our search) observational study indicate that receiving Covid-19 vaccine after getting infected might offer some protection against long Covid-19 outcomes (74). Other studies we identified from our search or have identified thereafter did not meet inclusion criteria (e.g., vaccination before infection (75, 76), indeterminate timing of vaccination (77), entire sample had post Covid-19 condition (78, 79)). Variations in definition of post Covid-19 outcomes and timing of assessment make it difficult to draw conclusions or compare the findings of our review with other studies. Additionally, a lack of well-designed randomized or quasi-randomized trials significantly limits our understanding of possible effects of any potentially preventive interventions. The living systematic review by a Cochrane group shows that the focus of scientific research is shifting from treatment of acute Covid-19 to post-acute and chronic phases of Covid-19 (68). Thus, continuous review and assessment of the rapidly emerging evidence is important to better shape our understanding as the body of evidence grows.

### Strengths and limitations of our review

We followed established guidelines for systematic reviews to provide methodologically rigorous syntheses of the available evidence on risk factors associated with post Covid-19 conditions and potentially preventive interventions. The review questions and outcomes were informed by input from clinical experts, stakeholders, and a patient panel. A wide range of risk factors and preventive interventions were considered in our reviews. Our findings are based on our certainty about associations/magnitudes of effect reaching predefined thresholds, which is informed by several factors, rather than reliance of statistical significance that is commonly used across the literature.

Despite these strengths, there are some limitations in our review. As with other reviews, there is a possibility of missing relevant studies by our search, although this was mitigated through searching grey literature resources and references of the included studies and relevant reviews. Involving experienced reviewers and selecting studies in duplicate further reduced the possibility of any important studies being missed. Our search was limited to English or French studies and this might have resulted in missing studies from jurisdictions where other languages are commonly used for publication. Further, although we selected decision thresholds for our certainty assessments, the findings might change if different thresholds were selected.

## CONCLUSION

Being a female is probably (moderate certainty) associated with a small-to-moderate increase in post Covid-19 non-recovery, fatigue and dyspnea, and having a severe or critical acute Covid-19 illness (versus not severe/critical) probably has a large (2-fold or more) association with increased cognitive impairment, a small-to-moderate association with more non-recovery, and little-to-no association with dyspnea. Though with low certainty, hospitalization during the acute phase may have large associations with more non-recovery and dyspnea and less return to work. All other evidence on risk factors was low or very low certainty. Evidence on possible preventive interventions was mostly in hospitalized patients and observational in nature, and all provided very low certainty evidence. Continuous assessment of the rapidly emerging evidence is important to better shape our understanding as the body of evidence grows. Sufficiently powered prospective trials of preventive interventions are warranted.

## Policy implications

Post Covid-19 condition is becoming a public health challenge for many communities and governments. To be effective and efficient, public health policies and programs that will be rolled out in response to this challenge need careful evaluation and assessment of available resources. Synthesizing high-quality evidence to identify risk factors of persistent Covid-19 symptoms and possible preventive interventions is therefore important in highlighting areas where policy-makers can take action to mitigate the longer term outcomes of this pandemic. The wide range of post Covid-19 condition outcomes and associated risk factors emphasize the need for a coordinated multidisciplinary approach to assessment and management strategies. We found that females and people with severe or critical acute-phase Covid-19 are at greater risk of developing post-Covid complications. These findings imply that public health guidelines in relation to surveillance, screening services, and other services such as, access to sickness and disability benefits, might need to prioritize these groups. Interventions targeting fatigue, dyspnea, and cognitive impairment (especially in those with previous severe illness) may be good to prioritize for development and evaluation and to provide evidence on their effects. Further, inputs from patients and primary care providers should be taken into account when developing new care pathways and appropriate services, including long-term follow-up, rehabilitation and support groups, to ensure management and treatment strategies are tailored to patient needs and the disease manifestations.

## Contributors

JP and LH designed the study. SR, SG and AW screened citations for inclusion and were involved in data extraction and interpretation. JP, SR, SG and AW were involved is risk of bias assessments. All authors were involved in interpretation of the data. JP wrote the draft manuscript with input from all authors. All authors approved the final version of the manuscript. JP is the guarantors of this manuscript and accepts full responsibility for the work and the conduct of the study, had access to the data, and controlled the decision to publish. The corresponding author attests that all listed authors meet authorship criteria and that no others meeting the criteria have been omitted.

## Data Availability

All data produced in the present work are contained in the manuscript

## Acknowledgements

We would like to thank Sara Khangura and Shannon Hill (Canadian Agency for Drugs and Technologies in Health [CADTH]), Nana Amankwah and Gareth Leung (PHAC) for their assistance with conducting the reviews, the working group members (Alejandra Jaramillo Garcia, Francesca Reyes Domingo, Lisa Waddell, Naomi Tschirhart, Gisele Contreras, Patricia Corrin [PHAC]; Gino De Angelis [CADTH]; Curtis Cooper [The Ottawa Hospital, University of Ottawa]; Angela Cheung [University Health Network/Sinai Health System]) for their assistance with scoping the review, review of the protocol, and rating the outcomes, and the PHAC post Covid-19 condition patient panel (Jonah McGarva, Ruth Castellanos, Nicole Keefler, Tracey Thompson, Susan Goulding) for rating the outcomes. Alejandra Jaramillo Garcia, Francesca Reyes Domingo, and Naomi Tschirhart provided input on a draft version of this manuscript. We also thank information specialists Kate Merucci and Lynda Gamble (Health Canada) for conducting the database searches, Robyn Hocking (Health Canada) and Tara Landry (University of Alberta) for peer review), and Carolyn Spry and Nina Frey (CADTH) for conducting the grey literature search.

## Funding Acknowledgement(s) & Disclaimer

The Public Health Agency of Canada (PHAC) funded this work. The funder had no role in the conduct of this research, nor did it decide whether or not to publish the results.

Dr. Hartling is supported by a Canada Research Chair in Knowledge Synthesis and Translation, and is a Distinguished Researcher with the Stollery Science Lab supported by the Stollery Children’s Hospital Foundation.

## Competing interests

No author declares any competing interests.

## Ethics approval

This research did not involve human research participants and thus did not require ethics approval.

## Transparency declaration

The lead author of this work, JP, affirms that the manuscript is an honest, accurate, and transparent account of the study being reported; that no important aspects of the study have been omitted; and that any discrepancies from the study as originally planned in the protocol have been explained.

## Data sharing

All of the data extracted for this review are included in the manuscript and associated supplementary files.

## Notes

### Competing Interest Statement

The authors have declared no competing interest.

## References

1. World Health Organization. WHO Coronavirus (COVID-19) Dashboard: WHO; 2021 [cited 2021 December 10]. Available from: https://covid19.who.int/

2. Tenforde MW, Kim SS, Lindsell CJ, Billig Rose E, Shapiro NI, Files DC, et al. Symptom Duration and Risk Factors for Delayed Return to Usual Health Among Outpatients with COVID-19 in a Multistate Health Care Systems Network -United States, March-June 2020. MMWR Morb Mortal Wkly Rep. 2020;69(30):993–8.

3. Davis HE, Assaf GS, McCorkell L, Wei H, Low RJ, Re’em Y, et al. Characterizing long COVID in an international cohort: 7 months of symptoms and their impact. EClinicalMedicine. 2021;38:101019.

4. Kayaaslan B, Eser F, Kalem AK, Kaya G, Kaplan B, Kacar D, et al. Post-COVID syndrome: A single-center questionnaire study on 1007 participants recovered from COVID-19. J Med Virol. 2021;93(12):6566–74.

5. Centers for Disease Control and Prevention. Post-COVID Conditions Washington (DC): CDC; 2021 [cited 2021 December 14]. Available from: https://www.cdc.gov/coronavirus/2019-ncov/long-term-effects/index.html?CDC_AA_refVal=https%3A%2F%2Fwww.cdc.gov%2Fcoronavirus%2F2019-ncov%2Flong-term-effects.html.

6. Shah W, Hillman T, Playford ED, Hishmeh L. Managing the long term effects of covid-19: summary of NICE, SIGN, and RCGP rapid guideline. BMJ. 2021;372:n136.

7. The Lancet (Editorial). Facing up to long COVID. Lancet (London, England). 2020;396(10266):1861.

8. Nalbandian A, Sehgal K, Gupta A, Madhavan MV, McGroder C, Stevens JS, et al. Post-acute COVID-19 syndrome. Nat Med. 2021;27(4):601–15.

9. World Health Organization. A clinical case definition of post COVID-19 condition by a Delphi consensus, 6 October 2021. 2021.

10. Akbarialiabad H, Taghrir MH, Abdollahi A, Ghahramani N, Kumar M, Paydar S, et al. Long COVID, a comprehensive systematic scoping review. Infection. 2021;49(6):1163–86.

11. Office for National Statistics. Prevalence of ongoing symptoms following coronavirus (COVID-19) infection in the UK : 2 December 2021. Estimates of the prevalence of self-reported long COVID and associated activity limitation, using UK Coronavirus (COVID-19) Infection Survey data. UK: Office for National Statistics 2021.

12. Domingo FR, Waddell LA, Cheung AM, Cooper CL, Belcourt VJ, Zuckermann AME, et al. Prevalence of long-term effects in individuals diagnosed with COVID-19: an updated living systematic review2021 [cited 2021 December 14]. Available from: https://www.medrxiv.org/content/10.1101/2021.06.03.21258317v2.article-info.

13. Michelen M, Manoharan L, Elkheir N, Cheng V, Dagens A, Hastie C, et al. Characterising long COVID: a living systematic review. BMJ Glob Health. 2021;6(9).

14. Huang C, Huang L, Wang Y, Li X, Ren L, Gu X, et al. 6-month consequences of COVID-19 in patients discharged from hospital: a cohort study. Lancet. 2021;397(10270):220–32.

15. Sigfrid L, Drake TM, Pauley E, Jesudason EC, Olliaro P, Lim WS, et al. Long Covid in adults discharged from UK hospitals after Covid-19: A prospective, multicentre cohort study using the ISARIC WHO Clinical Characterisation Protocol. Lancet Reg Health Eur. 2021;8:100186.

16. Boscolo-Rizzo P, Guida F, Polesel J, Marcuzzo AV, Capriotti V, D’Alessandro A, et al. Long COVID In Adults at 12 Months After Mild-to-Moderate SARS-CoV-2 Infection. [Pre-print]. In press 2021.

17. Menges D, Ballouz T, Anagnostopoulos A, Aschmann HE, Fehr JS, Puhan MA, et al. Burden of post-COVID-19 syndrome and implications for healthcare service planning: A population-based cohort study. PLoS ONE. 2021;16(7):e0254523.

18. Peghin M, Palese A, Venturini M, De Martino M, Gerussi V, Graziano E, et al. Post-COVID-19 symptoms 6 months after acute infection among hospitalized and non-hospitalized patients. Clin Microbiol Infect. 2021;27(10):1507–13.

19. Frontera JA, Yang D, Lewis A, Patel P, Medicherla C, Arena V, et al. A prospective study of long-term outcomes among hospitalized COVID-19 patients with and without neurological complications. J Neurol Sci. 2021;426:117486.

20. Carson G. Research priorities for Long Covid: refined through an international multi-stakeholder forum. BMC Med. 2021;19(1):84.

21. Wise J. Long covid: WHO calls on countries to offer patients more rehabilitation. BMJ. 2021;372(405).

22. National Institutes of Health. NIH launches new initiative to study “Long COVID”: NIH; 2021 [cited 2021 December 15]. Available from: https://www.nih.gov/about-nih/who-we-are/nih-director/statements/nih-launches-new-initiative-study-long-covid.

23. Higgins JPT, Thomas J, Chandler J, Cumpston M, Li T, Page MJ, et al. Cochrane Handbook for Systematic Reviews of Interventions. 2nd ed. Higgins JPTTJ, Chandler J, Cumpston M, Li T, Page MJ, Welch VA, editor. Chichester (UK): John Wiley & Sons; 2019.

24. Moher D, Liberati A, Tetzlaff J, Altman DG, Group P. Preferred reporting items for systematic reviews and meta-analyses: the PRISMA statement. PLoS Med. 2009;6(7):e1000097.

25. Page MJ, McKenzie JE, Bossuyt PM, Boutron I, Hoffmann TC, Mulrow CD, et al. The PRISMA 2020 statement: an updated guideline for reporting systematic reviews. BMJ. 2021;372:n71.

26. Gates M, Pillay J, Wingert A, Guitard S, Rahman S, Zakher B, et al. Risk factors associated with severe outcomes of COVID-19: An updated rapid review to inform national guidance on vaccine prioritization in Canada. medRxiv. 2021.

27. WHO Working Group on the Clinical Characterisation and Management of COVID-19 infection. A minimal common outcome measure set for COVID-19 clinical research. Lancet Infect Dis. 2020;20(8):e192–e7.

28. Guyatt G, Oxman AD, Akl EA, Kunz R, Vist G, Brozek J, et al. GRADE guidelines: 1. Introduction-GRADE evidence profiles and summary of findings tables. J Clin Epidemiol. 2011;64(4):383–94.

29. McGowan J, Sampson M, Salzwedel DM, Cogo E, Foerster V, Lefebvre C. PRESS peer review of electronic search strategies: 2015 guideline statement. J Clin Epidemiol. 2016;75:40–6.

30. National Institute for Health and Care Excellence (NICE), Scottish Intercollegiate Guidelines Network (SIGN), Royal College of General Practitioners (RCGP). Covid-19 rapid guideline: managing the long-term effects of Covid-19 UK: NICE; 2021.

31. Tufanaru C, Munn Z, Aromataris E, Campbell J, Hopp L. Chapter 3: Systematic reviews of effectiveness. In: Aromataris E MZ, editor. Joanna Briggs Institute Reviewer’s Manual: The Joanna Briggs Institute; 2017.

32. Sterne JAC, Savović J, Page MJ, Elbers RG, Blencowe NS, Boutron I, et al. RoB 2: a revised tool for assessing risk of bias in randomised trials. BMJ. 2019;366:l4898.

33. Murad MH, Mustafa RA, Schünemann HJ, Sultan S, Santesso N. Rating the certainty in evidence in the absence of a single estimate of effect. Evid Based Med. 2017;22(3):85–7.

34. Iorio A, Spencer FA, Falavigna M, Alba C, Lang E, Burnand B, et al. Use of GRADE for assessment of evidence about prognosis: rating confidence in estimates of event rates in broad categories of patients. BMJ. 2015;350:h870.

35. Foroutan F, Guyatt G, Zuk V, Vandvik PO, Alba AC, Mustafa R, et al. GRADE Guidelines 28: Use of GRADE for the assessment of evidence about prognostic factors: rating certainty in identification of groups of patients with different absolute risks. J Clin Epidemiol. 2020;121:62–70.

36. Blomberg B, Mohn KG-I, Brokstad KA, Zhou F, Linchausen DW, Hansen B-A, et al. Long COVID in a prospective cohort of home-isolated patients. Nat Med. 2021.

37. Liu Y-H, Wang Y-R, Wang Q-H, Chen Y, Zhu J, Li W, et al. Post-infection cognitive impairments in a cohort of elderly patients with COVID-19. Molecular Neurodegeneration. 2021;16(1):48.

38. Munblit D, Bobkova P, Spiridonova E, Shikhaleva A, Gamirova A, Blyuss O, et al. Incidence and risk factors for persistent symptoms in adults previously hospitalised for COVID-19. Clin Exp Allergy. 2021;51:1107– 20.

39. Nehme M, Braillard O, Chappuis F, Guessous I, Courvoisier DS. Prevalence of Symptoms More Than Seven Months After Diagnosis of Symptomatic COVID-19 in an Outpatient Setting. Ann Intern Med. 2021;174(9):1252–60.

40. Osmanov IM, Spiridonova E, Bobkova P, Gamirova A, Shikhaleva A, Andreeva M, et al. Risk factors for long covid in previously hospitalised children using the ISARIC Global follow-up protocol: A prospective cohort study. Eur Respir J. 2021;59(2):2101341.

41. Qin W, Chen S, Hu B, Zhu Z, Li F, Wang X, et al. Diffusion capacity abnormalities for carbon monoxide in patients with COVID-19 at 3-month follow-up. Eur Respir J. 2021;58(1):036772020.

42. Qu G, Sun Y, Zhen Q, Wang W, Yu Y, Yong L, et al. Health-related quality of life of COVID-19 patients after discharge: A multicenter follow-up study. J Clin Nurs. 2021;30(11-12):1742-50.

43. Shang YF, Yu JN, Xu XR, Zhou FL, Liu T, Wang XH, et al. Half-year follow-up of patients recovering from severe COVID-19: Analysis of symptoms and their risk factors. J Intern Med. 2021.

44. Stavem K, Einvik G, Ghanima W, Olsen MK, Gilboe HM. Prevalence and determinants of fatigue after covid-19 in non-hospitalized subjects: A population-based study. Int J Environ Health Res. 2021;18(4):1–11.

45. Westerlind E, Palstam A, Sunnerhagen KS, Persson HC. Patterns and predictors of sick leave after Covid-19 and long Covid in a national Swedish cohort. BMC Public Health. 2021;21(1):1023.

46. Amini A, Vaezmousavi M, Shirvani H. The effectiveness of cognitive-motor training on reconstructing cognitive health components in older male adults, recovered from the COVID-19. Neurol Sci. 2022;43(2):1395–403.

47. Anastasio F, Barbuto S, Scarnecchia E, Cosma P, Fugagnoli A, Rossi G, et al. Medium-term impact of COVID-19 on pulmonary function, functional capacity and quality of life. Eur Respir J. 2021;58(3).

48. Benzakour L, Braillard O, Mazzola V, Gex D, Nehme M, Perone SA, et al. Impact of peritraumatic dissociation in hospitalized patients with COVID-19 pneumonia: A longitudinal study. J Psychiatr Res. 2021;140:53–9.

49. Feng G, Shi L, Huang T, Ji N, Zheng Y, Lin H, et al. Human Umbilical Cord Mesenchymal Stromal Cell Treatment of Severe COVID-19 Patients: A 3-Month Follow-Up Study Following Hospital Discharge. Stem Cells Dev. 2021;30(15):773–81.

50. Gherlone EFP, E./Tete, G./De Lorenzo, R./Magnaghi, C./Rovere Querini, P./Ciceri, F. Frequent and Persistent Salivary Gland Ectasia and Oral Disease After COVID-19. J Dent Res. 2021;100(5):464–71.

51. Jain E, Harmon EY, Sonagere MB. Functional outcomes and post-discharge care sought by patients with COVID-19 compared to matched controls after completing inpatient acute rehabilitation. PM R. 2021;13(6):618–25.

52. Kataria S, Sharma P, Ram JP, Deswal V, Singh M, Rana R, et al. A pilot clinical study of an add-on Ayurvedic formulation containing Tinospora cordifolia and Piper longum in mild to moderate COVID-19. J Ayurveda Integr Med. 2022;13(2):100454.

53. Li L, Gou CY, Li XM, Song WY, Wang XJ, Li HY, et al. Effects of Chinese Medicine on Symptoms, Syndrome Evolution, and Lung Inflammation Absorption in COVID-19 Convalescent Patients during 84-Day Follow-up after Hospital Discharge: A Prospective Cohort and Nested Case-Control Study. Chin J Integr Med. 2021;27(4):245–51.

54. Martin I, Braem F, Baudet L, Poncin W, Fizaine S, Aboubakar F, et al. Follow-up of functional exercise capacity in patients with COVID-19: It is improved by telerehabilitation. Respir Med. 2021;183:106438.

55. Meije Y, Duarte-Borges A, Sanz X, Clemente M, Ribera A, Ortega L, et al. Long-term outcomes of patients following hospitalization for coronavirus disease 2019: a prospective observational study. Clin Microbiol Infect. 2021;27(8):1151–7.

56. Vetrici MA, Mokmeli S, Bohm AR, Monici M, Sigman SA. Evaluation of Adjunctive Photobiomodulation (PBMT) for COVID-19 Pneumonia via Clinical Status and Pulmonary Severity Indices in a Preliminary Trial. J Inflamm Res. 2021;14:965–79.

57. Wu X, Liu X, Zhou Y, Yu H, Li R, Zhan Q, et al. 3-month, 6-month, 9-month, and 12-month respiratory outcomes in patients following COVID-19-related hospitalisation: a prospective study. Lancet Respir Med. 2021;9(7):747–54.

58. Xiong Q, Xu M, Li J, Liu Y, Zhang J, Xu Y, et al. Clinical sequelae of COVID-19 survivors in Wuhan, China: a single-centre longitudinal study. Clin Microbiol Infect. 2021;27(1):89–95.

59. Zhao YM, Shang YM, Song WB, Li QQ, Xie H, Xu QF, et al. Follow-up study of the pulmonary function and related physiological characteristics of COVID-19 survivors three months after recovery. EClinicalMedicine. 2020;25:100463.

60. Castanares-Zapatero D, Kohn L, Dauvrin M, Detollenaere J, Maertens de Noordhout C, Primus-de Jong C, et al. Long COVID: Pathophysiology – epidemiology and patient needs. Brussels: Health Services Research (HSR) Brussels: Belgian Health Care Knowledge Centre (KCE). KCE Reports 344. D/2021/10.273/31; 2021. Contract No.: KCE Reports 344. D/2021/10.273/31.

61. Covid-19 Scientific Advisory Group. Rapid Evidence Report: Updated review of prolonged symptoms after acute COVID-19 infection. Alberta: Alberta Health Services 2021.

62. Sullivan GM, Feinn R. Using Effect Size-or Why the P Value Is Not Enough. J Grad Med Educ. 2012;4(3):279–82.

63. Office for National Statistics. Which occupations have the highest potential exposure to the coronavirus (COVID-19)? UK: Office for National Statistics 2020 [cited 2022 January 05]. Available from: https://www.ons.gov.uk/employmentandlabourmarket/peopleinwork/employmentandemployeetypes/articles/whichoccupationshavethehighestpotentialexposuretothecoronaviruscovid19/2020-05-11.

64. Ayoubkhani D, Khunti K, Nafilyan V, Maddox T, Humberstone B, Diamond I, et al. Post-covid syndrome in individuals admitted to hospital with covid-19: retrospective cohort study. BMJ (Clinical research ed). 2021;372:n693.

65. Sudre CH, Murray B, Varsavsky T, Graham MS, Penfold RS, Bowyer RC, et al. Attributes and predictors of long COVID. Nat Med. 2021;27(4):626–31.

66. Hermans G, Van den Berghe G. Clinical review: intensive care unit acquired weakness. Crit Care. 2015;19(1):274.

67. English KC, Espinoza J, Pete D, Tjemsland A. A Comparative Analysis of Telephone and In-Person Survey Administration for Public Health Surveillance in Rural American Indian Communities. J Public Health Manag Pract. 2019;25 Suppl 5, Tribal Epidemiology Centers: Advancing Public Health in Indian Country for Over 20 Years:S70-S6.

68. de Sire A, Andrenelli E, Negrini F, Patrini M, Lazzarini SG, Ceravolo MG. Rehabilitation and COVID-19: a rapid living systematic review by Cochrane Rehabilitation Field updated as of December 31st, 2020 and synthesis of the scientific literature of 2020. Eur J Phys Rehabil Med. 2021;57(2):181–8.

69. Curci C, Negrini F, Ferrillo M, Bergonzi R, Bonacci E, Camozzi DM, et al. Functional outcome after inpatient rehabilitation in postintensive care unit COVID-19 patients: findings and clinical implications from a real-practice retrospective study. Eur J Phys Rehabil Med. 2021;57(3):443–50.

70. Liu K, Zhang W, Yang Y, Zhang J, Li Y, Chen Y. Respiratory rehabilitation in elderly patients with COVID-19: A randomized controlled study. Complement Ther Clin Pract. 2020;39:101166.

71. Hameed F, Palatulan E, Jaywant A, Said R, Lau C, Sood V, et al. Outcomes of a COVID-19 recovery program for patients hospitalized with SARS-CoV-2 infection in New York City: A prospective cohort study. PM R. 2021;13(6):609–17.

72. Belcaro G, Cornelli U, Cesarone MR, Scipione C, Scipione V, Hu S, et al. Preventive effects of Pycnogenol(R) on cardiovascular risk factors (including endothelial function) and microcirculation in subjects recovering from coronavirus disease 2019 (COVID-19). Minerva Med. 2021.

73. Cesarone MR, Hu S, Belcaro G, Cornelli U, Feragalli B, Corsi M, et al. Pycnogenol(R)-Centellicum(R) supplementation improves lung fibrosis and post-COVID-19 lung healing. Minerva Med. 2022;113(1):135–40.

74. Office for National Statistics. Coronavirus (COVID-19) vaccination and self-reported long COVID in the UK: 25 October 2021. UK: Office of National Statistics 2021 25 October 2021.

75. Antonelli M, Penfold RS, Merino J, Sudre CH, Molteni E, Berry S, et al. Risk factors and disease profile of post-vaccination SARS-CoV-2 infection in UK users of the COVID Symptom Study app: a prospective, community-based, nested, case-control study. Lancet Infect Dis. 2022;22(1):43–55.

76. Taquet M, Dercon Q, Harrison PJ. Six-month sequelae of post-vaccination SARS-CoV-2 infection: a retrospective cohort study of 10,024 breakthrough infections. medRxiv. 2021:2021.10.26.21265508.

77. Kuodi P, Gorelik Y, Zayyad H, Wertheim O, Wiegler KB, Jabal KA, et al. Association between vaccination status and reported incidence of post-acute COVID-19 symptoms in Israel: a cross-sectional study of patients tested between March 2020 and November 2021. medRxiv. 2022:2022.01.05.22268800.

78. Arnold D, Milne A, Samms E, Stadon L, Maskell N, Hamilton F. Symptoms After COVID-19 Vaccination in Patients With Persistent Symptoms After Acute Infection: A Case Series. Ann Intern Med. 2021;174(9):1334–6.

79. Massey D, Berrent D, Akrami A, Assaf G, Davis H, Harris K, et al. Change in Symptoms and Immune Response in People with Post-Acute Sequelae of SARS-Cov-2 Infection (*PASC*) After SARS-Cov-2 Vaccination. medRxiv. 2021:2021.07.21.21260391.

