## Supplemental file for "Risk factors and preventive interventions for post Covid-19 condition: systematic reviews"

|  | **Page number** |
| --- | --- |
| **Table S1. Eligibility for Key Question 1** | **2** |
| **Table S2. Eligibility for Key Question 2** | **4** |
| **Search Strategy for Key Question 1** | **6** |
| **Search Strategy for Key Question 2** | **11** |
| **Excluded Studies List for Key Question 1** | **43** |
| **Excluded Studies List for Key Question 2** | **81** |
| **Table S3. Study Characteristics for Key Question 1** | **101** |
| **Table S4. Risk of Bias Assessments for Key Question 1** | **108** |
| **Table S5. Detailed Summary of Findings for Risk Factors of Post-Covid Condition in Children** | **110** |
| **Table S6. Detailed Summary of Findings for Socio-demographic Risk Factors for Post Covid-19 Condition** | **110** |
| **Table S7. Detailed Summary of Findings for Risk Factors Related to Pre-existing Conditions** | **117** |
| **Table S8. Detailed Summary of Findings for Risk Factors Related to Covid-19 Illness Severity** | **121** |
| **Table S9. Study Charactertistcis for Key Quesiton 2** | **127** |
| **Table S10: Risk of Bias for Key Question 2 Cohort Studies** | **139** |
| **Table S11: Risk of Bias for Key Question 2 Quasi-experimental Studies** | **143** |
| **Table S12: Risk of Bias for Key Question 2 Randomized Trials** | **144** |

**Table S1. Eligibility for Key Question 1**

| Domain | Inclusion criteria | Exclusion criteria |
| --- | --- | --- |
| Population | 1. People of any age from general population 2. People of any age who have had Covid-19   Minimum sample size 300 people who have had Covid-19  Subgroup analysis (for Covid-19 patients):   - Laboratory confirmed (using RT-PCR or antigen test) via records (≥90%) vs otherwise - ICU vs hospitalized vs mixed vs out-patient (<10% hospitalized) during acute phase | All/majority (>50%) of patients who have received assessment and advice/follow-up at a long/post Covid-19 clinic or similar.  All participants experiencing/seeking care at 12+ weeks post Covid-19 for symptoms  Most of population (≥80%) in ICU |
| Exposure & Comparison(s) | Risk factors  Pre-existing: age (e.g., <18 vs 19-49 vs 50-69 vs ≥70); sex (M vs F); gender identity (as reported by authors); BMI (e.g., <30 vs 30-39 vs ≥40 kg/m2); race/ethnicity (any as reported vs Caucasian); ≥2 and 1 vs 0 comorbidities; specific chronic conditions affecting a relatively large population and shown to lead to more severe Covid-19, based on clinical input and from having large associations with severe Covid-19 (Gates et al. MedRxiv doi: https://doi.org/10.1101/2021.04.23.21256014): i.e., diabetes, asthma, chronic lung disease, immunodeficiency (including use of immunosuppressants and transplantation), autoimmune conditions, psychiatric disorder, active/treated cancer); pregnancy; disability; vaccination status (without/before having had Covid-19); income; place of residence (e.g., remote, overcrowding, homeless, institutionalization); occupation; and immigration or refugee status; others will be considered.  Clinical factors during acute phase: presence of dyspnea (self-reported or based on tests), presence of binding antibodies to SARS-CoV-2, number of symptoms/symptom severity (e.g. <5 vs ≥5), need for hospitalization, need for intensive ventilation (e.g, grade 4 or 5 WHO Covid severity), need for ICU admission | Laboratory findings other than used for determining dyspnea or binding antibodies  Variations of severity or duration of chronic conditions (e.g. controlled vs uncontrolled diabetes) |
| Outcomes | 1. Non-recovery or full recovery to pre-Covid health (not having 1+ ongoing Covid-related symptoms; other author-defined recovery rates) 2. major cardiovascular event or organ impairment 3. experiencing moderate/severe or persistent (≥3 wks) fatigue 4. experiencing moderate/severe or persistent (≥3 wks) breathlessness/dyspnea (e.g. ≥2 on MRC breathlessness scale) 5. experiencing important impact on quality of life (e.g., ≥0.1 change on 1 point scale [EuroQuol5D or VAS, other composite scores on validated scales covering multiple domains of quality of life] from pre-Covid) 6. experiencing clinical/pathological levels of psychopathology i.e., anxiety, depression, PTSD 7. experiencing moderate/severe or persistent (≥3 wks) impairment in functional capacity (e.g. ≥ grade 2 on Post-Covid-19 Functional Status [PCFS] scale) 8. experiencing moderate/severe or persistent (≥3 wks) cognitive impairment 9. experiencing moderate/severe or persistent (≥3 wks) sleep disturbances 10. experiencing moderate/severe or persistent (≥3 wks) pain including headaches and chest pain 11. unable to return to full-time work/school/education or caring role (as applicable)   Data for slightly different outcomes (e.g. mild-to-moderate impairments, author-defined clinically meaningful difference thresholds) may be included, as feasible. |  |
| Timing of events/outcome assessment | Minimum 12 weeks after Covid-19 diagnosis (if only mean duration reported we will accept if mean – 1 SD ≥12 wks)  When authors did not report duration from symptom onset we made assumptions that symptom onset to hospital admission was 7 days and to hospital discharge was 4 weeks.  Subgroup analyses:   - Early in post Covid-19 phase (12-22 wks postCovid-19) - Long-term ($\geq$22 wks/6 mos post Covid-19) |  |
| Study design, analyses & publication type | Prospective and retrospective observational studies using participant-level data, including usual care control groups in intervention studies or trials  Requiring adjustment for at least 2 of the following: age, sex, pre-existing comorbidities, Covid-19 illness severity. Studies may stratify by one variable e.g. age and adjust for another e.g. sex in the same analysis.  Peer-reviewed articles or pre-prints/reports of original research | Case series where all participants have risk factor of interest e.g. comorbid condition (with exception of hospitalization)  Studies where the exposure to the risk factor is not defined as occurring before the outcome assessment |
| Country | Any  Subgroup analysis:   - High-income OECD countries vs other countries |  |
| Language of full text | English or French | We will document studies possibly eligible based on tiles/abstracts but not included because of publications in other languages. |

**Tables S2. Eligibility for Key Question 2**

| Domain | Inclusion criteria | Exclusion criteria |
| --- | --- | --- |
| Population | People of any age who had Covid-19 up to 8 weeks previous to enrollment/baseline data collection (we will accept 20% of participants at 8-12 weeks in post-acute phase)  Minimum sample size 30 per group, will accept smaller samples (n>5) to fill gaps where no other studies exist for the intervention  Subgroup analysis:   - Acute (<4 wks since diagnosis or symptom onset [≥80%]) vs post-acute/ongoing symptomatic phase (4-12 wks) - Laboratory confirmed via records (≥90%) vs otherwise - ICU vs hospitalized vs mixed vs out-patient (<10% hospitalized) during acute phase - Age (e.g. <18 vs 19-49 vs 50-64 vs ≥65), sex (M vs F), gender, race/ethnicity, SES, comorbid conditions - Symptom severity (as defined by authors) | Most (≥80%) of population was in ICU |
| Interventions | Interventions starting up to 8 weeks post-Covid (in ≥80% participants):   - Vaccination after Covid-19 diagnosis - Single and combinations of medications (e.g. dexamethasone, specific antivirals) and/or vitamins/supplements (e.g., vitamin D, zinc) - Self-management advice - Self-management programs, with or without tailoring & support groups - Multidisciplinary care models/pathways with comprehensive centralized screening/triage, and, if indicated, assessment and direct treatment and/or referrals to community/primary care or specialist clinics - Primary care treatment or referrals with/without screening/assessment - Rehabilitation clinics (focused on cardiopulmonary intervention(s)) - Single discipline interventions, e.g. psychology, psychiatry, cardiology, occupational therapy, nutritional counselling - Others (e.g., Indigenous health practices, Ayurvedic medicine, combinations of above)   When authors did not report duration from symptom onset we made assumptions that symptom onset to hospital admission was 7 days and to hospital discharge was 4 weeks.  Subgroup analysis:  Online vs in-person; screening/assessment component (Y/N); timing of intervention (≤4 vs >4-8 wks after Covid-19 diagnosis) | Studies focused on post-ICU syndrome  Studies starting the intervention ≥8 wks post-Covid will be excluded but documented |
| Comparators | Usual medical care  No comparator, if feasible and no other studies with comparators for intervention, but reporting baseline and follow-up or change scores for continuous outcomes  We will consider including studies with active control arms, if feasible. |  |
| Outcomes | A. Patient-important benefits    1. Non-recovery or full recovery to pre-Covid health (not having 1+ ongoing Covid-19 related symptoms) 2. major cardiovascular event or organ impairment 3. experiencing moderate/severe or persistent (≥3 wks) fatigue 4. experiencing moderate/severe or persistent (≥3 wks) breathlessness/dyspnea (e.g. ≥2 on MRC breathlessness scale) 5. experiencing important impact on quality of life (e.g., ≥0.1 change on 1 point scale [EuroQuol5D or VAS, other validated scales covering multiple domains of quality of life) from pre-Covid) 6. experiencing clinical/pathological levels of psychopathology i.e., anxiety, depression, PTSD 7. experiencing moderate/severe or persistent (≥3 wks) impairment in functional capacity (e.g. ≥ grade 2 on Post-Covid-19 Functional Status [PCFS] scale) 8. experiencing moderate/severe or persistent (≥3 wks) cognitive impairment 9. experiencing moderate/severe or persistent (≥3 wks) sleep disturbances 10. experiencing moderate/severe or persistent (≥3 wks) pain 11. unable to return to full-time work/school/education or caring role (as applicable)   Data for slightly different outcomes (e.g. mild-to-moderate impairments, author-defined clinically meaningful difference thresholds) may be included, as feasible.   B. Patient-important harms attributed to the intervention 1. Any adverse effects of the interventions (i.e., number having 1+ adverse effects) 2. Serious AEs [number and descriptions]) C. Secondary outcomes via healthcare system/utilization outcomes 1. all-cause hospital admission 2. emergency department visits 3. requiring urgent care outside of hospital 4. requiring referral for specialist care for physical and/or mental health (may include new onset of disease such as diabetes) 5. requiring pulmonary rehabilitation and/or long-term oxygen therapy D. Implementation outcomes (if one of A to C above is also reported), e.g. adherence, time for assessment, referral times, case load | Studies focused on post-ICU syndrome  Studies excluded for not measuring one of the outcomes will be documented |
| Timing of follow-up | - Minimum follow-up 21 days/3 wks - Outcomes measured ≥ 12 wks post Covid-19 (in ≥80%)   Subgroup analysis: outcomes measured 12-16 wks vs longer |  |
| Study design, analyses & publication type | - Randomized studies - Quasi-randomized or experimental studies (e.g. controlled before-after, interrupted time series, before-after studies) - Prospective cohort studies with control groups - Retrospective cohort studies with control groups and case-control studies - Case series/uncontrolled cohorts, if baseline and follow-up scores reported for patient-reported measures   Peer reviewed articles or pre-prints of original research | Case reports and series ≤5 pts |
| Country | Any  Subgroup analysis:   - High-income OECD countries vs other countries |  |
| Language of full text | English or French | We will document studies possibly eligible based on tiles/abstracts but not included because of publications in other languages. |

**Search Strategy for Key Question 1**

Ovid Medline(R) ALL

| **#** | **Searches** | **Results** |
| --- | --- | --- |
| 1 | (COVID or COVID-19 or COVID19 or coronavirus* or corona virus* or 2019-nCoV or 19nCoV or 2019nCoV or nCoV or n-CoV or SARS-CoV-2 or SARS-CoV2 or SARSCoV-2 or SARSCoV2 or 2019-novel CoV or Sars-coronavirus2 or novel CoV or pandemic or PASC).ti. | 165745 |
| 2 | (((long or longterm or longhaul or post acute or postacute or after acute or sequela* or protracted or post-infect* or post-viral or post-discharg* or non-recover* or nonrecover* or PASC) adj7 (COVID or COVID-19 or COVID19 or coronavirus* or corona virus* or 2019-nCoV or 19nCoV or 2019nCoV or nCoV or n-CoV or SARS-CoV-2 or SARS-CoV2 or SARSCoV-2 or SARSCoV2 or 2019-novel CoV or Sars-coronavirus2 or novel CoV)) not (long term care* or longterm care)).mp. | 2947 |
| 3 | ((chronic or persist* or linger* or continuing or continual) adj3 (COVID or COVID-19 or COVID19 or coronavirus* or corona virus* or 2019-nCoV or 19nCoV or 2019nCoV or nCoV or n-CoV or SARS-CoV-2 or SARS-CoV2 or SARSCoV-2 or SARSCoV2 or 2019-novel CoV or Sars-coronavirus2 or novel CoV)).mp. | 979 |
| 4 | ((long or longterm or longhaul* or ongoing or chronic* or lengthy or protracted or sustained or persist* or linger* or continu* or post-acute or postacute or post-viral or post-discharg* or post-infect* or residual) adj3 (outcome* or symptom* or morbidit* or manifestation* or issue* or effect? or difficulties or challeng* or problem? or complication* or disturbance* or consequence* or impair* or dysfunction* or dysfunct* or function* or abnormalit* or dizziness or headache* or dyspnea* or fatigue or breath or breathing or lung or lungs or respiratory or tachycardi* or palpitation* or neuro* or concentration or concentrating or brain fog or cough* or ache* or pain* or taste* or tasting or smell* or olfact* or gustatory)).mp. and (COVID or COVID-19 or COVID19 or coronavirus* or corona virus* or 2019-nCoV or 19nCoV or 2019nCoV or nCoV or n-CoV or SARS-CoV-2 or SARS-CoV2 or SARSCoV-2 or SARSCoV2 or 2019-novel CoV or Sars-coronavirus2 or novel CoV).tw. | 4501 |
| 5 | (("90" or ninety or "120" or "180" or hundred*) adj2 day?).tw,kf. | 68638 |
| 6 | (("12" or twelve or "13" or thirteen or "14" or fourteen or "15" or "fifteen" or "16" or sixteen or "17" or seventeen or "18" or eighteen or "19" or nineteen or "20" or "twenty" or "21" or "22" or "23" or "24") adj3 (week? or wk or wks)).tw,kf. | 294228 |
| 7 | (("3" or three or "4" or four or "5" or five or "6" or six or "7" or seven or "8" or eight or "9" or nine or "10" or ten or "11" or eleven or "12" or twelve or "13" or thirteen or "14" or fourteen or "15" or "fifteen" or "16" or sixteen or "17" or seventeen or "18" or eighteen or "19" or nineteen or "20" or "twenty" or "21" or "22" or "23" or "24") adj3 (month? or mo or mos)).tw,kf. | 1340656 |
| 8 | (("1" or one or "2" or two or half or multi* or full) adj3 (year? or yr or yrs)).tw,kf. | 809570 |
| 9 | (("more than" or longer or "at least" or "after" or follow* or upward* or later* or prospectiv* or retrospectiv* or review* or longitud* or post or beyond) adj5 (day? or week? or wk or wks or month? or mo or mos or year? or yr or yrs)).tw,kf. | 2518578 |
| 10 | (COVID or COVID-19 or COVID19 or coronavirus* or corona virus* or 2019-nCoV or 19nCoV or 2019nCoV or nCoV or n-CoV or SARS-CoV-2 or SARS-CoV2 or SARSCoV-2 or SARSCoV2 or 2019-novel CoV or Sars-coronavirus2 or novel CoV).mp. | 180757 |
| 11 | (or/5-8) and 9 and 10 | 3557 |
| 12 | (postcovid or postcorona* or ((post* or survivor*) adj3 (COVID or COVID-19 or COVID19 or coronavirus* or corona virus* or 2019-nCoV or 19nCoV or 2019nCoV or nCoV or n-CoV or SARS-CoV-2 or SARS-CoV2 or SARSCoV-2 or SARSCoV2 or 2019-novel CoV or Sars-coronavirus2 or novel CoV))).tw,kf. | 3398 |
| 13 | (outcome* or symptom* or morbidit* or manifestation* or issue* or effect? or difficulties or challeng* or problem? or complication* or disturbance* or consequence* or impair* or dysfunction* or disfunction* or function* or abnormalit* or dizziness or headache* or dyspnea* or fatigue or breath or breathing or lung or lungs or respiratory or tachycardi* or palpitation* or neuro* or concentration or concentrating or brain fog or cough* or ache* or pain* or taste* or tasting or smell* or olfact* or gustatory).tw,kf. | 15615917 |
| 14 | 12 and 13 | 2411 |
| 15 | (COVID-19/ or Coronavirus infections/ or SARS-CoV-2/) and syndrome/ | 145 |
| 16 | 2 or 3 or 4 or 11 or 14 or 15 | 11782 |
| 17 | (Risk factor* or relative risk or odds ratio or between group* or Regression or multi-variate or multivaria* or covariate or univariate or co-variate or matching or ANOVA or Analysis of variance or ANCOVA or Correlation or Covariance or Principal Component Analysis or cohort* or follow-up or prognos* or predict*).mp. | 6309693 |
| 18 | exp cohort studies/ | 2188722 |
| 19 | (cohort* or longitudinal* or ((prospective* or retrospective*) adj3 (study or studies or research* or participant* or trial* or enrol* or data))).tw,kw,kf. | 1554956 |
| 20 | ("Associated with" or "Association of" or "impact of" or "Correlated with" or "Impact* on" or characteristics or characterise or features).ti. | 1005321 |
| 21 | (clinical data or (clinical adj5 (characteristics or features or manifestations))).tw,kf. | 422063 |
| 22 | 17 or 18 or 19 or 20 or 21 | 7755026 |
| 23 | 1 and 16 and 22 | 5270 |
| 24 | (exp animals/ or exp "models, animal"/ or exp animal experimentation/) not (human* or person*).hw. | 4858428 |
| 25 | (animal* or amphib* or amphipod* or ape or apes or avian or bat or bats or beagle* or bird or birds or bivalve* or boar? or bovine* or canin* or cat or cats or catfish* or cattle or chicken* or chimpanzee* or cow or cows or crustacean* or dog or dogs or equine or ewe? or fish or fishes or feline* or frog or frogs or fruit fly or fruit flies or gamefish* or gastropod* or gerbil* or goat* or guineapig* or hamster? or hare or hares or horse* or macaque* or litter? or lizard* or mammal* or marmoset* or mice or mollusk* or monkey* or mouse or murine or ostrich* or ovine or pig or piglet* or pigs or porcine or pork* or primate* or rabbit* or rat or rats or reptil* or rodent? or sheep or sprague-dawley or songbird* or squirrel* or sow or sows or swine* or toad? or veterinar* or worm* or zebrafish* or zebra fish*).ti. | 2839482 |
| 26 | (address or autobiography or bibliography or biography or case reports or classical article or clinical trial, veterinary or clinical trials, veterinary as topic or editorial or historical article or interview or news or newspaper article or observational study, veterinary).pt. | 3397375 |
| 27 | case reports/ or (case-stud* or case-report*).jw. or (case* and report*).ti. | 2322852 |
| 28 | (MIS-C or multisystem-inflammatory syndrome or lockdown* or lock* down* or restrictions or shelter-in-place or stay-at-home or school clos* or personal protective equipment or ppe or hygiene or close contact* or self isolat* or social distanc* or mask or masks).ti. | 41676 |
| 29 | 23 not (or/24-28) | 4559 |
| 30 | limit 29 to yr="2021 -Current" | 3062 |
| 31 | (202101* or 202102* or 202103* or 202104* or 202105* or 202106* or 202107* or 202108* or 202109* or 20211* or 2022*).ez,dt,ed. | 1579740 |
| 32 | 29 and 31 | 3211 |
| 33 | 30 or 32 | 3264 |

Ovid Embase

| **#** | **Searches** | **Results** |
| --- | --- | --- |
| 1 | (COVID or COVID-19 or COVID19 or coronavirus* or corona virus* or 2019-nCoV or 19nCoV or 2019nCoV or nCoV or n-CoV or SARS-CoV-2 or SARS-CoV2 or SARSCoV-2 or SARSCoV2 or 2019-novel CoV or Sars-coronavirus2 or novel CoV or pandemic or PASC).ti. | 166234 |
| 2 | (((long or longterm or longhaul or post acute or postacute or after acute or sequela* or protracted or post-infect* or post-viral or post-discharg* or non-recover* or nonrecover* or PASC) adj7 (COVID or COVID-19 or COVID19 or coronavirus* or corona virus* or 2019-nCoV or 19nCoV or 2019nCoV or nCoV or n-CoV or SARS-CoV-2 or SARS-CoV2 or SARSCoV-2 or SARSCoV2 or 2019-novel CoV or Sars-coronavirus2 or novel CoV)) not (long term care* or longterm care)).mp. | 2915 |
| 3 | ((chronic or persist* or linger* or continuing or continual) adj3 (COVID or COVID-19 or COVID19 or coronavirus* or corona virus* or 2019-nCoV or 19nCoV or 2019nCoV or nCoV or n-CoV or SARS-CoV-2 or SARS-CoV2 or SARSCoV-2 or SARSCoV2 or 2019-novel CoV or Sars-coronavirus2 or novel CoV)).mp. | 980 |
| 4 | ((long or longterm or longhaul* or ongoing or chronic* or lengthy or protracted or sustained or persist* or linger* or continu* or post-acute or postacute or post-viral or post-discharg* or post-infect* or residual) adj3 (outcome* or symptom* or morbidit* or manifestation* or issue* or effect? or difficult* or challeng* or problem* or complication* or disturbance* or consequence* or impair* or dysfunction* or disfunction* or function* or abnormalit* or dizziness or headache* or dyspnea* or fatigue or breath or breathing or lung or lungs or respiratory or tachycardi* or palpitation* or neuro* or concentration or concentrating or brain fog or cough* or ache* or pain* or taste* or tasting or smell* or olfact* or gustatory)).mp. and (COVID or COVID-19 or COVID19 or coronavirus* or corona virus* or 2019-nCoV or 19nCoV or 2019nCoV or nCoV or n-CoV or SARS-CoV-2 or SARS-CoV2 or SARSCoV-2 or SARSCoV2 or 2019-novel CoV or Sars-coronavirus2 or novel CoV).tw. | 7363 |
| 5 | (("90" or ninety or "120" or "180" or hundred*) adj2 day?).tw,kw. | 112746 |
| 6 | (("12" or twelve or "13" or thirteen or "14" or fourteen or "15" or "fifteen" or "16" or sixteen or "17" or seventeen or "18" or eighteen or "19" or nineteen or "20" or "twenty" or "21" or "22" or "23" or "24") adj3 (week? or wk or wks)).tw,kw. | 455587 |
| 7 | (("3" or three or "4" or four or "5" or five or "6" or six or "7" or seven or "8" or eight or "9" or nine or "10" or ten or "11" or eleven or "12" or twelve or "13" or thirteen or "14" or fourteen or "15" or "fifteen" or "16" or sixteen or "17" or seventeen or "18" or eighteen or "19" or nineteen or "20" or "twenty" or "21" or "22" or "23" or "24") adj3 (month? or mo or mos)).tw,kw. | 2081779 |
| 8 | (("1" or one or "2" or two or half or multi* or full) adj3 (year? or yr or yrs)).tw,kw. | 1269157 |
| 9 | (("more than" or longer or "at least" or "after" or follow* or upward* or later* or prospectiv* or retrospectiv* or review* or longitud* or post or beyond) adj5 (day? or week? or wk or wks or month? or mo or mos or year? or yr or yrs)).tw,kw. | 3759265 |
| 10 | (COVID or COVID-19 or COVID19 or coronavirus* or corona virus* or 2019-nCoV or 19nCoV or 2019nCoV or nCoV or n-CoV or SARS-CoV-2 or SARS-CoV2 or SARSCoV-2 or SARSCoV2 or 2019-novel CoV or Sars-coronavirus2 or novel CoV).mp. | 191954 |
| 11 | (or/5-8) and 9 and 10 | 4888 |
| 12 | (postcovid or postcorona* or ((post* or survivor*) adj3 (COVID or COVID-19 or COVID19 or coronavirus* or corona virus* or 2019-nCoV or 19nCoV or 2019nCoV or nCoV or n-CoV or SARS-CoV-2 or SARS-CoV2 or SARSCoV-2 or SARSCoV2 or 2019-novel CoV or Sars-coronavirus2 or novel CoV))).tw,kw. | 3563 |
| 13 | (outcome* or symptom* or morbidit* or manifestation* or issue* or effect? or difficult* or challeng* or problem* or complication* or disturbance* or consequence* or impair* or dysfunction* or disfunction* or function* or abnormalit* or dizziness or headache* or dyspnea* or fatigue or breath or breathing or lung or lungs or respiratory or tachycardi* or palpitation* or neuro* or concentration or concentrating or brain fog or cough* or ache* or pain* or taste* or tasting or smell* or olfact* or gustatory).tw,kw. | 19486315 |
| 14 | 12 and 13 | 2628 |
| 15 | ("coronavirus disease 2019"/ or "severe acute respiratory syndrome coronavirus 2"/ or Coronavirus infection/) and syndrome/ | 44 |
| 16 | 2 or 3 or 4 or 11 or 14 or 15 | 15681 |
| 17 | (Risk factor* or relative risk or odds ratio or between group* or Regression or multi-variate or multivaria* or covariate or univariate or co-variate or matching or ANOVA or Analysis of variance or ANCOVA or Correlation or Covariance or Principal Component Analysis or cohort* or follow-up or prognos* or predict*).mp. | 8659147 |
| 18 | cohort analysis/ | 737503 |
| 19 | (cohort* or longitudinal* or ((prospective* or retrospective*) adj3 (study or studies or research* or participant* or trial* or enrol* or data))).tw,kw. | 2441812 |
| 20 | ("Associated with" or "Association of" or "impact of" or "Correlated with" or "Impact* on" or characteristics or characterise or features).ti. | 1291059 |
| 21 | (clinical data or (clinical adj5 (characteristics or features or manifestations))).tw,kw. | 649142 |
| 22 | 17 or 18 or 19 or 20 or 21 | 10112851 |
| 23 | 1 and 16 and 22 | 7464 |
| 24 | (exp animal/ or exp animal model/ or exp animal experiment/) not (human* or person*).hw. | 5081976 |
| 25 | (animal* or amphib* or amphipod* or ape or apes or avian or bat or bats or beagle* or bird or birds or bivalve* or boar? or bovine* or canin* or cat or cats or catfish* or cattle or chicken* or chimpanzee* or cow or cows or crustacean* or dog or dogs or equine or ewe? or fish or fishes or feline* or frog or frogs or fruit fly or fruit flies or gamefish* or gastropod* or gerbil* or goat* or guineapig* or hamster? or hare or hares or horse* or macaque* or litter? or lizard* or mammal* or marmoset* or mice or mollusk* or monkey* or mouse or murine or ostrich* or ovine or pig or piglet* or pigs or porcine or pork* or primate* or rabbit* or rat or rats or reptil* or rodent? or sheep or sprague-dawley or songbird* or squirrel* or sow or sows or swine* or toad? or veterinar* or worm* or zebrafish* or zebra fish*).ti. | 2938483 |
| 26 | (conference abstract or conference paper or "conference review" or editorial).pt. | 5621765 |
| 27 | case report/ or (case-stud* or case-report*).jw. or (case adj3 (report* or stud*)).ab. or (case* and report*).ti. | 3026467 |
| 28 | (MIS-C or multisystem-inflammatory syndrome or lockdown* or lock* down* or restrictions or shelter-in-place or stay-at-home or school clos* or personal protective equipment or ppe or hygiene or close contact* or self isolat* or social distanc* or mask or masks).ti. | 39595 |
| 29 | 23 not (or/24-28) | 5079 |
| 30 | limit 29 to yr="2021 -Current" | 3159 |
| 31 | (202101* or 202102* or 202103* or 202104* or 202105* or 202106* or 202107* or 202108* or 202109* or 20211* or 2022*).dc,dd,dp. | 1568590 |
| 32 | 29 and 31 | 3676 |
| 33 | 30 or 32 | 3713 |

**Search Strategy for Key Question 2**

Ovid Medline(R) ALL

| **#** | **Searches** | **Results** |
| --- | --- | --- |
| 1 | (long adj (COVID or COVID-19 or COVID19 or coronavirus* or corona virus* or 2019-nCoV or 19nCoV or 2019nCoV or nCoV or n-CoV or "CoV 2" or CoV2 or SARS-CoV-2 or SARS-CoV2 or SARSCoV-2 or SARSCoV2 or SARS2 or SARS-2 or severe acute respiratory syndrome coronavirus 2 or 2019-novel CoV or Sars-coronavirus2 or Sars-coronavirus-2 or SARS-like coronavirus* or novel coronavirus* or novel corona virus* or novel CoV or OC43 or NL63 or 229E or HKU1 or HCoV* or Sarscoronavirus*)).tw,kf. | 335 |
| 2 | ((longterm or long-term) adj (COVID or COVID-19 or COVID19 or coronavirus* or corona virus* or 2019-nCoV or 19nCoV or 2019nCoV or nCoV or n-CoV or "CoV 2" or CoV2 or SARS-CoV-2 or SARSCoV2 or SARSCoV-2 or SARSCoV2 or SARS2 or SARS-2 or severe acute respiratory syndrome coronavirus 2 or 2019-novel CoV or Sars-coronavirus2 or Sars-coronavirus-2 or SARS-like coronavirus* or novel coronavirus* or novel corona virus* or novel CoV or OC43 or NL63 or 229E or HKU1 or HCoV* or Sars-coronavirus*)).tw,kf. | 33 |
| 3 | ((postacute or post-acute) adj (COVID or COVID-19 or COVID19 or coronavirus* or corona virus* or 2019-nCoV or 19nCoV or 2019nCoV or nCoV or n-CoV or "CoV 2" or CoV2 or SARS-CoV-2 or SARS-CoV2 or SARSCoV-2 or SARSCoV2 or SARS2 or SARS-2 or severe acute respiratory syndrome coronavirus 2 or 2019-novel CoV or Sars-coronavirus2 or Sars-coronavirus-2 or SARS-like coronavirus* or novel coronavirus* or novel corona virus* or novel CoV or OC43 or NL63 or 229E or HKU1 or HCoV* or Sars-coronavirus*)).tw,kf. | 86 |
| 4 | (chronic* adj2 (COVID or COVID-19 or COVID19 or coronavirus* or corona virus* or 2019-nCoV or 19nCoV or 2019nCoV or nCoV or n-CoV or "CoV 2" or CoV2 or SARS-CoV-2 or SARS-CoV2 or SARSCoV-2 or SARSCoV2 or SARS2 or SARS-2 or severe acute respiratory syndrome coronavirus 2 or 2019-novel CoV or Sars-coronavirus2 or Sars-coronavirus-2 or SARS-like coronavirus* or novel coronavirus* or novel corona virus* or novel CoV or OC43 or NL63 or 229E or HKU1 or HCoV* or Sarscoronavirus*)).tw,kf. | 105 |
| 5 | (("90" or ninety or "120" or "180" or hundred*) adj2 day?).tw,kf,tc. | 68372 |
| 6 | (("12" or twelve or "13" or thirteen or "14" or fourteen or "15" or "fifteen" or "16" or sixteen or "17" or seventeen or "18" or eighteen or "19" or nineteen or "20" or "twenty" or "21" or "22" or "23" or "24") adj3 (week? or wk or wks)).tw,kf,tc. | 293500 |
| 7 | (("3" or three or "4" or four or "5" or five or "6" or six or "7" or seven or "8" or eight or "9" or nine or "10" or ten or "11" or eleven or "12" or twelve or "13" or thirteen or "14" or fourteen or "15" or "fifteen" or "16" or sixteen or "17" or seventeen or "18" or eighteen or "19" or nineteen or "20" or "twenty" or "21" or "22" or "23" or "24") adj3 (month? or mo or mos)).tw,kf,tc. | 1337459 |
| 8 | (("1" or one or "2" or two or half or multi* or full) adj3 (year? or yr or yrs)).tw,kf,tc. | 807366 |
| 9 | (("more than" or longer or "at least" or "after" or follow* or upward* or later* or prospectiv* or retrospectiv* or review* or longitud* or post or beyond) adj5 (day? or week? or wk or wks or month? or mo or mos or year? or yr or yrs)).tw,kf,tc. | 2512485 |
| 10 | (COVID or COVID-19 or COVID19 or coronavirus* or corona virus* or 2019-nCoV or 19nCoV or 2019nCoV or nCoV or n-CoV or SARS-CoV-2 or SARS-CoV2 or SARSCoV-2 or SARSCoV2 or 2019-novel CoV or Sars-coronavirus2 or novel CoV).tw,kf,tc. | 169269 |
| 11 | (5 or 6 or 7 or 8) and 9 and 10 | 3324 |
| 12 | COVID-19/ and Syndrome/ | 113 |
| 13 | SARS-CoV-2/ and Syndrome/ | 88 |
| 14 | 1 or 2 or 3 or 4 or 11 or 12 or 13 [LONG COVID - PT 1] | 3859 |
| 15 | COVID-19/ | 94576 |
| 16 | SARS-CoV-2/ | 73509 |
| 17 | Coronavirus/ | 4747 |
| 18 | Betacoronavirus/ | 33232 |
| 19 | Coronavirus Infections/ | 45007 |
| 20 | (COVID-19 or COVID19).tw,kf. | 141353 |
| 21 | ((coronavirus* or corona virus*) and (hubei or wuhan or beijing or shanghai)).tw,kf. | 5095 |
| 22 | (wuhan adj5 virus*).tw,kf. | 242 |
| 23 | (2019-nCoV or 19nCoV or 2019nCoV).tw,kf. | 1772 |
| 24 | (nCoV or n-CoV or "CoV 2" or CoV2).tw,kf. | 54353 |
| 25 | (SARS-CoV-2 or SARS-CoV2 or SARSCoV-2 or SARSCoV2 or SARS2 or SARS-2 or severe acute respiratory syndrome coronavirus 2).tw,kf. | 55171 |
| 26 | (2019-novel CoV or Sars-coronavirus2 or Sars-coronavirus-2 or SARS-like coronavirus* or ((novel or new or nouveau) adj2 (CoV or nCoV or covid or coronavirus* or corona virus or Pandemi*2)) or (coronavirus* and pneumonia)).tw,kf. | 19049 |
| 27 | (novel coronavirus* or novel corona virus* or novel CoV).tw,kf. | 9787 |
| 28 | ((coronavirus* or corona virus*) adj2 "2019").tw,kf. | 32748 |
| 29 | ((coronavirus* or corona virus*) adj2 "19").tw,kf. | 5292 |
| 30 | (coronavirus 2 or corona virus 2).tw,kf. | 17314 |
| 31 | (OC43 or NL63 or 229E or HKU1 or HCoV* or Sars-coronavirus*).tw,kf. | 3733 |
| 32 | COVID-19.rx,px,ox. or severe acute respiratory syndrome coronavirus 2.os. | 4656 |
| 33 | (coronavirus* or corona virus*).ti. | 22486 |
| 34 | or/15-33 [COVID-19] | 172141 |
| 35 | ((post or after or following) adj (COVID or COVID-19 or COVID19 or coronavirus* or corona virus* or 2019-nCoV or 19nCoV or 2019nCoV or nCoV or n-CoV or "CoV 2" or CoV2 or SARS-CoV-2 or SARS-CoV2 or SARSCoV-2 or SARSCoV2 or SARS2 or SARS-2 or severe acute respiratory syndrome coronavirus 2 or 2019-novel CoV or Sars-coronavirus2 or Sars-coronavirus-2 or SARS-like coronavirus* or novel coronavirus* or novel corona virus* or novel CoV or OC43 or NL63 or 229E or HKU1 or HCoV* or Sarscoronavirus*)).tw,kf,tc. | 3488 |
| 36 | (abnormalities or ache* or brain fog or breath* or challeng* or "co morbid*" or comorbid* or complications or concentrat* or condition* or consequences or convalescen* or cough or difficulties or disease* or disorder* or disturbances or dizziness or dysfunction or dyspnea or effects or fatigue or function* or gustatory or headache* or illness* or impairments or issues or lung or manifestation* or morbidit* or "multi morbid*" or multimorbid* or neuro* or olfaction or olfactory or outcome? or pain* or palpitation* or problem? or prognos* or recuperat* or respiratory or risk* or sickness* or sign or signs or smell* or surviv* or symptom* or syndrome* or tachycardia or taste?).tw,kf,tc. | 18170163 |
| 37 | 35 and 36 | 2626 |
| 38 | ((chronic* or continuous* or continual* or continuing* or delay* or endur* or extend* or fluctuat* or gradual* or lasting* or legacy* or lengthy* or linger* or long* or "medium* term*" or mediumterm* or multisystem* or "multi system*" or ongoing* or permanent* or persist* or prolong* or protract* or relaps* or remission* or remit* or residual* or slow* or subacute* or "sub acute*") adj3 recover*).tw,kf. | 33552 |
| 39 | ((after discharg* or following discharg* or postacute* or "post acute*" or postdischarg* or "post discharge" or "post discharging" or posthospital* or post-hospital* or postinfect* or "post infection" or "post infective*" or postviral* or "post viral*" or postvirus* or "post virus*" or postcritical or post-critical or postintensive or post-intensive or post-ICU) adj3 recover*).tw,kf. | 552 |
| 40 | ((chronic* or continuous* or continual* or continuing* or delay* or endur* or extend* or fluctuat* or gradual* or lasting* or legacy* or lengthy* or linger* or long* or "medium* term*" or mediumterm* or multisystem* or "multi system*" or ongoing or permanent* or persist* or prolong* or protract* or relaps* or remission* or remit* or residual* or slow* or subacute* or "sub acute*") adj5 (complication? or consequence? or convalescen* or disabilit* or feature* or illness* or prognos* or sequela* or sign or signs or suffering? or symptom* or recuperat*)).tw,kf. | 327882 |
| 41 | ((after discharg* or following discharg* or postacute* or "post acute*" or postdischarg* or "post discharge" or "post discharging" or posthospital* or post-hospital* or postinfect* or "post infection" or "post infective*" or postviral* or "post viral*" or postvirus* or "post virus*" or postcritical or post-critical or postintensive or post-intensive or post-ICU) adj5 (complication? or consequence? or convalescen* or disabilit* or feature* or illness* or prognos* or sequela* or sign or signs or suffering? or symptom* or recuperat*)).tw,kf. | 2836 |
| 42 | (nonrecover* or "non recover*" or "not recover*").tw,kf. | 7274 |
| 43 | ("long* haul*" or longhaul* or "long* tail*" or longtail* or longduration* or "long duration*" or longlast* or "long last*" or longstanding* or "long standing*" or "medium* term*" or mediumterm*).tw,kf. | 116227 |
| 44 | 37 or 38 or 39 or 40 or 41 or 42 or 43 [LONG-TERM ILLNESS, PROTRACTED RECOVERY, ETC.] | 476312 |
| 45 | 34 and 44 [LONG COVID - PT 2] | 6613 |
| 46 | Long Term Adverse Effects/ | 691 |
| 47 | "Recovery of Function"/ | 56171 |
| 48 | Convalescence/ | 3777 |
| 49 | or/46-48 [POST-COVID RECOVERY PERIOD] | 60460 |
| 50 | 34 and 49 [LONG COVID - PT 3] | 381 |
| 51 | 14 or 45 or 50 [LONG COVID - LONG COVID HEDGE: DECARY ET AL, 2021] | 9834 |
| 52 | *ambulatory care/ or *patient care management/ or *preventive health services/ or *diagnostic services/ or *early medical intervention/ or exp *health education/ or *primary prevention/ or *secondary prevention/ or *tertiary prevention/ or *health services/ or *adolescent health services/ or exp *community health services/ or *emergency medical services/ or hotlines/ or *health services for persons with disabilities/ or *health services for the aged/ or *health services for transgender persons/ or *personal health services/ or exp *pharmaceutical services/ or exp *rural health services/ or *suburban health services/ or *urban health services/ or *women's health services/ or *therapeutics/ or *drug therapy/ or *patient care/ or *public health practice/ | 532518 |
| 53 | (intervention* or service* or health care* or healthcare* or health system* or program* or strategy* or strategies or framework* or policy or policies or guideline* or treat* or therap* or prevent* or rehab* or pharmaceutic* or medication* or drug* or hotline* or clinic or clinics or health centre* or health center* or hospital* or practice*).ti,kf. | 4815473 |
| 54 | 52 or 53 [BROAD LEVEL INTERVENTIONS] | 5093044 |
| 55 | exp *aftercare/ or *outpatients/ | 129506 |
| 56 | (aftercare or after care or follow-up care or referral?).tw,kf. | 132933 |
| 57 | 55 or 56 [AFTER CARE] | 259328 |
| 58 | *models, organizational/ or exp *delivery of health care/ | 693554 |
| 59 | ((healthcare or health care or clinical* or public health or community) adj4 (approach* or model? or service? or deliver* or system* or distribut* or framework? or guideline?)).tw,kf. | 406896 |
| 60 | 58 or 59 [HEALTHCARE MODELS] | 1030936 |
| 61 | exp Vaccines/ | 242192 |
| 62 | exp Immunization/ | 185328 |
| 63 | post-exposure prophylaxis/ | 1439 |
| 64 | (vaccine* or vaccinat* or revaccin* or immunis* or immuniz* or inoculat*).tw,kf. | 526689 |
| 65 | (prophyla* or prevent* or protect* or postexpos* or (expos* adj3 (post or after or document* or suspect* or positiv* or confirm*))).tw,kf. | 2497381 |
| 66 | (postcontact* or (contact* adj3 (post or after or document* or suspect* or positiv* or confirm*))).tw,kf. | 13605 |
| 67 | or/61-66 [VACCINATION] | 2962965 |
| 68 | exp vitamins/ | 332864 |
| 69 | exp vitamin b 6/ | 16320 |
| 70 | exp vitamin b 12/ | 22609 |
| 71 | exp riboflavin/ | 14732 |
| 72 | exp niacinamide/ | 15462 |
| 73 | exp thiamine/ | 12027 |
| 74 | exp folic acid/ | 39240 |
| 75 | exp ascorbic acid/ | 43392 |
| 76 | exp Dietary Supplements/ | 85357 |
| 77 | vitamin?.tw,kf. | 232926 |
| 78 | riboflavin.tw,kf. | 10875 |
| 79 | (niacinamide or enduramide or nicobion or nicotinamide or papulex).tw,kf. | 23419 |
| 80 | (thiamine or aneurin or thiamin).tw,kf. | 13731 |
| 81 | (folic acid or folacin or folate or folvite or pteroylglutamic acid).tw,kf. | 44294 |
| 82 | (ascorbic acid or ferrous ascorbate or hybrin or "l-ascorbic acid" or magnesium ascorbate or magnesium ascorbicum or "magnesium di-l-ascorbate" or magnorbin or sodium ascorbate).tw,kf. | 35550 |
| 83 | ((diet* or nutrition* or herbal* or food?) adj3 supplement*).tw,kf. | 66286 |
| 84 | or/68-83 [NUTRTIONAL SUPPLEMENTS] | 579241 |
| 85 | exp anti-inflammatory agents/ | 529663 |
| 86 | (((antiinflammator* or anti inflammator*) adj2 (drug? or pharmaceutical? or agent? or substance? or medicin* or prescription?)) or NSAID or NSAIDs).tw,kf. | 68785 |
| 87 | exp adrenal cortex hormones/ | 406349 |
| 88 | (adrenal cortex hormone? or corticoid? or cortical steroid? or cortico steroid? or corticosteroid? or dermocorticosteroid?).tw,kf. | 124174 |
| 89 | (dexamethasone? or decaject? or decameth? or decaspray? of dexasone? or dexpak? or hexadecadrol? or hexadrol? or maxidex? or methylfluorprednisolone? or millicorten? or oradexon?).tw,kf. | 59981 |
| 90 | (prednisone or cortan or cortancyl or cutason or dacortin or decortin or decortisyl or dehydrocortisone or deltasone or encorton or encortone or enkortolon or kortancyl or liquid pred or meticorten or orasone or panafcort or panasol or predni tablinen or prednidib or predniment or prednison acsis or prednison galen or prednison hexal or pronisone or rectodelt or sone or sterapred or ultracorten or winpred or delta cortisone).tw,kf. | 30400 |
| 91 | (methylprednisolone or methylprednisolone or medrol or metipred or urbason).tw,kf. | 17506 |
| 92 | (hydrocortisone or "acticort" or "aeroseb hc" or "ala-cort" or "ala-scalp" or "alfacort" or "algicortis" or "alkindi" or "alpha derm" or "alphaderm" or "anucort-hc" or "anumed-hc" or "anutone-hc" or "aquanil hc" or "balneol-hc" or "barseb hc" or "beta-hc" or "biacort" or "cetacort" or "cobadex" or "colocort" or "compound f" or "cordicare lotion" or "coripen" or "cort dome" or "cortef" or "cortef cream" or "cortenema" or "cortibel" or "corticorenol" or "cortifan" or "cortiphate" or "cortisol" or "cortisole" or "cortispray" or "cortoderm" or "cortril" or "cotacort" or "covocort" or "cremicort-h" or "cutaderm" or "dermacrin hc lotion" or "dermaid" or "derm-aid cream" or "dermaid soft cream" or "dermocare" or "dermocortal" or "dermolate" or "dioderm" or "eczacort" or "ef cortelan" or "efcortelan" or "egocort" or "egocort cream" or "eksalb" or "eldecort" or "emo-cort" or "epicort" or "ficortril" or "filocot" or "flexicort" or "glycort" or "gly-cort" or "hc no. 1" or "hc no. 4" or "h-cort" or "hebcort" or "hebcort v" or "hemorrhoidal hc" or "hemril-30" or "hemril-hc uniserts" or "hi-cor" or "hidrotisona" or "hycor" or "hycort" or "hydracort" or "hydrasson" or "hydro ricortex" or "hydrocort" or "hydrocorticosteroid" or "hydrocortisate" or "hydrocortison" or "hydrocortisonum" or "hydrocortisyl" or "hydrocortone" or "hydrogalen" or "hydrokort" or "hydrokortison" or "hydro-rx" or "hydrotopic" or "hysone" or "hytisone" or "hytone" or "hytone lotion" or "incortin h" or "instacort 10" or "kyypakkaus" or "lacticare hc" or "lemnis fatty cream hc" or "lenirit" or "medihaler cort" or "medihaler duo" or "medrocil" or "mildison" or "mitocortyl demangeaisons" or "munitren" or "nogenic hc" or "novohydrocort" or "nsc 10483" or "nsc 741" or "nsc10483" or "nutracort" or "optef" or "otosone f" or "penecort" or "plenadren" or "prepcort" or "prevex hc" or "pro cort" or "procort" or "proctocort" or "proctosert hc" or "proctosol-hc" or "proctosone" or "proctozone hc" or "procutan" or "rectasol-hc" or "rectocort" or "rederm" or "sanatison" or "scalp-aid" or "schericur" or "schericur 0.25%" or "scherosone f" or "sistral hydrocort" or "skincalm" or "stie-cort" or "substance m" or "synacort" or "texacort" or "triburon-hc" or "unicort" or "vasocort").tw,kf. | 81737 |
| 93 | nitric oxide/ | 91752 |
| 94 | (nitric oxide or endogenous nitrate vasodilator or mononitrogen monoxide or nitrogen monoxide or genosyl or inomax or noxivent).tw,kf. | 154983 |
| 95 | exp prednisolone/ | 52218 |
| 96 | (prednisolone or "adelcort" or "antisolon" or "antisolone" or "aprednislon" or "aprednislone" or "benisolon" or "benisolone" or "berisolon" or "berisolone" or "caberdelta" or "capsoid" or "codelcortone" or "co-hydeltra" or "compresolon" or "cortadeltona" or "cortadeltone" or "cortalone" or "cortelinter" or "cortisolone" or "cotolone" or "dacortin" or "dacortin h" or "dacrotin" or "decaprednil" or "decortin h" or "decortril" or "dehydro cortex" or "dehydro hydrocortison" or "dehydro hydrocortisone" or "dehydrocortex" or "dehydrocortisol" or "dehydrocortisole" or "dehydrohydrocortison" or "dehydrohydrocortisone" or "delcortol" or "delta 1 hydrocortisone" or "delta cortef" or "delta cortril" or "delta ef cortelan" or "delta f" or "delta hycortol" or "delta hydrocortison" or "delta hydrocortisone" or "delta ophticor" or "delta stab" or "delta1 dehydrocortisol" or "delta1 dehydrohydrocortisone" or "delta1 hydrocortisone" or "deltacortef" or "deltacortenolo" or "deltacortil" or "deltacortoil" or "deltacortril" or "deltaderm" or "deltaglycortril" or "deltahycortol" or "deltahydrocortison" or "deltahydrocortisone" or "deltaophticor" or "deltasolone" or "deltastab" or "deltidrosol" or "deltisilone" or "deltisolon" or "deltisolone" or "deltolasson" or "deltolassone" or "deltosona" or "deltosone" or "depo-predate" or "dermosolon" or "dhasolone" or "di adreson f" or "di adresone f" or "diadreson f" or "diadresone f" or "dicortol" or "domucortone" or "encortelon" or "encortelone" or "encortolon" or "equisolon" or "fernisolone-p" or "glistelone" or "hefasolon" or "hostacortin h" or "hostacortin h vet" or "hydeltra" or "hydeltrone" or "hydrelta" or "hydrocortancyl" or "hydrocortidelt" or "hydrodeltalone" or "hydrodeltisone" or "hydroretrocortin" or "hydroretrocortine" or "inflanefran" or "insolone" or "keteocort h" or "key-pred" or "lenisolone" or "leocortol" or "liquipred" or "lygal kopftinktur n" or "mediasolone" or "meprisolon" or "meprisolone" or "metacortalon" or "metacortalone" or "metacortandralon" or "metacortandralone" or "metacortelone" or "meti derm" or "meticortelone" or "metiderm" or "morlone" or "mydrapred" or "neo delta" or "nisolon" or "nisolone" or "nsc 9120" or "nsc9120" or "opredsone" or "panafcortelone" or "panafcortolone" or "panafort" or "paracortol" or "phlogex" or "pre cortisyl" or "preconin" or "precortalon" or "precortancyl" or "precortisyl" or "predacort 50" or "predaject-50" or "predalone 50" or "predartrina" or "predartrine" or "predate-50" or "predeltilone" or "predisole" or "predisyr" or "pred-ject-50" or "predne dome" or "prednecort" or "prednedome" or "prednelan" or "predni coelin" or "predni h tablinen" or "prednicoelin" or "prednicort" or "prednicortelone" or "prednifor drops" or "predni-helvacort" or "predniment" or "predniretard" or "prednis" or "prednisil" or "prednisolon" or "prednisolona" or "prednisolone alcohol" or "prednisolone h" or "prednisolone oleosae sr 82" or "prednivet" or "prednorsolon" or "prednorsolone" or "predonine" or "predorgasolona" or "predorgasolone" or "prelon" or "prelone" or "prenilone" or "prenin" or "prenolone" or "preventan" or "prezolon" or "rubycort" or "scherisolon" or "scherisolona" or "serilone" or "solondo" or "solone" or "solupren" or "soluprene" or "spiricort" or "spolotane" or "sterane" or "sterolone" or "supercortisol" or "supercortizol" or "taracortelone" or "walesolone" or "wysolone").tw,kf. | 30072 |
| 97 | exp aspirin/ | 46245 |
| 98 | (aspirin or acetylsalicylic acid or "8-hour bayer" or "acenterine" or "acesal" or "acetan" or "acetard" or "aceticil" or "aceticyl" or "acetilum" or "acetonyl" or "acetophen" or "acetosal" or "acetosalicylic acid" or "acetosalin" or "acetosalum" or "acetyl salicylate" or "acetyl salicylic acid" or "acetylic salicylic acid" or "acetylin" or "acetylo" or "acetylo salicylic acid" or "acetylon" or "acetylosalicylic acid" or "acetylsal" or "acetylsalicyclic acid" or "acetylsalicyl" or "acetylsalicylate" or "acetylsalicylate strontium" or "acetylsalicylic acid plus glycine" or "acetylsalicylic acid sodium salt" or "acetylsalicylic acid strontium salt" or "acetylsalycic acid" or "acetylsalycylic acid" or "acetysal" or "acidulatum" or "acidum acetyl salicylicum" or "acidum acetylosalicylicum" or "acidum acetylsalicylicum" or "actorin" or "acylpyrin" or "acylpyrine" or "acytosal" or "adiro" or "alabukun" or "alasil" or "albyl e" or "albyl minor" or "alka seltzer" or "alkaspirin" or "anasprin" or "andol" or "anopyrin" or "ansin" or "anthrom" or "aptor" or "arthralgyl" or "arthritis strength bufferin" or "asacard" or "asaetta" or "asaflow" or "asaphen" or "asapor" or "asatard" or "asawin" or "aspec" or "aspent" or "aspergum" or "aspex" or "aspilets" or "aspirem" or "aspirgran" or "aspiricor" or "aspirina" or "aspirine" or "aspirinine" or "aspirisucre" or "aspisol" or "aspo cid" or "aspro" or "aspro cardio" or "aspro clear" or "asproflash" or "asrina" or "asrivo" or "asta" or "asteric" or "asteric acid" or "astrix" or "bamyl" or "bayaspirina" or "bebesan" or "biprin" or "bokey" or "boxazin" or "breoprin" or "bufferin" or "cafenol" or "cardioasa" or "cardioasae" or "cardioaspirina" or "cartia" or "caspirin" or "catalgine" or "catalgix" or "cemerit" or "cemirit" or "claradin" or "claragine" or "colfarit" or "comoprin" or "contrheuma" or "contrheuma retard" or "darosal" or "depot aspirin" or "dispirin" or "dolean" or "durlaza" or "dusil" or "easprin" or "ecasil" or "ecosprin" or "ecotrin" or "egalgic" or "emocin" or "empirin" or "encaprin" or "encine em" or "endosprin" or "entaprin" or "entericin" or "enteroprin" or "enterosarine" or "enterospirine" or "entrophen" or "eskotrin" or "euthermine" or "extren" or "flamasacard" or "genasprin" or "globentyl" or "godamed" or "gotosan" or "helicon" or "herz ass" or "hjertemagnyl" or "idotyl" or "infatabs a" or "istopirin" or "istopyrine" or "ivepirine" or "juvepirine" or "keypo" or "kilios" or "kinderaspirin" or "magnecyl brus" or "magnyl dak" or "mcn r 358" or "measurin" or "mejoral" or "melabon" or "micristin" or "micropyrin" or "migrasaa" or "mikristin" or "miniasal" or "mycristin" or "naspro" or "novasen" or "nu seal" or "nuseals" or "nu-seals" or "nu-seals asa" or "ortho acetoxybenzoate" or "ortho acetoxybenzoic acid" or "ortho acetyloxybenzoate" or "ortho acetyloxybenzoic acid" or "ostoprin" or "pancemol" or "para acetylsalicylic acid" or "paracin" or "paynocil" or "pengo" or "platet 300 cleartab" or "plewin" or "polopiryna" or "premaspin" or "primaspan" or "proprin" or "pyronoval" or "reumyl" or "rhodine" or "rhonal" or "ronal" or "salacetin" or "salacetogen" or "saletin" or "salisalido" or "salospir" or "sargepirine" or "sedergine" or "sedergine forte" or "sodium acetylsalicylate" or "sodium bicarbonate acetyl salicylate" or "sodium bicarbonate acetylsalicylate" or "soldral" or "solpyron" or "solucetyl" or "solupsa" or "spren" or "super tru" or "tapal" or "temagin" or "tevapirin" or "th 2152" or "thrombo-aspilets" or "toldex retard" or "treupahlin" or "treuphalin" or "tromalyt" or "tromcor" or "turivital" or "vazalore" or "verin" or "vitalink" or "xaxa" or "zorprin").tw,kf. | 61195 |
| 99 | (adalimumab or "abp 501" or "abp501" or "abrilada" or "abt d2e7" or "abtd2e7" or "adaly" or "amgevita" or "amjevita" or "amsparity" or "avt 02" or "avt02" or "bat 1406" or "bat1406" or "bax 2923" or "bax 923" or "bax2923" or "bax923" or "bi 695501" or "bi695501" or "chs 1420" or "chs1420" or "cinnora" or "ct p17" or "ctp17" or "cyltezo" or "da 3113" or "da3113" or "dmb 3113" or "dmb3113" or "exemptia" or "fkb 327" or "fkb327" or "fyzoclad" or "gp 2017" or "gp2017" or "hadlima" or "halimatoz" or "hefiya" or "hlx 03" or "hlx03" or "hulio" or "humira" or "hyrimoz" or "ibi 303" or "ibi303" or "idacio" or "imraldi" or "kromeya" or "lu 200134" or "lu200134" or "m 923" or "m923" or "mabura" or "monoclonal antibody d2e7" or "msb 11022" or "msb11022" or "ons 3010" or "ons3010" or "pf 06410293" or "pf 6410293" or "pf06410293" or "pf6410293" or "raheara" or "sb 5" or "sb5" or "solymbic" or "trudexa" or "zrc 3197" or "zrc3197").tw,kf. | 8028 |
| 100 | (celecoxib or "aclarex" or "artilog" or "artroxil" or "caditar" or "celcox" or "celebra" or "celebreks" or "celebrex" or "celecox" or "celib" or "celora" or "coxel" or "coxid" or "dilox" or "eliflam" or "elyxyb" or "lexfin" or "onsenal" or "sc 58635" or "sc58635" or "solexa" or "ym 177" or "ym177" or "zycel").tw,kf. | 7555 |
| 101 | (diclofenac or "abdiflam" or "abitren" or "acuflam" or "akis" or "algipatch" or "algistick" or "algopain eze" or "algoplast" or "allvoran" or "almiral" or "alonpin" or "antalcalm" or "apo-diclofenac ec" or "arcanafenac" or "arthrifen" or "artren" or "artrenac" or "artrites" or "assaren" or "athrofen" or "ba 47210" or "ba47210" or "berafen gel" or "berifen" or "betaren" or "bolabomin" or "calozan" or "catanac" or "catas" or "cencenag" or "clo-far" or "clofec" or "clofen" or "clonac" or "clonaren" or "clonodifen" or "cordralan" or "curinflam" or "ddl plaster" or "declophen" or "decrol" or "deflamat" or "deflam-k" or "delphinac" or "denaclof" or "depain" or "diceus" or "dicipan" or "diclac" or "diclax" or "diclo" or "diclobasan" or "diclobene" or "diclod" or "diclodent" or "diclodoc" or "diclodolor" or "diclofen" or "diclofen cremogel" or "diclofenac rekur" or "diclofenac resin" or "diclofenac resinate" or "diclofenac sodium" or "dicloflam" or "diclohexal" or "dicloin" or "diclomax" or "diclomol" or "diclon" or "diclopax" or "diclophenac sodium" or "diclopuren" or "dicloral" or "dicloran gel" or "diclorecip" or "dicloren" or "dicloreum" or "diclosan sr" or "diclosian" or "diclotec" or "diclowal" or "dicsnal" or "difen" or "difena" or "difenac" or "difenol gel" or "difnal k" or "dioxaflex" or "dioxaflex retard" or "divoltar" or "dixol" or "doflastad" or "doflex" or "dolaren" or "dolflam-retard" or "dolo voltaren" or "doloflam" or "dolotren" or "doragon" or "dosanac" or "duravolten" or "dycon sr" or "dyloject" or "ecofenac" or "econac" or "effekton" or "eflagen" or "epifenac" or "eslofen" or "evadol" or "evinopon" or "feloran" or "fenac" or "fenadium" or "fenaspec" or "flameril" or "flexagen" or "flogofenac" or "flogosin d" or "flogozan" or "fortfen sr" or "freejex" or "gp 45840" or "grofenac" or "hizemin" or "imflac" or "inac gel" or "indicam" or "inflamac" or "inflanac" or "isv 205" or "isv205" or "jonac gel" or "kadiflam" or "kinespir" or "klofen l" or "klotaren" or "kriplex" or "lesflam" or "leviogel" or "lifenac" or "lofenac" or "lotirac" or "magluphen" or "merflam" or "modifenac" or "monoflam" or "motifene" or "naboal" or "nac gel" or "naclof" or "nacoflar" or "nadifen" or "novapirina" or "novo-difenac" or "novolten" or "ofenac" or "olfen" or "optanac" or "orthophen" or "osteoflam" or "painstop" or "panamor" or "pennsaid" or "profenac" or "relaxyl gel" or "remethan" or "renvol emulgel" or "rewodina" or "rheufenac" or "rheumafen" or "rheumatac" or "rheumatac retard" or "rhewlin" or "rhewlin sr" or "rhumalgan" or "rolactin" or "sailib" or "savismin" or "sefnac" or "slofenac" or "sodium diclofenac" or "solaraze" or "sophenoderm" or "soproxen" or "spraymik" or "sr 318t" or "staren" or "sting gel" or "tabiflex" or "tds 943" or "tds943" or "tigen plaster" or "toraren" or "traulen" or "tsudohmin" or "uniclonax" or "uniren" or "valentac" or "vartelon" or "veral" or "voldal" or "voldic" or "volero" or "volfenac" or "volna-k" or "volsaid" or "volta" or "voltadex emulgel" or "voltadvance" or "voltalen" or "voltalen emulgel" or "voltalgan" or "voltaren" or "voltarene" or "voltarenspe" or "voltarol" or "voltine" or "voltral" or "voltrix" or "voren emulgel" or "votalen" or "voveran" or "vurdon" or "wergyl" or "xenid" or "yuren" or "zolterol" or "zorvolex").tw,kf. | 13885 |
| 102 | dimethyl sulfoxide/ | 15114 |
| 103 | (dimethyl sulfoxide or "damul" or "demasorb" or "demavet" or "demeso" or "demexide" or "dimethyl sulphoxide" or "dimethylsulfoxide" or "dimethylsulphoxide" or "dimexide" or "dms 70" or "dms 90" or "dms70" or "dms90" or "dmso" or "dolicur" or "domoso" or "dromisol" or "gamasal 90" or "hyadur" or "infiltrina" or "methyl sulfoxide" or "methylsulfoxide" or "nsc 763" or "nsc763" or "rimso 100" or "rimso 50" or "somipront" or "sq 9453" or "sq9453" or "syntexan").tw,kf. | 31512 |
| 104 | (indomethacin or " algiflam" or "algometacin" or "amuno" or "antalgin dialicels" or "apo-indomethacin" or "areumatin" or "argilex" or "arthrexin" or "articulen" or "artracin" or "artrilona s" or "artrinovo" or "artrocid" or "asimet" or "benocid" or "betacin" or "bonidon" or "boutycin" or "catlep" or "chrono indocid" or "chronoindocid" or "confortid" or "docin" or "dolazal" or "dolazol" or "dolcidium" or "dometin" or "durametacin" or "elmego spray" or "elmetacin" or "endometacin" or "flamaret" or "flexin continus" or "grindocin" or "helvecin" or "idicin" or "im-75" or "imbrilon" or "imet" or "inacid" or "indacin" or "indalgin" or "inderapollon" or "indicin" or "indo phlogont" or "indocap" or "indocid" or "indocin" or "indocolir" or "indocollyre" or "indogesic" or "indolag" or "indolar sr" or "indolemmon" or "indo-lemmon" or "indomecin" or "indomed" or "indomee" or "indomelan" or "indomelol" or "indomet retard" or "indometacin sodium" or "indometacine" or "indomethacin" or "indomethacine" or "indomethacinum" or "indomethegan" or "indometicina mckesson" or "indometin" or "indometin depot" or "indomexum" or "indomin" or "indono" or "indoptic" or "indoptol" or "indorektal" or "indorem" or "indos" or "indosan" or "indosima" or "indosmos" or "indo-tablinen" or "indotard" or "indovis" or "indoxen" or "indoy" or "indren" or "indrenin" or "indylon" or "inflazon" or "inmetsin" or "inteban" or "lauzit" or "luiflex" or "lyo indometacin trihydrate" or "malival" or "mcn r 1166" or "mcn r1166" or "metacen" or "methacin" or "methindol" or "methindole" or "methocaps" or "metindol" or "mezolin" or "miometacen" or "mk 615" or "mk615" or "mobilan" or "novomethacin" or "osmogit" or "osmosin" or "reumacid" or "reusin" or "rheumacid" or "rheumacin" or "salinac" or "servimeta" or "sidocin" or "tannex" or "taye" or "tivorbex" or "vi-gel" or "vonum").tw,kf. | 36868 |
| 105 | methotrexate/ | 39092 |
| 106 | (methotrexate or "abitrexate" or "amethopterin" or "amethopterine" or "ametopterine" or "antifolan" or "biotrexate" or "canceren" or "cl 14377" or "cl14377" or "emtexate" or "emthexat" or "emthexate" or "emtrexate" or "enthexate" or "farmitrexat" or "farmitrexate" or "farmotrex" or "folex" or "ifamet" or "imeth" or "intradose mtx" or "jylamvo" or "lantarel" or "ledertrexate" or "maxtrex" or "metex" or "methoblastin" or "methohexate" or "methotrate" or "methotrexat" or "methotrexato" or "methoxtrexate" or "methrotrexate" or "methylaminopterin" or "methylaminopterine" or "meticil" or "metoject" or "metothrexate" or "metotrexat" or "metotrexate" or "metotrexin" or "metrex" or "mexate" or "mpi 5004" or "mpi5004" or "neotrexate" or "nordimet" or "novatrex" or "nsc 740" or "nsc740" or "otrexup" or "otrexup pfs" or "rasuvo" or "reditrex" or "reumatrex" or "rheumatrex" or "texate" or "texorate" or "trexall" or "xaken" or "xatmep" or "zexate").tw,kf. | 44190 |
| 107 | (naproxen or " acusprain" or "aflamax" or "aflaxen" or "agilex" or "agilxen" or "aleve" or "alpoxen" or "alpron" or "anaprox" or "anexopen" or "apo-naproxen" or "apranax" or "apraxin" or "apronax" or "artagen" or "artron" or "artroxen" or "axer alfa" or "babel" or "bipronyl" or "bonyl" or "congex" or "crysanal" or "dafloxen" or "daprox" or "daprox entero" or "deflamox" or "dextro naproxen" or "diferbest" or "diocodal" or "dolormin fuer frauen" or "dolormin fur frauen" or "dolormin gs" or "dysmenalgit" or "dysmenalgit n" or "ec naprosyn" or "equiproxen" or "femex" or "feminax ultra" or "flanax" or "flanax forte" or "floginax" or "flonax" or "floxene" or "fuxen" or "galpharm period pain relief" or "gibixen" or "headlon" or "iraxen" or "laraflex" or "lasonil antinfiammatorio e antireumatico" or "lefaine" or "leniartil" or "levo naproxen" or "licorax" or "methoxypropiocin" or "miranax" or "momendol" or "nafasol" or "naixan" or "napolon" or "naposin" or "napreben" or "naprelan" or "napren" or "naprium" or "naprius" or "naproflam" or "naprogesic" or "naprong" or "naprontag" or "naprorex" or "naprossene" or "naprostad" or "naprosyn" or "naprosyne" or "naprovite" or "naproxeno" or "naproxi 250" or "naproxi 500" or "naproxyn" or "naprozyne" or "naprux" or "napsyn" or "napxen" or "narma" or "narocin" or "naxen" or "naxopren" or "naxyn" or "neprossin" or "norswel" or "novonaprox" or "novo-naprox" or "novuran" or "nuprafem" or "nycopren" or "pactens" or "prexan" or "priaxen" or "prodilor" or "pronaxen" or "proxen" or "proxidol" or "rahsen" or "rs 3540" or "rs 3650" or "rs3540" or "rs3650" or "sanomed" or "saritilron" or "seladin" or "shiprosyn" or "sutolin" or "synaprosyn" or "synflex" or "tohexen" or "uniflam" or "u-ritis" or "velsay" or "veradol" or "vinsen" or "wintrex" or "xenar" or "xenobid").tw,kf. | 6975 |
| 108 | or/85-107 [ANTI-INFLAMMATORY AGENTS] | 1073830 |
| 109 | exp antiviral agents/ | 373204 |
| 110 | ((antivir* or anti vir*) adj2 (drug? or pharmaceutical? or agent? or substance? or medicin* or prescription?)).tw,kf. | 23250 |
| 111 | remdesivir.tw,kf. | 1604 |
| 112 | (tocilizumab or actemra or atlizumab or lusinex or "r 1569" or r1569 or roactemra).tw,kf. | 4475 |
| 113 | (baricitinib or "incb 028050" or "incb 28050" or incb028050 or incb28050 or "ly 3009104" or ly300910 or olumiant).tw,kf. | 543 |
| 114 | ((anticoagulant* or anti coagulant* or antithrombo* or anti-thrombo*) adj2 (drug? or pharmaceutical? or agent? or substance? or medicin* or prescription?)).tw,kf. | 9639 |
| 115 | exp heparin, low-molecular-weight/ | 13165 |
| 116 | (low adj3 heparin).tw,kf. | 14080 |
| 117 | (choay or depolymerized heparin or traxyparine).tw,kf. | 101 |
| 118 | (enoxaparin or clexan or clexane or decipar or inhixa or klexane or ledraxen or lovenox or neoparin or qualiop klinik or thorinane).tw,kf. | 4809 |
| 119 | or/109-118 [ANTIVIRAL AGENTS] | 417899 |
| 120 | exp antibodies, monoclonal/ | 250377 |
| 121 | ((monoclonal or clonal or hybridoma) adj3 antibod*).tw,kf. | 203413 |
| 122 | (bamlanivimab or etesevimab or "bamlanivimab/etesevimab" or "2423943-37-5" or "LY-3819253" or "LY-COV555" or "LY3819253" or "UNII-45I6OFJ8QH" or "WHO 11876" or "2423948-94-9" or "anti-Sars-cov-2 antibody JS016" or "CB6" or "JS016" or "LY COV016" or "LY-3832479" or "LY-COV016" or "LY3832479" or "NP005" or "UNII-N7Q9NLF11I" or "WHO 11873").tw,kf. | 287 |
| 123 | (casirivimab or imdevimab or "casirivimab/imdevimab" or regen-cov or "REGN-COV2" or regn10933 or regn10987 or "anti-sars-cov-2 regn-cov2" or "2415933-42-3" or "REGN-10933" or "REGN10933" or "UNII-J0FI6WE1QN" or "WHO 11861" or "2415933-40-1" or "REGN-10987" or "REGN10987" or "UNII-2Z3DQD2JHM" or "WHO 11863").tw,kf. | 56 |
| 124 | ("Sotrovimab" or "2423014-07-5" or "GSK-4182136" or "GSK4182136" or "UNII-1MTK0BPN8V" or "VIR-7831" or "VIR7831").tw,kf. | 4 |
| 125 | ("BRII-196" or "BRII-198" or "DZIF-10c" or "BI 767551" or "SCTA01" or "Ty027" or "HLX70").tw,kf. | 5 |
| 126 | ("C144-LS" or "C-135-LS" or "C144-LS/C-135-LS").tw,kf. | 1 |
| 127 | ("COVI-GUARD" or "STI-1499").tw,kf. | 0 |
| 128 | ("COVI-AMG" or "sti-2020").tw,kf. | 1 |
| 129 | "HFB30132A".tw,kf. | 0 |
| 130 | ("ABBV-47D11" or "ABBV-2B04" or "AZD7442").tw,kf. | 1 |
| 131 | ("BI 767551" or "DZIF-10c" or "COR-101").tw,kf. | 3 |
| 132 | (baricitinib or olumiant or incb28050 or ly3009104).tw,kf. | 541 |
| 133 | or/120-132 [MONOCLONAL ANTIBODIES] | 339036 |
| 134 | ((immune adj3 (sera or serum? or plasma)) or antisera or antiserum or immun#serum).tw,kf. | 77890 |
| 135 | exp immunoglobulins/ | 920127 |
| 136 | (immune* globulin? or immuneglobulin? or immunoglobulin?).tw,kf. | 166596 |
| 137 | ((convalescent adj2 (plasma or serum or sera)) or (plasma adj3 therap*)).tw,kf. | 12428 |
| 138 | or/134-137 [IG THERAPY] | 1030904 |
| 139 | exp ibuprofen/ | 9284 |
| 140 | (ibuprofen or abfen or "aches-n-pain" or "act-3" or actiprofen or "adex 200" or adex liqui-gels or advil or afebril or aktren or aktren spezial or algiasdin or algifor or algofen or algoflex or allipen or alvofen express or "am-fam 400" or anadin or anadvil or analgyl or anbifen or anco or andran or anflagen or antalgil or antarene or antiflam or apo-ibuprofen or aragel or "atril 300" or attritin or balkaprofen or berlistar or bestafen or betaprofen or bifen or bluton or brufanic or brufedol or brufen or brufort or brugesic or brumare or brumed or brupro or buburone or bufect or bufohexal or bupogesic or burana or butacortelone or butifen or caldolor or calprofen or cap-profen or cenbufen or codral period pain or combiflam or contraneural or cuprofen or dalsy or "dc 7034" or "dc7034" or "dg 7034" or "dg7034" or dibufen or "diffutab sr 600" or dimidon or dolan fp or dolgit or dolobene ibu or dolocyl or dolodolgit or dolofen-f or dolomax or dolormin or dolval or donjust b or dorival or druisel or easifon or ecoprofen or emflam or epobron or ergix douleur et fievre or eudorlin extra or exidol or expanfen or febratic or febryn or femapirin or fenalgic or fenbid or flamicon or flarin or froben dolore or galprofen or gelufene or gyno-neuralgin or halprin or haltran or hemagene tailleur or h-loniten or ib-100 or ibalgin or ibofen or ibosure or ibu or ibuberl or ibucalm or ibudak or ibudol or ibudolor or ibufarmalid or ibufen or ibuflam or ibufug or ibugel or ibugesic or ibukern or ibuleve or ibulgan or ibuloid or ibumetin or ibumousse or ibunin or ibupen or ibupirac or ibuprin or ibuprocin or ibuprofene or ibuprohm or ibuprom or iburon or ibusal or ibuspray or ibustar or ibusynth or ibutop or ibux or ibuxin or idyl sr or ifenin or infant's motrin or infibu or inflanor or inflanor forte or ipren or irfen or junifen or junipro or kenfen or kontraneural or lamidon or librofem or lidifen or liptan or lopane or malafene or maxagesic or "mcn r 1451" or medicol or medipren or mediprin or mensoton or midol or momentact or motrin or mynosedin or nagifen-d or napacetin or neobrufen or neobrufen retard or nerofen or neutropain or nobfelon or nobgen or norflam-t or noritis or norton or novogent or novoprofen or nugin or nuprin or nureflex or nurofen or optifen or opturem or ostarin or ostofen or ozonol or paduden or panafen or pedea or pediacare fever or pediprofen or perdophen pediatrie or perofen or phorpain or phorpain gel or proartinal or profen or profeno or proff or proflex or proris or provin or provon or quadrax or rafen or ranofen or rapidophen or rapidophen forte or ratiodolor or rebugen or renidon or reuvol or rhelafen or roidenin or rufen or rupan or saridon n or schufen or seclodin or solufen lidose or solvium or spalt or syntofene or tabalon or tab-profen or taskine or tatanal or tofen or trendar or umafen or unipro or upfen or uprofen or urem or viamal febbre e dolore or zafen or zofen).tw,kf. | 16044 |
| 141 | 139 or 140 [IBUPROFEN] | 17529 |
| 142 | self-management/ or self-care/ | 37352 |
| 143 | (((patient or self) adj (manag* or treat*)) or self care).tw,kf. | 79427 |
| 144 | patient education as topic/ or patient medication knowledge/ or health literacy/ | 93112 |
| 145 | (patient* adj3 (educat* or knowledg* or literacy or literate or behavior* or behaviour* or attitud* or belief* or believ*)).tw,kf. | 93347 |
| 146 | self-help groups/ | 9318 |
| 147 | (((self help or support*) adj3 (group? or club? or servic* or social*)) or (therap* adj3 (club? or group* or social*))).tw,kf. | 123520 |
| 148 | or/142-147 [PATIENT SELF CARE] | 374116 |
| 149 | health services, indigenous/ | 3554 |
| 150 | ((indigenous or metis or aboriginal or inuit or (native adj (america* or alaska* or hawaii* or canad*)) or first nation* or first people*) and (healthcare or (health adj2 (care* or service* or program*)) or healing* or medic* or therap* or remed*)).tw,kf. | 15389 |
| 151 | 149 or 150 [INDIGENOUS THERAPY] | 17516 |
| 152 | exp complementary therapies/ or exp medicine, traditional/ or herbal medicine/ or plants, medicinal/ | 286138 |
| 153 | ((alternat* or complementary or folk or traditional or holistic or chinese or african or tribal) adj2 (medic* or therap* or healing* or treat* or remed*)).tw,kf. | 167250 |
| 154 | (hypnotism or hypnosis or hypnotherapy).tw,kf. | 9236 |
| 155 | (ayurved* or kampo or kanpo or acupunctur* or homeopath*).tw,kf. | 35188 |
| 156 | ((botanical* or herb* or plant or plants or plantlet* or root or roots or natural) adj2 (drug? or extract? or healing* or ingredient? or medic* or preparation* or product or products or remedies or remedy* or supplement* or treat* or therap*)).tw,kf. | 171861 |
| 157 | or/152-156 [ALTERNATIVE THERAPY] | 546523 |
| 158 | disease management/ or pain management/ | 76979 |
| 159 | ((manag* or cope or coping) adj3 (symptom* or disease* or condition* or pain or discomfort*)).tw,kf. | 133106 |
| 160 | 158 or 159 [DISEASE MANAGEMENT] | 193806 |
| 161 | exercise/ or exp running/ or swimming/ or exp walking/ or exp *physical fitness/ or yoga/ or "physical education and training"/ or physical therapy specialty/ | 237679 |
| 162 | ("physical activit*" or walk* or pedestrian* or bicycl* or cycling or cyclist* or biking or bike* or "active lifestyle*" or "aerobic fitness" or "aerobic exercise*" or (running not "running water") or runner* or jog* or swim* or yoga or (physical* adj2 (activit* or active or exercise*)) or ((exercise* or fitness or aerobic*) adj2 (regimen* or training or intervention* or program* or class* or course* or train* or rehab*))).tw,kf. | 500123 |
| 163 | (physiotherap* or ((physical or physio) adj1 (therap* or treat*))).tw,kf. | 56871 |
| 164 | or/161-163 [PHYSICAL THERAPY] | 638019 |
| 165 | exp mental health services/ or exp social work/ or psychiatric rehabilitation/ or psychiatric nursing/ | 129086 |
| 166 | ((mental health or emotion* or psych* or wellbeing or well-being or stress* or wellness* or anxiet* or depress* or obsess* or ocd or mood* or posttrauma* or post-trauma* or ptsd or schizo* or personality disorder* or bipolar* or adhd or attention deficit* or addict*) adj3 (care or service? or support? or treat* or therap* or psychotherap* or counsel* or hotline*)).tw,kf. | 317031 |
| 167 | 165 or 166 [MENTAL HEALTH SERVICES] | 412499 |
| 168 | 51 and (54 or 57 or 60 or 67 or 84 or 108 or 119 or 133 or 138 or 141 or 148 or 151 or 157 or 160 or 164 or 167) | 6197 |
| 169 | Epidemiologic studies/ | 8743 |
| 170 | exp case control studies/ | 1203937 |
| 171 | exp cohort studies/ | 2180275 |
| 172 | Case control.tw. | 135467 |
| 173 | (cohort adj (study or studies)).tw. | 242132 |
| 174 | Cohort analy$.tw. | 9280 |
| 175 | (Follow up adj (study or studies)).tw. | 51575 |
| 176 | (observational adj (study or studies)).tw. | 125247 |
| 177 | (prospect* adj (study or studies)).tw. | 185733 |
| 178 | Longitudinal.tw. | 270765 |
| 179 | Retrospective.tw. | 603898 |
| 180 | Cross sectional.tw. | 405969 |
| 181 | case series.tw. | 84989 |
| 182 | Cross-sectional studies/ | 378623 |
| 183 | method*.tw. | 6987797 |
| 184 | or/169-183 [Adapted from OVID observational studies filter] | 8466765 |
| 185 | (Randomized Controlled Trial or Controlled Clinical Trial or Pragmatic Clinical Trial or Equivalence Trial or Clinical Trial, Phase III).pt. | 631801 |
| 186 | exp Randomized Controlled Trial/ | 539567 |
| 187 | exp Randomized Controlled Trials as Topic/ | 150066 |
| 188 | Controlled Clinical Trial/ | 94308 |
| 189 | exp Controlled Clinical Trials as Topic/ | 155631 |
| 190 | Random Allocation/ | 105654 |
| 191 | Double-Blind Method/ | 165998 |
| 192 | Single-Blind Method/ | 30598 |
| 193 | Placebos/ | 35585 |
| 194 | Control Groups/ | 1758 |
| 195 | (random* or sham or placebo*).ti,ab,hw,kf,kw. | 1611458 |
| 196 | ((singl* or doubl*) adj (blind* or dumm* or mask*)).ti,ab,hw,kf. | 248712 |
| 197 | ((tripl* or trebl*) adj (blind* or dumm* or mask*)).ti,ab,hw,kf. | 1211 |
| 198 | (control* adj3 (study or studies or trial* or group*)).ti,ab,kf. | 1068773 |
| 199 | (Nonrandom* or non random* or non-random* or quasi-random* or quasirandom*).ti,ab,hw,kf. | 47867 |
| 200 | allocated.ti,ab,hw. | 72273 |
| 201 | ((open label or open-label) adj5 (study or studies or trial*)).ti,ab,hw,kf. | 38193 |
| 202 | ((equivalence or superiority or non-inferiority or noninferiority) adj3 (study or studies or trial*)).ti,ab,hw,kf. | 9675 |
| 203 | (pragmatic study or pragmatic studies).ti,ab,hw,kf,kw. | 478 |
| 204 | ((pragmatic or practical) adj3 trial*).ti,ab,hw,kf. | 6121 |
| 205 | ((quasiexperimental or quasi-experimental) adj3 (study or studies or trial*)).ti,ab,hw,kf. | 9314 |
| 206 | (phase adj3 (III or "3") adj3 (study or studies or trial*)).ti,hw,kf. | 30973 |
| 207 | or/185-206 [Adapted from "Strings attached: CADTH database search filters"] | 2314035 |
| 208 | 168 and (184 or 207) | 3380 |
| 209 | case reports/ or (case-stud* or case-report*).jw. or (case* and report*).ti. | 2318346 |
| 210 | (address or autobiography or bibliography or biography or case reports or classical article or clinical trial, veterinary or clinical trials, veterinary as topic or editorial or historical article or interview or news or newspaper article or observational study, veterinary).pt. | 3390505 |
| 211 | 208 not (209 or 210) | 3262 |
| 212 | limit 211 to yr="2020-Current" | 3174 |
| 213 | (202006* or 202007* or 202008* or 202009* or 20201* or 2021* or 2022*).ez,dt,ed. | 2496927 |
| 214 | (2020 06* or 2020 07* or 2020 08* or 2020 09* or 2020 1* or 2021* or 2022*).dp. | 1402458 |
| 215 | 212 and (213 or 214) | 3148 |

Ovid Embase

| **#** | **Searches** | **Results** |
| --- | --- | --- |
| 1 | (long adj (COVID or COVID-19 or COVID19 or coronavirus* or corona virus* or 2019-nCoV or 19nCoV or 2019nCoV or nCoV or n-CoV or "CoV 2" or CoV2 or SARS-CoV-2 or SARS-CoV2 or SARSCoV-2 or SARSCoV2 or SARS2 or SARS-2 or severe acute respiratory syndrome coronavirus 2 or 2019-novel CoV or Sars-coronavirus2 or Sars-coronavirus-2 or SARS-like coronavirus* or novel coronavirus* or novel corona virus* or novel CoV or OC43 or NL63 or 229E or HKU1 or HCoV* or Sars- coronavirus*)).tw,kw. | 316 |
| 2 | ((longterm or long-term) adj (COVID or COVID-19 or COVID19 or coronavirus* or corona virus* or 2019-nCoV or 19nCoV or 2019nCoV or nCoV or n-CoV or "CoV 2" or CoV2 or SARS-CoV-2 or SARS-CoV2 or SARSCoV-2 or SARSCoV2 or SARS2 or SARS-2 or severe acute respiratory syndrome coronavirus 2 or 2019-novel CoV or Sars-coronavirus2 or Sars-coronavirus-2 or SARS-like coronavirus* or novel coronavirus* or novel corona virus* or novel CoV or OC43 or NL63 or 229E or HKU1 or HCoV* or Sars-coronavirus*)).tw,kw. | 30 |
| 3 | ((postacute or post-acute) adj (COVID or COVID-19 or COVID19 or coronavirus* or corona virus* or 2019-nCoV or 19nCoV or 2019nCoV or nCoV or n-CoV or "CoV 2" or CoV2 or SARS-CoV-2 or SARS-CoV2 or SARSCoV-2 or SARSCoV2 or SARS2 or SARS-2 or severe acute respiratory syndrome coronavirus 2 or 2019-novel CoV or Sars-coronavirus2 or Sars-coronavirus-2 or SARS-like coronavirus* or novel coronavirus* or novel corona virus* or novel CoV or OC43 or NL63 or 229E or HKU1 or HCoV* or Sars-coronavirus*)).tw,kw. | 85 |
| 4 | (chronic* adj2 (COVID or COVID-19 or COVID19 or coronavirus* or corona virus* or 2019-nCoV or 19nCoV or 2019nCoV or nCoV or n-CoV or "CoV 2" or CoV2 or SARS-CoV-2 or SARS-CoV2 or SARSCoV-2 or SARSCoV2 or SARS2 or SARS-2 or severe acute respiratory syndrome coronavirus 2 or 2019-novel CoV or Sars-coronavirus2 or Sars-coronavirus-2 or SARS-like coronavirus* or novel coronavirus* or novel corona virus* or novel CoV or OC43 or NL63 or 229E or HKU1 or HCoV* or Sars- coronavirus*)).tw,kw. | 108 |
| 5 | (("90" or ninety or "120" or "180" or hundred*) adj2 day?).tw,kw. | 112182 |
| 6 | (("12" or twelve or "13" or thirteen or "14" or fourteen or "15" or "fifteen" or "16" or sixteen or "17" or seventeen or "18" or eighteen or "19" or nineteen or "20" or "twenty" or "21" or "22" or "23" or "24") adj3 (week? or wk or wks)).tw,kw. | 454237 |
| 7 | (("3" or three or "4" or four or "5" or five or "6" or six or "7" or seven or "8" or eight or "9" or nine or "10" or ten or "11" or eleven or "12" or twelve or "13" or thirteen or "14" or fourteen or "15" or "fifteen" or "16" or sixteen or "17" or seventeen or "18" or eighteen or "19" or nineteen or "20" or "twenty" or "21" or "22" or "23" or "24") adj3 (month? or mo or mos)).tw,kw. | 2074505 |
| 8 | (("1" or one or "2" or two or half or multi* or full) adj3 (year? or yr or yrs)).tw,kw. | 1264922 |
| 9 | (("more than" or longer or "at least" or "after" or follow* or upward* or later* or prospectiv* or retrospectiv* or review* or longitud* or post or beyond) adj5 (day? or week? or wk or wks or month? or mo or mos or year? or yr or yrs)).tw,kw. | 3747544 |
| 10 | (COVID or COVID-19 or COVID19 or coronavirus* or corona virus* or 2019-nCoV or 19nCoV or 2019nCoV or nCoV or n-CoV or SARS-CoV-2 or SARS-CoV2 or SARSCoV-2 or SARSCoV2 or 2019-novel CoV or Sars-coronavirus2 or novel CoV).tw,kw. | 169019 |
| 11 | (5 or 6 or 7 or 8) and 9 and 10 | 4142 |
| 12 | (or/1-4) or 11 [LONG COVID - PT 1] | 4570 |
| 13 | *coronavirus disease 2019/ | 114430 |
| 14 | *severe acute respiratory syndrome coronavirus 2/ | 14698 |
| 15 | *Coronavirinae/ | 1778 |
| 16 | *Betacoronavirus/ | 2660 |
| 17 | *coronavirus infection/ | 4490 |
| 18 | (COVID-19 or COVID19).tw,kw. | 139541 |
| 19 | ((coronavirus* or corona virus*) and (hubei or wuhan or beijing or shanghai)).tw,kw. | 5166 |
| 20 | (wuhan adj5 virus*).tw,kw. | 290 |
| 21 | (2019-nCoV or 19nCoV or 2019nCoV).tw,kw. | 1754 |
| 22 | (nCoV or n-CoV or "CoV 2" or CoV2).tw,kw. | 53061 |
| 23 | (SARS-CoV-2 or SARS-CoV2 or SARSCoV-2 or SARSCoV2 or SARS2 or SARS-2 or severe acute respiratory syndrome coronavirus 2).tw,kw. | 54000 |
| 24 | (2019-novel CoV or Sars-coronavirus2 or Sars-coronavirus-2 or SARS-like coronavirus* or ((novel or new or nouveau) adj2 (CoV or nCoV or covid or coronavirus* or corona virus or Pandemi*2)) or (coronavirus* and pneumonia)).tw,kw. | 19625 |
| 25 | (novel coronavirus* or novel corona virus* or novel CoV).tw,kw. | 9606 |
| 26 | ((coronavirus* or corona virus*) adj2 "2019").tw,kw. | 30723 |
| 27 | ((coronavirus* or corona virus*) adj2 "19").tw,kw. | 4963 |
| 28 | (coronavirus 2 or corona virus 2).tw,kw. | 16237 |
| 29 | (OC43 or NL63 or 229E or HKU1 or HCoV* or Sars-coronavirus*).tw,kw. | 4129 |
| 30 | (coronavirus* or corona virus*).ti. | 22755 |
| 31 | or/13-30 [COVID-19] | 168950 |
| 32 | ((post or after or following) adj (COVID or COVID-19 or COVID19 or coronavirus* or corona virus* or 2019-nCoV or 19nCoV or 2019nCoV or nCoV or n-CoV or "CoV 2" or CoV2 or SARS-CoV-2 or SARS-CoV2 or SARSCoV-2 or SARSCoV2 or SARS2 or SARS-2 or severe acute respiratory syndrome coronavirus 2 or 2019-novel CoV or Sars-coronavirus2 or Sars-coronavirus-2 or SARS-like coronavirus* or novel coronavirus* or novel corona virus* or novel CoV or OC43 or NL63 or 229E or HKU1 or HCoV* or Sarscoronavirus*)).tw,kw. | 3554 |
| 33 | (abnormalities or ache* or brain fog or breath* or challeng* or "co morbid*" or comorbid* or complications or concentrat* or condition* or consequences or convalescen* or cough or difficulties or disease* or disorder* or disturbances or dizziness or dysfunction or dyspnea or effects or fatigue or function* or gustatory or headache* or illness* or impairments or issues or lung or manifestation* or morbidit* or "multi morbid*" or multimorbid* or neuro* or olfaction or olfactory or outcome? or pain* or palpitation* or problem? or prognos* or recuperat* or respiratory or risk* or sickness* or sign or signs or smell* or surviv* or symptom* or syndrome* or tachycardia or taste?).tw,kw. | 22221480 |
| 34 | 32 and 33 | 2747 |
| 35 | ((chronic* or continuous* or continual* or continuing* or delay* or endur* or extend* or fluctuat* or gradual* or lasting* or legacy* or lengthy* or linger* or long* or "medium* term*" or mediumterm* or multisystem* or "multi system*" or ongoing* or permanent* or persist* or prolong* or protract* or relaps* or remission* or remit* or residual* or slow* or subacute* or "sub acute*") adj3 recover*).tw,kw. | 45527 |
| 36 | ((after discharg* or following discharg* or postacute* or "post acute*" or postdischarg* or "post discharge" or "post discharging" or posthospital* or post-hospital* or postinfect* or "post infection" or "post infective*" or postviral* or "post viral*" or postvirus* or "post virus*" or postcritical or post-critical or postintensive or post-intensive or post-ICU) adj3 recover*).tw,kw. | 807 |
| 37 | ((chronic* or continuous* or continual* or continuing* or delay* or endur* or extend* or fluctuat* or gradual* or lasting* or legacy* or lengthy* or linger* or long* or "medium* term*" or mediumterm* or multisystem* or "multi system*" or ongoing or permanent* or persist* or prolong* or protract* or relaps* or remission* or remit* or residual* or slow* or subacute* or "sub acute*") adj3 (complication? or consequence? or convalescen* or disabilit* or feature* or illness* or prognos* or sequela* or sign or signs or suffering? or symptom* or recuperat*)).tw,kw. | 350235 |
| 38 | ((after discharg* or following discharg* or postacute* or "post acute*" or postdischarg* or "post discharge" or "post discharging" or posthospital* or post-hospital* or postinfect* or "post infection" or "post infective*" or postviral* or "post viral*" or postvirus* or "post virus*" or postcritical or post-critical or postintensive or post-intensive or post-ICU) adj3 (complication? or consequence? or convalescen* or disabilit* or feature* or illness* or prognos* or sequela* or sign or signs or suffering? or symptom* or recuperat*)).tw,kw. | 2769 |
| 39 | (nonrecover* or "non recover*" or "not recover*").tw,kw. | 9452 |
| 40 | ("long* haul*" or longhaul* or "long* tail*" or longtail* or longduration* or "long duration*" or longlast* or "long last*" or longstanding* or "long standing*" or "medium* term*" or mediumterm*).tw,kw. | 150126 |
| 41 | 34 or 35 or 36 or 37 or 38 or 39 [LONG-TERM ILLNESS, PROTRACTED RECOVERY, ETC.] | 406115 |
| 42 | 31 and 41 [LONG COVID - PT 2] | 5313 |
| 43 | *convalescence/ | 7191 |
| 44 | 31 and 43 [LONG COVID - PT 3] | 279 |
| 45 | 12 or 42 or 44 [LONG COVID LONG COVID HEDGE: DECARY ET AL, 2021] | 9257 |
| 46 | exp *ambulatory care/ or exp *patient care/ or exp *preventive health service/ or *community care/ or *community based rehabilitation/ or exp *community health nursing/ or *community integration/ or exp *community mental health service/ or *community program/ or *health education/ or *health literacy/ or exp *health promotion/ or *patient education/ or *primary prevention/ or *secondary prevention/ or *tertiary prevention/ or *quaternary prevention/ or *mass drug administration/ or exp *prophylaxis/ or *health service/ or exp *emergency health service/ or exp *hospital service/ or *public health service/ or exp *child health care/ or *hotline/ or exp *elderly care/ or exp *rural health care/ or *therapy/ or exp *drug therapy/ or exp *rehabilitation/ | 1880476 |
| 47 | (intervention* or service* or health care* or healthcare* or health system* or program* or strategy* or strategies or framework* or policy or policies or guideline* or treat* or therap* or prevent* or rehab* or pharmaceutic* or medication* or drug* or hotline* or clinic or clinics or health centre* or health center* or hospital* or practice*).ti,kw. | 5875547 |
| 48 | 46 or 47 [BROAD LEVEL INTERVENTIONS] | 6957562 |
| 49 | exp *aftercare/ or *outpatient/ or *outpatient care/ | 73378 |
| 50 | (aftercare or after care or follow-up care or referral?).tw,kw. | 213478 |
| 51 | 49 or 50 [AFTER CARE] | 281870 |
| 52 | *health care delivery/ or *nonbiological model/ | 65008 |
| 53 | ((healthcare or health care or clinical* or public health or community) adj4 (approach* or model? or service? or deliver* or system* or distribut* or framework? or guideline?)).tw,kw. | 548109 |
| 54 | 52 or 53 [HEALTHCARE MODELS] | 599420 |
| 55 | exp *vaccine/ | 203675 |
| 56 | exp *severe acute respiratory syndrome vaccine/ | 2801 |
| 57 | (vaccine* or vaccinat* or revaccin* or immunis* or immuniz* or inoculat*).tw,kw. | 590031 |
| 58 | (prophyla* or prevent* or protect* or postexpos* or (expos* adj3 (post or after or document* or suspect* or positiv* or confirm*))).tw,kw. | 3211362 |
| 59 | (postcontact* or (contact* adj3 (post or after or document* or suspect* or positiv* or confirm*))).tw,kw. | 18037 |
| 60 | or/55-59 [VACCINATION] | 3659808 |
| 61 | exp *vitamin/ | 278247 |
| 62 | exp *folic acid/ | 20438 |
| 63 | *dietary supplement/ | 5975 |
| 64 | vitamin?.tw,kw. | 283759 |
| 65 | riboflavin.tw,kw. | 11219 |
| 66 | (niacinamide or enduramide or nicobion or nicotinamide or papulex).tw,kw. | 24975 |
| 67 | (thiamine or aneurin or thiamin).tw,kw. | 13203 |
| 68 | (folic acid or folacin or folate or folvite or pteroylglutamic acid).tw,kw. | 54742 |
| 69 | (ascorbic acid or ferrous ascorbate or hybrin or "l-ascorbic acid" or magnesium ascorbate or magnesium ascorbicum or "magnesium di-l-ascorbate" or magnorbin or sodium ascorbate).tw,kw. | 38979 |
| 70 | ((diet* or nutrition* or herbal* or food?) adj3 supplement*).tw,kw. | 83371 |
| 71 | or/61-70 [NUTRTIONAL SUPPLEMENTS] | 572793 |
| 72 | exp *antiinflammatory agent/ | 728124 |
| 73 | (((antiinflammator* or anti inflammator*) adj2 (drug? or pharmaceutical? or agent? or substance? or medicin* or prescription?)) or NSAID or NSAIDs).tw,kw. | 104335 |
| 74 | exp *corticosteroid/ | 300508 |
| 75 | (adrenal cortex hormone? or corticoid? or cortical steroid? or cortico steroid? or corticosteroid? or dermocorticosteroid?).tw,kw. | 172525 |
| 76 | (dexamethasone? or decaject? or decameth? or decaspray? of dexasone? or dexpak? or hexadecadrol? or hexadrol? or maxidex? or methylfluorprednisolone? or millicorten? or oradexon?).tw,kw. | 83010 |
| 77 | (prednisone or cortan or cortancyl or cutason or dacortin or decortin or decortisyl or dehydrocortisone or deltasone or encorton or encortone or enkortolon or kortancyl or liquid pred or meticorten or orasone or panafcort or panasol or predni tablinen or prednidib or predniment or prednison acsis or prednison galen or prednison hexal or pronisone or rectodelt or sone or sterapred or ultracorten or winpred or delta cortisone).tw,kw. | 53605 |
| 78 | (methylprednisolone or methylprednisolone or medrol or metipred or urbason).tw,kw. | 30972 |
| 79 | (hydrocortisone or "acticort" or "aeroseb hc" or "ala-cort" or "ala-scalp" or "alfacort" or "algicortis" or "alkindi" or "alpha derm" or "alphaderm" or "anucort-hc" or "anumed-hc" or "anutone-hc" or "aquanil hc" or "balneol-hc" or "barseb hc" or "beta-hc" or "biacort" or "cetacort" or "cobadex" or "colocort" or "compound f" or "cordicare lotion" or "coripen" or "cort dome" or "cortef" or "cortef cream" or "cortenema" or "cortibel" or "corticorenol" or "cortifan" or "cortiphate" or "cortisol" or "cortisole" or "cortispray" or "cortoderm" or "cortril" or "cotacort" or "covocort" or "cremicort-h" or "cutaderm" or "dermacrin hc lotion" or "dermaid" or "derm-aid cream" or "dermaid soft cream" or "dermocare" or "dermocortal" or "dermolate" or "dioderm" or "eczacort" or "ef cortelan" or "efcortelan" or "egocort" or "egocort cream" or "eksalb" or "eldecort" or "emo-cort" or "epicort" or "ficortril" or "filocot" or "flexicort" or "glycort" or "gly-cort" or "hc no. 1" or "hc no. 4" or "h-cort" or "hebcort" or "hebcort v" or "hemorrhoidal hc" or "hemril-30" or "hemril-hc uniserts" or "hi-cor" or "hidrotisona" or "hycor" or "hycort" or "hydracort" or "hydrasson" or "hydro ricortex" or "hydrocort" or "hydrocorticosteroid" or "hydrocortisate" or "hydrocortison" or "hydrocortisonum" or "hydrocortisyl" or "hydrocortone" or "hydrogalen" or "hydrokort" or "hydrokortison" or "hydro-rx" or "hydrotopic" or "hysone" or "hytisone" or "hytone" or "hytone lotion" or "incortin h" or "instacort 10" or "kyypakkaus" or "lacticare hc" or "lemnis fatty cream hc" or "lenirit" or "medihaler cort" or "medihaler duo" or "medrocil" or "mildison" or "mitocortyl demangeaisons" or "munitren" or "nogenic hc" or "novohydrocort" or "nsc 10483" or "nsc 741" or "nsc10483" or "nutracort" or "optef" or "otosone f" or "penecort" or "plenadren" or "prepcort" or "prevex hc" or "pro cort" or "procort" or "proctocort" or "proctosert hc" or "proctosol-hc" or "proctosone" or "proctozone hc" or "procutan" or "rectasol-hc" or "rectocort" or "rederm" or "sanatison" or "scalp-aid" or "schericur" or "schericur 0.25%" or "scherosone f" or "sistral hydrocort" or "skincalm" or "stie-cort" or "substance m" or "synacort" or "texacort" or "triburon-hc" or "unicort" or "vasocort").tw,kw. | 99531 |
| 80 | (nitric oxide or endogenous nitrate vasodilator or mononitrogen monoxide or nitrogen monoxide or genosyl or inomax or noxivent).tw,kw. | 198085 |
| 81 | (prednisolone or "adelcort" or "antisolon" or "antisolone" or "aprednislon" or "aprednislone" or "benisolon" or "benisolone" or "berisolon" or "berisolone" or "caberdelta" or "capsoid" or "codelcortone" or "co-hydeltra" or "compresolon" or "cortadeltona" or "cortadeltone" or "cortalone" or "cortelinter" or "cortisolone" or "cotolone" or "dacortin" or "dacortin h" or "dacrotin" or "decaprednil" or "decortin h" or "decortril" or "dehydro cortex" or "dehydro hydrocortison" or "dehydro hydrocortisone" or "dehydrocortex" or "dehydrocortisol" or "dehydrocortisole" or "dehydrohydrocortison" or "dehydrohydrocortisone" or "delcortol" or "delta 1 hydrocortisone" or "delta cortef" or "delta cortril" or "delta ef cortelan" or "delta f" or "delta hycortol" or "delta hydrocortison" or "delta hydrocortisone" or "delta ophticor" or "delta stab" or "delta1 dehydrocortisol" or "delta1 dehydrohydrocortisone" or "delta1 hydrocortisone" or "deltacortef" or "deltacortenolo" or "deltacortil" or "deltacortoil" or "deltacortril" or "deltaderm" or "deltaglycortril" or "deltahycortol" or "deltahydrocortison" or "deltahydrocortisone" or "deltaophticor" or "deltasolone" or "deltastab" or "deltidrosol" or "deltisilone" or "deltisolon" or "deltisolone" or "deltolasson" or "deltolassone" or "deltosona" or "deltosone" or "depo-predate" or "dermosolon" or "dhasolone" or "di adreson f" or "di adresone f" or "diadreson f" or "diadresone f" or "dicortol" or "domucortone" or "encortelon" or "encortelone" or "encortolon" or "equisolon" or "fernisolone-p" or "glistelone" or "hefasolon" or "hostacortin h" or "hostacortin h vet" or "hydeltra" or "hydeltrone" or "hydrelta" or "hydrocortancyl" or "hydrocortidelt" or "hydrodeltalone" or "hydrodeltisone" or "hydroretrocortin" or "hydroretrocortine" or "inflanefran" or "insolone" or "keteocort h" or "key-pred" or "lenisolone" or "leocortol" or "liquipred" or "lygal kopftinktur n" or "mediasolone" or "meprisolon" or "meprisolone" or "metacortalon" or "metacortalone" or "metacortandralon" or "metacortandralone" or "metacortelone" or "meti derm" or "meticortelone" or "metiderm" or "morlone" or "mydrapred" or "neo delta" or "nisolon" or "nisolone" or "nsc 9120" or "nsc9120" or "opredsone" or "panafcortelone" or "panafcortolone" or "panafort" or "paracortol" or "phlogex" or "pre cortisyl" or "preconin" or "precortalon" or "precortancyl" or "precortisyl" or "predacort 50" or "predaject-50" or "predalone 50" or "predartrina" or "predartrine" or "predate-50" or "predeltilone" or "predisole" or "predisyr" or "pred-ject-50" or "predne dome" or "prednecort" or "prednedome" or "prednelan" or "predni coelin" or "predni h tablinen" or "prednicoelin" or "prednicort" or "prednicortelone" or "prednifor drops" or "predni-helvacort" or "predniment" or "predniretard" or "prednis" or "prednisil" or "prednisolon" or "prednisolona" or "prednisolone alcohol" or "prednisolone h" or "prednisolone oleosae sr 82" or "prednivet" or "prednorsolon" or "prednorsolone" or "predonine" or "predorgasolona" or "predorgasolone" or "prelon" or "prelone" or "prenilone" or "prenin" or "prenolone" or "preventan" or "prezolon" or "rubycort" or "scherisolon" or "scherisolona" or "serilone" or "solondo" or "solone" or "solupren" or "soluprene" or "spiricort" or "spolotane" or "sterane" or "sterolone" or "supercortisol" or "supercortizol" or "taracortelone" or "walesolone" or "wysolone").tw,kw. | 43179 |
| 82 | (aspirin or acetylsalicylic acid or "8-hour bayer" or "acenterine" or "acesal" or "acetan" or "acetard" or "aceticil" or "aceticyl" or "acetilum" or "acetonyl" or "acetophen" or "acetosal" or "acetosalicylic acid" or "acetosalin" or "acetosalum" or "acetyl salicylate" or "acetyl salicylic acid" or "acetylic salicylic acid" or "acetylin" or "acetylo" or "acetylo salicylic acid" or "acetylon" or "acetylosalicylic acid" or "acetylsal" or "acetylsalicyclic acid" or "acetylsalicyl" or "acetylsalicylate" or "acetylsalicylate strontium" or "acetylsalicylic acid plus glycine" or "acetylsalicylic acid sodium salt" or "acetylsalicylic acid strontium salt" or "acetylsalycic acid" or "acetylsalycylic acid" or "acetysal" or "acidulatum" or "acidum acetyl salicylicum" or "acidum acetylosalicylicum" or "acidum acetylsalicylicum" or "actorin" or "acylpyrin" or "acylpyrine" or "acytosal" or "adiro" or "alabukun" or "alasil" or "albyl e" or "albyl minor" or "alka seltzer" or "alkaspirin" or "anasprin" or "andol" or "anopyrin" or "ansin" or "anthrom" or "aptor" or "arthralgyl" or "arthritis strength bufferin" or "asacard" or "asaetta" or "asaflow" or "asaphen" or "asapor" or "asatard" or "asawin" or "aspec" or "aspent" or "aspergum" or "aspex" or "aspilets" or "aspirem" or "aspirgran" or "aspiricor" or "aspirina" or "aspirine" or "aspirinine" or "aspirisucre" or "aspisol" or "aspo cid" or "aspro" or "aspro cardio" or "aspro clear" or "asproflash" or "asrina" or "asrivo" or "asta" or "asteric" or "asteric acid" or "astrix" or "bamyl" or "bayaspirina" or "bebesan" or "biprin" or "bokey" or "boxazin" or "breoprin" or "bufferin" or "cafenol" or "cardioasa" or "cardioasae" or "cardioaspirina" or "cartia" or "caspirin" or "catalgine" or "catalgix" or "cemerit" or "cemirit" or "claradin" or "claragine" or "colfarit" or "comoprin" or "contrheuma" or "contrheuma retard" or "darosal" or "depot aspirin" or "dispirin" or "dolean" or "durlaza" or "dusil" or "easprin" or "ecasil" or "ecosprin" or "ecotrin" or "egalgic" or "emocin" or "empirin" or "encaprin" or "encine em" or "endosprin" or "entaprin" or "entericin" or "enteroprin" or "enterosarine" or "enterospirine" or "entrophen" or "eskotrin" or "euthermine" or "extren" or "flamasacard" or "genasprin" or "globentyl" or "godamed" or "gotosan" or "helicon" or "herz ass" or "hjertemagnyl" or "idotyl" or "infatabs a" or "istopirin" or "istopyrine" or "ivepirine" or "juvepirine" or "keypo" or "kilios" or "kinderaspirin" or "magnecyl brus" or "magnyl dak" or "mcn r 358" or "measurin" or "mejoral" or "melabon" or "micristin" or "micropyrin" or "migrasaa" or "mikristin" or "miniasal" or "mycristin" or "naspro" or "novasen" or "nu seal" or "nuseals" or "nu-seals" or "nu-seals asa" or "ortho acetoxybenzoate" or "ortho acetoxybenzoic acid" or "ortho acetyloxybenzoate" or "ortho acetyloxybenzoic acid" or "ostoprin" or "pancemol" or "para acetylsalicylic acid" or "paracin" or "paynocil" or "pengo" or "platet 300 cleartab" or "plewin" or "polopiryna" or "premaspin" or "primaspan" or "proprin" or "pyronoval" or "reumyl" or "rhodine" or "rhonal" or "ronal" or "salacetin" or "salacetogen" or "saletin" or "salisalido" or "salospir" or "sargepirine" or "sedergine" or "sedergine forte" or "sodium acetylsalicylate" or "sodium bicarbonate acetyl salicylate" or "sodium bicarbonate acetylsalicylate" or "soldral" or "solpyron" or "solucetyl" or "solupsa" or "spren" or "super tru" or "tapal" or "temagin" or "tevapirin" or "th 2152" or "thrombo-aspilets" or "toldex retard" or "treupahlin" or "treuphalin" or "tromalyt" or "tromcor" or "turivital" or "vazalore" or "verin" or "vitalink" or "xaxa" or "zorprin").tw,kw. | 136083 |
| 83 | (adalimumab or "abp 501" or "abp501" or "abrilada" or "abt d2e7" or "abtd2e7" or "adaly" or "amgevita" or "amjevita" or "amsparity" or "avt 02" or "avt02" or "bat 1406" or "bat1406" or "bax 2923" or "bax 923" or "bax2923" or "bax923" or "bi 695501" or "bi695501" or "chs 1420" or "chs1420" or "cinnora" or "ct p17" or "ctp17" or "cyltezo" or "da 3113" or "da3113" or "dmb 3113" or "dmb3113" or "exemptia" or "fkb 327" or "fkb327" or "fyzoclad" or "gp 2017" or "gp2017" or "hadlima" or "halimatoz" or "hefiya" or "hlx 03" or "hlx03" or "hulio" or "humira" or "hyrimoz" or "ibi 303" or "ibi303" or "idacio" or "imraldi" or "kromeya" or "lu 200134" or "lu200134" or "m 923" or "m923" or "mabura" or "monoclonal antibody d2e7" or "msb 11022" or "msb11022" or "ons 3010" or "ons3010" or "pf 06410293" or "pf 6410293" or "pf06410293" or "pf6410293" or "raheara" or "sb 5" or "sb5" or "solymbic" or "trudexa" or "zrc 3197" or "zrc3197").tw,kw. | 23056 |
| 84 | (celecoxib or "aclarex" or "artilog" or "artroxil" or "caditar" or "celcox" or "celebra" or "celebreks" or "celebrex" or "celecox" or "celib" or "celora" or "coxel" or "coxid" or "dilox" or "eliflam" or "elyxyb" or "lexfin" or "onsenal" or "sc 58635" or "sc58635" or "solexa" or "ym 177" or "ym177" or "zycel").tw,kw. | 12657 |
| 85 | (diclofenac or "abdiflam" or "abitren" or "acuflam" or "akis" or "algipatch" or "algistick" or "algopain eze" or "algoplast" or "allvoran" or "almiral" or "alonpin" or "antalcalm" or "apo-diclofenac ec" or "arcanafenac" or "arthrifen" or "artren" or "artrenac" or "artrites" or "assaren" or "athrofen" or "ba 47210" or "ba47210" or "berafen gel" or "berifen" or "betaren" or "bolabomin" or "calozan" or "catanac" or "catas" or "cencenag" or "clo-far" or "clofec" or "clofen" or "clonac" or "clonaren" or "clonodifen" or "cordralan" or "curinflam" or "ddl plaster" or "declophen" or "decrol" or "deflamat" or "deflam-k" or "delphinac" or "denaclof" or "depain" or "diceus" or "dicipan" or "diclac" or "diclax" or "diclo" or "diclobasan" or "diclobene" or "diclod" or "diclodent" or "diclodoc" or "diclodolor" or "diclofen" or "diclofen cremogel" or "diclofenac rekur" or "diclofenac resin" or "diclofenac resinate" or "diclofenac sodium" or "dicloflam" or "diclohexal" or "dicloin" or "diclomax" or "diclomol" or "diclon" or "diclopax" or "diclophenac sodium" or "diclopuren" or "dicloral" or "dicloran gel" or "diclorecip" or "dicloren" or "dicloreum" or "diclosan sr" or "diclosian" or "diclotec" or "diclowal" or "dicsnal" or "difen" or "difena" or "difenac" or "difenol gel" or "difnal k" or "dioxaflex" or "dioxaflex retard" or "divoltar" or "dixol" or "doflastad" or "doflex" or "dolaren" or "dolflam-retard" or "dolo voltaren" or "doloflam" or "dolotren" or "doragon" or "dosanac" or "duravolten" or "dycon sr" or "dyloject" or "ecofenac" or "econac" or "effekton" or "eflagen" or "epifenac" or "eslofen" or "evadol" or "evinopon" or "feloran" or "fenac" or "fenadium" or "fenaspec" or "flameril" or "flexagen" or "flogofenac" or "flogosin d" or "flogozan" or "fortfen sr" or "freejex" or "gp 45840" or "grofenac" or "hizemin" or "imflac" or "inac gel" or "indicam" or "inflamac" or "inflanac" or "isv 205" or "isv205" or "jonac gel" or "kadiflam" or "kinespir" or "klofen l" or "klotaren" or "kriplex" or "lesflam" or "leviogel" or "lifenac" or "lofenac" or "lotirac" or "magluphen" or "merflam" or "modifenac" or "monoflam" or "motifene" or "naboal" or "nac gel" or "naclof" or "nacoflar" or "nadifen" or "novapirina" or "novo-difenac" or "novolten" or "ofenac" or "olfen" or "optanac" or "orthophen" or "osteoflam" or "painstop" or "panamor" or "pennsaid" or "profenac" or "relaxyl gel" or "remethan" or "renvol emulgel" or "rewodina" or "rheufenac" or "rheumafen" or "rheumatac" or "rheumatac retard" or "rhewlin" or "rhewlin sr" or "rhumalgan" or "rolactin" or "sailib" or "savismin" or "sefnac" or "slofenac" or "sodium diclofenac" or "solaraze" or "sophenoderm" or "soproxen" or "spraymik" or "sr 318t" or "staren" or "sting gel" or "tabiflex" or "tds 943" or "tds943" or "tigen plaster" or "toraren" or "traulen" or "tsudohmin" or "uniclonax" or "uniren" or "valentac" or "vartelon" or "veral" or "voldal" or "voldic" or "volero" or "volfenac" or "volna-k" or "volsaid" or "volta" or "voltadex emulgel" or "voltadvance" or "voltalen" or "voltalen emulgel" or "voltalgan" or "voltaren" or "voltarene" or "voltarenspe" or "voltarol" or "voltine" or "voltral" or "voltrix" or "voren emulgel" or "votalen" or "voveran" or "vurdon" or "wergyl" or "xenid" or "yuren" or "zolterol" or "zorvolex").tw,kw. | 22045 |
| 86 | (dimethyl sulfoxide or "damul" or "demasorb" or "demavet" or "demeso" or "demexide" or "dimethyl sulphoxide" or "dimethylsulfoxide" or "dimethylsulphoxide" or "dimexide" or "dms 70" or "dms 90" or "dms70" or "dms90" or "dmso" or "dolicur" or "domoso" or "dromisol" or "gamasal 90" or "hyadur" or "infiltrina" or "methyl sulfoxide" or "methylsulfoxide" or "nsc 763" or "nsc763" or "rimso 100" or "rimso 50" or "somipront" or "sq 9453" or "sq9453" or "syntexan").tw,kw. | 40713 |
| 87 | (indomethacin or " algiflam" or "algometacin" or "amuno" or "antalgin dialicels" or "apo-indomethacin" or "areumatin" or "argilex" or "arthrexin" or "articulen" or "artracin" or "artrilona s" or "artrinovo" or "artrocid" or "asimet" or "benocid" or "betacin" or "bonidon" or "boutycin" or "catlep" or "chrono indocid" or "chronoindocid" or "confortid" or "docin" or "dolazal" or "dolazol" or "dolcidium" or "dometin" or "durametacin" or "elmego spray" or "elmetacin" or "endometacin" or "flamaret" or "flexin continus" or "grindocin" or "helvecin" or "idicin" or "im-75" or "imbrilon" or "imet" or "inacid" or "indacin" or "indalgin" or "inderapollon" or "indicin" or "indo phlogont" or "indocap" or "indocid" or "indocin" or "indocolir" or "indocollyre" or "indogesic" or "indolag" or "indolar sr" or "indolemmon" or "indo-lemmon" or "indomecin" or "indomed" or "indomee" or "indomelan" or "indomelol" or "indomet retard" or "indometacin sodium" or "indometacine" or "indomethacin" or "indomethacine" or "indomethacinum" or "indomethegan" or "indometicina mckesson" or "indometin" or "indometin depot" or "indomexum" or "indomin" or "indono" or "indoptic" or "indoptol" or "indorektal" or "indorem" or "indos" or "indosan" or "indosima" or "indosmos" or "indo-tablinen" or "indotard" or "indovis" or "indoxen" or "indoy" or "indren" or "indrenin" or "indylon" or "inflazon" or "inmetsin" or "inteban" or "lauzit" or "luiflex" or "lyo indometacin trihydrate" or "malival" or "mcn r 1166" or "mcn r1166" or "metacen" or "methacin" or "methindol" or "methindole" or "methocaps" or "metindol" or "mezolin" or "miometacen" or "mk 615" or "mk615" or "mobilan" or "novomethacin" or "osmogit" or "osmosin" or "reumacid" or "reusin" or "rheumacid" or "rheumacin" or "salinac" or "servimeta" or "sidocin" or "tannex" or "taye" or "tivorbex" or "vi-gel" or "vonum").tw,kw. | 45614 |
| 88 | (methotrexate or "abitrexate" or "amethopterin" or "amethopterine" or "ametopterine" or "antifolan" or "biotrexate" or "canceren" or "cl 14377" or "cl14377" or "emtexate" or "emthexat" or "emthexate" or "emtrexate" or "enthexate" or "farmitrexat" or "farmitrexate" or "farmotrex" or "folex" or "ifamet" or "imeth" or "intradose mtx" or "jylamvo" or "lantarel" or "ledertrexate" or "maxtrex" or "metex" or "methoblastin" or "methohexate" or "methotrate" or "methotrexat" or "methotrexato" or "methoxtrexate" or "methrotrexate" or "methylaminopterin" or "methylaminopterine" or "meticil" or "metoject" or "metothrexate" or "metotrexat" or "metotrexate" or "metotrexin" or "metrex" or "mexate" or "mpi 5004" or "mpi5004" or "neotrexate" or "nordimet" or "novatrex" or "nsc 740" or "nsc740" or "otrexup" or "otrexup pfs" or "rasuvo" or "reditrex" or "reumatrex" or "rheumatrex" or "texate" or "texorate" or "trexall" or "xaken" or "xatmep" or "zexate").tw,kw. | 73788 |
| 89 | (naproxen or " acusprain" or "aflamax" or "aflaxen" or "agilex" or "agilxen" or "aleve" or "alpoxen" or "alpron" or "anaprox" or "anexopen" or "apo-naproxen" or "apranax" or "apraxin" or "apronax" or "artagen" or "artron" or "artroxen" or "axer alfa" or "babel" or "bipronyl" or "bonyl" or "congex" or "crysanal" or "dafloxen" or "daprox" or "daprox entero" or "deflamox" or "dextro naproxen" or "diferbest" or "diocodal" or "dolormin fuer frauen" or "dolormin fur frauen" or "dolormin gs" or "dysmenalgit" or "dysmenalgit n" or "ec naprosyn" or "equiproxen" or "femex" or "feminax ultra" or "flanax" or "flanax forte" or "floginax" or "flonax" or "floxene" or "fuxen" or "galpharm period pain relief" or "gibixen" or "headlon" or "iraxen" or "laraflex" or "lasonil antinfiammatorio e antireumatico" or "lefaine" or "leniartil" or "levo naproxen" or "licorax" or "methoxypropiocin" or "miranax" or "momendol" or "nafasol" or "naixan" or "napolon" or "naposin" or "napreben" or "naprelan" or "napren" or "naprium" or "naprius" or "naproflam" or "naprogesic" or "naprong" or "naprontag" or "naprorex" or "naprossene" or "naprostad" or "naprosyn" or "naprosyne" or "naprovite" or "naproxeno" or "naproxi 250" or "naproxi 500" or "naproxyn" or "naprozyne" or "naprux" or "napsyn" or "napxen" or "narma" or "narocin" or "naxen" or "naxopren" or "naxyn" or "neprossin" or "norswel" or "novonaprox" or "novo-naprox" or "novuran" or "nuprafem" or "nycopren" or "pactens" or "prexan" or "priaxen" or "prodilor" or "pronaxen" or "proxen" or "proxidol" or "rahsen" or "rs 3540" or "rs 3650" or "rs3540" or "rs3650" or "sanomed" or "saritilron" or "seladin" or "shiprosyn" or "sutolin" or "synaprosyn" or "synflex" or "tohexen" or "uniflam" or "u-ritis" or "velsay" or "veradol" or "vinsen" or "wintrex" or "xenar" or "xenobid").tw,kw. | 11438 |
| 90 | or/72-89 [ANTI-INFLAMMATORY AGENTS] | 1421692 |
| 91 | exp *antivirus agent/ | 339793 |
| 92 | ((antivir* or anti vir*) adj2 (drug? or pharmaceutical? or agent? or substance? or medicin* or prescription?)).tw,kw. | 30362 |
| 93 | remdesivir.tw,kw. | 1797 |
| 94 | *tocilizumab/ | 4384 |
| 95 | (tocilizumab or actemra or atlizumab or lusinex or "r 1569" or r1569 or roactemra).tw,kw. | 10531 |
| 96 | *baricitinib/ | 739 |
| 97 | (baricitinib or "incb 028050" or "incb 28050" or incb028050 or incb28050 or "ly 3009104" or ly300910 or olumiant).tw,kw. | 1267 |
| 98 | ((anticoagulant* or anti coagulant* or antithrombo* or anti-thrombo*) adj2 (drug? or pharmaceutical? or agent? or substance? or medicin* or prescription?)).tw,kw. | 14776 |
| 99 | exp *low molecular weight heparin/ | 16289 |
| 100 | (low adj3 heparin).tw,kw. | 22134 |
| 101 | (choay or depolymerized heparin or traxyparine).tw,kw. | 362 |
| 102 | (enoxaparin or clexan or clexane or decipar or inhixa or klexane or ledraxen or lovenox or neoparin or qualiop klinik or thorinane).tw,kw. | 11770 |
| 103 | or/91-102 [ANTIVIRAL AGENTS] | 415832 |
| 104 | exp *monoclonal antibody/ | 238023 |
| 105 | ((monoclonal or clonal or hybridoma) adj3 antibod*).tw,kw. | 255171 |
| 106 | (bamlanivimab or etesevimab or "bamlanivimab/etesevimab" or "2423943-37-5" or "LY-3819253" or "LY-COV555" or "LY3819253" or "UNII-45I6OFJ8QH" or "WHO 11876" or "2423948-94-9" or "anti-Sars-cov-2 antibody JS016" or "CB6" or "JS016" or "LY COV016" or "LY-3832479" or "LY-COV016" or "LY3832479" or "NP005" or "UNII-N7Q9NLF11I" or "WHO 11873").tw,kw. | 305 |
| 107 | (casirivimab or imdevimab or "casirivimab/imdevimab" or regen-cov or "REGN-COV2" or regn10933 or regn10987 or "anti-sars-cov-2 regn-cov2" or "2415933-42-3" or "REGN-10933" or "REGN10933" or "UNII-J0FI6WE1QN" or "WHO 11861" or "2415933-40-1" or "REGN-10987" or "REGN10987" or "UNII-2Z3DQD2JHM" or "WHO 11863").tw,kw. | 56 |
| 108 | ("Sotrovimab" or "2423014-07-5" or "GSK-4182136" or "GSK4182136" or "UNII-1MTK0BPN8V" or "VIR-7831" or "VIR7831").tw,kw. | 9 |
| 109 | ("BRII-196" or "BRII-198" or "DZIF-10c" or "BI 767551" or "SCTA01" or "Ty027" or "HLX70").tw,kw. | 8 |
| 110 | ("C144-LS" or "C-135-LS" or "C144-LS/C-135-LS").tw,kw. | 1 |
| 111 | ("COVI-GUARD" or "STI-1499").tw,kw. | 2 |
| 112 | ("COVI-AMG" or "sti-2020").tw,kw. | 0 |
| 113 | "HFB30132A".tw,kw. | 0 |
| 114 | ("ABBV-47D11" or "ABBV-2B04" or "AZD7442").tw,kw. | 0 |
| 115 | ("BI 767551" or "DZIF-10c" or "COR-101").tw,kw. | 3 |
| 116 | *baricitinib/ | 739 |
| 117 | (baricitinib or olumiant or incb28050 or ly3009104).tw,kw. | 1183 |
| 118 | or/104-117 [MONOCLONAL ANTIBODIES] | 402826 |
| 119 | exp *antiserum/ | 8959 |
| 120 | ((immune adj3 (sera or serum? or plasma)) or antisera or antiserum or immun#serum).tw,kw. | 75643 |
| 121 | exp *immunoglobulin/ | 155050 |
| 122 | (immune* globulin? or immuneglobulin? or immunoglobulin?).tw,kw. | 207464 |
| 123 | exp *convalescent blood product/ | 466 |
| 124 | ((convalescent adj2 (plasma or serum or sera)) or (plasma adj3 therap*)).tw,kw. | 16768 |
| 125 | or/119-124 [IG THERAPY] | 387633 |
| 126 | *ibuprofen/ | 13840 |
| 127 | (ibuprofen or abfen or "aches-n-pain" or "act-3" or actiprofen or "adex 200" or adex liqui-gels or advil or afebril or aktren or aktren spezial or algiasdin or algifor or algofen or algoflex or allipen or alvofen express or "am-fam 400" or anadin or anadvil or analgyl or anbifen or anco or andran or anflagen or antalgil or antarene or antiflam or apo-ibuprofen or aragel or "atril 300" or attritin or balkaprofen or berlistar or bestafen or betaprofen or bifen or bluton or brufanic or brufedol or brufen or brufort or brugesic or brumare or brumed or brupro or buburone or bufect or bufohexal or bupogesic or burana or butacortelone or butifen or caldolor or calprofen or cap-profen or cenbufen or codral period pain or combiflam or contraneural or cuprofen or dalsy or "dc 7034" or "dc7034" or "dg 7034" or "dg7034" or dibufen or "diffutab sr 600" or dimidon or dolan fp or dolgit or dolobene ibu or dolocyl or dolodolgit or dolofen-f or dolomax or dolormin or dolval or donjust b or dorival or druisel or easifon or ecoprofen or emflam or epobron or ergix douleur et fievre or eudorlin extra or exidol or expanfen or febratic or febryn or femapirin or fenalgic or fenbid or flamicon or flarin or froben dolore or galprofen or gelufene or gyno-neuralgin or halprin or haltran or hemagene tailleur or h-loniten or ib-100 or ibalgin or ibofen or ibosure or ibu or ibuberl or ibucalm or ibudak or ibudol or ibudolor or ibufarmalid or ibufen or ibuflam or ibufug or ibugel or ibugesic or ibukern or ibuleve or ibulgan or ibuloid or ibumetin or ibumousse or ibunin or ibupen or ibupirac or ibuprin or ibuprocin or ibuprofene or ibuprohm or ibuprom or iburon or ibusal or ibuspray or ibustar or ibusynth or ibutop or ibux or ibuxin or idyl sr or ifenin or infant's motrin or infibu or inflanor or inflanor forte or ipren or irfen or junifen or junipro or kenfen or kontraneural or lamidon or librofem or lidifen or liptan or lopane or malafene or maxagesic or "mcn r 1451" or medicol or medipren or mediprin or mensoton or midol or momentact or motrin or mynosedin or nagifen-d or napacetin or neobrufen or neobrufen retard or nerofen or neutropain or nobfelon or nobgen or norflam-t or noritis or norton or novogent or novoprofen or nugin or nuprin or nureflex or nurofen or optifen or opturem or ostarin or ostofen or ozonol or paduden or panafen or pedea or pediacare fever or pediprofen or perdophen pediatrie or perofen or phorpain or phorpain gel or proartinal or profen or profeno or proff or proflex or proris or provin or provon or quadrax or rafen or ranofen or rapidophen or rapidophen forte or ratiodolor or rebugen or renidon or reuvol or rhelafen or roidenin or rufen or rupan or saridon n or schufen or seclodin or solufen lidose or solvium or spalt or syntofene or tabalon or tab-profen or taskine or tatanal or tofen or trendar or umafen or unipro or upfen or uprofen or urem or viamal febbre e dolore or zafen or zofen).tw,kw. | 24966 |
| 128 | 126 or 127 [IBUPROFEN] | 28619 |
| 129 | exp *self care/ | 30071 |
| 130 | (((patient or self) adj (manag* or treat*)) or self care).tw,kw. | 118335 |
| 131 | exp *patient attitude/ or *health literacy/ or *patient education/ | 136470 |
| 132 | (patient* adj3 (educat* or knowledg* or literacy or literate or behavior* or behaviour* or attitud* or belief* or believ*)).tw,kw. | 143953 |
| 133 | (((self help or support*) adj3 (group? or club? or servic* or social*)) or (therap* adj3 (club? or group* or social*))).tw,kw. | 172580 |
| 134 | or/129-133 [PATIENT SELF CARE] | 536640 |
| 135 | *indigenous health care/ | 329 |
| 136 | ((indigenous or metis or aboriginal or inuit or (native adj (america* or alaska* or hawaii* or canad*)) or first nation* or first people*) and (healthcare or (health adj2 (care* or service* or program*)) or healing* or medic* or therap* or remed*)).tw,kw. | 21735 |
| 137 | 135 or 136 [INDIGENOUS THERAPY] | 21895 |
| 138 | exp *alternative medicine/ or *traditional healer/ or exp *traditional medicine/ or exp *medicinal plant/ or *herbaceous agent/ or exp *plant medicinal product/ | 902707 |
| 139 | ((alternat* or complementary or folk or traditional or holistic or chinese or african or tribal) adj2 (medic* or therap* or healing* or treat* or remed*)).tw,kw. | 245894 |
| 140 | *hypnosis/ | 7799 |
| 141 | (hypnotism or hypnosis or hypnotherapy).tw,kw. | 9609 |
| 142 | exp *acupuncture/ or *ayurvedic drug/ | 34037 |
| 143 | (ayurved* or kampo or kanpo or acupunctur* or homeopath*).tw,kw. | 52233 |
| 144 | ((botanical* or herb* or plant or plants or plantlet* or root or roots or natural) adj2 (drug? or extract? or healing* or ingredient? or medic* or preparation* or product or products or remedies or remedy* or supplement* or treat* or therap*)).tw,kw. | 236789 |
| 145 | or/138-144 [ALTERNATIVE THERAPY] | 1222198 |
| 146 | exp *disease management/ | 452528 |
| 147 | ((manag* or cope or coping) adj3 (symptom* or disease* or condition* or pain or discomfort*)).tw,kw. | 192610 |
| 148 | 146 or 147 [DISEASE MANAGEMENT] | 634299 |
| 149 | exp *exercise/ or exp *sport/ or exp *physical activity/ or *fitness/ or exp *physical medicine/ | 612785 |
| 150 | ("physical activit*" or walk* or pedestrian* or bicycl* or cycling or cyclist* or biking or bike* or "active lifestyle*" or "aerobic fitness" or "aerobic exercise*" or (running not "running water") or runner* or jog* or swim* or yoga or (physical* adj2 (activit* or active or exercise*)) or ((exercise* or fitness or aerobic*) adj2 (regimen* or training or intervention* or program* or class* or course* or train* or rehab*))).tw,kw. | 650329 |
| 151 | (physiotherap* or ((physical or physio) adj1 (therap* or treat*))).tw,kw. | 90322 |
| 152 | or/149-151 [PHYSICAL THERAPY] | 1122592 |
| 153 | exp *mental health care/ or *social work/ or *psychosocial rehabilitation/ | 74241 |
| 154 | ((mental health or emotion* or psych* or wellbeing or well-being or stress* or wellness* or anxiet* or depress* or obsess* or ocd or mood* or posttrauma* or post-trauma* or ptsd or schizo* or personality disorder* or bipolar* or adhd or attention deficit* or addict*) adj3 (care or service? or support? or treat* or therap* or psychotherap* or counsel* or hotline*)).tw,kw. | 415353 |
| 155 | 153 or 154 [MENTAL HEALTH SERVICES] | 460130 |
| 156 | 45 and (48 or 51 or 54 or 60 or 71 or 90 or 103 or 118 or 125 or 128 or 134 or 137 or 145 or 148 or 152 or 155) | 6024 |
| 157 | Clinical study/ | 155914 |
| 158 | Case control study/ | 175300 |
| 159 | Family study/ | 25317 |
| 160 | Longitudinal study/ | 158436 |
| 161 | Retrospective study/ | 1106653 |
| 162 | Prospective study/ | 699966 |
| 163 | Randomized controlled trials/ | 207740 |
| 164 | 162 not 163 | 691987 |
| 165 | Cohort analysis/ | 730992 |
| 166 | (Cohort adj (study or studies)).mp. | 358844 |
| 167 | (Case control adj (study or studies)).tw. | 145610 |
| 168 | (follow up adj (study or studies)).tw. | 66583 |
| 169 | (observational adj (study or studies)).tw. | 195203 |
| 170 | (epidemiologic$ adj (study or studies)).tw. | 111912 |
| 171 | (cross sectional adj (study or studies)).tw. | 257929 |
| 172 | (prospect* adj (study or studies)).tw. | 280089 |
| 173 | Longitudinal.tw. | 364969 |
| 174 | Retrospective.tw. | 1001302 |
| 175 | method*.tw. | 10379479 |
| 176 | case series.tw. | 118483 |
| 177 | or/157-161,164-176 [Adapted from OVID observational studies filter] | 11631706 |
| 178 | Randomized Controlled Trial/ | 667737 |
| 179 | "Randomized Controlled Trial (topic)"/ | 207740 |
| 180 | Controlled Clinical Trial/ | 463491 |
| 181 | "Controlled Clinical Trial (topic)"/ | 11796 |
| 182 | Randomization/ | 91306 |
| 183 | Double Blind Procedure/ | 185912 |
| 184 | Single Blind Procedure/ | 43247 |
| 185 | placebo/ | 368850 |
| 186 | Control Group/ | 110011 |
| 187 | (random* or sham or placebo*).ti,ab,hw,kw. | 2217320 |
| 188 | ((singl* or doubl*) adj (blind* or dumm* or mask*)).ti,ab,hw,kw. | 324531 |
| 189 | ((tripl* or trebl*) adj (blind* or dumm* or mask*)).ti,ab,hw,kw. | 1600 |
| 190 | (control* adj3 (study or studies or trial* or group*)).ti,ab,kw. | 1490184 |
| 191 | (Nonrandom* or non random* or non-random* or quasi-random* or quasirandom*).ti,ab,hw,kw. | 60215 |
| 192 | allocated.ti,ab,hw. | 93247 |
| 193 | ((open label or open-label) adj5 (study or studies or trial*)).ti,ab,hw,kw. | 70852 |
| 194 | ((equivalence or superiority or non-inferiority or noninferiority) adj3 (study or studies or trial*)).ti,ab,hw,kw. | 14217 |
| 195 | (pragmatic study or pragmatic studies).ti,ab,hw,kw. | 709 |
| 196 | ((pragmatic or practical) adj3 trial*).ti,ab,hw,kw. | 6460 |
| 197 | ((quasiexperimental or quasi-experimental) adj3 (study or studies or trial*)).ti,ab,hw,kw. | 14923 |
| 198 | (phase adj3 (III or "3") adj3 (study or studies or trial*)).ti,hw,kw. | 102696 |
| 199 | or/178-198 [Adapted from "Strings attached: CADTH database search filters"] | 3296002 |
| 200 | 156 and (177 or 199) | 3869 |
| 201 | *case report/ or (case-stud* or case-report*).jw. or (case* and report*).ti. | 508029 |
| 202 | (editorial or conference abstract or conference paper or conference review).pt. | 5605998 |
| 203 | 200 not (201 or 202) | 2665 |
| 204 | limit 203 to yr="2020-Current" | 2597 |
| 205 | (202006* or 202007* or 202008* or 202009* or 20201* or 2021* or 2022*).dc,dd,dp. | 2618960 |
| 206 | 204 and 205 | 2569 |

**Excluded Studies List for Key Question 1**

**Not primary research (n=50)**

1.	Acevedo-Aguilar L, Torres-Llinas D, Gaitan-Herrera G, Lozada-Martinez I, Moscote-Salazar L, Rahman M. Letter to the Editor Regarding "Perioperative COVID-19 Incidence and Outcomes in Neurosurgical Patients at Two Tertiary Care Centers in Washington, DC, During a Pandemic: A 6-Month Follow-up". *World Neurosurg*. 2021;146:391-2.

2.	Alcazar-Navarrete B, Molina-Paris J, Martin-Sanchez F. Management and follow up of respiratory patients in the post-Covid-19 era: Are we ready yet? *Arch Bronconeumol*. 2020;56(10):685-6.

3.	Al-Jahdhami I, Al-Naamani K, Al-Mawali A. The post-acute COVID-19 syndrome (Long COVID). *Oman Med J.* 2021;36(1):1-2.

4.	Atkins D. Unraveling the Connection Between Obesity and Outcomes in COVID-19. *Obesity*. 2021;29(5):786-7.

5.	Azadnajafabad S, Ghasemi E, Moghaddam SS, Rezaei N, Farzadfar F. Non-communicable diseases' contribution to the COVID-19 mortality: A global warning on the emerging syndemics. *Arch Iran Med*. 2021;24(5):445-6.

6.	Brodsky MB, Freeman-Sanderson A, Brenner MJ. Voice, Swallow, and Airway Impairment After Late Tracheostomy: Defining Features of COVID-19 Survivorship. *Laryngoscope.* 2021;131(7):E2311.

7.	Carli G, Cecchi L, Farsi A, Stebbing J, Parronchi P. Asthma phenotypes, comorbidities, and disease activity in COVID-19: The need of risk stratification. *Allergy*. 2021;76(3):955-6.

8.	Dani M, Dirksen A, Taraborrelli P, et al. Autonomic dysfunction in 'long COVID': rationale, physiology and management strategies. *Clin Med*. 2021;21(1):E63-E7.

9.	de Erausquin GA, Snyder H, Carrillo M, et al. The chronic neuropsychiatric sequelae of COVID-19: The need for a prospective study of viral impact on brain functioning. *Alzheimers Dement.* 2021;17(6):1056-65.

10.	Delanaye P, Huart J, Jouret F, Bouquegneau A. Long-term effects of COVID-19 on kidney function. *Lancet*. 2021;397(10287):1807.

11.	Foschi M, D'Anna L, Abdelhak A, et al. Ongoing challenges in unravelling the association between COVID-19 and Guillain-Barre syndrome. *Brain.* 2021;144(5):e44.

12.	Gandotra S, Russell D. The long and the short of it: Is "long covid" more than slow resolution of the acute disease? *Ann Am Thorac Soc*. 2021;18(6):948-50.

13.	Garg P, Arora U, Kumar A, Wig N. The "post-COVID" syndrome: How deep is the damage? *J Med Virol.* 2021;93(2):673-4.

14.	Gonzalez-Herazo M, Silva-Munoz D, Guevara-Martinez P, Lozada-Martinez I. Post-COVID 19 Neurological Syndrome: a fresh challenge in neurological management. *Neurol Neurochir Pol.* 2021.

15.	Hacker KA, Briss PA, Richardson L, Wright J, Petersen R. COVID-19 and Chronic Disease: The Impact Now and in the Future. *Prev Chronic Dis.* 2021;18:E62.

16.	Jordan A, Sivapalan P, Jensen J-U. Does inhaled corticosteroid use affect the risk of covid-19-related death? *Breathe*. 2021;17(1):1-3.

17.	Kalemci S, Sarihan A, Zeybek A. Does asthma affect outcomes of patients with COVID-19 infections? *J Allergy Clin Immunol Pract.* 2021;9(1):594.

18.	Khunti K, Davies MJ, Kosiborod MN, Nauck MA. Long COVID - metabolic risk factors and novel therapeutic management. *Nat Rev Endocrinol.* 2021;17(7):379-80.

19.	Konstantinidis I, Kitsios GD, Morris A. The Impact of Acute Illness Severity on Post-COVID-19 Sequelae Remains an Unsettled Question. *Ann Am Thorac Soc.* 2021.

20.	Kutsuna S. Clinical Manifestations of Coronavirus Disease 2019. *JMA*. 2021;4(2):76-80.

21.	Ludvigsson JF. Spanish telemedicine data on 8 children support concept of 'long covid' in children. *Acta Paediatr Int J Paediatr.* 2021;110(7):2284.

22.	Lunn MP, Carr AC, Keddie S, Pakpoor J, Pipis M, Willison HJ. Reply: Guillain-Barre syndrome, SARS-CoV-2 and molecular mimicry and Ongoing challenges in unravelling the association between COVID-19 and Guillain-Barre syndrome and Unclear association between COVID-19 and Guillain-Barre syndrome and Currently available data regarding the potential association between COVID-19 and Guillain-Barre syndrome. *Brain*. ;144(5):e47.

23.	Martinez MA, Franco S. Impact of COVID-19 in Liver Disease Progression. *Hepatol Commun.* 2021;5(7):1138-50.

24.	Mathian A, Amoura Z. Response to: Patients with lupus with COVID-19: University of Michigan experience' by *Wallace et al. Ann Rheum Dis*. 2021;80(3):e36.

25.	Murugan AK, Alzahrani AS. SARS-CoV-2 plays a pivotal role in inducing hyperthyroidism of Graves' disease. *Endocrine.* 2021;73(2):243-54.

26.	Musanabaganwa C, Cubaka V, Mpabuka E, et al. One hundred thirty-three observed COVID-19 deaths in 10 months: unpacking lower than predicted mortality in Rwanda. *BMJ Glob Health*. 2021;6(2).

27.	Nagel A, Laszewska A, Simon J, Haidinger G. The first 8 weeks of the Austrian SARS-CoV-2 epidemic. *Wien Klin Wochenschr.* 2021;133(7-8):364-76.

28.	Napoli R. Diabetes and Covid-19: An interplay difficult to dissect. *Diabetes Metab Res Rev*. 2021;37(1):e3387.

29.	Oelsner EC, Allen NB, Ali T, et al. Collaborative Cohort of Cohorts for COVID-19 Research (C4R) Study: Study Design. *medRxiv.* 2021.

30.	Perico N, Perico L, Ronco C, Remuzzi G. COVID-19 and the Kidney: Should Nephrologists Care about COVID-19 rather than Maintaining Their Focus on Renal Patients? *Contrib Nephrol.* 2021;199:1-15.

31.	Rai N, Baidya DK. Severe Acute Respiratory Syndrome Coronavirus 2 (SARS-CoV-2) Pandemic: Is Sequela the Bigger Threat? *Indian J Crit Care Med*. 2021;25(2):245-6.

32.	Raw RK, Kelly C, Rees J, Wroe C, Chadwick D. Previous COVID-19 infection, but not Long-COVID, is associated with increased adverse events following BNT162b2/Pfizer vaccination. *J Infect*. 2021.

33.	Roe K. The Symptoms and Clinical Manifestations Observed in COVID-19 Patients/Long COVID-19 Symptoms that Parallel Toxoplasma gondii Infections. *J Neuroimmune Pharmacol*. 2021.

34.	Sathish T, Anton MC, Sivakumar T. New-onset diabetes in "long COVID". *J Diabetes*. 2021;13(8):693-4.

35.	Serna-Hernandez J, Benavides-Guerrero J, Navarro-Padilla L, Medina-Sanchez C, Bolano-Romero M. Letter to the editor regarding "Coexistence of neurological diseases with Covid-19 pneumonia during the pandemic period". *J Clin Neurosci*. 2021.

36.	Skevaki C, Karsonova A, Karaulov A, et al. SARS-CoV-2 infection and COVID-19 in asthmatics: a complex relationship. *Nat Rev Immunol*. 2021;21(4):202-3.

37.	Solomon JJ, Heyman B, Ko JP, Condos R, Lynch DA. CT of Postacute Lung Complications of COVID-19. *Radiology*. 2021:211396.

38.	Stewart-Patterson C, Bourgeois R, Martin DW. The Importance of Keeping Patients with Post-Acute Sequelae of SARS-CoV-2 Infection (Long COVID) Engaged in Work. *Am Fam Physician.* 2021;103(12):710.

39.	Thompson LA, Rasmussen SA. One Year Later, How Does COVID-19 Affect Children? *JAMA Pediatr.* 2021;175(2):216.

40.	Townsend L, Dowds J, O'Brien K, Martin-Loeches I, Nadarajan P, Bannan C. Reply: The Impact of Acute Illness Severity on Post-COVID-19 Sequelae Remains an Unsettled Question. *Ann Am Thorac Soc.* 2021.

41.	Vehar S, Ntiamoah P, Biehl M, Boushra M. Post-acute sequelae of SARS-CoV-2 infection: Caring for the 'long-haulers'. *Clin J Med.* 2021;88(5):267-72.

42.	Walter K. An Inside Look at a Post-COVID-19 Clinic. *JAMA*. 2021;325(20):2036-7.

43.	Wijeratne T, Crewther S. COVID-19 and long-term neurological problems: Challenges ahead with Post-COVID-19 Neurological Syndrome. *Aust J Gen Pract.* 2021;50.

44.	Wostyn P. Anosmia as a predictor for post-COVID-19 fatigue syndrome. *Lancet.* 2021;7:100162.

45.	Yelin D, Margalit I, Yahav D, Runold M, Bruchfeld J. Long COVID-19-it's not over until? *Clin Microbiol Infect.* 2021;27(4):506-8.

46.	Zhang B, Liu S, Zhang L, Zhang S, Dong Y. Previous cardiovascular surgery significantly increases the risk of developing critical illness in patients with COVID-19. *J Infect*. 2021;82(2):282-327.

47.	Zhang P. Comment on the review entitled "Risk factors for mortality in patients with Coronavirus disease 2019 (COVID-19) infection: a systematic review and meta-analysis of observational studies". *Aging Male*. 2021;23(5):1527.

48.	Zhou Y, Chi J, Lv W, Wang Y. Obesity and diabetes as high-risk factors for severe coronavirus disease 2019 (Covid-19). *Diabetes Metab Res Rev*. 2021;37(2):e3377.

49.	Zipfel S, Stengel A, Malek N, Goepel S. Long Haulers-What Is the Evidence for Post-COVID Fatigue? *Front Psychiatr*. 2021;12:677934.

50.	Zuliani G, Zuin M, Rigatelli G, Roncon L. Fatigue as long-term consequence of ARDS in COVID-19 patients. *Anaesth Crit Care Pain Med.* 2021;40(1):100787.

**Language (n=6)**

1.	Aguila Gordo D, Martinez Del Rio J, Piqueras Flores J. Changes in antihypertensive treatment in surviving patients SARS-CoV-2 respiratory infection and its cardiovascular impact after one year of follow-up. *Med Clin.* 2021.

2.	Demidova TY, Lobanova KG, Oynotkinova OS, Perekhodov SN, Antsiferov MB. Clinical and laboratory characteristics of patients with COVID-19 and concomitant type 2 diabetes. *Cardiovasc Ther Prev (Russ. Fed.).* 2021;20(1):47-58.

3.	Hernando JEC. Follow-up of patients with non-respiratory sequelae of COVID-19. *FMC Form Medica Contin*. 2021;28(2):81-9.

4.	Kolditz M, Beyer-Westendorf J, von Bonin S, Koschel DS. Persistent dyspnea after COVID-19: Suggestions for follow-up care. *MMW Fortschr Med*. 2021;163(8):52-5.

5.	Lynnyk M, Gumeniuk M, Liskina I, Gumeniuk G, Ignatieva V, Tarasenko O. [FEATURES OF THE COMPLICATED COURSE OF NON-HOSPITAL VIRAL COVID-19 PNEUMONIA]. *Georgian Med News*. 2021(315):129-35.

6.	Saldias Penafiel F, Penaloza Tapia A, Farias Nesvadba D, et al. [Clinical features and predictors of severity in 1022 adults with COVID-19]. *Rev Med Chile*. 2020;148(10):1387-97.

**No/Wrong risk factor (n=34)**

1.	Amanat M, Salehi M, Rezaei N, et al. Neurological manifestations as the predictors of severity and mortality in hospitalized individuals with COVID-19: a multicenter prospective clinical study. *BMC Neurol*. 2021;21(1):116.

2.	Barker R, Kelly PA, Gonzalez E, et al. Three-month follow-up of pulmonary embolism in patients with COVID-19. *Thromb Res*. 2021;201:113-5.

3.	Bordin A, Gaudioso P, Nicolai P, et al. Comparison of self-reported symptoms and psychophysical tests in coronavirus disease 2019 (COVID-19) subjects experiencing long-term olfactory dysfunction: a 6-month follow-up study. *Int Forum Allergy Rhinol*. 2021.

4.	Cadegiani FA, Goren A, McCoy J, Wambier CG. Early COVID-19 therapy with azithromycin plus nitazoxanide, ivermectin or hydroxychloroquine in outpatient settings significantly improved COVID-19 outcomes compared to known outcomes in untreated patients. *New Microbes and New Infect*. 2021;43:100915.

5.	Chen X, Jiang Q, Ma Z, Hu W, Cao Q, Mo P, et al. Clinical Characteristics of Hospitalized Patients with SARS-CoV-2 and Hepatitis B Virus Co-infection. *Virol Sin*. 2020;35(6):842-5.

6.	Darley DR, Dore GJ, Cysique L, et al. Persistent symptoms up to four months after community and hospital-managed SARS-CoV-2 infection. *Med J Aust.* 2021;214(6):279-80.

7.	Davis HE, Assaf GS, McCorkell L, et al. Characterizing long COVID in an international cohort: 7 months of symptoms and their impact. *EClinicalMed.* 2021:101019.

8.	Havervall S, Rosell A, Phillipson M, et al. Symptoms and Functional Impairment Assessed 8 Months After Mild COVID-19 Among Health Care Workers. *JAMA*. 2021;325(19):2015-6.

9.	Humbir A, Tiwary P, Mishra M, et al. Recovery after critical illness in COVID-19 ICU survivors. *Br J Anaesth*. 2021;126(6):e217-e9.

10.	Iftikhar H, Doherty WL, Sharp C. Long-term COVID-19 complications: A multidisciplinary clinic follow-up approach. *Clin Med.* 2021;21.

11.	Kashif A, Chaudhry M, Fayyaz T, et al. Follow-up of COVID-19 recovered patients with mild disease. *Sci Rep.* 2021;11(1):13414.

12.	Logue JK, Franko NM, McCulloch DJ, et al. Sequelae in Adults at 6 Months after COVID-19 Infection. *JAMA Netw Open.* 2021;4(2):e210830.

13.	Mandal S, Brill SE, Jarvis HC, et al. Long-COVID': A cross-sectional study of persisting symptoms, biomarker and imaging abnormalities following hospitalisation for COVID-19. *Thorax*. 2021;76(4):396-8.

14.	Mata-Vazquez E, Martin-Toledano M, Lopez-Larramona G, et al. Long-term outcomes of patients with coronavirus disease 2019 at one year after hospital discharge. *J Clin Med.* 2021;10(13):2945.

15.	McPeake J, Shaw M, MacTavish P, et al. Long-term Outcomes Following Severe COVID-19 Infection: A Multicenter Cohort Study of Family Member Outcomes. *Ann Am Thorac Soc*. 2021.

16.	Moraleda C, Serna-Pascual M, Soriano-Arandes A, et al. Multi-inflammatory Syndrome in Children Related to Severe Acute Respiratory Syndrome Coronavirus 2 (SARS-CoV-2) in Spain. *Clin Infect Dis.* 2021;72(9):e397-e401.

17.	Morin L, Savale L, Montani D, et al. Four-Month Clinical Status of a Cohort of Patients after Hospitalization for COVID-19. *JAMA.* 2021;325(15):1525-34.

18.	Oh TK, Song I-A, Park HY. Risk of psychological sequelae among coronavirus disease-2019 survivors: A nationwide cohort study in South Korea. *Depress Anxiety*. 2021;38(2):247-54.

19.	Radin JM, Quer G, Ramos E, et al. Assessment of Prolonged Physiological and Behavioral Changes Associated With COVID-19 Infection. *JAMA Netw Open*. 2021;4(7):e2115959.

20.	Remy-Jardin M, Felloni P, Faivre J-B, Remy J, Duthoit L, Fry S, et al. Assessment of pulmonary arterial circulation 3 months after hospitalization for SARS-CoV-2 pneumonia: Dual-energy CT (DECT) angiographic study in 55 patients. *EClinicalMed*. 2021;34:100778.

21.	Renaud M, Thibault C, Le Normand F, et al. Clinical Outcomes for Patients with Anosmia 1 Year After COVID-19 Diagnosis. *JAMA Netw Open*. 2021;4(6):e2115352.

22.	Romero-Duarte A, Rivera-Izquierdo M, Guerrero-Fernandez de Alba I, et al. Sequelae, persistent symptomatology and outcomes after COVID-19 hospitalization: the ANCOHVID multicentre 6-month follow-up study. *BMC Med*. 2021;19(1):129.

23.	Say D, Crawford N, McNab S, Wurzel D, Steer A, Tosif S. Post-acute COVID-19 outcomes in children with mild and asymptomatic disease. *Lancet Child Adolesc Health*. 2021;5(6):e22-e3.

24.	Schoneveld L, Ladang A, Cavalier E, et al. YKL-40 as a new promising prognostic marker of severity in COVID infection. *Crit Care*. 2021;25(1):66.

25.	Singer ME, Taub IB, Kaelber DC. Risk of Myocarditis from COVID-19 Infection in People Under Age 20: A Population-Based Analysis. *medRxiv.* 2021.

26.	Stockmann H, Hardenberg J-HB, Aigner A, et al. High rates of long-term renal recovery in survivors of coronavirus disease 2019-associated acute kidney injury requiring kidney replacement therapy. *Kidney Int*. 2021;99(4):1021-2.

27.	Strohbehn IA, Zhao S, Seethapathy H, et al. Acute kidney injury incidence, recovery, and long-term kidney outcomes among hospitalized patients with COVID-19 and influenza. *Kidney Int Rep*. 2021.

28.	Sykes DL, Holdsworth L, Jawad N, Gunasekera P, Morice AH, Crooks MG. Post-COVID-19 Symptom Burden: What is Long-COVID and How Should We Manage It? *Lung*. 2021;199(2):113-9.

29.	Taghioff SM, Slavin BR, Singh D, Holton T. Examining the potential benefits of the influenza vaccine against SARS-CoV-2: A retrospective cohort analysis of 74,754 patients. *PLoS ONE*. 2021;16(8):e0255541.

30.	Taquet M, Geddes JR, Harrison PJ, Luciano S. Bidirectional associations between COVID-19 and psychiatric disorder: retrospective cohort studies of 62 354 COVID-19 cases in the USA. *Lancet Psychiatr*. 2021;8(2):130-40.

31.	Tholin B, Ghanima W, Einvik G, et al. Incidence of thrombotic complications in hospitalised and non-hospitalised patients after COVID-19 diagnosis. *Br J Haematol*. 2021;194(3):542-6.

32.	van Gassel RJJ, Bels JLM, Raafs A, al. High prevalence of pulmonary sequelae at 3 months after hospital discharge in mechanically ventilated survivors of COVID-19. *Am J Respir Crit Care Med*. 2021;203(3):371-4.

33.	Xiong L, Cao X, Xiong H, et al. Dynamic changes of functional fitness, antibodies to SARS-CoV-2 and immunological indicators within 1 year after discharge in Chinese health care workers with severe COVID-19: a cohort study. *BMC Med*. 2021;19(1):163.

34.	Zhu F, Li X. Lactate-dehydrogenase associated with mortality in hospitalized patients with COVID-19 in Mexico. *Ann Hepatol*. 2021;24:100348.

**No outcome of interest (n=72)**

1.	Achiron A, Gurevich M, Falb R, Dreyer-Alster S, Sonis P, Mandel M. SARS-CoV-2 antibody dynamics and B-cell memory response over time in COVID-19 convalescent subjects. *Clin Microbiol Infect*. 2021.

2.	Alroy-Preis S, Haas EJ, Angulo FJ, et al. Impact and effectiveness of mRNA BNT162b2 vaccine against SARS-CoV-2 infections and COVID-19 cases, hospitalisations, and deaths following a nationwide vaccination campaign in Israel: an observational study using national surveillance data. *Lancet*. 2021;397(10287):1819-29.

3.	Amjadi MF, O'Connell SE, Armbrust T, et al. Fever, Diarrhea, and Severe Disease Correlate with High Persistent Antibody Levels against SARS-CoV-2. *medRxiv*. 2021.

4.	An J, Liao X, Wang H, et al. Distinct kinetics of immunoglobulin isotypes reveal early diagnosis and disease severity of COVID-19: A 6-month follow-up. *Clin Transl Med*. 2021;11(3):e342.

5.	Banham GD, Godlee A, Faustini SE, Cunningham AF, Richter A, Harper L, et al. Hemodialysis Patients Make Long-Lived Antibodies against SARS-CoV-2 that May Be Associated with Reduced Reinfection. *J Am Soc Nephrol.* 2021.

6.	Batty GD, Deary IJ, Gale CR. Pre-pandemic cognitive function and COVID-19 mortality: prospective cohort study. *Eur J Epidemiol*. 2021;36(5):559-64.

7.	Bhaskaran K, Rentsch CT, Schultze A, et al. HIV infection and COVID-19 death: a population-based cohort analysis of UK primary care data and linked national death registrations within the OpenSAFELY platform. *Lancet HIV*. 2021;8(1):e24-e32.

8.	Bhatta S, Sharma S, Sharma D, et al. Study of Hearing Status in COVID-19 Patients: A Multicentered Review. *Indian J Otolaryngol Head Neck Surg*. 2021:1-7.

9.	Broche-Perez Y, Medina-Navarro C. Neuropsychological & cognitive sequelae in COVID-19 patients. *MEDICC Rev.* 2021;23(2):78-9.

10.	Brojakowska A, Eskandari A, Bisserier M, et al. Comorbidities, sequelae, blood biomarkers and their associated clinical outcomes in the Mount Sinai Health System COVID-19 patients. *PLoS ONE*. 2021;16(7):e0253660.

11.	Chemaitelly H, Abu-Raddad L, Coyle P, et al. SARS-CoV-2 antibody-positivity protects against reinfection for at least seven months with 95% efficacy. *EClinicalMed.* 2021;35:100861.

12.	Cifuentes MP, Rojas-Botero M, Rodriguez-Villamizar L, Alvarez-Moreno C, Fernandez-Nino J. Socioeconomic inequalities associated with mortality for COVID-19 in Colombia: a cohort nationwide study. *J Epidemiol Community Health.* 2021.

13.	Cotter MP, Houston AC, Habibi MS, Breathnach AS, Riley PA, Planche TD. Prior COVID-19 significantly reduces the risk of subsequent infection, but reinfections are seen after eight months. *J Infect*. 2021;82(4):e11-e2.

14.	Dai M, Tao L, Chen Z, et al. Influence of Cigarettes and Alcohol on the Severity and Death of COVID-19: A Multicenter Retrospective Study in Wuhan, China. *Front Physiol.* 2020;11:588553.

15.	den Hartog G, Schepp RM, Vos ERA, et al. Persistence of antibodies to SARS-CoV-2 in relation to symptoms in a nationwide prospective study. *Clin Infect Dis.* 2021.

16.	Ekstrom S, Andersson N, Lovquist A, et al. COVID-19 among young adults in Sweden: self-reported long-term symptoms and associated factors. *Scand J Public Health*. 2021:14034948211025425.

17.	Espasandin-Dominguez J, Gude-Sampedro F, Fernandez-Merino C, et al. Development and validation of a prognostic model based on comorbidities to predict COVID-19 severity: A population-based study. *Int J Epidemiol.* 2021;50(1):64-74.

18.	Estiri H, Klann JG, Wagholikar KB, Strasser ZH, Naseri P, Murphy SN. Predicting COVID-19 mortality with electronic medical records. *npj Dig Med*. 2021;4(1):15.

19.	Estiri H, Strasser ZH, Brat GA, et al. Evolving Phenotypes of non-hospitalized Patients that Indicate Long Covid. *medRxiv.* 2021.

20.	Fernandez-de-las-Penas C, Guijarro C, Plaza-Canteli S, Hernandez-Barrera V, Torres-Macho J. Prevalence of Post-COVID-19 Cough One Year After SARS-CoV-2 Infection: A Multicenter Study. *Lung*. 2021;199(3):249-53.

21.	Foulkes S, Atti A, Monk EJM, et al. SARS-CoV-2 infection rates of antibody-positive compared with antibody-negative health-care workers in England: a large, multicentre, prospective cohort study (SIREN). *Lancet*. 2021;397(10283):1459-69.

22.	Frey S, Gugenheim J, Iannelli A, et al. The Impact of Previous History of Bariatric Surgery on Outcome of COVID-19. A Nationwide Medico-Administrative French Study. *Obes Surg*. 2021;31(4):1455-63.

23.	Gerkin RC, Ohla K, Veldhuizen MG, et al. Recent smell loss is the best predictor of COVID-19 among individuals with recent respiratory symptoms. *Chem Senses*. 2021;46.

24.	Gist RE, Pinto R, Ahmed YE, Kissoon N, Daniel P, Hamele M. Repurposing a PICU for Adult Care in a State Mandated COVID-19 Only Hospital: Outcome Comparison to the MICU Cohort to Determine Safety and Effectiveness. *Front Pediatr*. 2021;9:665350.

25.	Graham MS, Antonelli M, Murray B, et al. Changes in symptomatology, reinfection, and transmissibility associated with the SARS-CoV-2 variant B.1.1.7: an ecological study. *Lancet Public Health.* 2021;6(5):e335-e45.

26.	Hashim MJ, Alsuwaidi AR, Khan G. Population risk factors for COVID-19 mortality in 93 countries. *J Epidemiol Glob Health.* 2020;10(3):204-8.

27.	Hopkins C, Surda P, Safarian M, et al. Six month follow-up of self-reported loss of smell during the COVID-19 pandemic. *Rhinology.* 2021;59(1):26-31.

28.	Husain Q, Houston S, Shargorodsky J, Kokinakos K, Kuo Y-H, Zaidi F. Characteristics of COVID-19 smell and taste dysfunction in hospitalized patients. *Am J Otolaryngol.* 2021;42(6):103068.

29.	Jon R, Veronica F, Anne R, Christopher B, Kenneth C. Incidence of COVID-19 recurrence among large cohort of healthcare employees. *Ann Epidemiol*. 2021;60:8-14.

30.	Journe F, Circiu MP, Hans S, et al. Prevalence and 6-month recovery of olfactory dysfunction: a multicentre study of 1363 COVID-19 patients. *J Int Med.* 2021;290(2):451-61.

31.	Karaismailoglu E, Karaismailoglu S. Two novel nomograms for predicting the risk of hospitalization or mortality due to COVID-19 by the naive Bayesian classifier method. *J Med Virol*. 2021;93(5):3194-201.

32.	Kim S, Jung C-G, Lee JY, et al. Characterization of asthma and risk factors for delayed SARS-CoV-2 clearance in adult COVID-19 inpatients in Daegu. *Allergy.* 2021;76(3):918-21.

33.	Kons ZA, Coelho DH, Reiter ER, Costanzo RM, Budd SG, Shin Y. Quality of life and safety impact of COVID-19 associated smell and taste disturbances. *Am J Otolaryngol.* 2021;42(4):103001.

34.	Lebreton X, Chavarot N, Scemla A, et al. Decline and loss of anti-SARS-CoV-2 antibodies in kidney transplant recipients in the 6 months following SARS-CoV-2 infection. *Kidney Int*. 2021;99(2):486-8.

35.	Lu Q-B, Jiang W-L, Zhang X, et al. Comorbidities for fatal outcome among the COVID-19 patients: A hospital-based case-control study. *J Infect*. 2021;82(1):159-98.

36.	Macias-Munoz L, Wijngaard R, Gonzalez-de la Presa B, Bedini JL, Morales-Ruiz M, Jimenez W. Value of clinical laboratory test for early prediction of mortality in patients with COVID-19: the BGM score. *J Circ Biomark*. 2021;10:1-8.

37.	Maniscalco M, Fuschillo S, Ambrosino P, et al. Bronchodilator Response as a Possible Predictor of Lung Function Improvement After Pulmonary Rehabilitation in Post-COVID-19 Patients. *Arch Bronconeumol*. 2021.

38.	Masia M, Padilla S, Fernandez-Gonzalez M, Gutierrez F, Galiana A. Incidence of delayed asymptomatic COVID-19 recurrences in a 6-month longitudinal study. *J Infect*. 2021;82(6):276-316.

39.	Melenotte C, Drancourt M, Amrane S, et al. SARS-CoV-2 persistent viral shedding in the context of hydroxychloroquine-azithromycin treatment. *Viruses*. 2021;13(5):890.

40.	Meyerholz DK, Perlman S. Does common cold coronavirus infection protect against severe SARS-CoV-2 disease? *J Clin Invest.* 2021;131(1):e144807.

41.	Padilla-Raygoza N, Sandoval-Salazar C, Ramirez-Gomez X, et al. Status of novel coronavirus disease and analysis of mortality in Mexico, until June 30th, 2020: An ecological study. *Biomed Pharmacol J.* 2020;13(4):1781-90.

42.	Petrocelli M, Cutrupi S, Salzano G, et al. Six-month smell and taste recovery rates in coronavirus disease 2019 patients: A prospective psychophysical study. *J Laryngol Otol*. 2021;135(5):436-41.

43.	Pizzonia KL, Koscinski B, Suhr JA, Accorso C, Allan DM, Allan NP. Insomnia during the COVID-19 pandemic: the role of depression and COVID-19-related risk factors. *Cogn Behav Ther*. 2021;50(3):246-60.

44.	Pourhomayoun M, Shakibi M. Predicting mortality risk in patients with COVID-19 using machine learning to help medical decision-making. *Smart Health*. 2021;20:100178.

45.	Pourhoseingholi MA, Jafari R, Jafari NJ, et al. Predicting 1-year post-COVID-19 mortality based on chest computed tomography scan. *J Med Virol.* 2021.

46.	Qureshi AI, Huang W, Lobanova I, Baskett WI, Naqvi SH, Shyu C-R. Re-infection with SARS-CoV-2 in Patients Undergoing Serial Laboratory Testing. *Clin Infect Dis*. 2021.

47.	Raad RA, Papagiannopoulos P, LoSavio P, et al. Temporal patterns of nasal symptoms in patients with mild severity SARS-CoV-2 infection. *Am J Otolaryngol*. 2021;42(6):103076.

48.	Ramirez-Aldana R, Gomez-Verjan J, Bello-Chavolla O, Garcia-Pena C. Spatial epidemiological study of the distribution, clustering, and risk factors associated with early COVID-19 mortality in Mexico. *PloS ONE*. 2021;16(7):e0254884.

49.	Riestra-Ayora J, Yanes-Diaz J, Esteban-Sanchez J, et al. Long-term follow-up of olfactory and gustatory dysfunction in COVID-19: 6 months case-control study of health workers. *Eur Arch Otorhinolaryngol.* 2021.

50.	Salgado-Aranda R, Nunez-Gil I, Orozco AJ, et al. Influence of Baseline Physical Activity as a Modifying Factor on COVID-19 Mortality: A Single-Center, Retrospective Study. *Infect Dis Ther.* 2021;10(2):801-14.

51.	Sami R, Soltaninejad F, Amra B, et al. A one-year hospital-based prospective COVID- 19 open-cohort in the Eastern Mediterranean region: The Khorshid COVID Cohort (KCC) study. *PLoS ONE.* 2020;15(11):e0241537.

52.	Sands KE, McLean LE, Korwek KM, et al. Patient characteristics and admitting vital signs associated with coronavirus disease 2019 (COVID-19)-related mortality among patients admitted with noncritical illness. *Infect Control Hosp Epidemiol*. 2021;42(4):399-405.

53.	Seghers F, Perlot Q, Desmet C, et al. A longitudinal, 3-month serologic assessment of sars-cov-2 infections in a belgian hemodialysis facility. *Clin J Am Soc Nephrol.* 2021;16(4):613-4.

54.	Sheehan MM, Reddy AJ, Rothberg MB. Reinfection Rates among Patients who Previously Tested Positive for COVID-19: a Retrospective Cohort Study. *Clin Infect Dis.* 2021.

55.	Silva PV, Oliveira SB, Escalante JJC, et al. Risk Factors for Death Among 120,804 Hospitalized Patients with Confirmed COVID-19 in Sao Paulo, Brazil. *Am J Trop Med.* 2021.

56.	Silveira EC. Chronic neurological disease as a independent risk factor for death in severe covid-19 cases. *Rom J Neurol.* 2021;20(2):224-7.

57.	Strohbehn IA, Zhao S, Seethapathy H, et al. Acute kidney injury incidence, recovery, and long-term kidney outcomes among hospitalized patients with COVID-19 and influenza. *Kidney Int Rep*. 2021.

58.	Sun LL, Wang J, Hu PF, et al. Symptomatic features and prognosis of 932 hospitalized patients with coronavirus disease 2019 in Wuhan. *J Dig Dis*. 2021;22(5):271-81.

59.	Tabrizi S, Trippa L, Cagney D, et al. Assessment of Simulated SARS-CoV-2 Infection and Mortality Risk Associated With Radiation Therapy Among Patients in 8 Randomized Clinical Trials. *JAMA Netw Open*. 2021;4(3):e213304.

60.	Tandan M, Acharya Y, Pokharel S, Timilsina M. Discovering symptom patterns of COVID-19 patients using association rule mining. *Comput Biol Med*. 2021;131:104249.

61.	Thomason ME, Hendrix CL, Werchan D, Brito NH. Social determinants of health exacerbate disparities in COVID-19 illness severity and lasting symptom complaints. *medRxiv*. 2021.

62.	Topless RK, Phipps-Green A, Dalbeth N, et al. Gout, Rheumatoid Arthritis, and the Risk of Death Related to Coronavirus Disease 2019: An Analysis of the UK Biobank. *ACR Open Rheumatol.* 2021;3(5):333-40.

63.	Trivedi C, Khan N, Mahmud N, Reinisch W, Lewis JD. Risk factors for SARS-CoV-2 infection and course of COVID-19 disease in patients with IBD in the Veterans Affair Healthcare System. *Gut.* 2021.

64.	Veronica F, Anne R, Christopher B, Kenneth C, Jon R. Incidence of COVID-19 recurrence among large cohort of healthcare employees. *Ann Epidemiol*. 2021;60:8-14.

65.	Walker AJ, MacKenna B, Inglesby P, et al. Clinical coding of long COVID in English primary care: a federated analysis of 58 million patient records in situ using OpenSAFELY. *Br J Gen Pract*. 2021.

66.	Wang H, Yuan Y, Xiao M, et al. Dynamics of the SARS-CoV-2 antibody response up to 10 months after infection. *Cell Molec Microbiol*. 2021;18(7):1832-4.

67.	Wang QQ, Kaelber DC, Xu R, Volkow ND. COVID-19 risk and outcomes in patients with substance use disorders: analyses from electronic health records in the United States. *Molec Psychiatr*. 2021;26(1):30-9.

68.	Wang Q, Xu R, Volkow ND. Increased risk of COVID-19 infection and mortality in people with mental disorders: analysis from electronic health records in the United States. *World Psychiatr*. 2021;20(1):124-30.

69.	Wilkins JT, Hirschhorn LR, Gray EL, et al. Serologic Status and SARS CoV-2 Infection over 6-Months of Follow-Up in Healthcare Workers in Chicago: A Cohort Study. *Infect Control Hosp Epidemiol*. 2021:1-29.

70.	Zare F, Teimouri M, Khosravi A, et al. COVID-19 re-infection in Shahroud, Iran: a follow-up study. *Epidemiol Infect*. 2021;149:e159.

71.	Zarif A, Joy M, Sherlock J, et al. The impact of primary care supported shielding on the risk of mortality in people vulnerable to COVID-19: English sentinel network matched cohort study. *J Infect*. 2021;83(2):228-36.

72.	Zhang D, Zhang C, Zhao J, et al. Thin-section computed tomography findings and longitudinal variations of the residual pulmonary sequelae after discharge in patients with COVID-19: a short-term follow-up study. *Eur Radiol.* 2021.

**Population not post-COVID (n=241)**

1.	Abu Esba LC, Alqahtani RA, Alswaidan L, Mardawi G, Thomas A, Shamas N. Ibuprofen and NSAID Use in COVID-19 Infected Patients Is Not Associated with Worse Outcomes: A Prospective Cohort Study. *Infect Dis Ther.* 2021;10(1):253-68.

2.	Achonu C, Lee B, Whelan M, et al. Epidemiology, clinical characteristics, household transmission, and lethality of severe acute respiratory syndrome coronavirus-2 infection among healthcare workers in Ontario, Canada. *PLoS ONE*. 2020;15(12):e0244477.

3.	Aguilar RB, McGoohan J, Osorio M, et al. Managed Care COVID-19 Outcomes in a Population Health Program. *Am J Manag Care*. 2021;27(6).

4.	Ahrenfeldt LJ, Nielsen CR, Lindahl-Jacobsen R, Moller S, Christensen K. Burden and prevalence of risk factors for severe COVID-19 in the ageing European population - a SHARE-based analysis. *J Public Health.* 2021.

5.	Ak C, Sayar S, Ozdil K, Polat ZP, Kilic ET. Clinical and laboratory factors associated with severe disease course in Turkish patients with COVID-19 infection. *Iran Red Screscent Med J*. 2021;23(2):e283.

6.	Akter F, Mannan A, Salauddin A, et al. Clinical characteristics and short term outcomes after recovery from COVID-19 in patients with and without diabetes in Bangladesh. *Diabetes Metab Syndr Clin Res Rev*. 2020;14(6):2031-8.

7.	Alali AS, Alshehri AO, Alshammari MA, et al. Demographics, comorbidities, and outcomes among young and middle-aged COVID-19 patients in Saudi Arabia. *Saudi Pharm J*. 2021.

8.	Al-Aly Z, Xie Y, Bowe B. High-dimensional characterization of post-acute sequelae of COVID-19. *Nature*. 2021;594(7862):259-64.

9.	Almubark R, Memish Z, Tamim H, et al. Natural history and clinical course of symptomatic and asymptomatic COVID-19 patients in the Kingdom of Saudi Arabia. *Saudi J Med Med Sci*. 2021;9(2):118-24.

10.	Al-Salameh A, Desailloud R, Lalau J-D, et al. Characteristics and outcomes of COVID-19 in hospitalized patients with and without diabetes. *Diabetes Metab Res Rev*. 2021;37(3):e3388.

11.	Alser O, Mokhtari A, Naar L, et al. Multisystem outcomes and predictors of mortality in critically ill patients with COVID-19: Demographics and disease acuity matter more than comorbidities or treatment modalities. *J Trauma Acute Care Surg*. 2021;90(5):880-90.

12.	Alwafi H, Naser AY, Qanash S, et al. Predictors of length of hospital stay, mortality, and outcomes among hospitalised COVID-19 patients in Saudi Arabia: A cross-sectional study. *J Multidiscip Healthc*. 2021;14:839-52.

13.	Amin S, Rahim F, Noor M, Bahadur S, Mahmood A, Gul H. The effect of tocilizumab on inflammatory markers in survivors and non-survivors of severe COVID-19. *J Coll Physicians Surg Pak*. 2021;31:S7-S10.

14.	Aminian A, Bena J, Pantalone KM, Burguera B. Association of obesity with postacute sequelae of COVID-19. *Diabetes Obes Metab*. 2021.

15.	Andreano A, Murtas R, Tunesi S, Gervasi F, Magnoni P, Russo AG. Sviluppo di un modello predittivo del rischio di decesso sulla base delle comorbidita in una coorte di 18.286 casi confermati di COVID-19 con almeno 40 anni d'eta, Development of a multivariable model predicting mortality risk from comorbidities in an Italian cohort of 18,286 confirmed COVID-19 cases aged 40 years or older. *Epidemiol Prev.* 2021;45(1-2):100-9.

16.	Apea VJ, Dhairyawan R, Orkin CM, et al. Ethnicity and outcomes in patients hospitalised with COVID-19 infection in East London: An observational cohort study. *BMJ Open.* 2021;11(1):e042140.

17.	Arman A, Tajik M, Nazemipour M, et al. Risk factors of developing critical conditions in Iranian patients with COVID-19. *Global Epidemiol.* 2021;3:100046.

18.	Aslaner H, Aslaner HA, Gokcek MB, Benli AR, Yildiz O. The Effect of Chronic Diseases, Age and Gender on Morbidity and Mortality of COVID-19 Infection. *Iran J Public Health*. 2021;50(4):721-7.

19.	Atalla E, Kalligeros M, Giampaolo G, Mylona EK, Shehadeh F, Mylonakis E. Readmissions among patients with COVID-19. *Int J Clin Pract*. 2021;75(3):e13700.

20.	Azad MH, Mousavi GS, Khorrami F, et al. Clinical and epidemiological characteristics of hospitalized COVID-19 patients in Hormozgan, Iran: A retrospective, multicenter study. *Arch Iran Med*. 2021;24(5):434-44.

21.	Bahadorimonfared A, Sohrabi M-R, Amin R, et al. Sociodemographic determinants and clinical risk factors associated with COVID-19 severity: a cross-sectional analysis of over 200,000 patients in Tehran, Iran. *BMC Infect Dis.* 2021;21(1):474.

22.	Baigorria F, Castilla J, Guevara M, et al. Risk factors of infection, hospitalization and death from SARS-CoV-2: A population-based cohort study. *J Clin Med*. 2021;10(12):2608.

23.	Baillargeon J, Polychronopoulou E, Kuo Y-F, Raji MA. The impact of substance use disorder on COVID-19 outcomes. *Psychiatr Serv.* 2021;72(5):578-81.

24.	Battagin T, Zaramella B, Leal FE, et al. Post-infection depressive, anxiety and post-traumatic stress symptoms: A prospective cohort study in patients with mild COVID-19. *Prog Neuropsychopharmacol Biol Psychiatry*. 2021;111:110341.

25.	Becerra-Munoz V, Gomez-Doblas J, Nunez-Gil I, et al. Clinical profile and predictors of in-hospital mortality among older patients hospitalised for COVID-19. *Age Ageing*. 2021;50(2):326-34.

26.	Bell ML, Catalfamo CJ, Farland LV, et al. Post-acute sequelae of COVID-19 in a non-hospitalized cohort: Results from the Arizona CoVHORT. *PLoS ONE*. 2021;16(8):e0254347.

27.	Bello-Chavolla O, Antonio-Villa N, Vargas-Vazquez A, et al. Validation and repurposing of the MSLCOVID- 19 score for prediction of severe COVID-19 using simple clinical predictors in a triage setting: The Nutri-CoV score. *PLoS ONE*. 2020;15(12):e0244051.

28.	Bello-Chavolla O, Bahena-Lopez J, Antonio-Villa N, et al. Predicting Mortality Due to SARS-CoV-2: A Mechanistic Score Relating Obesity and Diabetes to COVID-19 Outcomes in Mexico. *J Clin Endocrinol Metab.* 2020;105(8).

29.	Bennett KE, Mullooly M, O'Loughlin M, et al. Underlying conditions and risk of hospitalisation, ICU admission and mortality among those with COVID-19 in Ireland: A national surveillance study. *Lancet Reg healthEurope*. 2021;5:100097.

30.	Bergman J, Nordstrom P, Ballin M, Nordstrom A. Risk factors for COVID-19 diagnosis, hospitalization, and subsequent all-cause mortality in Sweden: a nationwide study. *Eur J Epidemiol*. 2021;36(3):287-98.

31.	Berkowitsch A, Cremer S, Zeiher AM, et al. Elevated markers of thrombo-inflammatory activation predict outcome in patients with cardiovascular comorbidities and COVID-19 disease: insights from the LEOSS registry. *Clin Res Cardiol*. 2021;110(7):1029-40.

32.	Bertaina M, Nunez-Gil I, Fernandez-Ortiz A, et al. Non-invasive ventilation for SARS-CoV-2 acute respiratory failure: A subanalysis from the HOPE COVID-19 registry. *Emerg J Med.* 2021;38(5):359-65.

33.	Bigelow BF, Zenilman JM, Katz MJ, et al. Outcomes of Nursing Home COVID-19 Patients by Initial Symptoms and Comorbidity: Results of Universal Testing of 1970 Residents. *J Am Med Dir Assoc.* 2020;21(12):1767.

34.	Bongiovanni M, Marra AM, De Lauretis A, et al. Clinical characteristics and outcome of COVID-19 pneumonia in elderly subjects. *J Infect*. 2021;82(2):e33-e4.

35.	Bowles KH, McDonald M, Barron Y, Kennedy E, O'Connor M, Mikkelsen M. Surviving COVID-19 After Hospital Discharge: Symptom, Functional, and Adverse Outcomes of Home Health Recipients. *Ann Intern Med*. 2021;174(3):316-25.

36.	Broberg M, Ruotsalainen S, Strausz S, et al. Sleep apnoea is a risk factor for severe COVID-19. *BMJ Open Respir Res*. 2021;8(1):000845.

37.	Calmes D, Graff S, Frix A-N, et al. Asthma and COPD Are Not Risk Factors for ICU Stay and Death in Case of SARS-CoV2 Infection. *J Allergy Clin Immunol*. 2021;9(1):160-9.

38.	Campos-Murguia A, Roman-Calleja B, Gonzalez-Regueiro J, et al. Liver fibrosis in patients with metabolic associated fatty liver disease is a risk factor for adverse outcomes in COVID-19. *Dig Liver Dis*. 2021;53(5):525-33.

39.	Candilio L, Weeraman D, Vlachou M, et al. Pulmonary thrombosis in Covid-19: before, during and after hospital admission. *J Thromb Thrombolysis*. 2021;51(4):978-84.

40.	Cantenys-Molina S, Francos P, Fernandez-Cruz E, Gil-Herrera J, Lopez Bernaldo de Quiros JC, Munoz P. Lymphocyte subsets early predict mortality in a large series of hospitalized COVID-19 patients in Spain. *Clin Exp Immunol*. 2021;203(3):424-32.

41.	Carlson N, Nelveg-Kristensen K, Freese Ballegaard E, et al. Increased vulnerability to COVID-19 in chronic kidney disease. *J Intern Med*. 2021;290(1):166-78.

42.	Carroll F, Dunn G, Fremeaux A, et al. Maternal and perinatal outcomes of pregnant women with SARS-CoV-2 infection at the time of birth in England: national cohort study. *Am J Obstet Gynecol*. 2021.

43.	Castelino L, Murphy M, Parker K, et al. Chronic anticoagulation is not associated with a reduced risk of acute kidney injury in hospitalised Covid-19 patients. *BMC Nephrol*. 2021;22(1):224.

44.	Castilla J, Guevara M, Miqueleiz A, et al. Risk Factors of Infection, Hospitalization and Death from SARS-CoV-2: A Population-Based Cohort Study. *J Clin Med.* 2021;10(12).

45.	Cereda A, Toselli M, Giannini F, et al. Is pleural effusion in COVID-19 interstitial pneumonia related to in-hospital mortality? *Ital J Med.* 2021;15(1):56-8.

46.	Chan KW, Hung IF-N, Tsang OT-Y, et al. Mass Screening Is Associated with Low Rates of Acute Kidney Injury among COVID-19 Patients in Hong Kong. *Am J Nephrol*. 2021;52(2):161-72.

47.	Chen B, Zhu S, Gao Q, et al. Prevalence and risk factors of disability and anxiety in a retrospective cohort of 432 survivors of Coronavirus Disease-2019 (Covid-19) from China. *PLoS ONE*. 2020;15(12):e0243883.

48.	Chen B, Lu C, Luo X, et al. Serum Uric Acid Concentrations and Risk of Adverse Outcomes in Patients With COVID-19. *Front Endocrinol.* 2021;12:633767.

49.	Chen K, Lei Y, He Y, et al. Clinical outcomes of hospitalized COVID-19 patients with renal injury: a multi-hospital observational study from Wuhan. *Sci Rep*. 2021;11(1):15205.

50.	Cheng D, Skyllberg E, Ainley A, Calderwood C. Clinical characteristics and outcomes of adult patients admitted with COVID-19 in East London: A retrospective cohort analysis. *BMJ Open Resp Res.* 2021;8(1):e000813.

51.	Cheng K, Li H, Xu J, Xiong W, Zhou X, Zheng J. Diagnosis and treatment of 471 patients with 2019 novel coronavirus disease (COVID-19). *Ann Transl Med.* 2021;9(2):163.

52.	Cho KH, Park JW, Do JY, Kang SH, Kim SW. Effect of sex on clinical outcomes in patients with coronavirus disease: A population-based study. *J Clin Med*. 2021;10(1):1-12.

53.	Cho SI, Kim YE, Jo SJ. Association of COVID-19 with skin diseases and relevant biologics: a cross-sectional study using nationwide claim data in South Korea*. *Br J Dermatol*. 2021;184(2):296-303.

54.	Choi H, Lee H, Lee S-K, et al. Impact of bronchiectasis on susceptibility to and severity of COVID-19: a nationwide cohort study. *Ther Adv Respir Dis.* 2021;15.

55.	Choi YJ, Suh J, Song JY, et al. Variable effects of underlying diseases on the prognosis of patients with COVID-19. *PLoS ONE.* 2021;16(7):e0254258.

56.	Choubey A, Sagar D, Cawley P, Miller K. Retrospective review analysis of COVID-19 patients co-infected with Mycoplasma pneumoniae. *Lung India*. 2021;38:S22-S6.

57.	Chua GT, Rosa Duque JS, et al. Clinical Characteristics and Transmission of COVID-19 in Children and Youths during 3 Waves of Outbreaks in Hong Kong. *JAMA Netw Open*. 2021.

58.	Clark JR, Batra A, Shlobin NA, et al. Acute-care hospital reencounters in COVID-19 patients. *GeroScience*. 2021.

59.	Cui N, Zhao J, Yan R, Qin C. Clinical Characteristics and Immune Responses of 137 Deceased Patients With COVID-19: A Retrospective Study. *Front Cell Infect Microbiol*. 2020;10:595333.

60.	da Cal MS, Monnerat LB, Litrento PF, et al. One-month outcomes of patients with SARS-CoV-2 infection and their relationships with lung ultrasound signs. *Ultrasound J*. 2021;13(1):19.

61.	da Silva PV, Tsuha DH, Cortez Escalante JJ, et al. Risk factors for death among 120,804 hospitalized patients with confirmed COVID-19 in Sao Paulo, Brazil. *Am J Trop Med*. 2021;105(1):88-92.

62.	Dai SP, Wu J-h, Zhao X. Effects of Comorbidities on the Elderly Patients with COVID-19: Clinical Characteristics of Elderly Patients Infected with COVID-19 from Sichuan, China. *J Nutr Health Aging.* 2021;25(1):18-24.

63.	Daugherty SE, Guo Y, Dasmarinas MC, et al. Risk of clinical sequelae after the acute phase of SARS-CoV-2 infection: Retrospective cohort study. *BMJ*. 2021;373:n1098.

64.	de la Cruz-Benito B, Rivas-Pollmar M, Alvarez Roman MT, et al. Paradoxical effect of SARS-CoV-2 infection in patients with immune thrombocytopenia. *Br J Haematol.* 2021;192(6):973-7.

65.	Deng H, Ye B, Zhao H, Liang J, Ke L, Li W. Association between an increase in blood urea nitrogen at 24 h and worse outcomes in COVID-19 pneumonia. *Ren Fail.* 2021;43(1):347-50.

66.	Di Castelnuovo A, Gialluisi A, Bonaccio M, et al. Disentangling the association of hydroxychloroquine treatment with mortality in covid-19 hospitalized patients through hierarchical clustering. *J Healthc Eng.* 2021;2021:5556207.

67.	Di Mascio D, Vena F, Giancotti A, et al. Risk factors associated with adverse fetal outcomes in pregnancies affected by Coronavirus disease 2019 (COVID-19): A secondary analysis of the WAPM study on COVID-19. *J Perinat Med.* 2020;48(9):950-8.

68.	Doganay F, Elkonca F, Seyhan AU, Yilmaz E, Ak R, Batirel A. Shock index as a predictor of mortality among the Covid-19 patients. *Am J Emerg Med*. 2021;40:106-9.

69.	Dres M, Hajage D, Lebbah S, et al. Characteristics, management, and prognosis of elderly patients with COVID-19 admitted in the ICU during the first wave: insights from the COVID-ICU study : Prognosis of COVID-19 elderly critically ill patients in the ICU. *Ann Intensive Care*. 2021;11(1):77.

70.	Feingold D, Tzur Bitan D, Krieger I, et al. COVID-19 Prevalence and Mortality Among Schizophrenia Patients: A Large-Scale Retrospective Cohort Study. *Schizophr Bull*. 2021.

71.	Ferdenzi C, Bousquet C, Aguera P-E, et al. Recovery From COVID-19-Related Olfactory Disorders and Quality of Life: Insights From an Observational Online Study. *Chem Senses*. 2021;46.

72.	Fernandez-de-las-Penas C, Palacios-Cena D, Rodriuez-Jimenez J, et al. Long-term post-COVID symptoms and associated risk factors in previously hospitalized patients: A multicenter study. *J Infect.* 2021;83(2):237-79.

73.	Ferro D, Violi F, Pignatelli P, Ceccarelli G, et al. Arterial and venous thrombosis in coronavirus 2019 disease (Covid-19): relationship with mortality. *Intern Emerg Med*. 2021.

74.	Foglesong A, Michel A, Gill I, et al. Epidemiology, clinical characteristics, and outcomes of a large cohort of covid-19 outpatients in Michigan. *Int J Gen Med*. 2021;14:1555-63.

75.	Forsblom E, Kortela E, Jarvinen A, et al. Male predominance in disease severity and mortality in a low Covid-19 epidemic and low case-fatality area-a population-based registry study. *Infect Dis*. 2021.

76.	Gamba P, Zaniboni A. Smell and taste in CoViD-19 patients: The forgotten sense. *Recenti Progressi Med.* 2020;111(10):614-8.

77.	Garazzino S, Denina M, Tovo P-A, et al. Epidemiology, Clinical Features and Prognostic Factors of Pediatric SARS-CoV-2 Infection: Results From an Italian Multicenter Study. *Front Pediatr.* 2021;9:649358.

78.	George CE, Inbaraj LR, Chandrasingh S, de Witte LP. High seroprevalence of COVID-19 infection in a large slum in South India; what does it tell us about managing a pandemic and beyond? *Epidemiol Infect*. 2021;149:e39.

79.	Geriatric Medicine RC, Collaborative C, Welch C. Age and frailty are independently associated with increased COVID-19 mortality and increased care needs in survivors: results of an international multi-centre study. *Age Ageing.* 2021;50(3):617-30.

80.	Giannis D, Kohn N, Barish MA, et al. Incidence of Venous Thromboembolism and Mortality in Patients with Initial Presentation of COVID-19. *J Thromb Thrombolysis*. 2021;51(4):897-901.

81.	Gisondi P, Piaserico S, Naldi L, et al. Incidence rates of hospitalization and death from COVID-19 in patients with psoriasis receiving biological treatment: A Northern Italy experience. *J Allergy Clin Immunol*. 2021;147(2):558.

82.	Glickman JW, Guttman-Yassky E, Pavel AB, Miller RL. The role of circulating eosinophils on COVID-19 mortality varies by race/ethnicity. *Allergy*. 2021;76(3):925-7.

83.	Gomez Antunez M, Muino Miguez A, Bendala Estrada AD, et al. Clinical Characteristics and Prognosis of COPD Patients Hospitalized with SARS-CoV-2 *Int J Chron Obstruct Pulmon Dis*. 2020;15:3433-45.

84.	Gunster C, Busse R, Spoden M, et al. 6-month mortality and readmissions of hospitalized COVID-19 patients: A nationwide cohort study of 8,679 patients in Germany. *PloS ONE.* 2021;16(8):e0255427.

85.	Gupta A, Garg I, Iqbal A, et al. Long-Term X-ray Findings in Patients With Coronavirus Disease-2019. *Cureus*. 2021;13(5):e15304.

86.	Gupta A, Nayan N, Nair R, et al. Diabetes Mellitus and Hypertension Increase Risk of Death in Novel Corona Virus Patients Irrespective of Age: a Prospective Observational Study of Co-morbidities and COVID-19 from India. *SN Compr Clin Med.* 2021;3(4):937-44.

87.	Gurol-Urganci I, Jardine JE, Carroll F, et al. Maternal and perinatal outcomes of pregnant women with SARS-CoV-2 infection at the time of birth in England: national cohort study. *Am J Obstet Gynecol*. 2021.

88.	Haase N, Perner A, Plovsing R, et al. Characteristics, interventions, and longer term outcomes of COVID-19 ICU patients in Denmark-A nationwide, observational study. *Acta Anaesthesiol Scand*. 2021;65(1):68-75.

89.	Hampshire A, Trender W, Chamberlain SR, et al. Cognitive deficits in people who have recovered from COVID-19. *EClinicalMed*. 2021:101044.

90.	Han CH, Park SC, Lee SC, Son KJ, Jung JY. Impact of COPD on COVID-19 prognosis: A nationwide population-based study in South Korea. *Sci Rep*. 2021;11(1):3735.

91.	Hansen ESH, Backer V, Moeller AL, et al. Severe outcomes of covid-19 among patients with copd and asthma. *ERJ Open Res.* 2021;7(1):1-9.

92.	Hasani Azad M, Khorrami F, Kazemi Jahromi M, et al. Clinical and Epidemiological Characteristics of Hospitalized COVID-19 Patients in Hormozgan, Iran: A Retrospective, Multicenter Study. *Arch Iran Med.* 2021;24(5):434-44.

93.	He J, Xu J, Qian H, et al. The prognostic value of myocardial injury in COVID-19 patients and associated characteristics. *Immun Inflamm Dis*. 2021.

94.	He W, Liu B, Spokes P, Kaldor J. High risk groups for severe COVID-19 in a whole of population cohort in Australia. *BMC Infect Dis*. 2021;21(1):685.

95.	Hedberg P, Karlsson Valik J, Van Der Werff S, et al. Clinical phenotypes and outcomes of SARS-CoV-2, influenza, RSV and seven other respiratory viruses: A retrospective study using complete hospital data. *Thorax*. 2021.

96.	Hermel DJ, Spierling Bagsic SR, Costantini CL, Mason JR, Gahvari ZJ, Saven A. ABO phenotype and clinical correlates of COVID-19 severity in hospitalized patients. *Future Sci OA.* 2021;7(8):0045.

97.	Hirschtick JL, Titus AR, Slocum E, et al. Population-based estimates of post-acute sequelae of SARS-CoV-2 infection (PASC) prevalence and characteristics. *Clin Infect Dis*. 2021.

98.	Hsu TYT, D'Silva KM, Patel NJ, et al. Laboratory trends, hyperinflammation, and clinical outcomes for patients with a systemic rheumatic disease admitted to hospital for COVID-19: a retrospective, comparative cohort study. *Lancet Rheumatol.* 2021.

99.	Huang Q-M, Zhou J-M, Liu D, et al. Genetic Risk and COPD Independently Predict the Risk of Incident Severe COVID-19. *Ann Am Thorac Soc*. 2021.

100.	Huang Y, Pinto MD, Borelli JL, et al. COVID Symptoms, Symptom Clusters, and Predictors for Becoming a Long-Hauler: Looking for Clarity in the Haze of the Pandemic. *medRxiv*. 2021.

101.	Hussein AAM, Moustafa M, Saad M, et al. Post-COVID-19 functional status: Relation to age, smoking, hospitalization, and previous comorbidities. *Ann Thorac Med.* 2021;16(3):260-5.

102.	Iaccarino G, Grassi G, Borghi C, Ferri C, Salvetti M, Volpe Massimo M. Age and multimorbidity predict death among COVID-19 Patients: Results of the SARS-RAS study of the Italian society of hypertension. *Hypertension*. 2020;76(2):15324.

103.	Iftimie S, Lopez-Azcona A, Parra S, et al. First and second waves of coronavirus disease-19: A comparative study in hospitalized patients in Reus, Spain. *PLoS ONE*. 2021;16(3):e0248029.

104.	Jaafar S, Morton J, Sangha G, et al. Incidence of symptomatic, image-confirmed venous thromboembolism following hospitalization for COVID-19 with 90-day follow-up. *Blood Adv*. 2020;4(24):6230-9.

105.	Jegatheeswaran V, Chan MWK, Chakrabarti S, Chen YA, Fawcett A. Neuroimaging Findings of Hospitalized Covid-19 Patients: A Canadian Retrospective Observational Study. *Can Assoc Radiol J*. 2021.

106.	Ji H, Dai Q, Xu K, et al. Epidemiology of 631 cases of COVID-19 identified in Jiangsu Province between January 1st and March 20th 2020: Factors associated with disease severity and analysis of zero mortality.*Med Sci Monitor*. 2021;27:e929986.

107.	Kang IS, Kong KA. Body mass index and severity/fatality from coronavirus disease 2019: A nationwide epidemiological study in Korea. *PLoS ONE.* 2021;16(6):e0253640.

108.	Khoonsari M, Zamani F, Niya MHK, et al. Liver enzymes and inpatient deaths in COVID-19 hospitalized patients. *Hepat Mon.* 2020;20(11):1-6.

109.	Kim E, Kim H, Kim YC, Park JY, Jung J, Lee JP. Evaluation of the prognosis of covid-19 patients according to the presence of underlying diseases and drug treatment. *Int J Environ Public Health.* 2021;18(10):5342.

110.	Kim H-S, Kang M, Kang G. Renin-angiotensin system modulators and other risk factors in COVID-19 patients with hypertension: a Korean perspective. *BMC Infect Dis*. 2021;21(1):175.

111.	Kim S-R, Nam S-H, Kim Y-R. Risk factors on the progression to clinical outcomes of covid-19 patients in South Korea: Using national data. *Int J Environ Public Health*. 2020;17(23):1-9.

112.	Kokturk N, Ulukavak Ciftci T, Oguzulgen IK, et al. The predictors of COVID-19 mortality in a nationwide cohort of Turkish patients. *Resp Med.* 2021;183:106433.

113.	Kompaniyets L, Agathis NT, Nelson JM, et al. Underlying Medical Conditions Associated with Severe COVID-19 Illness among Children. *JAMA Netw Open*. 2021:e2111182.

114.	Kotwica A, Knights H, Mayor N, Russell-Jones E, Dassios T, Russell-Jones D. Intrapulmonary shunt measured by bedside pulse oximetry predicts worse outcomes in severe COVID-19. *Eur Resp J*. 2021;57(4):2003841.

115.	Kumar B, Mittal M, Gopalakrishnan M, Garg MK, Misra S. Effect of plasma glucose at admission on covid-19 mortality: Experience from a tertiary hospital. *Endocr Connect.* 2021;10(6):589-98.

116.	Kumar KK, Sampritha UC, Prakash AA, et al. Ophthalmic manifestations in the COVID-19 clinical spectrum. *Indian J Opthalmol*. 2021;69(3):691-4.

117.	Kuo C-L, Kuchel GA, Pilling LC, et al. Biological Aging Predicts Vulnerability to COVID-19 Severity in UK Biobank Participants. *J Gerontol A Biol Sci*. 2021;76(8):e133-e41.

118.	Kwon JS, Park S-H, Jeon H-L, Shin J-Y. Association of mental disorders with SARS-CoV-2 infection and severe health outcomes: Nationwide cohort study. *Br J Psychiatr*. 2021;218(6):344-51.

119.	Laires PA, Dias S, Gama A, et al. The Association Between Chronic Disease and Serious COVID-19 Outcomes and Its Influence on Risk Perception: Survey Study and Database Analysis. *JMIR*. 2021;7(1):e22794.

120.	Lampl BMJ, Buczovsky M, Martin G, Schmied H, Leitzmann M, Salzberger B. Clinical and epidemiological data of COVID-19 from Regensburg, Germany: a retrospective analysis of 1084 consecutive cases. *Infection*. 2021;49(4):661-9.

121.	Lang M, Geller AS, Diffenderfer MR, et al. Corona Virus Disease-19 serology, inflammatory markers, hospitalizations, case finding, and aging. *PLoS ONE*. 2021;16(6):e0252818.

122.	LaPlant Q, Shaverdian N, Shin JY, et al. Association of prior radiation dose to the cardiopulmonary system with COVID-19 outcomes in patients with cancer. *Radiother Oncol*. 2021;161:115-7.

123.	Lee SW, Moon SY, Ha EK, et al. Severe clinical outcomes of COVID-19 associated with proton pump inhibitors: A nationwide cohort study with propensity score matching. *Gut*. 2021;70(1):76-84.

124.	Lee SW, Moon SY, Yang JM, et al. Association between mental illness and COVID-19 susceptibility and clinical outcomes in South Korea: a nationwide cohort study. *Lancet Psychiatr*. 2020;7(12):1025-31.

125.	Lei J, Gong G, Zhang M, et al. Mortality risk of COVID-19 in elderly males with comorbidities: a multi-country study. *Aging.* 2020;13(1):27-60.

126.	Leijte WT, de Kruif MD, Jongbloed M, Linssen CFM, van den Hoogen BG, van Gorp ECM. Clinical impact of human metapneumovirus infections before and during the COVID-19 pandemic. *Infect Dis*. 2021;53(7):488-97.

127.	Leite VF, Rampim DB, Jorge VC, et al. Persistent Symptoms and Disability After COVID-19 Hospitalization: Data From a Comprehensive Telerehabilitation Program. *Arch Phys Med Rehabil.* 2021;102(7):1308-16.

128.	Li HL, Cheung BMY. The Proportion of Adult Americans at Risk of Severe COVID-19 Illness. *J Gen Intern Med* 2021;36(1):259-61.

129.	Li Y, Rao X, Wang B, Wang D, et al. Corticosteroid therapy in critically ill patients with COVID-19: a multicenter, retrospective study. *Crit Care*. 2020;24(1):698.

130.	Lim A, Merle U, Radujkovic A, Weigand MA. Soluble receptor for advanced glycation end products (sRAGE) as a biomarker of COVID-19 disease severity and indicator of the need for mechanical ventilation, ARDS and mortality. *Ann Intensive Care.* 2021;11(1):50.

131.	Lindson N, Tan PS, Clift AK, et al. Association between pre-existing respiratory disease and its treatment, and severe COVID-19: a population cohort study. *Lancet Respir Med.* 2021;9(8):909-23.

132.	Liu B, Jayasundara D, Pye V, et al. Whole of population-based cohort study of recovery time from COVID-19 in New South Wales Australia. *Lancet.* 2021;12:100193.

133.	Liu C, Wen Y, Wan W, Lei J, Jiang X. Clinical characteristics and antibiotics treatment in suspected bacterial infection patients with COVID-19. *Int Immunopharmacol*. 2021;90:107157.

134.	Liu X, Li W, Chen J, Xie R, et al. Clinical and epidemiological features of 46 children <1 year old with coronavirus disease 2019 in Wuhan, China: A descriptive study. *J Infect Dis*. 2020;222(8):1293-7.

135.	Loerinc LB, Scheel AM, Evans ST, Shabto JM, O'Keefe GA, O'Keefe JB. Discharge characteristics and care transitions of hospitalized patients with COVID-19. *Healthcare*. 2021;9(1):100512.

136.	Lopez Bernal J, Andrews N, Gower C, et al. Effectiveness of the Pfizer-BioNTech and Oxford-AstraZeneca vaccines on covid-19 related symptoms, hospital admissions, and mortality in older adults in England: test negative case-control study. *BMJ.* 2021;373:n1088.

137.	Lozano-Cruz O, Jimenez JV, Ortiz-Brizuela E, et al. Adverse Effects Associated With the Use of Antimalarials During The COVID-19 Pandemic in a Tertiary Care Center in Mexico City. *Front Pharmacol*. 2021;12:668678.

138.	Ma X, Ma H, Feng K, et al. Characteristics of 1738 Patients with Coronavirus Disease 2019 (COVID-19) in Wuhan, China. *Disaster Med Public Health*. 2021:1-20.

139.	Macedo MCF, Araujo OAC, Pinheiro IM, et al. Correlation between hospitalized patients' demographics, symptoms, comorbidities, and COVID-19 pandemic in Bahia, Brazil. *PLoS ONE*. 2020;15(12):e0243966.

140.	Mahajan NN, Gajbhiye RK, Bahirat S, et al. Co-infection of malaria and early clearance of SARS-CoV-2 in healthcare workers. *J Med Virol*. 2021;93(4):2431-8.

141.	Mahmud R, Rassel MA, Rahman MM, et al. Post-COVID-19 syndrome among symptomatic COVID-19 patients: A prospective cohort study in a tertiary care center of Bangladesh. *PLoS ONE*. 2021;16(4):e0249644.

142.	Mallia P, Meghji J, Wong B, et al. Symptomatic, biochemical and radiographic recovery in patients with COVID-19. *BMJ Open Respir Res*. 2021;8(1).

143.	Maltezou HC, Raftopoulos V, Vorou R, et al. Association Between Upper Respiratory Tract Viral Load, Comorbidities, Disease Severity, and Outcome of Patients With SARS-CoV-2 Infection. *J Infect Dis*. 2021;223(7):1132-8.

144.	Mandal S, Barnett J, Brill SE, et al. 'Long-COVID': a cross-sectional study of persisting symptoms, biomarker and imaging abnormalities following hospitalisation for COVID-19. *Thorax*. 2021;76(4):396-8.

145.	Mangone L, Gioia F, Mancuso P, et al. Cumulative COVID-19 incidence, mortality and prognosis in cancer survivors: A population-based study in Reggio Emilia, Northern Italy. *Int J Cancer Res*. 2021.

146.	Mannan A, Mehedi HMH, Rob MA, et al. A multi-centre, cross-sectional study on coronavirus disease 2019 in Bangladesh: clinical epidemiology and short-term outcomes in recovered individuals. *New Microbes New Infect.* 2021;40:100838.

147.	Marcolino MS, Souza-Silva M, Nascimento IJB, et al. Clinical characteristics and outcomes of patients hospitalized with COVID-19 in Brazil: Results from the Brazilian COVID-19 registry. *Int J Infect Dis*. 2021;107:300-10.

148.	Marcon CEM, Trevisol DJ, Schuelter-Trevisol F, et al. Assessment of patients with COVID-19 hospitalized in southern Santa Catarina. *Rev Soc Bras Med*. 2020;53:1-5.

149.	Maripuu M, Bendix M, Ohlund L, Werneke U, Widerstrom M. Death Associated With Coronavirus (COVID-19) Infection in Individuals With Severe Mental Disorders in Sweden During the Early Months of the Outbreak-An Exploratory Cross-Sectional Analysis of a Population-Based Register Study. *Front Psychiatry*. 2020;11:609579.

150.	Marjot T, Barnes E, Kennedy J, et al. SARS-CoV-2 infection in patients with autoimmune hepatitis. *J Hepatol*. 2021;74(6):1335-43.

151.	Marquez-Gonzalez H, Klunder-Klunder M, De La Rosa-Zamboni D, et al. Risk conditions in healthcare workers of a pediatric COVID center in Mexico city. *Bol Med Hosp Infant Mex.* 2021;78(2):110-5.

152.	Marron RM, Zheng M, Romero GF, et al. Impact of chronic obstructive pulmonary disease and emphysema on outcomes of hospitalized patients with COVID-19 pneumonia. *COPD*. 2021;8(2).

153.	Martinot M, Gravier S, Bonijoly T, et al. Predictors of mortality, ICU hospitalization, and extrapulmonary complications in COVID-19 patients. *Infect Dis Now*. 2021.

154.	Martins-Filho P, de Souza Araujo AA, Quintans-Junior L, et al. Factors associated with mortality among hospitalized patients with covid-19: A retrospective cohort study. *Am J Trop Med*. 2021;104(1):103-5.

155.	Martos-Benitez F, Soler-Morejon C, Garcia-Del Barco D. Chronic comorbidities and clinical outcomes in patients with and without COVID-19: a large population-based study using national administrative healthcare open data of Mexico. *Intern Emerg Med*. 2021.

156.	Mash RJ, Presence-Vollenhoven M, Hendrikse A, et al. Evaluation of patient characteristics, management and outcomes for COVID-19 at district hospitals in the Western Cape, South Africa: Descriptive observational study. *BMJ Open*. 2021;11(1):e047016.

157.	Mathur R, Rentsch CT, Schultze A, et al. Ethnic differences in SARS-CoV-2 infection and COVID-19-related hospitalisation, intensive care unit admission, and death in 17 million adults in England: an observational cohort study using the OpenSAFELY platform. *Lancet*. 2021;397(10286):1711-24.

158.	Mercuriali L, Politi J, Martin-Sanchez M, et al. Epidemiological characteristics and outcomes of COVID-19 cases: Mortality inequalities by socioeconomic status, Barcelona, Spain, 24 February to 4 May 2020. *Eurosurveillance*. 2021;26(20).

159.	Meyerholz DK, Perlman S. Does common cold coronavirus infection protect against severe SARS-CoV-2 disease? *J Clin Investig*. 2021;131(1):e144807.

160.	Mikkelsen S, Kaspersen KA, Jespersen S, et al. Symptoms reported by SARS-CoV-2 seropositive and seronegative healthcare and administrative employees in Denmark from May to August 2020. *Int J Infect Dis*. 2021;109:17-23.

161.	Moftakhar L, Moftakhar P, Piraee E, Ghaem H, Valipour A, Azarbakhsh H. Epidemiological characteristics and outcomes of COVID-19 in diabetic versus non-diabetic patients. *Int J Diabetes Dev Ctries*. 2021.

162.	Mohamed NE, Okhawere KE, Korn TG, et al. Association between chronic kidney disease and COVID-19-related mortality in New York. *World J Urol.* 2021.

163.	Monari C, Sagnelli C, Camaioni C, et al. More Severe COVID-19 in Patients With Active Cancer: Results of a Multicenter Cohort Study. *Front Oncol*. 2021;11:662746.

164.	Monreal E, Sainz de la Maza S, Natera-Villalba E, et al. The Impact of Immunosuppression and Autoimmune Disease on Severe Outcomes in Patients Hospitalized with COVID-19. *J Clin Immunol*. 2021;41(2):315-23.

165.	Montefusco L, Ben Nasr M, D'Addio F, et al. Acute and long-term disruption of glycometabolic control after SARS-CoV-2 infection. *Nat Metab*. 2021;3(6):774-85.

166.	Morrissey H, Ball P, Nevil A, et al. COVID-19 in haematology patients: a multicentre West Midlands clinical outcomes analysis on behalf of the West Midlands Research Consortium. *Br J Hematol*. 2021;192(1):e11-e4.

167.	Mousavi Movahed SM, Akhavizadegan H, Dolatkhani F, et al. Different incidences of acute kidney injury (AKI) and outcomes in COVID-19 patients with and without non-azithromycin antibiotics: A retrospective study. *J Med Virol*. 2021;93(7):4411-9.

168.	Muller M, Reif A, Trudzinski NKF, et al. Residual symptoms and lower lung function in patients recovering from SARS-CoV-2 infection. *Eur Respir J*. 2021;57(2):03002-2020.

169.	Myall KJ, Mukherjee B, Castanheira AM, et al. Persistent post-COVID-19 interstitial lung disease: An observational study of corticosteroid treatment. *Ann Am Thorac Soc.* 2021;18(5):799-806.

170.	Naidu SB, Shah AJ, Saigal A, et al. The high mental health burden of "long covid" and its association with on-going physical and respiratory symptoms in all adults discharged from hospital. *Eur Respir J*. 2021;57(6):2004364.

171.	Nair V, Satapathy SK, Roth N, et al. An early experience on the effect of solid organ transplant status on hospitalized COVID-19 patients. *Am J Transplant*. 2021;21(7):2522-31.

172.	Neant N, Lingas G, Guedj J, et al. Modeling SARS-CoV-2 viral kinetics and association with mortality in hospitalized patients from the French COVID cohort. *PNAS*. 2021;118(8):e2017962118.

173.	Nogueira Lopez J, Grasa C, Calvo C, Garcia Lopez-Hortelano M. Long-term symptoms of COVID-19 in children. Acta Paediatrica, *Int J Pediatr*. 2021;110(7):2282-3.

174.	Oddy C, McCaul J, Keeling P, et al. Pharmacological Predictors of Morbidity and Mortality in COVID-19. *J Clin Pharmacol*. 2021.

175.	O'Keefe JB, Tong EJ, O'Keefe GD, Tong DC. Description of symptom course in a telemedicine monitoring clinic for acute symptomatic covid-19: A retrospective cohort study. *BMJ Open*. 2021;11(3):e044154.

176.	Osaghae I, Nguyen LK, Chung TH, et al. Prevalence and Factors Associated With Mental Health Symptoms in Adults Undergoing Covid-19 Testing. *J Prim Care Community Health.* 2021;12:21501327211027100.

177.	Osikomaiya B, Erinoso O, Wright KO, et al. 'Long COVID': persistent COVID-19 symptoms in survivors managed in Lagos State, Nigeria. *BMC Infect Dis*. 2021;21(1):304.

178.	Ozcan S, Emeksiz S, Perk O, Uyar E, Kanik Yuksek S. Severe Coronavirus Disease Pneumonia in Pediatric Patients in a Referral Hospital. *J Trop Pediatr.* 2021;67(3):fmab052.

179.	Paneroni M, Simonelli C, Saleri M, et al. Muscle Strength and Physical Performance in Patients Without Previous Disabilities Recovering From COVID-19 Pneumonia. *Am J Phys Med Rehabil*. 2021;100(2):105-9.

180.	Pannu AK, Kumar M, Singh P, et al. Severe acute respiratory infection surveillance during the initial phase of the covid-19 outbreak in north india: Comparison of covid-19 to other sari causes. *Indian J Crit Care Med*. 2021;25(7):761-7.

181.	Pennington AF, Kompaniyets L, Summers AD, et al. Risk of Clinical Severity by Age and Race/Ethnicity among Adults Hospitalized for COVID-19 - United States, March-September 2020. *Open Forum Infect Dis*. 2021;8(2):ofaa638.

182.	Pereda R, Gonzalez D, Rivero HB, et al. Therapeutic Effectiveness of Interferon Alpha 2b Treatment for COVID-19 Patient Recovery. *J Interferon Cytokine Res*. 2020;40(12):578-88.

183.	Perez-Garcia C, Enriquez-Vazquez D, Olmos C, et al. Article the sadden death study: Results from a pilot study in non-icu covid-19 spanish patients. *J Clin Med*. 2021;10(4):1-16.

184.	Perlis RH, Green J, Santillana M, et al. Persistence of symptoms up to 10 months following acute COVID-19 illness. *medRxiv*. 2021.

185.	Piazza G, Goldhaber SZ, Bikdeli B, et al. Intermediate-Dose versus Standard-Dose Prophylactic Anticoagulation in Patients with COVID-19 Admitted to the Intensive Care Unit: 90-Day Results from the INSPIRATION Randomized Trial. *Thromb Haemost*. 2021.

186.	Pinto-Sietsma S, Offerhaus J, Bleijendaal H, et al. Antihypertensive drugs in COVID-19 infection. *Eur Heart J Cardiovasc Pharmacother*. 2020;6(6):415-6.

187.	Politi J, Martin-Sanchez M, Mercuriali L, et al. Epidemiological characteristics and outcomes of COVID-19 cases: mortality inequalities by socio-economic status, Barcelona, Spain, 24 February to 4 May 2020. *Euro Surveill*. 2021;26(20).

188.	Priya S, Selva Meena M, Brinda Priyadharshini C, Vijay Anand V, Sangumani J, Rathinam P. "Factors influencing the outcome of COVID-19 patients admitted in a tertiary care hospital, Madurai.- a cross-sectional study". *Clin Epidemiology Glob Health*. 2021;10:100705.

189.	Queiroz NSF, de Almeida Martins C, Damiao AOMC, et al. Risk stratification and geographical mapping of Brazilian inflammatory bowel disease patients during the COVID-19 outbreak: Results from a nationwide survey. *World J Gastroenterol.* 2021;27(12):1226-39.

190.	Rahim BIH, Babakir-Mina M. Clinical characteristics of covid-19 in patients: A meta-analysis. *Indian J Forensic Med Toxicol*. 2021;15(3):2672-6.

191.	Ramani C, Davis EM, Kim JS, Enfield KB, Provencio JJ, Kadl A. Post-ICU COVID-19 Outcomes: A Case Series. *Chest*. 2021;159(1):215-8.

192.	Rana MS, Usman M, Alam MM, et al. Clinical and laboratory characteristics of recovered versus deceased COVID-19 patients in Islamabad, Pakistan. *J Infect*. 2021;82(4):84-123.

193.	Ray JG, Park AL, Vermeulen MJ, Schull MJ. ABO Blood Group, SARS-CoV-2 Infection, and Risk of Venous Thromboembolism: Population-Based Cohort Study. *Clin Appl Thromb Hemost*. 2021;27.

194.	Righi E, Mirandola M, Mazzaferri F, et al. Long-Term Patient-Centred Follow-up in a Prospective Cohort of Patients with COVID-19. *Infect Dis Ther*. 2021;10(3):1579-90.

195.	Robinson LB, Long AA, Wang L, Fet al. COVID-19 severity in asthma patients: a multi-center matched cohort study. *J Asthma.* 2020.

196.	Rodeles LM, Peverengo LM, Benitez R, et al. Seroprevalence of anti-SARS-CoV-2 IgG in asymptomatic and pauci-symptomatic people over a 5 month survey in Argentina. *Rev Panam Salud Publica*. 2021;45:e66.

197.	Romagnolo A, Balestrino R, Imbalzano G, et al. Neurological comorbidity and severity of COVID-19. *J Neurol*. 2021;268(3):762-9.

198.	Rosales-Castillo A, Garcia de los Rios C, Mediavilla Garcia JD. Persistent symptoms after acute COVID-19 infection: importance of follow-up. *Med Clin*. 2021;156(1):35-6.

199.	Russo E, Esposito P, Saio M, et al. Kidney disease and all-cause mortality in patients with COVID-19 hospitalized in Genoa, Northern Italy. *J Nephrol*. 2021;34(1):173-83.

200.	Sahin M, Rifat E, Haymana C, et al. The clinical outcomes of COVID-19 infection in patients with a history of thyroid cancer: A nationwide study. *Clin Endocrinol*. 2021.

201.	Schavemaker R, Lagrand WK, Schultz MJ, van Slobbe-Bijlsma ER, Neto AS, Paulus F. Associations of body mass index with ventilation management and clinical outcomes in invasively ventilated patients with ards related to covid-19-insights from the provent-covid study. *J Clin Med*. 2021;10(6):1-14.

202.	Seligman B, Ferranna M, Bloom DE. Social determinants of mortality from COVID-19: A simulation study using NHANES. *PLoS Med*. 2021;18(1):e1003490.

203.	Sen P, Majumdar U, Hatipoglu U, Attaway AH, Zein J. Inhaled corticosteroids do not adversely impact outcomes in COVID-19 positive patients with COPD: An analysis of Cleveland Clinic's COVID-19 registry. *PLoS ONE*. 2021;16(6):e0252576.

204.	Sharif N, Ahmed SN, Opu RR, et al. Prevalence and impact of diabetes and cardiovascular disease on clinical outcome among patients with COVID-19 in Bangladesh. *Diabetes Metab Syndr.* 2021;15(3):1009-16.

205.	Shin EK, Choi HY, Hayes N. The anatomy of COVID-19 comorbidity networks among hospitalized Korean patients. *Epidemiol Health*. 2021;43:e2021035.

206.	Singer ME, Taub IB, Kaelber DC. Risk of Myocarditis from COVID-19 Infection in People Under Age 20: A Population-Based Analysis. *medRxix.* 2021.

207.	Song J, Zeng M, Wang H, et al. Distinct effects of asthma and COPD comorbidity on disease expression and outcome in patients with COVID-19. *Allergy*. 2021;76(2):483-96.

208.	Soria MLJB, Quiwa LQ, Calvario MKJS, Duya JED, Punongbayan RB, Ting FIL. The philippine coronavirus disease 2019 (Covid-19) profile study: Clinical profile and factors associated with mortality of hospitalized patients. *Philipp J Intern Med*. 2021;59(1):37-58.

209.	Sormani MP, Battaglia MA, Nozzolillo A, et al. Disease-Modifying Therapies and Coronavirus Disease 2019 Severity in Multiple Sclerosis. *Ann Neurol*. 2021;89(4):780-9.

210.	Steidler S, Martinenghi CMA, Mushtaq J, et al. Initial chest radiographs and artificial intelligence (AI) predict clinical outcomes in COVID-19 patients: analysis of 697 Italian patients. *Eur Radiol*. 2021;31(3):1770-9.

211.	Sudre CH, Murray B, Varsavsky T, et al. Attributes and predictors of long COVID. *Nat Med*. 2021;27(4):626-31.

212.	Taylor MF, Urena MC, Torres M, Martinoia A, Ciappa JM, Mir GA. Covid-19 in renal transplant patients, on the waiting list and under evaluation for transplantation. Experience in a public hospital in argentina. *Rev de Nefrol Dialysis y Transpl.* 2021;41(2):119-24.

213.	Turan O, Arpinar Yigitbas B, Turan PA, Mirici A. Clinical characteristics and outcomes of hospitalized COVID-19 patients with COPD. *Expert Rev Respir Med*. 2021.

214.	Tzur Bitan D, Kridin K, Cohen AD, Weinstein O. COVID-19 hospitalisation, mortality, vaccination, and postvaccination trends among people with schizophrenia in Israel: a longitudinal cohort study. *Lancet Psychiatry*. 2021.

215.	Ungaro RC, Agrawal M, Park S, et al. Autoimmune and Chronic Inflammatory Disease Patients with COVID-19. *ACR Open Rheumatol*. 2021;3(2):111-5.

216.	Valent F. Age, comorbidities, nursing home stay and outcomes of SARS-CoV-2 infection in a Northern Italian cohort. *J Gerontol Geriatr*. 2021;69(2):114-9.

217.	Valenzuela RG, Bracey A, Cruz P, et al. Outcomes in Hispanics With COVID-19 Are Similar to Those of Caucasian Patients in Suburban New York. *Acad Emerg Med*. 2020;27(12):1260-9.

218.	Valverde-Monge M, Canas JA, Barroso B, et al. Eosinophils and Chronic Respiratory Diseases in Hospitalized COVID-19 Patients. *Front Immunol*. 2021;12:668074.

219.	Venturelli S, Benatti SV, Casati M, et al. Surviving COVID-19 in Bergamo Province: A post-Acute outpatient re-evaluation. *Epidemiol Infect*. 2021.

220.	Vera-Zertuche J, Mancilla-Galindo J, Aguirre-Garcia M, et al. Obesity is a strong risk factor for short-term mortality and adverse outcomes in Mexican patients with COVID-19: A national observational study. *Epidemiol Infect*. 2021.

221.	Verderese JP, Stepanova M, Lam B, et al. Neutralizing Monoclonal Antibody Treatment Reduces Hospitalization for Mild and Moderate COVID-19: A Real-World Experience. *Clin Infect Dis*. 2021.

222.	Vergara A, Molina-Van Den Bosch M, Toapanta N, et al. The impact of age on mortality in chronic haemodialysis popu-lation with covid-19. *J Clin Med*. 2021;10(14):3022.

223.	Verma R, Morin DP, Manzo MA, et al. Impact of Preinfection Left Ventricular Ejection Fraction on Outcomes in COVID-19 Infection. *Curr Probl Cardiol*. 2021:100845.

224.	Villalba GC, Catala P, Aparisi A, et al. Impact of the presence of heart disease, cardiovascular medications and cardiac events on outcome in covid-19. *Cardiol J*. 2021;28(3):360-8.

225.	Villamanan E, Sobrino C, Moreno M, et al. Inhaled bronchodilators use and clinical course of adult inpatients with Covid-19 pneumonia in Spain: A retrospective cohort study. *Pulm Pharmacol Ther*. 2021;69:102007.

226.	Wang M, Xiong N, Jiang N, et al. Sex-Disaggregated Data on Clinical Characteristics and Outcomes of Hospitalized Patients With COVID-19: A Retrospective Study. *Front Cell Infect Microbiol*. 2021;11:680422.

227.	Wang S-M, Na H-R, Lim HK, et al. Association between dementia and clinical outcome after covid-19: A nationwide cohort study with propensity score matched control in south korea. *Psychiatry Investig.* 2021;18(6):523-9.

228.	Wang X, Zhang H, Du H, Ma R, Nan Y, Zhang T. Risk Factors for COVID-19 in Patients with Hypertension. *Can J Infect Dis Med Microbiol*. 2021;2021:5515941.

229.	Wu Q, Huang W, Chen Z, et al. The potential indicators for pulmonary fibrosis in survivors of severe COVID-19. *J Infect*. 2021;82(2):e5-e7.

230.	Yacobitti A, Otero L, Doldan Arrubarrena V, et al. Clinical characteristics of vulnerable populations hospitalized and diagnosed with COVID-19 in Buenos Aires, Argentina. *Sci Rep*. 2021;11(1):9679.

231.	Yahyavi A, Hemmati N, Derakhshan P, et al. Angiotensin enzyme inhibitors and angiotensin receptor blockers as protective factors in COVID-19 mortality: a retrospective cohort study. *Intern Emerg Med*. 2021;16(4):883-93.

232.	Yang JM, Lee JY, Moon SY, Agalliu D, Yon DK, Lee SW. COVID-19 morbidity and severity in patients with age-related macular degeneration: a Korean nationwide cohort study. *Am J Ophthalmol*. 2021:11881.

233.	Yllescas M, Arriba JR, Aznar Munoz E, et al. Characteristics and predictors of death among 4035 consecutively hospitalized patients with COVID-19 in Spain. *Clin Microbiol Infect*. 2020;26(11):1525-36.

234.	Yoo J, Grewal P, Hotelling J, et al. Admission NT-proBNP and outcomes in patients without history of heart failure hospitalized with COVID-19. *ESC Heart Fail*. 2021.

235.	Yozgat A, Kasapoglu B, Can G, et al. Long-Term Proton Pump Inhibitor Use is a Risk Factor for Mortality in Patients Hospitalized for COVID-19. *Turk J Med Sci*. 2021.

236.	Yu B, Gutierrez VP, Carlos A, et al. Empiric use of anticoagulation in hospitalized patients with COVID-19: a propensity score-matched study of risks and benefits. *Biomark Res*. 2021;9(1):29.

237.	Yun K, Lee JS, Kim EY, Chandra H, Oh B-L, Oh J. Severe COVID-19 Illness: Risk Factors and Its Burden on Critical Care Resources. *Front Med*. 2020;7:583060.

238.	Zarif A, Joy M, Sherlock J, et al. The impact of primary care supported shielding on the risk of mortality in people vulnerable to COVID-19: English sentinel network matched cohort study. *J Infect*. 2021;83(2):228-36.

239.	Zettersten E, Bell M, Jaderling G, et al. Long-term outcome after intensive care for COVID-19: differences between men and women-a nationwide cohort study. *Crit Care.* 2021;25(1):86.

240.	Zheng M, Song L. Shift in the distributions of pre-existing medical condition, Gender and age across different COVID-19 outcomes. *Aging Dis*. 2021;12(2):327-9.

241.	Zhou M, Cai J, Sun W, et al. Do post-COVID-19 symptoms exist? A longitudinal study of COVID-19 sequelae in Wenzhou, China. *Ann Med Psychol*. 2021.

**All ICU population (n=2)**

1.	Herzog AL, Von Jouanne-Diedrich HK, Wanner C, et al. COVID-19 and the kidney: A retrospective analysis of 37 critically ill patients using machine learning. *PLoS ONE*. 2021;16(5):e0251932.

2.	Taboada M, Carinena A, Moreno E, et al. Quality of life, functional status, and persistent symptoms after intensive care of COVID-19 patients. *Br J Anaesth*. 2021;126(3):e110-e3.

**Case series (n=7)**

1.	Chai C, Feng X, Lu M, et al. One-Year Mortality and Consequences of COVID-19 in Cancer Patients: a Cohort Study. *IUBMB Life*. 2021.

2.	Evans JS, Chung C, Pinato DJ, et al. Determinants of enhanced vulnerability to coronavirus disease 2019 in UK patients with cancer: a European study. *Eur J Cancer*. 2021;150:190-202.

3.	Manson B, Clark SG, Hendra H, et al. Identifying prognostic risk factors for poor outcome following COVID-19 disease among in-centre haemodialysis patients: role of inflammation and frailty. *J Nephrol*. 2021;34(2):315-23.

4.	Meshram HS, Kute VB, Chauhan S, Desai S. Mucormycosis in post-COVID-19 renal transplant patients: A lethal complication in follow-up. *Transpl Infect Dis*. 2021.

5.	Pena JE-dl, Gonzalez-Figueroa E, Medina-Gomez O, et al. Hypertension, Diabetes and Obesity, Major Risk Factors for Death in Patients with COVID-19 in Mexico. *Arch Med Res*. 2021;52(4):443-9.

6.	Sun M, Wang P, Zheng S, et al. Clinical characteristics of 30 COVID-19 patients with epilepsy: A retrospective study in Wuhan. *Int J Infect Dis.* 2021;103:647-53.

7.	Zhang X, Brenner EJ, Kappelman MD, et al. Presence of Comorbidities Associated with Severe Coronavirus Infection in Patients with Inflammatory Bowel Disease. *Dig Dis Sci*. 2021.

**N <300 with COVID-19 (n=55)**

1.	Abbas HM, Nassir KF, Al Khames Aga QA, et al. Presenting the characteristics, smoking versus diabetes, and outcome among patients hospitalized with COVID-19. *J Med Virol.* 2021;93(3):1556-67.

2.	Ashimov J, Kudaiberdiev T, Gaybyldaev J, Zaripov D, Akhmedova I, Abibillaev D. The effects of tocilizumab on clinical and laboratory features of patients with severe COVID-19: A single center experience. *Heart Vessels Transpl*. 2020;4(4):220.

3.	Bliddal S, Banasik K, Pedersen OB, et al. Acute and persistent symptoms in non-hospitalized PCR-confirmed COVID-19 patients. *Sci Rep*. 2021;11(1):13153.

4.	Bordin A, Gaudioso P, Nicolai P, et al. Comparison of self-reported symptoms and psychophysical tests in coronavirus disease 2019 (COVID-19) subjects experiencing long-term olfactory dysfunction: a 6-month follow-up study. *Int Forum Allergy Rhinol*. 2021.

5.	Boscolo-Rizzo P, Guida F, Marcuzzo AV, et al. Self-reported smell and taste recovery in coronavirus disease 2019 patients: a one-year prospective study. *Eur Arch Otorhinolaryngol*. 2021.

6.	Brandao Neto D, Dib C, Di Francesco RC, et al. Chemosensory Dysfunction in COVID-19: Prevalences, Recovery Rates, and Clinical Associations on a Large Brazilian Sample. *Otolaryngol Head Neck Surg.* 2021;164(3):512-8.

7.	Claflin ES, Daunter AK, Bowman A, et al. Hospitalized Patients With COVID-19 and Neurological Complications Experience More Frequent Decline in Functioning and Greater Rehabilitation Needs. *Am J Phys Med Rehabil*. 2021;100(8):725-9.

8.	Cristillo V, Pilotto A, Cotti Piccinelli S, et al. Age and subtle cognitive impairment are associated with long-term olfactory dysfunction after COVID-19 infection. *J Am Geriatr Soc*. 2021.

9.	Damanti S, Bozzolo EP, Scotti R, et al. 6-Month Respiratory Outcomes and Exercise Capacity of COVID-19 Acute Respiratory Failure Patients Treated With CPAP. *Intern Med J*. 2021.

10.	Du Z, Guo Y, Guo W, et al. The clinical characteristics and prognosis of COVID-19 patients with comorbidities: A retrospective analysis of the infection peak in Wuhan. *Ann Transl Med*. 2021;9(4):280.

11.	Frontera JA, Lewis A, Melmed K, et al. Prevalence and Predictors of Prolonged Cognitive and Psychological Symptoms Following COVID-19 in the United States. *Front Aging Neurosci*. 2021;13:690383.

12.	Fu H, Zhang N, Zheng Y, et al. Risk stratification of cardiac sequelae detected using cardiac magnetic resonance in late convalescence at the six-month follow-up of recovered COVID-19 patients. *J Infect*. 2021;83(1):119-45.

13.	Guarin G, Lo KB, Bhargav R, et al. Factors associated with hospital readmissions among patients with COVID-19: A single-center experience. *J Med Virol*. 2021;93(9):5582-7.

14.	Han X, Fan Y, Alwalid O, et al. Fibrotic Interstitial Lung Abnormalities at 1-year Follow-up CT after Severe COVID-19. *Radiology*. 2021:210972.

15.	Hua-Huy T, Lorut C, Aubourg F, et al. Persistent nasal inflammation 5 months after acute anosmia in patients with COVID-19. *Am J Respir Crit Care Med*. 2021;203(10):1319-22.

16.	Ismaiel WF, Abdelazim MH, Eldsoky I, et al. The impact of COVID-19 outbreak on the incidence of acute invasive fungal rhinosinusitis. *Am J Otolaryngol*. 2021;42(6):103080.

17.	Kanberg N, Eden A, Andersson L-M, et al. Neurochemical signs of astrocytic and neuronal injury in acute COVID-19 normalizes during long-term follow-up. *EBioMedicine.* 2021;70:103512.

18.	Kashif A, Chaudhry M, Fayyaz T, et al. Follow-up of COVID-19 recovered patients with mild disease. *Sci Rep*. 2021;11(1):13414.

19.	Le Bon S-D, Saussez S, Khalife M, et al. Predictive factors of smell recovery in a clinical series of 288 coronavirus disease 2019 patients with olfactory dysfunction. *Eur J Neurol*. 2021.

20.	Li X, Li Y, Qing L, et al. Pulmonary fibrosis and its related factors in discharged patients with new corona virus pneumonia: a cohort study. *Respir Res*. 2021;22(1):203.

21.	Lim JWJ, Taylor JW, Saief T, et al. Smell and taste loss in COVID-19 patients: assessment outcomes in a Victorian population. *Acta Otolaryngol.* 2021;141(3):299-302.

22.	Lucidi D, Molinari G, Silvestri M, et al. Patient-reported olfactory recovery after SARS-CoV-2 infection: A 6-month follow-up study. *Int Forum Allergy Rhinol*. 2021;11(8):1249-52.

23.	Mallardo S, Marcellino A, Bloise S, et al. The comprehensive clinic, laboratory, and instrumental evaluation of children with COVID-19: A 6-months prospective study. *J Med Virol*. 2021;93(5):3122-32.

24.	McCue C, Cowan R, Quasim T, Puxty K, McPeake J. Long term outcomes of critically ill COVID-19 pneumonia patients: early learning. *Intensive Care Med*. 2021;47(2):240-1.

25.	McGroder CF, Zhang D, Choudhury MA, et al. Pulmonary fibrosis 4 months after COVID-19 is associated with severity of illness and blood leucocyte telomere length. *Thorax*. 2021.

26.	Miyazato Y, Akashi M, Osanai Y, et al. Prolonged and Late-Onset Symptoms of Coronavirus Disease 2019. *Open Forum Infect Dis*. 2020;7(11):ofaa507.

27.	Moreno-Perez O, Merino E, Boix V, et al. Post-acute COVID-19 syndrome. Incidence and risk factors: A Mediterranean cohort study. *J Infect*. 2021;82(3):378-83.

28.	Nguyen NN, Gautret P, Hoang VT, Lagier J-C, Raoult D. Long-term persistence of olfactory and gustatory disorders in COVID-19 patients. *Clin Microbiol Infect*. 2021;27(6):931-2.

29.	Noviello D, Costantino A, Vecchi M, et al. Functional gastrointestinal and somatoform symptoms five months after SARS-CoV-2 infection: A controlled cohort study. *Neurogastroenterol Motil.* 2021.

30.	O'Kelly B, Townsend L, Cheallaigh CN, et al. Persistent poor health after covid-19 is not associated with respiratory complications or initial disease severity. *Ann Am Thorac Soc*. 2021;18(6):997-1003.

31.	Olds H, Liu J, Luk K, Lim HW, Ozog D, Rambhatla PV. Telogen effluvium associated with COVID-19 infection. *Dermatol Ther*. 2021;34(2):e14761.

32.	Orieux A, Rubin S, Prevel R, Garric A, Gruson D, Boyer A. Severe COVID-19-induced AKI: a 3-month follow-up. *Clin Kidney J*. 2021;14(4):1289-90.

33.	Osikomaiya B, Erinoso O, Wright KO, et al. 'Long COVID': persistent COVID-19 symptoms in survivors managed in Lagos State, Nigeria. *BMC Infect Dis*. 2021;21(1):304.

34.	Ozcelik F, Tanoglu A, Guven BB, Keskin U, Kaplan M. Assessment of severity and mortality of COVID-19 with anti-A1 and anti-B IgM isohaemagglutinins, a reflection of the innate immune status. *Int J Clin Pract*. 2021.

35.	Pang NTP, Imon GN, Johoniki E, et al. Fear of COVID-19 and COVID-19 stress and association with sociodemographic and psychological process factors in cases under surveillance in a frontline worker population in borneo. *Int J Environ Res Public Health*. 2021;18(13):7210.

36.	Peluso MJ, Kelly JD, Lu S, et al. Rapid implementation of a cohort for the study of post-acute sequelae of SARS-CoV-2 infection/COVID-19. *medRxiv*. 2021.

37.	Pistea C, Enache I, Geny B, et al. Respiratory follow-up after hospitalization for COVID-19: Who and when? *Eur J Clin Investig*. 2021;51(8):e13603.

38.	Puentes-Gutierrez A, Sanchez-Casado M, Diaz-Jimenez M. [Shoulder pain as residual injury after hospital discharge in patients admitted to ICU for COVID-19 pneumonia]. *Med Clin*. 2021;156(6):301-2.

39.	Renaud M, Thibault C, Le Normand F, et al. Clinical Outcomes for Patients With Anosmia 1 Year After COVID-19 Diagnosis. *JAMA Netw Open*. 2021;4(6):e2115352.

40.	Sanz X, Clemente M, Ortega L, et al. Long-term outcomes of patients following hospitalization for coronavirus disease 2019: a prospective observational study. *Clin Microbiol Infect*. 2021.

41.	Shah AS, Wong AW, Johnston JC, et al. A prospective study of 12-week respiratory outcomes in COVID-19-related hospitalisations. *Thorax.* 2021;76(4):402-4.

42.	Shahrvini B, Prajapati DP, Said M, et al. Risk factors and characteristics associated with persistent smell loss in coronavirus disease 2019 (COVID-19) patients. *Int Forum* *Allergy Rhinol*. 2021.

43.	Singhania SVK, Simon C, Raut A, Parvatkar N. Pulmonary sequelae of moderate-to-severe COVID pneumonia, a 3-month follow-up study. *Lung India*. 2021;38(4):397-9.

44.	Skala M, Kopecky M, Koblizek V, et al. Heterogeneity of post-COVID impairment: interim analysis of a prospective study from Czechia. *Virol J.* 2021;18(1):73.

45.	Skjorten I, Ankerstjerne OAW, Trebinjac D, et al. Cardiopulmonary exercise capacity and limitations 3 months after COVID-19 hospitalisation. *Eur Respir J.* 2021.

46.	Smane L, Pucuka Z, Roge I, Pavare J, Stars I. Persistent clinical features in paediatric patients after SARS-CoV-2 virological recovery: A retrospective population-based cohort study from a single centre in Latvia. *BMJ Paediatr Open*. 2020;4(1):e000905.

47.	Sykes DL, Holdsworth L, Jawad N, Gunasekera P, Morice AH, Crooks MG. Post-COVID-19 Symptom Burden: What is Long-COVID and How Should We Manage It? *Lung.* 2021;199(2):113-9.

48.	Taboada M, Carinena A, Diaz-Vieito M, et al. Post-COVID-19 functional status six-months after hospitalization. *J Infect*. 2021;82(4):e31-e3.

49.	Talla A, Vasaikar SV, Lemos MP, et al. Longitudinal immune dynamics of mild COVID-19 define signatures of recovery and persistence. *bioRxiv*. 2021.

50.	Truffaut L, Demey L, Bruyneel AV, et al. Post-discharge critical COVID-19 lung function related to severity of radiologic lung involvement at admission. *Respir Res*. 2021;22(1):29.

51.	Wang S, Fu L, Huang K, Han J, Zhang R, Fu Z. Neutrophil-to-lymphocyte ratio on admission is an independent risk factor for the severity and mortality in patients with coronavirus disease 2019. *J Infect*. 2021;82(2):e16-e8.

52.	Yan X, Zhu Y, Huang H, et al. Follow-up study of pulmonary function among COVID-19 survivors 1 year after recovery. *J Infect*. 2021.

53.	Zhao Y, Liu Y, Yi F, et al. Type 2 diabetes mellitus impaired nasal immunity and increased the risk of hyposmia in COVID-19 mild pneumonia patients. *Int Immunopharmacol.* 2021;93:107406.

54.	Zhong C-H, Zhou Z-Q, Ye F, et al. Prognosis of Severe and Critical Patients Who Have Recovered from COVID-19: A Three-Month Follow-Up. *Respiration*. 2021;100(4):364-7.

55.	Zou J-N, Sun L, Wang B-R, et al. The characteristics and evolution of pulmonary fibrosis in COVID-19 patients as assessed by AI-assisted chest HRCT. *PloS ONE.* 2021;16(3):e0248957.

**No adjusted analysis (n=22)**

1.	Anastasio F, Barbuto S, Cosma P, et al. Medium-term impact of COVID-19 on pulmonary function, functional capacity and quality of life. *Eur Respir J*. 2021.

2.	Augustin M, Schommers P, Stecher M, et al. Post-COVID syndrome in non-hospitalised patients with COVID-19: a longitudinal prospective cohort study. *Lancet*. 2021;6:100122.

3.	de Cortina Camarero C, Gomez Mariscal E, Munoz Aguilera R, Espejo Bares V, Nunez Garcia A, Botas Rodriguez J. SARS-CoV-2 infection: A predisposing factor for acute coronary syndrome. *Med Clin*. 2021;157(3):114-7.

4.	Divanoglou A, Samuelsson APK, Levi PR, Sjodahl PER, Andersson C. Rehabilitation needs and mortality associated with the Covid-19 pandemic: a population-based study of all hospitalised and home-healthcare individuals in a Swedish healthcare region. *EClinicalMedicine*. 2021;36:100920.

5.	Eggert LE, He Z, Lee AS, et al. Asthma phenotypes, associated comorbidities, and long-term symptoms in COVID-19. *Allergy*. 2021.

6.	Eriksson KE, Campoccia-Jalde F, Rysz S, Rimes-Stigare C. Continuous renal replacement therapy in intensive care patients with COVID-19; survival and renal recovery. *J Crit Care*. 2021;64:125-30.

7.	Estiri H, Strasser ZH, Brat GA, et al. Evolving Phenotypes of non-hospitalized Patients that Indicate Long Covid. *medRxiv*. 2021.

8.	Ferdous MZ, Islam US, Islam MS, Mosaddek ASM, Potenza MN, Pardhan S. Treatment, persistent symptoms, and depression in people infected with covid-19 in bangladesh. *Int J Environ Res Public Health.* 2021;18(4):1-16.

9.	Hernandez-Romieu A, Leung S, Mbanya A, et al. Health Care Utilization and Clinical Characteristics of Nonhospitalized Adults in an Integrated Health Care System 28-180 Days After COVID-19 Diagnosis - Georgia, May 2020-March 2021. *Morb Mortal Wkly Rep*. 2021;70(17):644-50.

10.	Lund LC, Pottegard A, Hallas J, et al. Post-acute effects of SARS-CoV-2 infection in individuals not requiring hospital admission: a Danish population-based cohort study. *Lancet Infect Dis*. 2021.

11.	Mata-Vazquez E, Martin-Toledano M, Lopez-Larramona G, et al. Long-term outcomes of patients with coronavirus disease 2019 at one year after hospital discharge. *J Clin Med*. 2021;10(13):2945.

12.	Molinari C, Laurenzi A, Caretto A, et al. Dysglycemia after COVID-19 pneumonia: a six-month cohort study. *Acta Diabetol.* 2021.

13.	Moulson N, Petek BJ, Drezner JA, et al. SARS-CoV-2 Cardiac Involvement in Young Competitive Athletes. *Circulation*. 2021;144(4):256-66.

14.	Niedzwiedz CL, Benzeval M, Hainey K, Leyland AH, Katikireddi SV. Psychological distress among people with probable COVID-19 infection: Analysis of the UK Household Longitudinal Study. *BJPsych Open.* 2021;7(3):e104.

15.	Rogers-Brown J, Wanga V, Okoro C, et al. Outcomes Among Patients Referred to Outpatient Rehabilitation Clinics After COVID-19 diagnosis - United States, January 2020-March 2021. *Morb Mortal Wkly Rep*. 2021;70(27).

16.	Ros M, Martin M, Cachero M, et al. Impact of COVID-19 on nutritional status during the first wave of the pandemic. *Clin Nutr*. 2021.

17.	Shoucri SM, Purpura L, Theodore DA, et al. Characterising the long-term clinical outcomes of 1190 hospitalised patients with COVID-19 in New York City: A retrospective case series. *BMJ Open*. 2021;11(6):e049488.

18.	Taylor RR, Trivedi B, Patel N, et al. Post-COVID symptoms reported at asynchronous virtual review and stratified follow-up after COVID-19 pneumonia. *Clin Med.* 2021.

19.	Torralba Y, Sibila O, Albacar N, et al. Lung Function sequelae in COVID-19 Patients 3 Months After Hospital Discharge. *Arch Bronconeumol*. 2021;57:59-61.

20.	Tran V-T, Riveros C, Clepier B, et al. Development and validation of the long covid symptom and impact tools, a set of patient-reported instruments constructed from patients' lived experience. *Clin Infect Dis*. 2021.

21.	Wu Q, Hou X, Li H, et al. A follow-up study of respiratory and physical function after discharge in patients with redetectable positive SARS-CoV-2 nucleic acid results following recovery from COVID-19. *Int J Infect Dis*. 2021;107:5-11.

22.	Yom-Tov E, Lekkas D, Jacobson NC. Association of COVID19-induced anosmia and ageusia with depression and suicidal ideation. *J Affect Disord.* 2021;5:100156.

**Protocol (n=11)**

1.	Arevalos V, Ortega-Paz L, Fernandez-Rodriguez D, et al. Long-term effects of coronavirus disease 2019 on the cardiovascular system, CV COVID registry: A structured summary of a study protocol. *PloS ONE.* 2021;16(7):e0255263.

2.	Avramovic G, McHugh T, Connolly SP, Cullen W, Lambert JS. Anticipate study protocol: Baseline profile and care outcomes of patients attending Mater Misericordiae University Hospital with COVID-19 infection. *HRB Open Res*. 2020;3:52.

3.	Catalfamo CJ, Heslin KM, Shilen A, et al. Design of the Arizona CoVHORT: A Population-Based COVID-19 Cohort. *Front Public Health*. 2021;9:620060.

4.	Drapkina OM, Loukyanov MM, Martsevich SY, et al. Experience of creating and the first results of the prospective hospital registry of patients with suspected or confirmed coronavirus infection (COVID-19) and community-acquired pneumonia (TARGET-VIP). *Profil Med*. 2020;23(8):6-13.

5.	Forlenza OV, Damiano RF, Busatto GF, et al. Post-acute sequelae of SARS-CoV-2 infection (PASC): A protocol for a multidisciplinary prospective observational evaluation of a cohort of patients surviving hospitalisation in Sao Paulo, Brazil. *BMJ Open*. 2021;11(6):051706.

6.	Lyons J, Akbari A, Torabi F, et al. Understanding and responding to COVID-19 in Wales: Protocol for a privacy-protecting data platform for enhanced epidemiology and evaluation of interventions. *BMJ Open*. 2020;10(10):e043010.

7.	Molgaard Nielsen F, Klitgaard TL, Crescioli E, et al. Handling oxygenation targets in ICU patients with COVID-19 - protocol and statistical analysis plan in the HOT-COVID trial. *Acta Anaesthesiol Scand*. 2021.

8.	Rosa RG, Robinson CC, Veiga VC, et al. Quality of life and long-term outcomes after hospitalization for COVID-19: Protocol for a prospective cohort study (Coalition VII). *Rev Bras Ter Intensiva*. 2021;33(1):31-7.

9.	Rydwik E, Anmyr L, Regardt M, et al. ReCOV: recovery and rehabilitation during and after COVID-19 - a study protocol of a longitudinal observational study on patients, next of kin and health care staff. *BMC Sports Sci Med Rehabil*. 2021;13(1):70.

10.	Teece L, Gray LJ, Melbourne C, et al. United Kingdom Research study into Ethnicity And COVID-19 outcomes in Healthcare workers (UK-REACH): a retrospective cohort study using linked routinely collected data, study protocol. *BMJ Open*. 2021;11(6):e046392.

11.	Valk CMA, Swart P, Boers LS, et al. Practice of adjunctive treatments in critically ill COVID-19 patients-rational for the multicenter observational PRoAcT-COVID study in the Netherlands. *Ann Transl Med*. 2021;9(9):764.

**Relevant to KQ2 (n=3)**

1.	Lucidi D, Molinari G, Silvestri M, et al. Patient-reported olfactory recovery after SARS-CoV-2 infection: A 6-month follow-up study. *Int Forum Allergy Rhinol*. 2021;11(8):1249-52.

2.	Mokhtari M, Mohraz M, Gouya MM, et al. Clinical outcomes of patients with mild COVID-19 following treatment with hydroxychloroquine in an outpatient setting. *Int Immunopharmacol.* 2021;96:107636.

3.	Simplicio MIC, Ribeiro LB, Oliveira R, et al. Effect of Early Treatment with Hydroxychloroquine or Lopinavir and Ritonavir on Risk of Hospitalization among Patients with COVID-19: The TOGETHER Randomized Clinical Trial. *JAMA Netw Open*. 2021;4(4):e216468.

**Other (n=40)**

1.	Ayoubkhani D, Khunti K, Nafilyan V, et al. Post-covid syndrome in individuals admitted to hospital with covid-19: retrospective cohort study. *BMJ*. 2021;372:n693.

2.	Boari GEM, Turini D, Guarinoni V, et al. Prognostic factors and predictors of outcome in patients with COVID-19 and related pneumonia: a retrospective cohort study. *Biosci Rep*. 2020;40(12):BSR20203455.

3.	Briss PA, Richardson L, Wright J, Petersen R, Hacker KA. COVID-19 and Chronic Disease: The Impact Now and in the Future. *Prev Chronic Dis*. 2021;18:E62.

4.	Castro VM, Rosand J, Giacino JT, McCoy TH, Perlis RH. Case-control study of neuropsychiatric symptoms following COVID-19 hospitalization in 2 academic health systems. *medRxiv.* 2021.

5.	Chevinsky JR, Tao G, Lavery AM, et al. Late Conditions Diagnosed 1-4 Months Following an Initial Coronavirus Disease 2019 (COVID-19) Encounter: A Matched-Cohort Study Using Inpatient and Outpatient Administrative Data-United States, 1 March-30 June 2020. *Clin Infect Dis.* 2021;73:S5-S16.

6.	El Sayed S, Shokry D, Gomaa SM. Post-COVID-19 fatigue and anhedonia: A cross-sectional study and their correlation to post-recovery period. *Neuropsychopharmacol Rep*. 2021;41(1):50-5.

7.	Enghard P, Hardenberg J-H, Stockmann H, Hinze C, Eckardt K-U, Schmidt-Ott K. Long-term effects of COVID-19 on kidney function. *Lancet*. 2021;397(10287):1806-7.

8.	Griffin J, Henderson P, Strachan L, et al. Long-term Outcomes Following Severe COVID-19 Infection: A Multicenter Cohort Study of Family Member Outcomes. *Ann Am Thorac Soc*. 2021.

9.	Hampshire A, Trender W, Chamberlain SR, et al. Cognitive deficits in people who have recovered from COVID-19. *EClinicalMedicine*. 2021:101044.

10.	Hedberg P, Karlsson Valik J, Van Der Werff S, et al. Clinical phenotypes and outcomes of SARS-CoV-2, influenza, RSV and seven other respiratory viruses: A retrospective study using complete hospital data. *Thorax*. 2021.

11.	Hu B, Zhu Z, Li F, et al. Diffusion Capacity Abnormalities for Carbon Monoxide in Patients with COVID-19 At Three-Month Follow-up. *Eur Respir J*. 2021.

12.	Jeon J, Baruah G, Sarabadani S, Palanica A. Identification of risk factors and symptoms of COVID-19: Analysis of biomedical literature and social media data. *J Med Internet Res.* 2020;22(10):e20509.

13.	Lumlertgul N, Pirondini L, Cooney E, et al. Acute kidney injury prevalence, progression and long-term outcomes in critically ill patients with COVID-19: a cohort study. *Ann Intensive Care*. 2021;11(1):123.

14.	Mei Q, Wang F, Yang Y, et al. Health Issues and Immunological Assessment Related to Wuhan's COVID-19 Survivors: A Multicenter Follow-Up Study. *Front Med*. 2021;8:617689.

15.	Mei Q, Yuan X, Wang F, Wei L, Bryant A, Li J. Mental health problems among COVID-19 survivors in Wuhan, China. *World Psychiatry*. 2021;20(1):139-40.

16.	Nadjarian A, LeClair J, Mahoney TF, et al. Validation of a Crisis Standards of Care Model for Prioritization of Limited Resources During the Coronavirus Disease 2019 Crisis in an Urban, Safety-Net, Academic Medical Center. *Crit Care Med*. 2021.

17.	Perlis RH, Santillana M, Ognyanova K, et al. Comparison of post-COVID depression and major depressive disorder. *medRxiv.* 2021.

18.	Peters F, Behrendt C-A, Marschall U. Prevalence of COVID-19 Risk Factors and Risks of Severe Acute Respiratory Disease are Markedly Higher in Patients with Symptomatic Peripheral Arterial Occlusive Disease. *Eur J Vasc Endovasc Surg*. 2021;61(5):859-60.

19.	Pezzuto A, Tammaro A, Tonini G, Ciccozzi M. COPD influences survival in patients affected by COVID-19, comparison between subjects admitted to an internal medicine unit, and subjects admitted to an intensive care unit: An Italian experience. *J Med Virol*. 2021;93(3):1239-41.

20.	Pham T, Beduneau G, Guidet B, et al. Characteristics, management, and prognosis of elderly patients with COVID-19 admitted in the ICU during the first wave: insights from the COVID-ICU study: Prognosis of COVID-19 elderly critically ill patients in the ICU. *Ann Intensive Care*. 2021;11(1):77.

21.	Pinheiro MM, Pileggi GS, Reis-Neto E, et al. Incidence and risk factors for moderate/severe COVID-19 in rheumatic diseases patients on hydroxychloroquine: a 24-week prospective cohort. *Clin Exp Rheumatol*. 2021.

22.	Playan-Escribano J, Gomez-alvarez Z, Romero-Delgado T, Perez-Garcia C, Enriquez-Vazquez D, Vilacosta I. Cardiovascular comorbidity and death from covid-19: Prevalence and differential characteristics. *Cardiol J*. 2021;28(2):339-41.

23.	Rogers-Brown J, Wanga V, Okoro C, et al. Outcomes Among Patients Referred to Outpatient Rehabilitation Clinics After COVID-19 diagnosis - United States, January 2020-March 2021. *Morb Mortal Wkly Rep*. 2021;70(27).

24.	Saussez S, Sharma S, Thiriard A, et al. Predictive factors of smell recovery in a clinical series of 288 coronavirus disease 2019 patients with olfactory dysfunction. *Eur J Neurol*. 2021.

25.	Schmidt M, Hajage D, Demoule A, et al. Clinical characteristics and day-90 outcomes of 4244 critically ill adults with COVID-19: a prospective cohort study. *Intensive Care Med*. 2021;47(1):60-73.

26.	Shah W, Hillman T, Playford ED, Hishmeh L. Managing the long term effects of covid-19: Summary of NICE, SIGN, and RCGP rapid guideline. *BMJ*. 2021;372:n136.

27.	Shinohara T, Saida K, Tanaka S, Murayama A. Do lifestyle measures to counter COVID-19 affect frailty rates in elderly community dwelling? Protocol for cross-sectional and cohort study. *BMJ Open*. 2020;10(10):e040341.

28.	Tang L, Zeng X. One-year Follow-up after Severe SARS-CoV-2 Pneumonia. *Radiology*. 021;300(1):E310.

29.	Tao G, Lavery AM, Click ES, et al. Late Conditions Diagnosed 1-4 Months Following an Initial Coronavirus Disease 2019 (COVID-19) Encounter: A Matched-Cohort Study Using Inpatient and Outpatient Administrative Data-United States, 1 March-30 June 2020. *Clin Infect Dis*. 2021;73(1):S5-S16.

30.	Taquet M, Geddes JR, Husain M, Luciano S, Harrison PJ. 6-month neurological and psychiatric outcomes in 236 379 survivors of COVID-19: a retrospective cohort study using electronic health records. *Lancet Psychiatry*. 2021;8(5):416-27.

31.	Tedesco-Silva H, Medina-Pestana J, Requiao-Moura L, et al. High mortality among kidney transplant recipients diagnosed with coronavirus disease 2019: Results from the Brazilian multicenter cohort study. *PLoS ONE*. 2021;16(7):e0254822.

32.	Terrabuio DRB, Ferreira RMT, Cardoso AJA, et al. Insights in the approach of long-term liver transplant recipients with COVID-19. *Transpl Infect Dis*. 2021;23(1):e13424.

33.	Vaes AW, Delbressine JM, Houben-Wilke S, et al. Recovery from COVID-19: A sprint or marathon? 6-month follow-up data from online long COVID-19 support group members. *ERJ Open Res*. 2021;7(2):00141-2021.

34.	Venkatakrishnan AJ, Pawlowski C, Zemmour D, et al. Mapping each pre-existing condition's association to short-term and long-term COVID-19 complications. *NPJ Dig Med*. 2021;4(1):117.

35.	Welk B, Richard L, Braschi E, Averbeck MA. Is coronavirus disease 2019 associated with indicators of long-term bladder dysfunction? *Neurol Urodyn*. 2021;40(5):1200-6.

36.	Xiong L-J, Cao X-J, Xiong H-G, et al. Possible posttraumatic stress disorder in Chinese frontline healthcare workers who survived COVID-19 6 months after the COVID-19 outbreak: prevalence, correlates, and symptoms. *Transl Psychiatry*. 2021;11(1):374.

37.	Yang H, Xu J, Liang X, Shi L, Wang Y. Autoimmune diseases are independently associated with COVID-19 severity: Evidence based on adjusted effect estimates. *J Infect*. 2021;82(4):e23-e6.

38.	Yang H, Xu J, Liang X, Shi L, Wang Y. Chronic liver disease independently associated with COVID-19 severity: evidence based on adjusted effect estimates. *Hepatol Int*. 2021;15(1):217-22.

39.	Yasin R, Hassanein SA, Gomaa AAK, Ghazy T, Ibrahem RAl, Khalifa MH. Predicting lung fibrosis in post-COVID-19 patients after discharge with follow-up chest CT findings. *Egypt J Radiol Nucl Med*. 2021;52(1):118.

40.	Zampogna E, Ambrosino N, Saderi L, et al. Time course of exercise capacity in patients recovering from COVID-19-associated pneumonia. *J Bras Pneumol*. 2021;47(4):e20210076.

**Duplicates (n=2)**

1.	Hu B, Zhu Z, Li F, et al. Diffusion Capacity Abnormalities for Carbon Monoxide in Patients with COVID-19 At Three-Month Follow-up. *Eur Respir J*. 2021.

2.	Mandal S, Brill SE, Jarvis HC, et al. Long-COVID': A cross-sectional study of persisting symptoms, biomarker and imaging abnormalities following hospitalisation for COVID-19. *Thorax*. 2021;76(4):396-8.

**Excluded at DE, insufficient adjustment (n=6)**

1.	Aminian A, Bena J, Pantalone KM, Burguera B. Association of obesity with postacute sequelae of COVID-19. *Diabetes Obes Metab*. 2021.

2.	Augustin M, Schommers P, Stecher M, et al. Post-COVID syndrome in non-hospitalised patients with COVID-19: a longitudinal prospective cohort study. *Lancet.* 2021;6:100122.

3.	Estiri H, Strasser ZH, Brat GA, et al. Evolving Phenotypes of non-hospitalized Patients that Indicate Long Covid. *medRxiv*. 2021.

4.	Gunster C, Busse R, Spoden M, et al. 6-month mortality and readmissions of hospitalized COVID-19 patients: A nationwide cohort study of 8,679 patients in Germany. *PloS ONE.* 2021;16(8):e0255427.

5.	Mei Q, Wang F, Yang Y, et al. Health Issues and Immunological Assessment Related to Wuhan's COVID-19 Survivors: A Multicenter Follow-Up Study. *Front Med*. 2021;8:617689.

6.	Morin L, Savale L, Montani D, et al. Four-Month Clinical Status of a Cohort of Patients after Hospitalization for COVID-19. *JAMA*. 2021;325(15):1525-34.

**Author contact, unsuccessful (n=1)**

1.	Oh TK, Song I-A, Park HY. Risk of psychological sequelae among coronavirus disease-2019 survivors: A nationwide cohort study in South Korea. *Depress Anxiety.* 2021;38(2):247-54.

**SR included studies lists, excluded (n=17)**

1.	Bezzio C, Saibeni S, Variola A, et al. Outcomes of COVID-19 in 79 patients with IBD in Italy: an IG-IBD study. *Gut*. 2020;69(7):1213-7.

2.	Du RH, Liang LR, Yang CQ, et al. Predictors of mortality for patients with COVID-19 pneumonia caused by SARS-CoV-2: a prospective cohort study. *Eur Respir J*. 2020;55(5).

3.	Gregoriano C, Koch D, Haubitz S, et al. Characteristics, predictors and outcomes among 99 patients hospitalised with COVID-19 in a tertiary care centre in Switzerland: an observational analysis. *Swiss Med Wkly*. 2020;150:w20316.

4.	Lerum TV, Aaløkken TM, Brønstad E, et al. Dyspnoea, lung function and CT findings 3 months after hospital admission for COVID-19. *Eur Respir J*. 2021;57(4).

5.	Li X, Xu S, Yu M, et al. Risk factors for severity and mortality in adult COVID-19 inpatients in Wuhan. *J Allergy Clin Immunol*. 2020;146(1):110-8.

6.	Liang L, Yang B, Jiang N, et al. Three-month Follow-up Study of Survivors of Coronavirus Disease 2019 after Discharge*. J Korean Med Sci.* 2020;35(47):e418.

7.	Mandal S, Barnett J, Brill SE, et al. 'Long-COVID': a cross-sectional study of persisting symptoms, biomarker and imaging abnormalities following hospitalisation for COVID-19. *Thorax*. 2021;76(4):396-8.

8.	Merkler AE, Parikh NS, Mir S, et al. Risk of Ischemic Stroke in Patients With Coronavirus Disease 2019 (COVID-19) vs Patients With Influenza. *JAMA Neurol*. 2020;77(11):1-7.

9.	Moreno-Pérez O, Merino E, Leon-Ramirez JM, et al. Post-acute COVID-19 syndrome. Incidence and risk factors: A Mediterranean cohort study. *J Infect*. 2021;82(3):378-83.

10.	Nguyen NN, Hoang VT, Lagier J-C, Raoult D, Gautret P. Long-term persistence of olfactory and gustatory disorders in COVID-19 patients. *Clin Microbiol Infect*. 2021;27(6):931-2.

11.	Wang L, He W, Yu X, et al. Coronavirus disease 2019 in elderly patients: Characteristics and prognostic factors based on 4-week follow-up*. J Infect*. 2020;80(6):639-45.

12.	Wu C, Chen X, Cai Y, et al. Risk Factors Associated With Acute Respiratory Distress Syndrome and Death in Patients With Coronavirus Disease 2019 Pneumonia in Wuhan, China. *JAMA Intern Med*. 2020;180(7):934-43.

13.	Yang Q, Xie L, Zhang W, et al. Analysis of the clinical characteristics, drug treatments and prognoses of 136 patients with coronavirus disease 2019. *J Clin Pharm Ther*. 2020;45(4):609-16.

14.	Yang Q, Zhou Y, Wang X, et al. Effect of hypertension on outcomes of adult inpatients with COVID-19 in Wuhan, China: a propensity score-matching analysis. *Respir Res*. 2020;21(1):172.

15.	Yuan M, Yin W, Tao Z, Tan W, Hu Y. Association of radiologic findings with mortality of patients infected with 2019 novel coronavirus in Wuhan, China. *PLoS ONE*. 2020;15(3):e0230548.

16.	Zhang J, Wang X, Jia X, et al. Risk factors for disease severity, unimprovement, and mortality in COVID-19 patients in Wuhan, China. *Clin Microbiol Infect*. 2020;26(6):767-72.

17.	Zhao YM, Shang YM, Song WB, et al. Follow-up study of the pulmonary function and related physiological characteristics of COVID-19 survivors three months after recovery. *EClinicalMed*. 2020;25:100463.

**Excluded Studies List for Key Question 2**

**Not primary research (n=7)**

1.	Barone PD, Robert A. Convalescent plasma to treat coronavirus disease 2019 (COVID-19): considerations for clinical trial design. *Transfusion*. 2020;60(6):1123-7.

2.	Han GZ, Yi-Hua. Thinking more about therapy with convalescent plasma for COVID-19 patients. *Hum Vaccin Immunother*. 2020:2601-3.

3.	Joyner MJ, Senefeld JW, Klassen, SA, Fairweather D, Wright RS, Carter RE. In Reply - Limitations of Safety Update on Convalescent Plasma Transfusion in COVID-19 Patients. *Mayo Clin Proc*. 2020;95(12):2802-3.

4.	Kelly JR, Crockett MT, Alexander L, et al. Psychedelic science in post-COVID-19 psychiatry. *Ir J Psychol Med*. 2021;38(2):93-8.

5.	Leentjens JM, van Haaps TF, Wessels PF, Schutgens REG. COVID-19-associated coagulopathy and antithrombotic agents-lessons after 1 year. *Lancet Haematol.* 2021;8(7):e524-e33.

6.	Mussini CM, Falcone M, Nozza S, et al. Therapeutic strategies for severe COVID-19: a position paper from the Italian Society of Infectious and Tropical Diseases (SIMIT). *Clin Microbiol Infect*. 2021;27(3):389-95.

7.	Vink M, Vink-Niese A. Could Cognitive Behavioural Therapy Be an Effective Treatment for Long COVID and Post COVID-19 Fatigue Syndrome? Lessons from the Qure Study for Q-Fever Fatigue Syndrome. *Healthcare (Basel, Switzerland).* 2020;8(4):552.

**Language (n=8)**

1.	Gomez Herrero H, Caballero Garcia P, Vicaria Fernandez I, Galbete A. Residual lesions on chest-Xray after SARS-CoV-2 pneumonia: Identification of risk factors. Medicina clinica. 2021;S0025-7753(21):00205-0.

2.	Kovalchuk V. The role of the new coronavirus infection (COVID-19) in the progression and development of cerebrovascular diseases. A competent choice of pathogenic treatment is the key to success in treatment and prevention. An expert's view from the 'red zone'. *Nevrol Neiropsikhiatr Psikhosomatika.* 2021;13(1):57-66.

3.	Lopez-Barbeito B, Garcia-Martinex A, Coll-Vinent B, et al. Factors associated with revisits by patients with SARS-CoV-2 infection discharged from a hospital emergency department. *Emergencias : revista de la Sociedad Espanola de Medicina de Emergencias.* 2020;32(6):386-94.

4.	Marchenkova LA, Makarova EV, Yurova OV. The role of micronutrients in the comprehensive rehabilitation of patients with the novel coronavirus infection COVID-19. *Vopr Pitan*. 2021;90(2):40-9.

5.	Olenskaya TL, Nikolayeva AH, Piatsko VV, Azaronak MK, Yukhno YS. Application of altitude chamber adaptation as a component of the medical rehabilitation patients after COVID-19. *Profil Med.* 2021;24(4):76-82.

6.	Taboada M, Rodriguez N, Diaz-Vieito M, et al. [Quality of life and persistent symptoms after hospitalization for COVID-19. A prospective observational study comparing ICU with non-ICU Patients]. *Rev Esp Anestesiol Reanim*. 2021.

7.	Xiaofang G, Chunxia M, Yujie M, et al. Clinical efficacy and safety of different antiviral regimens in patients with coronavirus disease 2019. *Zhonghua Wei Zhong Bing Ji Jiu Yi Xue*. 2021;32(12):1423-7.

8.	Zolotovskaia IA, Shatskaia PR, Davydkin IL, Shavlovskaya OA. [Post-COVID-19 asthenic syndrome]. *Zh Nevrol Psikhiatr im S.S.Korsakova.* 2021;121(4):25-30.

**Protocol (n=36)**

1.	Bates A, Rushbrook S, Shapiro E, Grocott M, Cusack, R. CovEMERALD: Assessing the feasibility and preliminary effectiveness of remotely delivered Eye Movement Desensitisation and Reprocessing following Covid-19 related critical illness: A structured summary of a study protocol for a randomised controlled trial. *Trials*. 2020;21(1):929.

2.	Carvalho AC, Moreira J, Cubelo P, Cantista P, Branco CA, Guimaraes B. Therapeutic respiratory and functional rehabilitation protocol for intensive care unit patients affected by COVID-19: a structured summary of a study protocol for a randomised controlled trial. *Trials*. 2021;22(1):268.

3.	Daval M, Corre A, Palpacuer C, et al. Efficacy of local budesonide therapy in the management of persistent hyposmia in COVID-19 patients without signs of severity: A structured summary of a study protocol for a randomised controlled trial. *Trials*. 2020;21(1):666.

4.	Funk AL, Florin TA, Dalziel SR, et al. Prospective cohort study of children with suspected SARS-CoV-2 infection presenting to paediatric emergency departments: A Paediatric Emergency Research Networks (PERN) Study Protocol. *BMJ Open*. 2021;11(1):041711.

5.	Gao Y, Zhong LD, Quach B, et al. COVID-19 Rehabilitation With Herbal Medicine and Cardiorespiratory Exercise: Protocol for a Clinical Study. *JMIR Res Protoc*. 2021;10(5):e25556.

6.	Gorman E, Shankar-Hari M, Hopkins P, et al. Repair of Acute Respiratory Distress Syndrome by Stromal Cell Administration in COVID-19 (REALIST-COVID-19): A structured summary of a study protocol for a randomised, controlled trial. *Trials*. 2020;21(1):462.

7.	Grund S, Caljouw MAA, Haaksma ML, et al. Pan-European Study on Functional and Medical Recovery and Geriatric Rehabilitation Services of Post-COVID-19 Patients: Protocol of the EU-COGER Study. *J Nutr Health Aging.* 2021;25(5):668-74.

8.	Haran JP, Pinero JC, Zheng Y, Palma NA, Wingertzahn M. Virtualized clinical studies to assess the natural history and impact of gut microbiome modulation in non-hospitalized patients with mild to moderate COVID-19 a randomized, open-label, prospective study with a parallel group study evaluating the physiologic effects of KB109 on gut microbiota structure and function: a structured summary of a study protocol for a randomized controlled study. *Trials.* 2021;22(1):245.

9.	Benjamin K, Innes A, Holl M, et al. Scalable modEls of Community rehAbilitation for Individuals Recovering From COVID:19 reLated illnEss: A Longitudinal Service Evaluation Protocol-"SeaCole Cohort Evaluation". *Front Public Health.* 2021;9:628333.

10.	Janssen M, Schakel U, Foku CD, et al. A Randomized Open label Phase-II Clinical Trial with or without Infusion of Plasma from Subjects after Convalescence of SARS-CoV-2 Infection in High-Risk Patients with Confirmed Severe SARS-CoV-2 Disease (RECOVER): A structured summary of a study protocol for a randomised controlled trial. *Trials*. 2020;21(1):828.

11.	Verena K, Jensen B, Feldt T, et al. Reconvalescent plasma/camostat mesylate in early SARS-CoV-2 Q-PCR positive high-risk individuals (RES-Q-HR): a structured summary of a study protocol for a randomized controlled trial. *Trials*. 2021;22(1):343.

12.	Jieping L, Yang L, Wen G, Qumu S, Xiaoxia R, Yang T. Pulmonary Telerehabilitation and Efficacy among Discharged COVID-19 Patients: Rational and Design of a Prospective Real-world Study. *Clin Respir J.* 2021;15(11):1158-1167.

13.	Les Bujanda I, Loureiro-Amigo J, Bastons FC, et al. Treatment of COVID-19 pneumonia with glucocorticoids (CORTIVID): a structured summary of a study protocol for a randomised controlled trial. *Trials*. 2021;22(1):43.

14.	Jiawen L, Zang C, Wu Z, Wang G, Zhao H. The Mechanism and Clinical Outcome of patients with Corona Virus Disease 2019 Whose Nucleic Acid Test has changed from negative to positive, and the therapeutic efficacy of Favipiravir: A structured summary of a study protocol for a randomised controlled trial. *Trials*. 2020;21(1):488.

15.	Wu X, Wei L, Quan G, et al. Novel coronavirus pneumonia (COVID-19) combined with Chinese and Western medicine based on "Internal and External Relieving -Truncated Torsion" strategy. *Medicine*. 2020;99(51):e23874.

16.	Lu ZH, Yang CL, Yang GG, et al. Efficacy of the combination of modern medicine and traditional Chinese medicine in pulmonary fibrosis arising as a sequelae in convalescent COVID-19 patients: a randomized multicenter trial. *Infect Dis Poverty.* 2021;10(1):31.

17.	Lyngbakken MN, Berdal JE, Eskesen A, et al. Norwegian Coronavirus Disease 2019 (NO COVID-19) Pragmatic Open label Study to assess early use of hydroxychloroquine sulphate in moderately severe hospitalised patients with coronavirus disease 2019: A structured summary of a study protocol for a randomised controlled trial. *Trials*. 2020;21(1):485.

18.	Malaska J, Stasek J, Duska F, et al. Effect of dexamethasone in patients with ARDS and COVID-19 - prospective, multi-centre, open-label, parallel-group, randomised controlled trial (REMED trial): A structured summary of a study protocol for a randomised controlled trial. *Trials*. 2021;22(1):172.

19.	Misset B, Hoste E, Donneau AF, et al. A multicenter randomized trial to assess the efficacy of CONvalescent plasma therapy in patients with Invasive COVID-19 and acute respiratory failure treated with mechanical ventilation: the CONFIDENT trial protocol. *BMC Pulm Med*. 2020;20(1):317.

20.	Moreno-Gonzalez G, Mussetti A, Albasanz-Puig A, et al. A Phase I/II Clinical Trial to evaluate the efficacy of baricitinib to prevent respiratory insufficiency progression in onco-hematological patients affected with COVID19: A structured summary of a study protocol for a randomised controlled trial. *Trials*. 2021;22(1):116.

21.	Tan D, Chan AK, Juni P, et al. Post-exposure prophylaxis against SARS-CoV-2 in close contacts of confirmed COVID-19 cases (CORIPREV): study protocol for a cluster-randomized trial. *Trials.* 2021;22(1):224.

22.	Nanni O, Viale P, Vertogen B, et al.. PROTECT Trial: A cluster-randomized study with hydroxychloroquine versus observational support for prevention or early-phase treatment of Coronavirus disease (COVID-19): A structured summary of a study protocol for a randomized controlled trial. *Trials*. 2020;21(1):689.

23.	O'Brien H, Tracey MJ, Ottewill C, et al. An integrated multidisciplinary model of COVID-19 recovery care. *Ir J Med Sci*. 2021;190(2):461-8.

24.	Ojeda A, Calvo A, Cunat T, et al. Rationale and study design of an early care, therapeutic education, and psychological intervention program for the management of post-intensive care syndrome and chronic pain after COVID-19 infection (PAIN-COVID): study protocol for a randomized controlled trial. *Trials*. 2021;22(1):486.

25.	Payares-Herrera C, Martinez-Munoz ME, Valhonrat IL, et al. Double-blind, randomized, controlled, trial to assess the efficacy of allogenic mesenchymal stromal cells in patients with acute respiratory distress syndrome due to COVID-19 (COVID-AT): A structured summary of a study protocol for a randomised controlled trial. *Trials.* 2021;22(1):9.

26.	Petersen MW, Meyhoff TS, Helleberg M, et al. Low-dose hydrocortisone in patients with COVID-19 and severe hypoxia (COVID STEROID) trial-Protocol and statistical analysis plan. *Acta Anaesthesiol Scand*. 2020;64(9):1365-75.

27.	Rangnekar H, Patankar S, Suryawanshi K, Soni P. Safety and efficacy of herbal extracts to restore respiratory health and improve innate immunity in COVID-19 positive patients with mild to moderate severity: A structured summary of a study protocol for a randomised controlled trial. *Trials*. 2020;21(1):943.

28.	Rilinger J, Winfried VK, Duerschmied D, et al. A prospective, randomised, double blind placebo-controlled trial to evaluate the efficacy and safety of tocilizumab in patients with severe COVID-19 pneumonia (TOC-COVID): A structured summary of a study protocol for a randomised controlled trial. *Trials*. 2020;21(1):470.

29.	Rydwik E, Anmyr L, Regardt M, et al. ReCOV: recovery and rehabilitation during and after COVID-19 - a study protocol of a longitudinal observational study on patients, next of kin and health care staff. *BMC Sports Sci Med Rehabil*. 2021;13(1):70.

30.	McGregor G, Sandhu H, Bruce J, et al. Rehabilitation Exercise and psycholoGical support After covid-19 InfectioN' (REGAIN): a structured summary of a study protocol for a randomised controlled trial. *Trials*. 2021;22(1):8.

31.	Si MY, Xiao WJ, Pan C, et al. Mindfulness-based online intervention on mental health and quality of life among COVID-19 patients in China: an intervention design. *Infect Dis Poverty*. 2021;10(1):69.

32.	Sigfrid L, Cevik M, Jesudason E, et al. What is the recovery rate and risk of long-term consequences following a diagnosis of COVID-19? A harmonised, global longitudinal observational study protocol. *BMJ Open*. 2021;11(3):043887.

33.	Smith K, Pace A, Ortiz S, Kazani S, Rottinghaus S. A Phase 3 Open-label, Randomized, Controlled Study to Evaluate the Efficacy and Safety of Intravenously Administered Ravulizumab Compared with Best Supportive Care in Patients with COVID-19 Severe Pneumonia, Acute Lung Injury, or Acute Respiratory Distress Syndrome: A structured summary of a study protocol for a randomised controlled trial. *Trials*. 2020;21(1):639.

34.	Wright JK, Tan DHS, Walmsley SL, et al. Protecting Frontline Health Care Workers from COVID-19 with Hydroxychloroquine Pre-exposure Prophylaxis: A structured summary of a study protocol for a randomised placebo-controlled multisite trial in Toronto, Canada. *Trials*. 2020;21(1):647.

35.	Yadav B, Rai A, Mundada PS, et al. Safety and efficacy of Ayurvedic interventions and Yoga on long term effects of COVID-19: A structured summary of a study protocol for a randomized controlled trial. *Trials.* 2021;22(1):378.

36.	Ye Q, Wang H, Xia X, et al. Safety and efficacy assessment of allogeneic human dental pulp stem cells to treat patients with severe COVID-19: Structured summary of a study protocol for a randomized controlled trial (Phase I/II). *Trials*. 2020;21(1):520.

**Case reports/series ≤5 per group (n=3)**

1.	Vadasz I, Husain-Syed F, Dorfmuller P, et al. Severe organising pneumonia following COVID-19. Thorax. 2021;76(2):201-4.

2.	Unnimaya KL, Malavika TR, Sulfath TS, Ashique S. A case series of post covid lung fibrosis in dialysis patients; a prospective study. Int J Pharm Sci Rev Res. 2021;67(1):74-7.

3.	Wanschitz JV, Manuela K, Pfausler B, et al. Myasthenic crisis following SARS-CoV-2 infection and delayed virus clearance in a patient treated with rituximab: clinical course and 6-month follow-up. J Neurol. 2021;268(8):2700-2.

**Population not post-COVID (n=15)**

1.	Ajcevic M, Furlanis G, Maccarato M, et al. e-Health solution for home patient telemonitoring in early post-acute TIA/Minor stroke during COVID-19 pandemic. *Int J Med Inform*. 2021;152:104442.

2.	Altamura C, Cevoli S, Aurilla C, et al. Locking down the CGRP pathway during the COVID-19 pandemic lockdown: the PandeMig study. *Neurol Sci*. 2020;41(12):3385-9.

3.	Basar EZ, Sonmez HE, Oncel S, Yetimakman AF, Babaoglu K. Multisystemic inflammatory syndrome in children associated with covid-19: A single center experience in turkey. *Turk Pediatri Arsivi.* 2021;56(3):192-9.

4.	Beddok A, Chevrier M, Calugaru V, et al. Acute and late toxicities of patients infected with SARS-CoV-2 and treated for cancer with radiation therapy during the COVID-19 pandemic. *Int J Radiat Biol*. 2021:1-5.

5.	Carmi L, Ben-Arush O, Fostick L, Cohen H, Zohar J. Obsessive Compulsive Disorder during COVID-19 - two- and six-month follow-ups.OCD during COVID-19. *Int J Neuropsychopharmacol*. 2021;24(9):703-709.

6.	Bilir C, Cakir E, Gulbagci B, et al. COVID-19 prevalence and oncologic outcomes of asymptomatic patients with active cancer who received chemotherapy. *Acta Medica Mediterr*. 2021;37(1):667-71.

7.	BasaranNC, Uyaroglu OA, Dizman GT, et al. Outcome of noncritical COVID-19 patients with early hospitalization and early antiviral treatment outside the ICU. *Turk J Med Sci*. 2021;51(2):411-20.

8.	Lo Sasso B, Giglio RV, Vidali M, et al. Evaluation of anti-sars-cov-2 s-rbd igg antibodies after covid-19 mrna bnt162b2 vaccine. *Diagnostics*. 2021;11(7):1135.

9.	Jakubikova MT, Michaela/Rysankova, Irena/Novakova, Iveta/Doleckova, Kristyna/Dusek, Pavel/Pitha, Jiri/Tesar, Adam/Horakova, Magda/Vohanka, Stanislav/Bednarik, Josef/Vlazna, Daniela. Predictive factors for a severe course of COVID-19 infection in myasthenia gravis patients with an overall impact on myasthenic outcome status and survival. European Journal of Neurology. 2021doi: http://dx.doi.org/10.1111/ene.14951.

10.	Matheny ME, A. R./Druey, Kirk M./Maleque, Noble/Auld, Sara C./Channell, Natalie/Banerji, Aleena. Severe Exacerbations of Systemic Capillary Leak Syndrome After COVID-19 Vaccination: A Case Series. *Ann Intern Med*. 2021;174(10): 1476-1478.

11.	Namiki T, Takayama S, Arita R, et al. A structured summary of a study protocol for a multi-center, randomized controlled trial (RCT) of COVID-19 prevention with Kampo medicines (Integrative Management in Japan for Epidemic Disease by prophylactic study: IMJEDI P1 study). *Trials*. 2021;22(1):23.

12.	Pirjani R, Tahereh S, Dehpour AR, et al. Effect of hydroxychloroquine on prevention of COVID-19 virus infection among healthcare professionals: A structured summary of a study protocol for a randomised controlled trial. *Trials*. 2020;21(1):467.

13.	Rizzi S, Wensink MJ, Lindahl-Jacobsen R, Tian L, Lu Y, Eisenberg M. Risk of pre-term births and major birth defects resulting from paternal intake of COVID-19 medications prior to conception. *BMC Res Notes*. 2020;13(1):509.

14.	Shabaka A, Gruss E, Landaluce-Triska E, et al. Late thrombotic complications after SARS-CoV-2 infection in hemodialysis patients. *Hemodial Int*. 2021;25(4):507-514.

15.	Wichova H, Mia EM, Derebery MJ. Otologic Manifestations After COVID-19 Vaccination: The House Ear Clinic Experience. *Otol Neurotol*. 2021;42(9):e1213-e1218.

**Enrolment/intervention >8 wks post-Covid (n=11)**

1.	Albu S, Zozaya NR, Murillo N, Garcia-Molina A, Chacon C, Kumuru H. What's going on following acute COVID-19? Clinical characteristics of patients in an out-patient rehabilitation program. *NeuroRehabilitation*. 2021;48(4):469-80.

2.	Arnold DT, Milne A, Samms E, Stadon L, Maskell NA, Hamilton FW. Symptoms After COVID-19 Vaccination in Patients With Persistent Symptoms After Acute Infection: A Case Series. *Ann Intern Med*. 2021;174(9):1334-1336.

3.	Belcaro G, Umberto C, Cesarone MR, et al. Preventive effects of Pycnogenol on cardiovascular risk factors (including endothelial function) and microcirculation in subjects recovering from coronavirus disease 2019 (COVID-19). *Minerva Med*. 2021.

4.	D'Ascanio L, Vitelli F, Cingolani C, Maranzano M, Brenner MJ, Di Stadio A. Randomized clinical trial "olfactory dysfunction after COVID-19: Olfactory rehabilitation therapy vs. intervention treatment with Palmitoylethanolamide and Luteolin": Preliminary results. *Eur Rev Med Pharmacol Sci.* 2021;25(11):4156-62.

5.	Busatto GF, Ladeira de Araujo A, Duarte J, et al. Post-acute sequelae of SARS-CoV-2 infection (PASC): A protocol for a multidisciplinary prospective observational evaluation of a cohort of patients surviving hospitalisation in Sao Paulo, Brazil. *BMJ Open*. 2021;11(6):051706..

6.	Fortini A, Torrigiani A, Sbaragli S, et al. COVID-19: persistence of symptoms and lung alterations after 3-6 months from hospital discharge. *Infection*. 2021;49(5):1007-1015.

7.	Gerlis CC, Daynes E, Gerlis C, Chaplin E, Gardiner N, Singh SJ. Early experiences of rehabilitation for individuals post-COVID to improve fatigue, breathlessness exercise capacity and cognition - A cohort study. *Chron Respir Dis*. 2021;18:14799731211015691.

8.	Gloeckl R, Daniela L, Jarosch I, et al. Benefits of pulmonary rehabilitation in covid-19 - a prospective observational cohort study. *ERJ Open Res*. 2021;7(2).

9.	Liu M, Fajin L, Zheng Y, Xiao K. A prospective cohort study on radiological and physiological outcomes of recovered COVID-19 patients 6 months after discharge. *Quant Imaging Med Surg*. 2021;11(9):4181-92.

10.	Myall KJ, Mukherjee B, Castanheira AM, et al. Persistent post-COVID-19 interstitial lung disease: An observational study of corticosteroid treatment. *Ann Am Thorac Soc*. 2021;18(5):799-806.

11.	Zampogna E, Ambrosino N, Saderi L, et al. Time course of exercise capacity in patients recovering from COVID-19-associated pneumonia. *J Bras Pneumol.* 2021;47(4):e20210076.

**No intervention (n=63)**

1.	Hernandez-Piriz A, Tung-Chen Y, Jimminez-Virumbrales D, et al. Importance of lung ultrasound follow-up in patients who had recovered from coronavirus disease 2019: Results from a prospective study. *J Clin Med*. 2021;10(14):3196.

2.	Batisse DB, Gousseff M, Penot P, Gallay L, et al. Clinical recurrences of COVID-19 symptoms after recovery: Viral relapse, reinfection or inflammatory rebound? *J Infect*. 2020;81(5):816-46.

3.	Bedock D, Couffignal J, Lassen PB, et al. Evolution of nutritional status after early nutritional management in covid-19 hospitalized patients. *Nutrients.* 2021;13(7):2276.

4.	Bordin A, Mucignat-Caretta C, Gaudioso P, et al. Comparison of self-reported symptoms and psychophysical tests in coronavirus disease 2019 (COVID-19) subjects experiencing long-term olfactory dysfunction: a 6-month follow-up study. *Int Forum Allergy Rhinol*. 2021;11(11):1592-1595.

5.	Bourguignon A, Beaulieu C, Balkaid W, Desilets A, Blais N. Incidence of thrombotic outcomes for patients hospitalized and discharged after COVID-19 infection. *Thromb Res*. 2020;196:491-3.

6.	Boutou AK, Georgopoulou A, Pitsiou G, et al. Changes in the respiratory function of COVID-19 survivors during follow-up: A novel respiratory disorder on the rise? *Int J Clin Pract.* 2021.

7.	Broughan J, McCombe G, Avramovic G, et al. General practice attendances among patients attending a post-COVID-19 clinic: a pilot study. *BJGP Open*. 2021;5(3):1-7.

8.	Cao H, Lei R, Liu J, Liao W. The clinical characteristic of eight patients of COVID-19 with positive RT-PCR test after discharge. *J Med Virol*. 2020;92(10):2159-64.

9.	Crawford KHD, Dingens AS, Eguia R, et al. Dynamics of Neutralizing Antibody Titers in the Months After Severe Acute Respiratory Syndrome Coronavirus 2 Infection. *J Infect Dis*. 2021;223(2):197-205.

10.	Darley DR, Dore GJ, Cysique L, et al. Persistent symptoms up to four months after community and hospital-managed SARS-CoV-2 infection. *Med J Aust.* 2021;214(6):279-80.

11.	Davis HE, Gina S, McCorkell L, et al. Characterizing long COVID in an international cohort: 7 months of symptoms and their impact. *EClinicalMedicine*. 2021:101019.

12.	Debeaumont D, Boujibar F, Ferrand-Devogue E, et al. Cardiopulmonary Exercise Testing to Assess Persistent Symptoms at 6 Months in People With COVID-19 Who Survived Hospitalization: A Pilot Study. *Phys Ther*. 2021;101(6).

13.	Denina M, Giulia P, Carlo S, et al. Sequelae of COVID-19 in Hospitalized Children: A 4-Months Follow-Up. *J Pediatr Infect Dis*. 2020:E458-E9.

14.	Di Germanio C, Graham S, Kelly K, et al. SARS-CoV-2 antibody persistence in COVID-19 convalescent plasma donors: Dependency on assay format and applicability to serosurveillance. *Transfusion*. 2021;61(9):2677-2687.

15.	Divanoglou A, Samuelsson K, Sjodahl R, Andersson C, Levi R. Rehabilitation needs and mortality associated with the Covid-19 pandemic: a population-based study of all hospitalised and home-healthcare individuals in a Swedish healthcare region. *EClinicalMedicine*. 2021;36:100920.

16.	Engelen MM, Vandenbriele C, Balthazar T, et al. Venous Thromboembolism in Patients Discharged after COVID-19 Hospitalization. *Semin Thromb Hemost.* 2021;47(4):362-671.

17.	Erickson JL, Poterucha JT, Gende A, et al. Use of Electrocardiographic Screening to Clear Athletes for Return to Sports Following COVID-19 Infection. *Mayo Clin Proc*. 2021;5(2):368-76.

18.	Eser F, Bircan K, Guner R, et al. The Effect of prolonged PCR Positivity on patient Outcomes and Determination of Isolation period in COVID-19 patients. *Int J Clin Pract*. 2021;75(5):e14025.

19.	Garrigues E, Janvier P, Kherabi Y, et al. Post-discharge persistent symptoms and health-related quality of life after hospitalization for COVID-19. *J Infect*. 2020;81(6):e4-e6.

20.	Daynes E, Gerlis C, Briggs-Price S, Jones P, Singth SJ. COPD assessment test for the evaluation of COVID-19 symptoms. *Thorax*. 2021;76(2):185-7.

21.	Gharebaghi N, Farshid S, Boroofeh B, et al. Evaluation of epidemiology, clinical features, prognosis, diagnosis and treatment outcomes of patients with COVID-19 in West Azerbaijan Province. *Int J Clin Pract.* 2021;75(6):e14108.

22.	Giovannetti G, De Michele L, De Ceglie M, et al. Lung ultrasonography for long-term follow-up of COVID-19 survivors compared to chest CT scan. *Respir Med*. 2021;181:106384.

23.	Havervall S, Rosell A, Phillipson M, et al. Symptoms and Functional Impairment Assessed 8 Months After Mild COVID-19 Among Health Care Workers. *JAMA*. 2021;325(19):2015-6.

24.	Iftikhar H, Doherty WL, Sharp C. Long-term COVID-19 complications: a multidisciplinary clinic follow-up approach. *Clin Med (London, England).* 2021;21:3-4.

25.	Jack D, Damian D, Nolting A, Galazka A. COVID-19 in patients with multiple sclerosis treated with cladribine tablets: An update. *Mult Scler Relat Disord*. 2021;51:102929.

26.	Fitzpatrick V, Anne R, Christopher B, Kenneth C, Jon R.. Incidence of COVID-19 recurrence among large cohort of healthcare employees. *Ann Epidemiol*. 2021;60:8-14.

27.	Juncker HG, Romjin M, Loth VN, et al. Human Milk Antibodies Against SARS-CoV-2: A Longitudinal Follow-Up Study. *J Hum Lact*. 2021:8903344211030171.

28.	Labriola L, Scohy A, Seghers F, et al. A Longitudinal, 3-Month Serologic Assessment of SARS-CoV-2 Infections in a Belgian Hemodialysis Facility. *Clin J Am Soc Nephrol*. 2021;16(4):613-4.

29.	Lanham D, Roe J, Chauhan A, et al. Covid-19 emergency department discharges: An outcome study. *Clin Med*. 2021;21(2):E126-E31.

30.	Leite VFR, Danielle Bianchini/Jorge, Valeria Conceicao/de Lima, Maria do Carmo Correia/Cezarino, Leandro Goncalves/da Rocha, Cleber Nunes/Esper, Rodrigo Barbosa. Persistent Symptoms and Disability After COVID-19 Hospitalization: Data From a Comprehensive Telerehabilitation Program. *Arch Phys Med Rehabil.* 2021;102(7):1308-1316.

31.	Liu B, Yaling S, Zhang W, et al. Recovered COVID-19 patients with recurrent viral RNA exhibit lower levels of anti-RBD antibodies. *Cell Mol Immunol*. 2020;17(10):1098-100.

32.	Sergey T. L, Kirill/Tsaplin, Sergey/Schastlivtsev, Ilya/Zhuravlev, Sergey/Barinov, Victor/Caprini, Joseph A. The original and modified Caprini score equally predicts venous thromboembolism in COVID-19 patients. *J Vasc Surg Venous Lymphat Disord.* 2021;9(6):1371-1381.e4.

33.	Logue JK, Nicholas M, Franko BS, et al. Sequelae in Adults at 6 Months after COVID-19 Infection. *JAMA Netw Open*. 2021;4(2):e210830.

34.	Lund LC, Hallas J, Nielsen H, et al. Post-acute effects of SARS-CoV-2 infection in individuals not requiring hospital admission: a Danish population-based cohort study. *Lancet Infect Dis.* 2021;21(10):1373-1382.

35.	Malek LA, Marczak M, Milosz-Wieczorek B, et al. Cardiac involvement in consecutive elite athletes recovered from Covid-19: A magnetic resonance study. *J Magn Reson Imaging*. 2021;53(6):1723-9.

36.	Kerr C, Hughes G, McKenna L, Bergin C. Prevalence of smell and taste dysfunction in a cohort of CoVID19 outpatients managed through remote consultation from a large urban teaching hospital in Dublin, Ireland. *Infect Prev Pract*. 2020;2(3):100076.

37.	Nehme M, Braillard O, Alcoba G, et al. COVID-19 Symptoms: Longitudinal Evolution and Persistence in Outpatient Settings. *Ann Int Med*. 2021;174(5):723-5.

38.	Oygar PD, Ozsurekci Y, Gurlevik SL, et al. Longitudinal Follow-up of Antibody Responses in Pediatric Patients with COVID-19 up to 9 Months after Infection. *Pediatr Infect Dis J*. 2021:E294-E9.

39.	Pimlott NA, Agarwal P, McCarthy LM, et al. Clinical learnings from a virtual primary care program monitoring mild to moderate COVID-19 patients at home. *Fam Pract*. 2020;38(5):549-555.

40.	Plassmeyer M, Alpan O, Corley MJ, et al. Caspases and therapeutic potential of caspase inhibitors in moderate-severe SARS CoV2 infection and long COVID. *Allergy.* 2021;77(1):118-129.

41.	Raad RA, Ganti A, Goshtasbi K, et al. Temporal patterns of nasal symptoms in patients with mild severity SARS-CoV-2 infection. *Am J Otolaryngol*. 2021;42(6):103076.

42.	Ray A, Chaudhry R, Rai S, et al. Prolonged Oxygen Therapy Post COVID-19 Infection: Factors Leading to the Risk of Poor Outcome. *Cureus*. 2021;13(2):e13357.

43.	Gianella P, Rigamonti E, Marando M, et al. Clinical, radiological and functional outcomes in patients with SARS-CoV-2 pneumonia: a prospective observational study. *BMC Pulm Med.* 2021;21(1):136.

44.	Rosales-Castillo A, Rios C, Garica JD. [Persistent symptoms after acute COVID-19 infection: importance of follow-up]. *Med Clin*. 2021;156(1):35-6.

45.	Samimagham HR, Mohsen A, Hooshyar D, Kazemijahromi M. The association of non-steroidal anti-inflammatory drugs with COVID-19 severity and mortality. *Arch Clin Infect Dis*. 2020;15(4):1-5.

46.	Secchi M, Bazzigaluppi E, Brigatti C, et al. COVID-19 survival associates with the immunoglobulin response to the SARS-cov-2 spike receptor binding domain. *J Clin Invest*. 2020;130(12):6366-78.

47.	Wong AW, Shah AS, Johnston JC, Carlsten C, Ryerson CJ. Patient-reported outcome measures after COVID-19: A prospective cohort study. *Eur Respir J*. 2020;56(5):2003276.

48.	Shendy W, Ezzat MM, E’Laidy DA, Elsherif AA. Prevalence of fatigue in patients post Covid-19. *Eur J Mol Clin Med*. 2021;8(3):1330-40.

49.	Lee RJ, Wysocki O, Bhogal T, et al. Longitudinal characterisation of haematological and biochemical parameters in cancer patients prior to and during COVID-19 reveals features associated with outcome. *ESMO Open*. 2021;6(1):100005.

50.	Singhania SVK, Simon C, Abhijit R, Nikhil P. Pulmonary sequelae of moderate-to-severe COVID pneumonia, a 3-month follow-up study. *Lung India*. 2021;38(4):397-9.

51.	Skjorten I, Ankerstjerne OAW, Trebinjac D, et al. Cardiopulmonary exercise capacity and limitations 3 months after COVID-19 hospitalisation. *Eur Respir J*. 2021;59(1).

52.	Sibila O, Albacar N, Perea L, et al. Lung Function sequelae in COVID-19 Patients 3 Months After Hospital Discharge. *Arch Bronconeumol*. 2021;57:59-61.

53.	Nunez-Cortez R, Rivera-Lillo G, Arias-Campoverde M, Soto-Garcia D, Garcia-Palomera R, Torrres-Castro R. Use of sit-to-stand test to assess the physical capacity and exertional desaturation in patients post COVID-19. *Chron Respir Dis*. 2021;18.

54.	van den Borst B, Peters JB, Brink M, et al. Comprehensive health assessment three months after recovery from acute COVID-19. *Clin Infect Dis.* 2020;73(5):e1089-e1098.

55.	van den Heuvel FMA, Vos JL, van Bakel B, et al. Comparison between myocardial function assessed by echocardiography during hospitalization for COVID-19 and at 4 months follow-up. *Int J Cardiovascular Imaging*. 2021;37(12):3459-3467.

56.	Varghese J, Sandmann S, Ochs K, et al. Persistent symptoms and lab abnormalities in patients who recovered from COVID-19. *Sci Rep*. 2021;11(1):12775.

57.	von Meijenfeldt F, Havervall S, Adelmeijer J, et al. Sustained prothrombotic changes in COVID-19 patients 4 months after hospital discharge. *Blood Adv*. 2021;5(3):756-9.

58.	Wang H, Yuan Y, Xiao M, et al. Dynamics of the SARS-CoV-2 antibody response up to 10 months after infection. *Cell Mol Immunol*. 2021;18(7):1832-4.

59.	Wang H, Ruili L, Zhou Z, et al. Cardiac involvement in COVID-19 patients: mid-term follow up by cardiovascular magnetic resonance. *J Cardiovasc Magn Res*. 2021;23(1):14.

60.	Yan X, Huang H, Wang C, et al. Follow-up study of pulmonary function among COVID-19 survivors 1 year after recovery. *J Infect*. 2021;83(3):381-412.

61.	Younis SS, Savastano A, Crincoli E, Savastano MC, et al. Peripapillary retinal vascular involvement in early post-covid-19 patients. *J Clin Med*. 2020;9(9):1-16.

62.	Yu X. Impact of mitigating interventions and temperature on the instantaneous reproduction number in the COVID-19 pandemic among 30 US metropolitan areas. *One Health*. 2020;10:100160.

63.	Zhang D, Zhang C, Li X, et al. Thin-section computed tomography findings and longitudinal variations of the residual pulmonary sequelae after discharge in patients with COVID-19: a short-term follow-up study. *Eur Radiol*. 2021;31(9):7172-7183.

**Wrong intervention (n=10)**

1.	Pampols PA, Trujillo H, Melilli E, et al. Immunosuppression minimization in kidney transplant recipients hospitalized for COVID-19. *Clin Kidney J*. 2021;14(4):1229-35.

2.	Damanti S, Ramirez GA, Bozzolo EP, et al. 6-Month Respiratory Outcomes and Exercise Capacity of COVID-19 Acute Respiratory Failure Patients Treated With CPAP. *Intern Med J*. 2021; 10.1111/imj.15345.

3.	Islam MS, Ferdous MZ, Islam US, Mosaddek AS, Potenza MN, Pardhan S. Treatment, persistent symptoms, and depression in people infected with covid-19 in Bangladesh. *Int J Environ Res Public Health*. 2021;18(4):1-16.

4.	Jaywant A, Vanderlind WM, Alexopoulos GS, Fridman CB, Perlis RH, Gunning FM. Frequency and profile of objective cognitive deficits in hospitalized patients recovering from COVID-19. *Neuropsychopharmacol*. 2021;46:2235-2240.

5.	Le Bon S-D, Konopnicki D, Pisarski N, Prunier L, Lechien JR, Horoi M. Efficacy and safety of oral corticosteroids and olfactory training in the management of COVID-19-related loss of smell. *Eur Arch Oto-Rhino-Laryngol.* 2021;278(8):3113-7.

6.	Maestrini V, Birtolo LI, Francone M, et al. Cardiac involvement in consecutive unselected hospitalized COVID-19 population: In-hospital evaluation and one-year follow-up. *Int J Cardiol*. 2021;339:235-242.

7.	Mitra S, Janweja M, Sengupta A. Post-COVID-19 rhino-orbito-cerebral mucormycosis: a new addition to challenges in pandemic control. *Eur Arch Oto-rhino-laryngol.* 2021;1-6.

8.	Mohamad SA, Badawi AM, Mansour HF. Insulin fast-dissolving film for intranasal delivery via olfactory region, a promising approach for the treatment of anosmia in COVID-19 patients: Design, in-vitro characterization and clinical evaluation. *Int J Pharm*. 2021;601:120600.

9.	Uz C, Umay E, Gundogdu I, Uz FB, Bahtiyarca ZT. Factors affecting medium-term aerobic capacity in COVID-19 patients. *Acta Medica Mediterr*. 2021;37(2):847-53.

10.	Zhang S, Wenxue B, Yue J, et al. Eight months follow-up study on pulmonary function, lung radiographic, and related physiological characteristics in COVID-19 survivors. *Sci Rep*. 2021;11(1):13854.

**Active comparator (n=4)**

1.	Lee K-M, Ko H-J, Lee GH, Kim A-S, Lee D-W. A well-structured follow-up program is required after recovery from coronavirus disease 2019 (Covid-19); release from quarantine is not the end of treatment. *J Clin Med*. 2021;10(11):2329.

2.	Moreno Diaz R, Garcia MAM, Munoz FJT, et al. Does timing matter on tocilizumab administration? Clinical, analytical and radiological outcomes in COVID-19. *Eur J Hosp Pharm*. 2021:2020002669.

3.	Rutsch M, Frommhold J, Buhr-Schinner H, et al. Study protocol medical rehabilitation after COVID-19 disease: an observational study with a comparison group with obstructive airway disease / Re_Co. BMC Health Serv Res. 2021;21(1):373.

4.	Sakai T, Hoshino C, Yamaguchi R, Hirao M, Nakahara R, Okawa A. Remote rehabilitation for patients with COVID-19. *J Rehabil Med*. 2020;52(9):jrm00095.

**No/wrong outcome (n=10)**

1.	Acosta-Ampudia Y, Monsalve DM, Rojas M, et al. COVID-19 convalescent plasma composition and immunological effects in severe patients. *J Autoimmun*. 2021;118:102598.

2.	An Y-W, Yuan B, Wang J-C, et al. Clinical characteristics and impacts of traditional Chinese medicine treatment on the convalescents of COVID-19. Int *J Med Sci*. 2021;18(3):646-51.

3.	Archer SK, Iezzi CM, Gilpin L. Louisa. Swallowing and Voice Outcomes in Patients Hospitalized With COVID-19: An Observational Cohort Study. *Arch Phys Med Rehabil*. 2021;102(6):1084-90.

4.	Lee KS, Talenfeld AD, Browne WF, et al. Role of interventional radiology in the treatment of COVID-19 patients: Early experience from an epicenter. *Clin Imaging*. 2021;71:143-6.

5.	Guiling L, Du L, Cao X, et al. Follow-up study on serum cholesterol profiles and potential sequelae in recovered COVID-19 patients. *BMC Infect Dis*. 2021;21(1):299.

6.	Liu C, Dun Y, Liu P, et al. Associations of medications used during hospitalization and immunological changes in patients with COVID-19 during 3-month follow-up. *Int Immunopharmacol.* 2020;89:107121.

7.	Lucidi D, Molinari G, Silvestri M, et al. Patient-reported olfactory recovery after SARS-CoV-2 infection: A 6-month follow-up study. *Int Forum Allergy Rhinol*. 2021;11(8):1249-1252.

8.	Omma A, Abdulsamet E, Guven SC, Ates I, Kucuksahin O. Convalescent Plasma Reduces Endogenous Antibody Response in COVID-19: A Retrospective Cross-Sectional Study. *Turk J Haematol*. 2021;38(4):321-324.

9.	Parry AH, Wani AH, Shah NN, Jehangir M. Medium-term chest computed tomography (CT) follow-up of COVID-19 pneumonia patients after recovery to assess the rate of resolution and determine the potential predictors of persistent lung changes. *Egypt J Radiol Nucl Med*. 2021;52(1):55.

10.	Wu Q, Hou X, Li H, et al. A follow-up study of respiratory and physical function after discharge in patients with redetectable positive SARS-CoV-2 nucleic acid results following recovery from COVID-19. *Int J Infect Dis*. 2021;107:5-11.

**Outcome(s) <12 wks post-Covid (n=61)**

1.	Abdelalim AA, Mohamady AA, Elsayed RA, Elawady MA, Ghallab AF. Corticosteroid nasal spray for recovery of smell sensation in COVID-19 patients: A randomized controlled trial. *Am J Otolaryngol*. 2021;42(2):102884.

2.	Ahmed I, Inam AB, Belli S, Ahmad J, Khalil W, Jafar MM. Effectiveness of aerobic exercise training program on cardio-respiratory fitness and quality of life in patients recovered from COVID-19. *Eur J Physiother*. 2021

3.	Alemanno F, Houdayer E, Parma A, et al. COVID-19 cognitive deficits after respiratory assistance in the subacute phase: A COVID rehabilitation unit experience. *PLoS ONE*. 2021;16(2):e0246590.

4.	Jordan SC, Zakowski P, Tran HP, et al. Compassionate Use of Tocilizumab for Treatment of SARS-CoV-2 Pneumonia. *Clin Infect Dis*. 2020;71(12):3168-73.

5.	Arnold DT, Hamilton FW, Milne A, et al. Patient outcomes after hospitalisation with COVID-19 and implications for follow-up: Results from a prospective UK cohort. *Thorax*. 2021;76(4):399-401.

6.	Attia ZI, Kapa S, Noseworthy PA, Lopez-Jiminez F, Friedman PA. Artificial Intelligence ECG to Detect Left Ventricular Dysfunction in COVID-19: A Case Series. *Mayo Clin Proc*. 2020;95(11):2464-6.

7.	Barratt-Due A, Olsen IC, Nezvalova-Henriksen K, et al. Evaluation of the Effects of Remdesivir and Hydroxychloroquine on Viral Clearance in COVID-19 : A Randomized Trial. *Ann Intern Med*. 2021doi: https://dx.doi.org/10.7326/M21-0653.

8.	Bottino I, Patria MF, Milani GP, et al. Can Asymptomatic or Non-Severe SARS-CoV-2 Infection Cause Medium-Term Pulmonary Sequelae in Children? *Front Pediatr*. 2021;9:621019.

9.	Bowles KH, McDonald M, Barron Y, Kennedy E, O’Connor M, Mikkelsen M. Surviving COVID-19 After Hospital Discharge: Symptom, Functional, and Adverse Outcomes of Home Health Recipients. *Ann Intern Med*. 2021;174(3):316-25.

10.	Caballero Bermejo AF, Ruiz-Antoran B, Cruz AF, et al. Sarilumab versus standard of care for the early treatment of COVID-19 pneumonia in hospitalized patients: SARTRE: a structured summary of a study protocol for a randomised controlled trial. *Trials*. 2020;21(1):794.

11.	Canziani LM, Trovati S, Brunetta E, et al. Interleukin-6 receptor blocking with intravenous tocilizumab in COVID-19 severe acute respiratory distress syndrome: A retrospective case-control survival analysis of 128 patients. *J Autoimmun*. 2020;114:102511.

12.	Cao J, Zheng X, Wei W, et al. Three-month outcomes of recovered COVID-19 patients: prospective observational study. *Ther Adv Respir Dis*. 2021;15:17534666211009410.

13.	Cravedi P, Mothi SS, Azzi Y, et al. COVID-19 and kidney transplantation: Results from the TANGO International Transplant Consortium. *Am J Transplant*. 2020;20(11):3140-8.

14.	Declercq J, Bosteels C, Damme KV, et al. Zilucoplan in patients with acute hypoxic respiratory failure due to COVID-19 (ZILU-COV): A structured summary of a study protocol for a randomised controlled trial. *Trials.* 2020;21(1):934.

15.	Dheir H, Sipahi S, Yaylaci S, et al. Clinical course of COVID-19 disease in immunosuppressed renal transplant patients. *Turk J Med Sci*. 2021;51(2):428-34.

16.	Di Pietro DA, Comini L, Gazzi L, Luisa A, Vitacca M. Neuropsychological pattern in a series of post-acute COVID-19 patients in a rehabilitation unit: Retrospective analysis and correlation with functional outcomes. *Int J Environ Res Public Health.* 2021;18(11):5917.

17.	Matalon N, Dorman-Ilan S, Hasson-Ohayon I, et al. Trajectories of post-traumatic stress symptoms, anxiety, and depression in hospitalized COVID-19 patients: A one-month follow-up. *J Psychosom Res*. 2021;143:110399.

18.	Duvignaud A, Lhomme E, Pistone T, et al. Home Treatment of Older People with Symptomatic SARS-CoV-2 Infection (COVID-19): A structured Summary of a Study Protocol for a Multi-Arm Multi-Stage (MAMS) Randomized Trial to Evaluate the Efficacy and Tolerability of Several Experimental Treatments to Reduce the Risk of Hospitalisation or Death in outpatients aged 65 years or older (COVERAGE trial). *Trials.* 2020;21(1):846.

19.	Herrman ML, Hahn J-M, Walter-Frank B, et al.COVID-19 in persons aged 70+ in an early affected German district: Risk factors, mortality and post-COVID care needs-A retrospective observational study of hospitalized and non-hospitalized patients. *PLoS ONE*. 2021;16(6):e0253154.

20.	Hannah CR, Blyth KG, Burley G, et al. Glasgow Early Treatment Arm Favirpiravir (GETAFIX) for adults with early stage COVID-19: A structured summary of a study protocol for a randomised controlled trial. *Trials*. 2020;21(1):935.

21.	Franchini M, Glingani C, Morandi M, et al. Safety and Efficacy of Convalescent Plasma in Elderly COVID-19 Patients: The RESCUE Trial. *Mayo Clin Proc Innov Qual Outcomes*. 2021;5(2):403-12.

22.	Ghati N, Roy A, Bhatnagar S, et al. Atorvastatin and Aspirin as Adjuvant Therapy in Patients with SARS-CoV-2 Infection: A structured summary of a study protocol for a randomised controlled trial. *Trials*. 2020;21(1):902.

23.	Goldman DL, Aldrich ML, Hagmann SH, et al. Compassionate use of remdesivir in children with severe covid-19. *Pediatrics.* 2021;147(5):e2020047803.

24.	Gong Y, Guan L, Jin Z, Chen S, Xiang G, Gao B. Effects of methylprednisolone use on viral genomic nucleic acid negative conversion and CT imaging lesion absorption in COVID-19 patients under 50 years old. *J Med Virol*. 2020;92(11):2551-5.

25.	Ramakrishnan S, Nicolau Jr DV, Langford B, et al. Inhaled budesonide in the treatment of early COVID-19 (STOIC): a phase 2, open-label, randomised controlled trial. *Lancet Respir Med.* 2021;9(7):763-772.

26.	Hayward G, Butler C, Yu L-M, et al. Platform Randomised trial of INterventions against COVID-19 in older peoPLE (PRINCIPLE): Protocol for a randomised, controlled, open-label, adaptive platform, trial of community treatment of COVID-19 syndromic illness in people at higher risk. *BMJ Open*. 2021;11(6).

27.	Hermann M, Pekacka-Egli A-M, Witassek F, et al. Feasibility and Efficacy of Cardiopulmonary Rehabilitation After COVID-19. *Am J Phys Med Rehabil*. 2020;99(10):865-9.

28.	Salisbury R, Iotchkova V, Jaafar S, et al. Incidence of symptomatic, image-confirmed venous thromboembolism following hospitalization for COVID-19 with 90-day follow-up. *Blood Adv*. 2020;4(24):6230-9.

29.	Kartsios C, Lokare A, Osman H, et al. Diagnosis, management, and outcomes of venous thromboembolism in COVID-19 positive patients: a role for direct anticoagulants? *J Thromb Thrombolysis*. 2021;51(4):947-52.

30.	Paterson RW, Brown RL, Benjamin L, et al. The emerging spectrum of COVID-19 neurology: Clinical, radiological and laboratory findings. *Brain*. 2020;143(10):3104-20.

31.	Khan FR, Kazmi S, Iqbal NT, Iqbal J, Ali ST, Abbas SA. A quadruple blind, randomised controlled trial of gargling agents in reducing intraoral viral load among hospitalised COVID-19 patients: A structured summary of a study protocol for a randomised controlled trial. *Trials*. 2020;21(1):785.

32.	Puchner B, Sahanic S, Kirchmair R, et al. Beneficial effects of multi-disciplinary rehabilitation in postacute COVID-19: an observational cohort study. *Eur J Phys Rehabil Med*. 2021;57(2):189-98.

33.	Kireyev IV, Zhabotynska NV, Vladimirova IM, Ocheredko LV. Prevention of Asthenic Syndrome as Concomitant Circumstains in Post-Covid-19 Patients. *Wiadomosci Lek.* 2021;74(5):1104-8.

34.	Kulkarni S, Fisk M, Kostapanos M, et al. Repurposed immunomodulatory drugs for Covid-19 in pre-ICU patients - mulTi-Arm Therapeutic study in pre-ICu patients admitted with Covid-19 - Repurposed Drugs (TACTIC-R): A structured summary of a study protocol for a randomised controlled trial. *Trials*. 2020;21(1):626.

35.	Kumar S, De Souza R, Nadkar M, et al. A two-arm, randomized, controlled, multi-centric, open-label phase-2 study to evaluate the efficacy and safety of Itolizumab in moderate to severe ARDS patients due to COVID-19. *Expert Opin Biol Ther*. 2021;21(5):675-86.

36.	Li L, Zhang W, Hu Y, et al. Effect of Convalescent Plasma Therapy on Time to Clinical Improvement in Patients with Severe and Life-threatening COVID-19: A Randomized Clinical Trial. *JAMA*. 2020;324(5):460-70.

37.	Loinaz C, Marcacuzco A, Fernandez-Ruiz M, et al. Varied clinical presentation and outcome of SARS-CoV-2 infection in liver transplant recipients: Initial experience at a single center in Madrid, Spain. *Transpl Infect Dis*. 2020;22(5):e13372.

38.	Lugon CC, Smit M, Salamun J, et al. Novel outpatient management of mild to moderate COVID-19 spares hospital capacity and safeguards patient outcome: The Geneva PneumoCoV-Ambu study. *PLoS ONE*. 2021;16(3):e0247774.

39.	Lui DTW, Lee CH, Chow WS, et al. Long COVID in Patients with Mild to Moderate Disease: Do Thyroid Function and Autoimmunity Play a Role? *Endocr Pract*. 2021;27(9):894-902.

40.	Mahapatra S, Rattan R, Mohanty CBK. Convalescent Plasma Therapy in the management of COVID-19 patients-The newer dimensions. *Transfus Clin Biol*. 2021;28(3):246-253.

41.	Maksimov VA, Torshin IY, Chuchalin AG, et al. An experience of using Laennec in patients at high risk of a cytokine storm with COVID-19 and hyperferritinemia. *Pulmonologiya*. 2020;30(5):587-98.

42.	Maniscalco M, Ambrosino P, Fuschillo S, et al. Bronchodilator reversibility testing in post-COVID-19 patients undergoing pulmonary rehabilitation. *Respir Med*. 2021;182:106401.

43.	Yuksel M, Akturk H, Mizikoglu O, Toroslu E, Arikan C. A single-center report of COVID-19 disease course and management in liver transplanted pediatric patients. *Pediatr Transplant.* 2021;25(7):e14061.

44.	Olds H, Liu J, Luk K, Lim HW, Ozog D, Rambhatla PV. Telogen effluvium associated with COVID-19 infection. *Dermatol Ther*. 2021;34(2):e14761.

45.	Olezene CS, Hansen E, Steere HK, et al. Functional outcomes in the inpatient rehabilitation setting following severe COVID-19 infection. *PloS ONE*. 2021;16(3):e0248824. do

46.	Panda PK, Bandyopadhyay A, Singh BC, et al. Safety and efficacy of antiviral combination therapy in symptomatic patients of Covid-19 infection - a randomised controlled trial (SEV-COVID Trial): A structured summary of a study protocol for a randomized controlled trial. *Trials*. 2020;21(1):866.

47.	Piquet V, Luczak C, Seiler F, et al. Do Patients With COVID-19 Benefit from Rehabilitation? Functional Outcomes of the First 100 Patients in a COVID-19 Rehabilitation Unit. *Arch Phys Med Rehabil.* 2021;102(6):1067-74.

48.	Santeusanio AD, Menon M, Liu C, et al. Influence of patient characteristics and immunosuppressant management on mortality in kidney transplant recipients hospitalized with coronavirus disease 2019 (COVID-19). *Clin Transplant.* 2021;35(4):e14221.

49.	Spielmanns M, Pekacka-Egli A-M, Schoendorf S, Windisch W, Hermann M. Effects of a comprehensive pulmonary rehabilitation in severe post-covid-19 patients. *Int J Environ Res Public Health*. 2021;18(5):1-14.

50.	Sehgal K, Fadel HJ, Tande AJ, Pardi DS, Khannah S. Outcomes in Patients with SARS-CoV-2 and Clostridioides difficile Coinfection. *Infect Drug Resist*. 2021;14:1645-8.

51.	Joyner MJ, Carter RE, Senefeld JW, et al. Convalescent plasma antibody levels and the risk of death from covid-19. *N Engl J Med.* 2021;384(11):1015-27.

52.	Shah FA, Chaudhry ZR, Rasheed S, Shakir S, Rashid E, Rasheed F. Deltacortil (prednisolone) relieves symptoms of corona virus (COVID-19) infection. *Pak J Med Health Sci*. 2020;14(3):1052-328.

53.	Reis G, Silva E, Silva D, et al. Effect of Early Treatment with Hydroxychloroquine or Lopinavir and Ritonavir on Risk of Hospitalization among Patients with COVID-19: The TOGETHER Randomized Clinical Trial. *JAMA Netw Open.* 2021;4(4):e216468.

54.	Sohrabpour S, Heidari F, Ansari R, Tajdini A, Heidari F. Subacute Thyroiditis in COVID-19 Patients. *Eur Thyroid J*. 2021;9(6):321-3.

55.	Floencio FKZ, Tenorio M, Macedo A, Lima S. Aspirin with or without statin in the treatment of endotheliitis, thrombosis, and ischemia in coronavirus disease. *Rev Soc Bras Med Trop*. 2020;53:1-5.

56.	Trunfio M, Salvador E, Gaviraghi A, et al. Early low-molecular-weight heparin administration is associated with shorter time to SARS-CoV-2 swab negativity. *Antivir Ther*. 2021;25(6):327-33.

57.	Vaira LA, Hopkins C, Petrocelli M, et al. Efficacy of corticosteroid therapy in the treatment of long- lasting olfactory disorders in COVID-19 patients. *Rhinology.* 2021;59(1):21-5.

58.	Wang M, Zhao Y, Hu W, et al. Treatment of COVID-19 Patients with Prolonged Post-Symptomatic Viral Shedding with Leflunomide -- a Single-Center, Randomized, Controlled Clinical Trial. *Clin Infect Dis*. 2020;73(11):e4012-e4019.

59.	Yamamoto S, Saito M, Nagai E, et al. Seroconversion against SARS-CoV-2 occurred after the recovery in patients with COVID-19. *J Med Virol*. 2021;93(2):692-4.

60.	Yu M, Liu Y, Zhang R, Lan L, Xu H. Prediction of the development of pulmonary fibrosis using serial thin-section ct and clinical features in patients discharged after treatment for COVID-19 pneumonia. *Korean J Radiol*. 2020;21(6):746-55.

61.	Zha L, Xu X, Wang D, Qiao G, Zhuang W, Huang S. Modified rehabilitation exercises for mild cases of COVID-19. *Ann Palliat Med*. 2020;9(5):3100-6.

**No outcome data (n=1)**

1.	Tawfik HM, Shaaban HS, Tawfik AM. Post-covid-19 syndrome in egyptian healthcare staff: Highlighting the carers sufferings. *Electron J Gen Med.* 2021;18(3):em291.

**Other (n=3)**

1.	Kingstone T, Taylor AK, O’Donnell CA, Atherson H, Blane DN, Chew-Graham CA. Finding the 'right' GP: a qualitative study of the experiences of people with long-COVID. *BJGP Open*. 2020;4(5):1-12.

2.	Perreault J, Tremblay T, Fournier M-J, et al. Waning of SARS-CoV-2 RBD antibodies in longitudinal convalescent plasma samples within 4 months after symptom onset. *Blood*. 2020;136(22):2588-91.

3.	Sivan M, Halpin S, Hollingworth L, Snook N, Hickman K, Clifton I. Development of an integrated rehabilitation pathway for individuals recovering from COVID-19 in the community. *J Rehabil Med.* 2020;52(8):jrm00089.

**Author contact, unsuccessful (n=2)**

1.	Mokhtari M, Mohraz M, Gouya MM, et al. Clinical outcomes of patients with mild COVID-19 following treatment with hydroxychloroquine in an outpatient setting. *Int Immunopharmacol.* 2021;96:107636.

2.	Rogers-Brown JS, Wanga V, Okoro C, et al. Outcomes Among Patients Referred to Outpatient Rehabilitation Clinics After COVID-19 diagnosis - United States, January 2020-March 2021. *MMWR Morb Mort Wkly Rep*. 2021;70(27).

**Author contact, ineligible timing or no details (n=6)**

1.	Ferrario SR, Panzeri A, Cerutti P, Sacco D. The psychological experience and intervention in post-acute COVID-19 inpatients. *Neuropsychiatr Dis Treat*. 2021;17:413-22.

2.	Ferraro F, Calafiore D, Dambruoso F, Guidarini S, de Sire A. COVID-19 related fatigue: Which role for rehabilitation in post-COVID-19 patients? A case series. *J Med Virol*. 2021;93(4):1896-9.

3.	Hameed F, Palatulan E, Jaywant A, et al. Outcomes of a COVID-19 recovery program for patients hospitalized with SARS-CoV-2 infection in New York City: A prospective cohort study. *PM R*. 2021;13(6):609-17.

4.	Giannis D, Allen SL, Tsang J, et al. Postdischarge thromboembolic outcomes and mortality of hospitalized patients with COVID-19: the CORE-19 registry. *Blood.* 2021;137(20):2838-47.

5.	Vaes AW, Goertz YM, Herck MV, et al. Recovery from COVID-19: A sprint or marathon? 6-month follow-up data from online long COVID-19 support group members. *ERJ Open Res*. 2021;7(2):00141-2021.

6.	Weinbergerova B, Mayer J, Hrabovsky S, et al. COVID-19's natural course among ambulatory monitored outpatients. *Sci Rep*. 2021;11(1):10124.

**SR included studies lists, excluded (n=1)**

1.	Frontera JA, Ariane L, Kara M, et al. Prevalence and Predictors of Prolonged Cognitive and Psychological Symptoms Following COVID-19 in the United States. *Front Aging Neurosci*. 2021;13.

**All specific population eg in ICU (n=32)**

1.	Al Chikhanie Y, Veale D, Choeffler M, Pepin JL, Verges S, Herengt F. Effectiveness of pulmonary rehabilitation in COVID-19 respiratory failure patients post-ICU. *Respir Physiol Neurobiol.* 2021;287:103639.

2.	Alharthy A, Abuhamdah M, Balhamar A, et al. Residual Lung Injury in Patients Recovering From COVID-19 Critical Illness: A Prospective Longitudinal Point-of-Care Lung Ultrasound Study. *J Ultrasound Med*. 2020;40(9):1823-1838.

3.	Schandl A, Hedman A, Lynga P, et al. Long-term consequences in critically ill COVID-19 patients: A prospective cohort study. *Acta Anaesthesiol Scand*. 2021;65(9):1285-1292.

4.	Pancera S, Bianchi LNC, Porta R, Galeri S, Carrozza MC, Villafane JH. Feasibility of subacute rehabilitation for mechanically ventilated patients with COVID-19 disease: a retrospective case series. *Int J Rehabil Res*. 2021;44(1):77-81.

5.	Boers NSB, Botta M, Tsonas AM, et al. PRactice of VENTilation in Patients with Novel Coronavirus Disease (PRoVENT-COVID): rationale and protocol for a national multicenter observational study in The Netherlands. *Ann Transl Med*. 2020;8(19):1251.

6.	Fang J, Li R, Chen Y, et al. Extracorporeal Membrane Oxygenation Therapy for Critically Ill Coronavirus Disease 2019 Patients in Wuhan, China: A Retrospective Multicenter Cohort Study. *Curr Med Sci*. 2021;41(1):1-13.

7.	Cour MA, Amaz C, Bohe J, Rimmele T, Ovize M, Argaud L. Day-90 survival in critically-ill patients with COVID-19 and hydroxychloroquine: A propensity analysis. *Ann Transl Med*. 2021;9(7):A1.

8.	Curci C, Negrini F, Ferrillo M, et al. Functional outcome after inpatient rehabilitation in postintensive care unit COVID-19 patients: findings and clinical implications from a real-practice retrospective study. *Eur J Phys Rehabil Med*. 2021;57(3):443-50.

9.	Curci C, Pisano F, Bonacci E, et al. Early rehabilitation in post-acute COVID-19 patients: data from an Italian COVID-19 Rehabilitation Unit and proposal of a treatment protocol. *Eur J Phys Rehabil Med*. 2020;56(5):633-41.

10.	Boers NS, Botta M, Tsonas AM, et al. Practice of ventilation in patients with novel coronavirus disease (provent-covid): Rationale and protocol for a national multicenter observational study in the netherlands. *Ann Transl Med*. 2020;8(19):1251.

11.	Doyle AJ, Thomas W, Retter A, et al. Updated hospital associated venous thromboembolism outcomes with 90-days follow-up after hospitalisation for severe COVID-19 in two UK critical care units. *Thromb Res* 2020;196:454-6.

12.	Dreier EM, Malfertheiner MV, Dienemann T, et al. ECMO in COVID-19-prolonged therapy needed? A retrospective analysis of outcome and prognostic factors. *Perfusion.* 2021;36(6):582-591.

13.	Hassenpflug MS, Jun D, Nelson DR, Dolinay T. Post-COVID recovery: Characteristics of chronically critically ill patients admitted to a long-term acute care hospital. *F1000Res*. 2021;9:1241.

14.	Imamura M, Mirisola AR, Ribeiro F, et al. Rehabilitation of patients after COVID-19 recovery: An experience at the Physical and Rehabilitation Medicine Institute and Lucy Montoro Rehabilitation Institute. *Clinics*. 2021;76:e2804.

15.	Biancari F, Mariscalco G, Dalen M, et al. Six-Month Survival After Extracorporeal Membrane Oxygenation for Severe COVID-19. *J Cardiothorac Vasc Anesth*. 2021;35(7):1999-2006.

16.	Kotani T, Sugiyama M, Matsuzaki F, et al. Roles of early mobilization program in preventing muscle weakness and decreasing psychiatric disorders in patients with coronavirus disease 2019 pneumonia: A retrospective observational cohort study. *J Clin Med*. 2021;10(13):2941.

17.	Lallana S, Chen A, Requena M, et al. Posterior reversible encephalopathy syndrome (PRES) associated with COVID-19. *J Clin Neurosci*. 2021;88:108-12.

18.	Li Y, Meng Q, Rao X, et al. Corticosteroid therapy in critically ill patients with COVID-19: a multicenter, retrospective study. *Crit Care*. 2020;24(1):698.

19.	Magomedov A, Zickler D, Karaivanov S, et al. Viscoelastic testing reveals normalization of the coagulation profile 12 weeks after severe COVID-19. *Sci Rep*. 2021;11(1):13325.

20.	Nikam PP, Varadharajulu G. Effect of twist and raise walking technique on icu-acquired weakness in COVID-19 patients: a pre-post experimental study. *J Ecophysiol Occup Health*. 2020;20(3-4):155-8.

21.	Olivares-Gazca JC, Priesca-Marin JM, Ojeda-Laguana M, et al. Infusion of Convalescent Plasma is Associated with Clinical Improvement in Critically Ill Patients with Covid-19: a Pilot Study. *Rev Invest Clin*. 2020;72(3):159-64.

22.	Pantos C, Kostopanagiotou G, Armaganidis A, Trikas A, Tseti I, Mourouzis I. Triiodothyronine for the treatment of critically ill patients with COVID-19 infection: A structured summary of a study protocol for a randomised controlled trial. *Trials*. 2020;21(1):573.

23.	Parker AJ, Humbir A, Tiwary P, et al. Recovery after critical illness in COVID-19 ICU survivors. *Br J Anaesth*. 2021;126(6):e217-e9.

24.	Bikdeli B, Talasaz AH, Rashidi F, et al. Intermediate-Dose versus Standard-Dose Prophylactic Anticoagulation in Patients with COVID-19 Admitted to the Intensive Care Unit: 90-Day Results from the INSPIRATION Randomized Trial. *Thromb Haemost*. 2021.

25.	Eriksson KE, Campoccia-Jalde F, Rysz S, Rimes-Stigare C. Continuous renal replacement therapy in intensive care patients with COVID-19; survival and renal recovery. *J Crit Care.* 2021;64:125-30.

26.	Stockmann H, Hardenberg JH, Aigner A, et al. High rates of long-term renal recovery in survivors of coronavirus disease 2019-associated acute kidney injury requiring kidney replacement therapy. *Kidney Int*. 2021;99(4):1021-2.

27.	Valk CMA, Swart P, Boers LS, et al. Practice of adjunctive treatments in critically ill COVID-19 patients-rational for the multicenter observational PRoAcT-COVID study in the Netherlands. *Ann Transl Med*. 2021;9(9):764.

28.	van Gassel RJJ, Bels J, Remij L, et al. Functional Outcomes and Their Association With Physical Performance in Mechanically Ventilated Coronavirus Disease 2019 Survivors at 3 Months Following Hospital Discharge: A Cohort Study. *Crit Care Med*. 2021;49(10):1726-1738.

29.	Van Zeller C, Anwar A, Ramos-Bascon N, Barnes N, Madden B. Pulmonary function, computerized tomography features and six-minute walk test at three months in severe COVID-19 patients treated with intravenous pulsed methylprednisolone: a preliminary report. *Monaldi Arch Chest Dis*. 2021.

30.	Maskin LP, Olarte GL, Palizas Jr F, et al. High dose dexamethasone treatment for Acute Respiratory Distress Syndrome secondary to COVID-19: A structured summary of a study protocol for a randomised controlled trial. *Trials*. 2020;21(1):743.

31.	Klein MN, Wang EW, Zimand P, et al. Kinetics of SARS-CoV-2 antibody responses pre-COVID-19 and post-COVID-19 convalescent plasma transfusion in patients with severe respiratory failure: An observational case-control study. *J Clin Pathol*. 2021:207356.

32.	Wiertz CMH, Vints WAJ, Maas GJC, et al. COVID-19: Patient Characteristics in the First Phase of Postintensive Care Rehabilitation. *Arch Rehabil Res Clin Transl*. 2021;3(2):100108.

**Table S3. Study Characteristics for Key Question 1**

| **Study** | **Population** | **Risk factor** | **Outcome ^ʃ^** | **Method** |
| --- | --- | --- | --- | --- |
| Author & year;  Country;  Design;  Funding source | N analyzed and population;  Covid-19 ascertainment %;  Age (yr);  Male %;  SES^*^ | Relevant risk factors;  Data source | Relevant outcomes;  Outcome definition;  Assessment tool  Assessment timing; | Analysis method;  Variables adjusted for |
| **Non-hospitalized** |  |  |  |  |
| **Nehme 2021**  Ref ID: 2919  Switzerland  Prospective cohort  No funding | 410 persons who tested positive for Covid-19  100% RT-PCR  Age, mean (SD): 42.7 (12.9)  33% male  Profession: 26.3% healthcare worker; 73.7% non-healthcare worker | Age (18-39 [reference]; 40-59; ≥60)  Sex ^ǂ^  Self-reported | **Recovery/nonrecovery:** fatigue, anosmia and/or ageusia, dyspnea, headache, cough, digestive symptoms, and fever  **Fatigue:** ECOG performance scale (range: 0= no limitation to 4= completely disabled, totally confined to bed or chair)  **Dyspnea:** mMRC scale  **Cognitive impairment**  Difficulty concentrating (self-reported)  Memory loss (self-reported)  7-9 months post-diagnosis | Study reported stratified analysis only; estimates obtained using the marginal prediction of a logistic regression adjusted for age, sex, and the number of symptoms at baseline, corrected for attrition using inverse probability weighting. |
| **Stavem 2021**  Ref ID: 3870  Norway  Retrospective cohort  No funding | 458 non-hospitalized people who tested positive for Covid-19  100% PCR  Age, mean (SD): 49.5 (15.3)  44% male  Education: 53% university; 38% secondary school; 9% primary school  Ethnicity: 84% born in Norway | Age (per 10 yr)  Sex  BMI (kg/m^2^)  Number of comorbidities (0 [reference], 1, ≥2)  Previous depression  Number of Covid-19 symptoms (0-5 [reference], 6-9, 10-23)  Self-reported (mixed-mode survey, postal and web) | **Fatigue:** CFQ-11 fatigue scale (defined as ≥4)  Median 4 months after first symptoms | Multivariate logistic regression  Adjusted for age, sex, marital status, education, BMI, comorbidities, previous depression, number of Covid-19 symptoms, dyspnea and confusion during Covid-19, smoking, time since symptom onset |
| **Boscolo-Rizzo 2021**  From Ref ID: 1307  Italy  Prospective cohort  No funding | 304 mild-to-moderate symptomatic adult patients  100% RT-PCR  Age, median (range): 47.0 (18.0-76.0)  39% male  SES NR | Age (<40 [reference], 40-54, ≥55)  Sex  BMI (≥25 vs <25 [reference])  Comorbidity (any vs 0)  Number of symptoms (≤2 [reference], 3-7, 8+)  Self-reported (phone interview) | **Recovery/nonrecovery:** Persistent symptoms at 12-month follow-up (ad-hoc questions and structured questionnaire)  12 months after disease onset | Unconditional multivariate logistic regression  Adjusted for age, sex, number of symptoms during acute phase |
| **Hospitalized** |  |  |  |  |
| **Frontera 2021**  Ref ID: 1394  USA  Bidirectional cohort  Non-industry funding | 382 Covid-19 patients hospitalized with (cases) and without (controls) new neurological complications during index hospitalization  100% RT-PCR  Age, median: 68.5  65% male  Race: 43% White; 13% Black; 7% Asian | Age (per yr)  Race/ethnicity (White vs non-White)  History of atrial fibrillation (yes vs no)  History of dementia (yes vs no)  Baseline modified Rankin score, 0-5  SOFA illness severity score, worst and IQR  Hypotension requiring vasopressors and intubation (yes vs no)  Intubation (yes vs no)  Medical records | **Fatigue:** worse than average (T-score >50) based on Neuro-QoL short form self-reported measure  **Anxiety:** worse than average (T-score >50) based on Neuro-QoL short form self-reported measure  **Depression:** worse than average (T-score >50) based on Neuro-QoL short form self-reported measure  **Functional capacity:** modified Rankin scale (0 no symptom to 6 dead; cut-off NR)  **Cognitive impairment:** MoCA (≤18 abnormal cognition, 22 perfect score)  **Sleep quality:** worse than average (T-score >50) based on Neuro-QoL short form self-reported measure  **Return to work:** resumed work either in person or remotely, even if they changed employers (self-reported)  Median 6.7 months after onset of neurological symptoms (among cases) or onset of Covid-19 symptoms (among controls) | Ordinal logistic regression  Adjusted for, by outcome:  Fatigue (cases and controls matched on age, sex, baseline illness severity, hypotension requiring vasopressors and intubation)  Psychopathology – anxiety, depression (cases and controls matched on age, sex, baseline illness severity)  Cognitive impairment (age, race, education)  Functional capacity (neurological complications during hospitalization; age; baseline disability score; hospital length of stay)  Cognitive impairment (age, race, education)  Sleep quality (cases and controls were matched on age, sex, baseline illness severity, intubation)  Return to work (neurological complications during hospitalization, worst SOFA score, hospital length of stay, race) |
| **Liu 2021**  Ref ID: 2408  China  Retrospective cohort  Non-industry funding | 1,539 inpatients aged >60 yr discharged from hospital  100% WHO interim guidance  Age, median (IQR): 69.0 (66.0-75.0)  48% male  Education, median (IQR): 12 (9-12) years | Age (yr)  Sex  COPD (yes vs no)  Hypertension (yes vs no)  Covid-19 illness severity (severe vs non-severe)  Medical records and family members | **Cognitive decline, longitudinal:** Informant Questionnaire on Cognitive Decline in the Elderly (IQCODE) short form, Chinese version: score ≥3.5 cognitive decline  6 months after discharge | Multivariate logistic regression  Adjusted for, by outcome:  Cognitive status, decline (age, sex, education, severity, ICU admission, high flow oxygen therapy, delirium, diabetes, stroke, coronary heart disease, hypertension) |
| **Osmanov 2021**  Ref ID: 2736  Russia  Bidirectional cohort  No funding | 518, children (≤18 yr) previously hospitalized with Covid-19  100% RT-PCR  Age, median (IQR): 10.4 (3.0-15.2)  48% male  SES NR | Age (<2 reference, 2-5, 6-11, 12-18 yr)  Sex  Excessive weight and obesity (yes vs no)  Allergic disease (includes any of: asthma, allergic rhinitis, eczema, food allergy; yes vs no)  Pneumonia during Covid-19 (yes vs no)  Severe acute Covid-19 (having received invasive/non-invasive ventilation, or admission to PICU; yes vs no)  Medical records and parent interview | **Recovery/nonrecovery:** Persistent symptoms present at the time of the follow-up interview and lasting for over 5 months, assessed using ISARIC Covid-19 Health and Wellbeing Follow-Up Survey for Children; symptoms subcategories: respiratory, neurological, sensory, sleep, gastrointestinal, dermatological, cardiovascular, fatigue and musculoskeletal  Median 8.5 months after hospital discharge | Multivariate logistic regression without imputation for missing data  Adjusted for age, sex, comorbidities (neurological conditions, allergies, GI problems, excessive weight and obesity), pneumonia during Covid-19, severe acute Covid-19 |
| **Munblit 2021**  Ref ID: 2850  Russia  Bidirectional cohort  Non-industry funding | 2,649 previously hospitalized patients with Covid- 19  51% clinical diagnosis and RT-PCR  Age, median (IQR): 56.0 (46.0-66.0)  49% male  Employment: 57.5% working full-time; 34% retired/early retirement due to illness  Education: 77.4% university; 9.5% school | Age (continuous)  Sex  The following also include data for subset of lab-confirmed patients:  Obesity (yes vs no)  Asthma (yes vs no)  Diabetes (yes vs no)  Chronic cardiac disease (yes vs no)  Cardiac revascularization (yes vs no)  Chronic pulmonary disease (yes vs no)  Hypertension (yes vs no)  Malignant neoplasm (yes vs no)  Rheumatologic disorder (yes vs no)  Acute Covid-19 severity (mild [reference], moderate, severe)  Medical records | **Recovery/nonrecovery:** symptoms present since hospital discharge only, assessed using Tier 1 ISARIC Long- term Follow- up Study case report form (CRF); symptoms subcategories: respiratory, gastrointestinal, dermatological, chronic fatigue, neurological, mood and behavior, sensory  Median 7.3 months after hospital discharge | Multivariate logistic regression  Adjusted for age, sex, comorbidities, severity, RT-PCR |
| **Qin 2021**  Ref ID: 3276  China  Prospective cohort  Industry and non-industry funding | 647 hospitalized Covid-19 patients  100% RT-PCR  Age, mean (SD): 58.0 (15.0)  44% male  SES NR | Age (yr)  Sex  Severe pneumonia (yes vs no)  Data source NR | **Recovery/nonrecovery:** weakness, fatigue, palpitation, dyspnea, and chest pain were included among other symptoms)  3 months after hospital discharge | Multivariate logistic regression  Adjusted for age, sex, severe pneumonia, lab values, in-patient days |
| **Qu 2021**  Ref ID: 3279  China  Bidirectional cohort  Non-industry | 540 COVID-19 patients discharged from hospitals  100% PCR  Age, median (IQR): 47.5 (37.0-57.0)  50% male  SES NR | Age (< 60 vs ≥ 60)  Sex  Covid-19 severity (severe vs mild/moderate)  Data source NR | **Functional capacity:** assessed using SF-36 physical component summary (PCS) subscale, ranging from 0-100 (higher scores represent better quality)  Month 3 after discharge | Multivariate logistic regression  Adjusted for age, sex, physical symptoms (univariate estimates reported for Covid-19 severity) |
| **Shang 2021**  Ref ID: 3696  China  Retrospective cohort  Funding NR | 796 severe or critically ill Covid-19 patients who were recovered and discharged from hospital  100% RT-PCR  Age, median (IQR): 62.0 (51.0-69.0)  51% male  SES NR | Age (>65 vs ≤ 65)  Sex  Severe/critical acute Covid-19 (yes vs no)  Intubation ( yes vs no)  ICU admission (yes vs no)  Data source NR | **Recovery/nonrecovery:** self-reported; included among others: shortness of breath, chest pains, developing new hypertension, muscle pain, sleep disorder, fatigue, mental symptoms  **Fatigue:** self-reported, details not provided  **Sleep disorder:** self-reported, details not provided  **Hypomnesis:** self-reported, details not provided  6 months after discharge | Multivariate logistic regression  Adjusted for age, sex, Covid-19 severity/critical illness, ICU admission, intubation, in-hospital days |
| **Huang 2021**  Ref ID: 4558  China  Bidirectional cohort  Non-industry | 1,733 adult patients with confirmed Covid-19 discharged from hospital  100% confirmed Covid-19 (test NR)  Age, median (IQR): 57.0 (47.0-65.0)  52% male  Education: 68% middle school/lower; 32% college/higher | Age (per year)  Sex  Comorbidity (yes vs no)  Disease severity, range: scale 1 (i.e. not admitted to hospital) to scale 7 (i.e. death) (scale 3 [reference], scale 4, scale 5-6)  Electronic medical records and self-reported comorbidities | **Anxiety/depression:** self-reported using EuroQol five-dimension five-level (EQ-5D-5L), responses include no, slight, moderate, severe, and extreme anxiety/depression  **Fatigue/muscle weakness:** self-reported using symptom questionnaire (compared with pre-Covid-19)  Median 186 (175-199) days after symptom onset | Multivariate logistic regression  Adjusted for age, sex, smoking, education, comorbidities, disease severity, corticosteroids, antiviral, intravenous immunoglobulins |
| **Sigfrid 2021**  Identified from grey literature search  UK  Prospective cohort  Non-industry | 327 adults who were admitted to hospital with confirmed or highly suspected SARS-CoV-2 infection  98.7% RT-PCR  Age, median (IQR): 59.7 (51.7-67.7)  59% male  Ethnicity: 81% White; 4.6% Black; 6% other minorities | Age and sex interaction (male \| < 50 yrs [reference], male \| 50-69 yrs, male \| ≥ 70 yrs, female \| < 50 yrs, female \| 50-69 yrs, female \| ≥ 70 yrs)  Comorbidity (1 or more vs none)  Covid-19 severity (scale 3 i.e. not requiring supplemental O2 [reference], scale 4 i.e. requiring supplemental O2, scale 5 i.e. requiring high flow nasal cannula or noninvasive ventilation, scale 6 or 7 i.e. requiring invasive mechanical ventilation or critical care)  NR | **Recovery/nonrecovery: :** self-reported using symptom questionnaire at 3 to 12 months following initial Covid-19 symptoms  **Dyspnea:** Breathlessness on daily activities measured using the Medical Research Council (MRC) dyspnea scale (1= no breathlessness, 5= unable to undertake), analyzed as change in value reported by participants before Covid-19 onset compared to follow-up assessment  **Fatigue (continuous):** on a 1 to 10 visual analogue scale (VAS), 0= no fatigue and 10= worst possible fatigue  **Quality of Life (continuous):** EuroQol EQ5D-5L instrument (EQ5D-5L) summary index change (0= worse health, 1= perfect health)  **Disability:** New or worsened disability assessed using the Washington Disability Group (WG) Short Form on functioning (vision, hearing, mobility, cognition, self-care, communication)  Median 222 (IQR: 189-269; range: 112-343) days from symptom onset | Multivariate logistic (for dichotomous outcomes) and linear (for continuous outcomes) regression  Adjusted for age-sex interaction variable, comorbidities, acute Covid-19 severity |
| **Hospitalized and non-hospitalized** |  |  |  |  |
| **Blomberg 2021**  Ref ID: 513  Norway  Prospective cohort  Non-industry | 312 home-isolated and hospitalized Covid-19 patients  100% RT-PCR  Age, median (IQR): 46.0 (30.0-58.0)  49% male  SES NR | Age (median)  Sex  BMI (median)  Diabetes (yes vs no)  Asthma/COPD (yes vs no)  Hypertension (yes vs no)  Chronic heart disease (yes vs no)  Rheumatic disease (yes vs no)  Dyspnea (yes vs no)  Covid-19 illness severity (median, range from 0-8)  Self-reported | **Fatigue:** total bimodal score of ≥4 on 11 questions (Chalder fatigue score)  **Fatigue (continuous):** Chalder fatigue scale ranging from 0 (no fatigue) to 33 9worst possible fatigue) (diabetes, bypertension, and Rheumatic disease)  6 (±1) months after initial Covid-19 | Multivariate logistic regression (for dichotomous outcome) and negative binomial regression (for continuous outcome)  Adjusted for, by outcome:  Fatigue dichotomous (age, sex, BMI, comorbidities, days in hospital, symptoms at onset, Covid-19 severity, antibodies, antibiotic use)  Fatigue continuous (age, sex, BMI, other comorbidities, illness severity, antibodies at 2 months) |
| **Kayaaslan 2021**  Ref ID: 2048  Turkey  Retrospective cohort  Funding NR | 1,007 patients with Covid‐19 followed in hospital or outpatient clinics  PCR on a nasopharyngeal sample and/or typical pulmonary involvement on computed tomography (% NR)  Age, mean (SD): 45.0 (16.4)  54% male  Employment: 68.5% employed | At least one comorbidity (yes vs no)  Disease severity (i.e. severe/critical); yes vs no)  Hospitalization (yes vs no)  Patient interview and hospital automation system | **Recovery/nonrecovery:** symptoms (fatigue, pain, concentration/memory deficit, new onset anxiety and depression) persisting beyond 12 weeks after recovery; each symptom assessed using scale of Covid‐19 Yorkshire Rehabilitation Screening (C19‐YRS) Tool ranging from 0 (no problem) to 10 (extreme problem)  Median 20 (IQR: 19-22) weeks after first Covid-19 diagnosis. | Multivariate logistic regression  Adjusted for hospitalization, comorbidity, illness severity |
| **Menges 2021**  Ref ID: 2697  Switzerland  Retrospective cohort  Industry and non-industry | 431 adults with positive Covid-19 within contact tracing database  100% PCR  Age, median (IQR): 47.0 (33.0-58.0)  50% male  Employment: 75.4% employed; 3.3% students  Education: 43.5% vocational training/specialized baccalaureate | Age (18-39 [reference], 40- 64, ≥ 65)  Sex  Employment (employed [reference], student, retired, unemployed)  Income (<6,000 CHF [reference], 6,000-12,000 CHF, >12,000 CHF)  BMI (index)  Number of comorbidities (none vs at least 1)  Respiratory conditions (yes vs no)  Initial symptom severity (asymptomatic [reference], mild to moderate, severe to very severe)  Initial hospitalization (yes vs no)  Initial ICU stay (yes vs no)  Self-reported through REDCap | **Recovery/nonrecovery:** not having fully recovered, among initially symptomatic participants  **Fatigue:** Fatigue Assessment Scale (score ≥22 = presence of relevant fatigue)  **Dyspnea:** Modified Medical Research Council (mMRC) dyspnea scale grade ≥ 1  **Presence and severity of anxiety, depression, stress:** 21-item Depression, Anxiety and Stress Scale (DASS21)  Median 7.2 (range: 5.9-10.3) months after diagnosis | Multivariate logistic regression  Adjusted for, by outcome:  Non-recovery (age, sex, initial symptom severity, initial hospitalization, initial ICU stay, smoking, BMI, comorbidities)  Fatigue (age, sex, initial symptom severity, initial hospitalization, initial ICU stay, smoking, BMI, comorbidities)  Dyspnea (age, sex, initial symptom severity, initial hospitalization, initial ICU stay, smoking, BMI, respiratory conditions, comorbidities)  Presence and severity of anxiety, depression, stress (age, sex, initial symptom severity, initial hospitalization, initial ICU stay, smoking, BMI, comorbidities, education, employment, income) |
| **Peghin 2021**  Ref ID: 3136  Italy  Bidirectional cohort  Non-industry | 599 adult inpatients and outpatients with COVID-19 diagnosis  100% Nucleic acid amplification test  Age, mean (SD): 53.0 (15.8)  47% male  Ethnicity: 91.5% native Italian; 7.8% European  Employment: 42.2% work in contact with public and HCWs; 18.8% retired | Age (18-40 [reference], 41- 60, >60)  Sex  Employment (in contact with public, not in contact with public, retired, other [reference varied])  Number of symptoms of acute Covid-19 (0 symptoms vs ≥1 symptoms)  Acute Covid-19 management (outpatient/ward/ICU)  Data source NR | **Recovery/nonrecovery:** symptoms that developed during or after COVID-19, continued for ≥12 weeks, and were not explained by an alternative diagnosis.  Patient-reported (narratives) outcomes were categorized by four independent physicians (includes among others: dyspnea, fatigue, psychiatric/mood disorder, brain fog, upper respiratory tract involvement).  Mean 187 days (SD: 22) after the disease onset | Multivariate logistic regression  Adjusted for age, sex, number of symptoms of acute Covid-19, management of acute Covid-19 (ward, outpatient, ICU), IgG value at 6 months (for employment only^†^) |
| **Westerlind 2021**  Ref ID: 4338  Sweden  Retrospective cohort  Industry and non-industry | 11,955 people who started to receive sickness benefits for Covid-19  33% by ICD codes (U00-U49)  Age, mean (SD): 48.0 (11.3)  40.4% male  Education: 49.6% secondary school; 25.2% long university education; 14.7% short university education; 10.4% primary school  Income: 1000 SEK median (IQR): 212 (61) in low income; 288 (33) in medium income; 390 (100) in high income  Employment: 95.9% employed; 1.7% unemployed | Age (continuous)  Sex  Employment (employed [reference], self-employed, unemployed)  Income (low [reference], medium, high)  Birth country (Sweden [reference], Nordic, European, Outside Europe)  Need for hospitalization during acute Covid-19 phase (yes vs no)  Swedish Social Insurance Agency, National Patient Register, and Statistics Sweden | **Sick leave for long Covid-19:** sick leave for long Covid-19 (≥ 12 weeks); at least one period of sickness benefits due to Covid-19 diagnosis, comprised of maximum of 122 days (4 months of follow-up) and receiving sickness benefits regardless of amount  Data extracted from Swedish Social Insurance Agency database  Being on sick leave for at least 12 weeks | Multivariate logistic regression for overall sample and by subgroup of participants receiving inpatient care  Adjusted for age, sex, prior sick leave, country of birth, education, employment, income, marital status, inpatient care |

^*^ SES: socio-economic status; BMI: body mass index; CFQ: Chalder Fatigue Scale; ECOG: Eastern Cooperative Oncology Group performance scale; MoCA: Montreal Cognitive Assessment scale; mMRC: modified Medical Research Council (dyspnea scale); N: total number; OR: odds ratio; RAND: RAND-36 questionnaire (energy/fatigue scale); vs: versus; yr: year(s); O2: oxygen

**^ʃ^** All outcomes are dichotomous, unless otherwise stated.

^ǂ^ Male was the reference category in majority of the studies. We reversed the reported estimates if ‘female’ was the reference category in a study (Stavem 2021; Liu 2021; Qin 2021).

^†^ Association between employment and outcome was assessed in a subgroup of patients who had serological assessment performed at follow-up.

**Table S4. Risk of Bias Assessments for Key Question 1**

| **Author, year** | **Were the two groups similar and recruited from the same population?** | **Were the risk factors measured similarly to assign people to both exposed and unexposed groups?** | **Were the risk factors measured in a valid and reliable way?** | **Were confounding factors identified?** | **Were strategies to deal with confounding factors stated?** | **Is there sufficient indication that the outcome (especially the continuous outcomes) was attributable to long-COVID?** | **Were the outcomes measured in a valid and reliable way?** | **Was the follow up time reported and sufficient to be long enough for outcomes to occur?** | **Was follow up complete, and if not, were the reasons to loss to follow up described and explored?** | **Were strategies to address incomplete follow up utilized?** | **Was appropriate statistical analysis used (i.e. age, sex, comorbidities, illness severity)?** | **Was Covid confirmed via lab in ≥ 90% of participants?** | **Are there potential outcomes measured that were not reported?^*^** |
| --- | --- | --- | --- | --- | --- | --- | --- | --- | --- | --- | --- | --- | --- |
| **Blomberg, 2021** | + | + | - | + | + | + | + | + | + | NA | + | + | - |
| **Boscolo-Rizzo, 2021** | + | + | - | + | + | + | + | + | + | NA | - | + | + |
| **Frontera, 2021** | + | + | + | + | + | ? | + | + | - | - | + | + | + |
| **Huang, 2021** | + | + | + | + | + | + | - | + | - | - | + | + | + |
| **Kayaaslan, 2021** | + | + | - | + | + | + | + | + | + | NA | ? | + | - |
| **Liu, 2021** | + | + | + | + | + | + | + | + | + | NA | + | + | - |
| **Menges, 2021** | + | + | - | + | + | + | + | + | + | NA |  | + | - |
| **Munblit, 2021** | + | + | + | + | + | + | ? | + | - | - | + | - | - |
| **Nehme, 2021** | + | + | + | + | + | + | ? | + | - | + | - | + | + |
| **Osmanov, 2021** | + | + | + | + | + | + | + | + | - | - | + | + | - |
| **Peghin, 2021** | + | + | + | + | + | + | + | + | + | NA | - | + | - |
| **Qin, 2021** | + | ? | ? | + | + | ? | ? | + | + | NA | - | + | - |
| **Qu, 2021** | + | ? | ? | + | + | + | + | + | + | NA | - | + | - |
| **Shang, 2021** | + | ? | ? | - | + | ? | - | + | - | - | - | + | - |
| **Sigfrid, 2021** | + | + | + | + | + | + | + | + | - | - | + | + | - |
| **Stavem, 2021** | + | + | + | + | + | + | + | + | + | NA | + | + | - |
| **Westerlind, 2021** | + | + | + | + | + | + | + | ? | + | NA | - | - | - |

+: Yes/low risk of bias; -: No/high risk of bias; ?: Unclear; NA: Not applicable

* For this question a “-/No” response is considered as low risk of bias.

**Table S5. Detailed Summary of Findings for Risk Factors of Post-Covid Condition in Children**

| **Risk Factor** | **Outcome & Timing** | **Baseline severity (No. of Studies)** | **Author, year (RoB)** | **Findings, aOR (95% CI)** | **Conclusions** | **GRADE** |
| --- | --- | --- | --- | --- | --- | --- |
|  | **Non-recovery** | | | | | |
| **Age (continuous)** | ≥22 wks | Hospitalized (n = 1) | Osmanov, 2021  (Some concern) | 1.06 (1.03 to 1.10) | Small-to-moderate association with an increase in post-Covid condition among children ≥ 6 years | Low^a,c^ |
| **Age group (ref: < 2 yrs)** |  |  |  | 2-5 yrs: 0.93 (0.38 to 2.22)  6-11 yrs: 2.57 (1.29 to 5.36)  12-18 yrs: 2.52 (1.34 to 5.01) |  |  |
| **Sex (ref: Male)** |  |  |  | 1.18 (0.77 to 1.81) | Very uncertain | Very low^a,c,d^ |
| **Obesity** |  |  |  | 2.06 (0.86 to 4.89) | Very uncertain | Very low^a,c,d^ |
| **Allergies** |  |  |  | 1.67 (1.04 to 2.67) | Very uncertain | Very low^a,c,d^ |
| **Pneumonia during acute Covid-19** |  |  |  | 1.22 (0.79 to 1.89) | Very uncertain | Very low^a,c,d^ |
| **Severe acute Covid-19** |  |  |  | 2.58 (0.79 to 8.07) | Very uncertain | Very low^a,c,d^ |

**Table S6. Detailed Summary of Findings for Socio-demographic Risk Factors of Post-Covid Condition**

| **Risk Factor** | **Outcome & Timing** | **Baseline severity (No. of Studies)** | **Author, year (RoB)** | **Findings, aOR (95% CI)**  **Including results in subsets with laboratory confirmed Covid-19, if applicable** | **Conclusions** | **GRADE** |
| --- | --- | --- | --- | --- | --- | --- |
| **Age (continuous)** | **Non-recovery** | | | | | |
|  | 12-21 wks | Hospitalized (n = 1) | Qin, 2021  (Some concern) | **1.00 (0.99 to 1.01)** | Little-to-no association | Low^a,b^ |
|  | ≥22 wks | Hospitalized (n = 1) | Munblit, 2021  (Some concern) | **0.99 (0.98 to 0.99)**  Subset: 0.99 (0.98 to 1.00) |  |  |
|  |  |  | **Pooled (n =2):** 0.99 (0.98 to 1.00); I^2^: 48% | |  |  |
| **Age group** | ≥22 wks | Hospitalized (n = 2) | Shang, 2021  (High) | > 65 (ref: ≤ 65 yrs): aOR NR, adjusted p = 0.277  Univariate OR: 0.839 (0.619 to 1.1) | Little-to-no association in 40-60 and >60 yrs vs 18-40 | Low^a,c^ |
|  |  |  | Sigfrid, 2021  (Some concern) | Male\|<50 yrs: ref  Male\|50-69 yrs: 0.82 (0.21 to 3.3)  Male\|≥70 yrs: 0.74 (0.14 to 3.83)  Subset:  Male\|50-69 yrs: 0.80 (0.20 to 3.22)  Male\|≥70 yrs: 0.73 (0.14 to 3.78) |  |  |
|  |  | Mixed (n = 2) | Menges, 2021  (Some concern) | 18-39 yrs: ref  **40-64 yrs: 1.59 (0.93 to 2.73)**  **≥ 65 yrs: 0.97 (0.41 to 2.20)** |  |  |
|  |  |  | Peghin, 2021  (Some concern) | **41-60 vs 18-40 yrs: 1.00 (0.61 to 1.62)**  **> 60 vs 18-40 yrs: 1.03 (0.62 to 1.74)**  > 60 vs 41-60 yrs: 1.04 (0.67 to 1.60) |  |  |
|  |  | Non-hospitalized (n = 2) | Boscolo-Rizzo, 2021 (Some concern) | < 40 yrs: ref  **40-54 yrs: 1.92 (1.07 to 3.44)**  ≥55 yrs: 1.56 (0.84 to 2.90) |  |  |
|  |  |  | Nehme, 2021  (High) | 18-39 yrs: ref  **40-59 yrs: 1.13 (0.74 to 1.72)**  **≥ 60 yrs: 1.46 (0.72 to 2.94)** |  |  |
|  |  |  | **Pooled (n = 4):** (40-60 vs 18-40 yrs): 1.31 (0.99 to 1.74); I^2^: 20%  **Pooled (n = 3):** (>60 vs 18-40yrs): 1.12 (0.77 to 1.63); I^2^: 0%  **Pooled without Nehme (n = 3):** (40-60 vs 18-40 yrs): 1.42 (0.96 to 2.09); I^2^: 36%  **Pooled without Nehme (n = 2):** (>60 vs 18-40yrs): 1.01 (0.64 to 1.61); I^2^: 0% | |  |  |
| **Sex (ref: Male)** | 12-21 wks | Hospitalized (n = 1) | Qin, 2021  (Some concern) | **1.42 (0.90 to 2.00)** | Small-to-moderate association | Moderate^a^ |
|  | ≥22 wks | Hospitalized (n = 3) | Shang, 2021  (High) | aOR NR; adjusted p = 0.002  **Univariate OR: 1.62 (1.19 to 2.18)** |  |  |
|  |  |  | Munblit, 2021  (Some concern) | **1.83 (1.55 to 2.17)**  Subset: 1.88 (1.49 to 2.37) |  |  |
|  |  |  | Sigfrid, 2021  (Some concern) | In <50 yrs: 2.75 (0.26 to 28.92)  Subset:  In <50 yrs: 2.75 (0.26 to 29.00) |  |  |
|  |  | Mixed (n = 2) | Peghin, 2021  (Some concern) | **1.54 (1.05 to 2.27)** |  |  |
|  |  |  | Menges, 2021  (Some concern) | **1.89 (1.18 to 3.07)** |  |  |
|  |  | Non-hospitalized (n = 2) | Boscolo-Rizzo, 2021 (Some concern) | **1.64 (1.00 to 2.70)** |  |  |
|  |  |  | Nehme, 2021  (High) | **1.69 (1.09 to 2.61)** |  |  |
|  |  |  | **Pooled (n = 8):** 1.72 (1.53 to 1.94); I^2^: 0%  **Pooled without Shang, 2021 and Sigfrid, 2021 (n = 6):** 1.74 (1.53 to 1.98); I^2^: 0% | |  |  |
| **Occupation** | ≥22 wks | Mixed (n = 1) | Peghin, 2021  (Some concern) | In contact with public & HCWs vs not in contact with public: 0.84 (0.39 to 1.78)  In contact with public & HCWs vs retired: 1.27 (0.45 to 3.57)  In contact with public & HCWs vs other: 2.43 (0.85 to 7.14)  Not in contact with public vs other: 2.94 (0.93 to 9.09)  Not in contact with public vs retired: 1.52 (0.49 to 4.76)  Other vs retired: 0.52 (0.15 to 1.82) | Very uncertain | Very Low^a,C,d^ |
| **BMI (continuous)** | ≥22 wks | Mixed (n = 1) | Menges, 2021  (Some concern) | 1.04 (0.99 to 1.09) | Very uncertain | Very low^a,b,c^ |
| **Obesity** | ≥22 wks | Hospitalized (n = 1) | Munblit, 2021  (Some concern) | Obesity (as defined by clinical staff): 1.09 (0.89 to 1.34)  Subset: 0.87 (0.65 to 1.17) | Very uncertain | Very low^a,b,C^ |
| **Overweight** | ≥22 wks | Non-hospitalized (n = 1) | Boscolo-Rizzo, 2021 (Some concern) | BMI (≥25 vs <25): 1.67 (1.00 to 2.78) | Very uncertain | Very low^a,C,d^ |
| **Age (continuous)** | **Fatigue** | | | | | |
|  | 12-21 wks | Non-hospitalized (n = 1) | Stavem, 2021  (Some concern) | **1.02 (0.86 to 1.22)** | Little-to-no association | Low^a,b^ |
|  | ≥22 wks | Hospitalized (n = 2) | Munblit, 2021  (High) | **0.99 (0.99 to 1.00)**  Subset: 0.99 (0.98 to 1.00) |  |  |
|  |  |  | Huang, 2021  (Some concern) | **1.17 (1.07 to 1.27)** |  |  |
|  |  | Mixed (n = 1) | Blomberg, 2021  (Some concern) | **1.01 (0.99 to 1.03)** |  |  |
|  |  |  | **Pooled (n = 4):** 1.02 (0.98 to 1.06); I^2^: 81% | |  |  |
| **Age group** | ≥22 wks | Hospitalized (n = 2) | Shang, 2021  (High) | >65 vs ≤65 yrs: aOR NR; adjusted p = 0.525  Univariate OR: 0.89 (0.64 to 1.24) | Very uncertain | Very low ^a,b,c^ |
|  |  |  | Sigfrid, 2021  (Some concern) | Fatigue level (continuous), aMD (95% CI):  Male\|<50 yrs: ref  Male\|50-69 yrs: 0.44 (-0.56 to 1.44)  Male\|≥70 yrs: 0.38 (-0.84 to 1.60)  Subset, aMD (95% CI):  Male\|50-69 yrs: 0.43 (-0.58 to 1.44)  Male\|≥70 yrs: 0.35 (-0.89 to 1.58) |  |  |
|  |  | Mixed (n = 1) | Menges, 2021  (Some concern) | 18-39 yrs: ref  40-64 yrs: 0.59 (0.39 to 0.91)  ≥65 yrs: 0.41 (0.21 to 0.78) |  |  |
|  |  | Non-hospitalized (n = 1) | Nehme, 2021  (High) | 18-39 yrs: ref  40-59 yrs: 1.32 (0.79 to 2.22)  ≥ 60 yrs: 2.38 (1.10 to 5.17) |  |  |
| **Sex (ref: Male)** | 12-21 wks | Non-hospitalized (n = 1) | Stavem, 2021  (Some concern) | **2.04 (1.32 to 3.23)** | Small-to-moderate association | Moderate^a^ |
|  | ≥22 wks | Hospitalized (n = 4) | Munblit, 2021  (High) | **1.67 (1.39 to 2.02)**  Subset: 1.81 (1.40 to 2.34) |  |  |
|  |  |  | Huang, 2021  (Some concern) | **1.33 (1.05 to 1.67)** |  |  |
|  |  |  | Shang, 2021  (High) | aOR NR; adjusted p = 0.009  **Univariate OR: 1.53 (1.11 to 2.12)** |  |  |
|  |  |  | Sigfrid, 2021  (Some concern) | Fatigue level (continuous, range: 0-10), aMD (95% CI):  In <50 yrs: 2.06 (0.81 to 3.31)  Subset, aMD (95% CI):  In <50 yrs: 2.07 (0.82 to 3.33) |  |  |
|  |  | Mixed (n = 2) | Blomberg, 2021  (Some concern) | **2.02 (1.13 to 3.70)** |  |  |
|  |  |  | Menges, 2021  (Some concern) | **1.38 (0.94 to 2.04)** |  |  |
|  |  | Non-hospitalized (n = 1) | Nehme, 2021 (High) | **1.82 (1.05 to 3.15)** |  |  |
|  |  |  | **Pooled (n = 7):** 1.58 (1.41 to 1.77); I^2^: 0%  **Pooled without Shang (n = 6):** 1.58 (1.40 to 1.80); I^2^: 3% | |  |  |
| **BMI (continuous)** | 12-21 wks | Non-hospitalized (n = 1) | Stavem, 2021  (Some concern) | **1.03 (0.99 to 1.08)** | Little-to-no association | Low^a,b^ |
|  | ≥22 wks | Mixed (n = 2) | Blomberg, 2021  (Some concern) | **0.99 (0.91 to 1.07)** |  |  |
|  |  |  | Menges, 2021  (Some concern) | **1.04 (1.00 to 1.09)** |  |  |
|  |  |  | **Pooled (n = 3):** 1.03 (1.00 to 1.06); I^2^: 0% | |  |  |
| **Obesity** | ≥22 wks | Hospitalized (n = 1) | Munblit, 2021  (High) | 1.15 (0.91 to 1.44)  Subset: 1.03 (0.75 to 1.42) | Very uncertain | Very low^A,c^ |
| **Age (continuous)** | **Dyspnea** | | | | | |
|  | ≥22 wks | Hospitalized (n = 1) | Munblit, 2021  (Some concern) | 0.99 (0.98 to 1.00)  Subset: 0.99 (0.98 to 1.00) | Little-to-no association | Low^a,c^ |
| **Age group** | ≥22 wks | Hospitalized (n = 1) | Sigfrid 2021  (Some concern) | Change in dyspnea:  Male\|<50 yrs: ref  Male\|50-69 yrs: 2.2 (0.89 to 5.45)  Male\|≥70 yrs: 2.59 (0.84 to 7.95)  Subset:  Male\|50-69 yrs: 2.35 (0.95 to 5.83)  Male\|≥70 yrs: 2.43 (0.79 to 7.47) | Very uncertain | Very low^a,b,c^ |
|  |  | Mixed (n = 1) | Menges, 2021  (Some concern) | 18-39 yrs: ref  40-64 yrs: 0.79 (0.41 to 1.49)  ≥ 65 yrs: 0.90 (0.37 to 2.16) |  |  |
|  |  | Non-hospitalized (n = 1) | Nehme, 2021  (High) | 18-39 yrs: ref  40-59 yrs: 1.39 (0.73 to 2.65)  ≥ 60 yrs: 1.35 (0.47 to 3.19) |  |  |
| **Sex (ref: Male)** | ≥22 wks | Hospitalized (n = 2) | Munblit, 2021  (Some concern) | **1.31 (1.06 to 1.62)**  Subset: 1.21 (0.90 (1.62) | Small-to-moderate association | Moderate^a^ |
|  |  |  | Sigfrid 2021  (Some concern) | Change in dyspnea:  **In <50 yrs: 7.15 (2.24 to 22.8)**  Subset: In <50 yrs: 7.05 (2.22 to 22.43) |  |  |
|  |  | Mixed (n = 1) | Menges, 2021  (Some concern) | **2.24 (1.31 to 3.87)** |  |  |
|  |  | Non-hospitalized (n = 1) | Nehme, 2021  (High) | **2.00 (0.97 to 4.16)** |  |  |
|  |  |  | **Pooled (n = 4):** 2.12 (1.23 to 3.67); I^2^: 73%  **Pooled without Sigfrid (n = 3):** 1.71 (1.10 to 2.67); I^2^: 55% | |  |  |
| **BMI (continuous)** | ≥22 wks | Mixed (n = 1) | Menges, 2021  (Some concern) | 1.14 (1.08 to 1.20) | Very uncertain | Very low^a,b,c^ |
| **Obesity** | ≥22 wks | Hospitalized (n = 1) | Munblit, 2021  (Some concern) | 1.30 (1.00 to 1.67)  Subset: 1.10 (0.76 to 1.56) | Very uncertain | Very low^a,c,d^ |
| **Age group** | **Quality of Life (overall summary index)** | | | | | |
|  | ≥22 wks | Hospitalized (n = 1) | Sigfrid 2021  (Some concern | aMD (95% CI):  Male\|50-69 yrs: -0.05 (-0.11 to 0.02)  Male\|≥70 yrs: -0.04 (-0.12 to 0.04)  Subset, aMD (95% CI):  Male\|50-69 yrs: -0.04 (-0.11 to 0.02)  Male\|≥70 yrs: -0.04 (-0.12 to 0.04) | Very uncertain | Very low^a,b,c^ |
| **Sex (ref: Male)** | ≥22 wks | Hospitalized (n = 1) | Sigfrid 2021  (Some concern | aMD (95% CI):  In <50 yrs: -0.19 (-0.27 to -0.11)  Subset, aMD (95% CI):  In <50 yrs: -0.19 (-0.27 to -0.11) | Very uncertain | Very low^a,b,c^ |
| **Age (continuous)** | **Depression** | | | | | |
|  | ≥22 wks | Hospitalized (n = 2) | Munblit, 2021  (High) | **0.99 (0.98 to 1.00)**  Subset: 0.99 (0.98 to 1.00) | Little-to-no association | Low^a,b^ |
|  |  |  | Huang, 2021 (Some concern) | **0.96 (0.87 to 1.06)** |  |  |
|  |  |  | **Pooled (n = 2):** 0.99 (0.98 to 1.00); I^2^: 0% | |  |  |
| **Age group** | ≥22 wks | Mixed (n = 1) | Menges, 2021  (Some concern) | 18-39 yrs: ref  40-64 yrs: 0.97 (0.59 to 1.59)  ≥ 65 yrs: 1.18 (0.56 to 2.42) | Little-to-no association in adults > 40 years old | Low^a,c^ |
| **Sex (ref: Male)** | ≥22 wks | Hospitalized (n = 2) | Munblit, 2021  (High) | **1.83 (1.41 to 2.40)**  Subset: 1.73 (1.22 (2.47) | Very uncertain | Very low^a,C^ |
|  |  |  | Huang, 2021  (Some concern) | **1.80 (1.39 to 2.34)** |  |  |
|  |  | Mixed (n = 1) | Menges, 2021  (Some concern) | **0.74 (0.47 to 1.15)** |  |  |
|  |  |  | **Pooled (n = 3):** 1.40 (0.89 to 2.21); I^2^: 84%  **Pooled in hospitalized subgroup, (n = 2):** 1.81 (1.51 to 2.18); I^2^: 0% | |  |  |
| **Employment (ref: Employed)** | ≥22 wks | Mixed (n = 1) | Menges, 2021  (Some concern) | Student: 0.60 (0.09 to 2.41) | Very uncertain | Very low^a,c,d^ |
|  |  |  |  | Retired: 1.89 (0.58 to 6.11) | Very uncertain | Very low^a,c,d^ |
|  |  |  |  | Unemployed/other: 2.53 (1.12 to 5.63) | Very uncertain | Very low^a,b,c^ |
| **Income** | ≥22 wks | Mixed (n = 1) | Menges, 2021  (Some concern) | 6,000-12,000 CHF vs <6,000 CHF: 1.05 (0.61 to 1.81)  >12,000 CHF vs <6,000 CHF: 0.84 (0.45 to 1.55)  *Minimum wage is 3000 CHF | Very uncertain | Very low^a,c,d^ |
| **BMI (continuous)** | ≥22 wks | Mixed (n = 1) | Menges, 2021  (Some concern) | 1.02 (0.97 to 1.06) | Very uncertain | Very low^a,b,c^ |
| **Obesity** | ≥22 wks | Hospitalized (n = 1) | Munblit, 2021  (High) | 0.93 (0.67 to 1.28)  Subset: 0.85 (0.54 to 1.31) | Very uncertain | Very low^a,c,d^ |
| **Age (continuous)** | **Functional incapacity** | | | | | |
|  | ≥22 wks | Hospitalized (n = 1) | Frontera, 2021  (High) | Dependence/disability: 1.02 (1.00 to 1.04)  Abnormal ADL: 1.04 (1.01 to 1.06) | Very uncertain | Very low^A,b,c^ |
| **Age group** | 12-21 wks | Hospitalized (n = 2) | Qu, 2021  (Some concern) | Poor physical component score of SF-36: ≥60 yrs vs <60 yrs: 2.44 (1.33 to 4.47) | Small-to-moderate association with an increase in incapacity in ≥60 yrs old | Low^a,b^ |
|  |  |  | Sigfrid 2021  (Some concern) | New/worsened disability in at least one domain:  Male\|<50 yrs: ref  Male\|50-69 yrs: 1.66 (0.51 to 5.42)  Male\|≥70 yrs: 2.08 (0.55 to 7.96)  Subset:  Male\|50-69 yrs: 1.75 (0.53 to 5.72)  Male\|≥70 yrs: 2.17 (0.57 to 8.30) |  |  |
| **Sex (ref: Male)** | 12-21 wks | Hospitalized (n = 1) | Qu, 2021  (Some concern) | **Poor physical component score of SF-36: 1.79 (1.04 to 3.06)** | Small-to-moderate association | Low^a,d^ |
|  | ≥22 wks | Hospitalized (n = 1) | Sigfrid 2021  (Some concern) | **New/worsened disability in at least one domain:**  **In <50 yrs: 4.22 (1.12 to 15.94)**  Subset:  In <50 yrs: 4.23 (1.12 to 15.97) |  |  |
|  |  |  | **Pooled (n =2):** 2.20 (1.07 to 4.51); I^2^: 27% | |  |  |
| **Age group** | **Sleep disturbance** | | | | | |
|  | ≥22 wks | Hospitalized (n = 1) | Shang, 2021  (High) | >65 vs ≤ 65 yrs: aOR NR; adjusted p = 0.921  Univariate OR: 1.01 (0.72 to 1.41) | Very uncertain | Very low^A,c^ |
|  |  | Non-hospitalized (n = 1) | Nehme, 2021  (High) | 18-39 yrs: ref  40-59 yrs: 2.95 (1.06 to 8.24)  ≥ 60 yrs: 2.80 (0.64 to 12.25) |  |  |
| **Sex (ref: Male)** | ≥22 wks | Hospitalized (n = 1) | Shang, 2021  (High) | aOR NR; adjusted p = 0.008  Univariate OR: 1.57 (1.12 to 2.18) | Very uncertain | Very low^A,c^ |
|  |  | Non-hospitalized (n = 1) | Nehme, 2021  (High) | 0.75 (0.32 to 1.78) |  |  |
| **Age group** | **Chest pain** | | | | | |
|  | ≥22 wks | Non-hospitalized (n = 1) | Nehme, 2021  (High) | 18-39 yrs: ref  40-59 yrs: 1.05 (0.31 to 3.50)  ≥ 60 yrs: 1.82 (0.34 to 9.73) | Very uncertain | Very low ^A,c^ |
| **Sex (ref: Male)** | ≥22 wks | Non-hospitalized (n = 1) | Nehme, 2021  (High) | 1.11 (0.34 to 3.67) | Very uncertain | Very low ^A,c^ |
| **Age (continuous)** | **Cognitive impairment** | | | | | |
|  | ≥22 wks | Hospitalized (n = 2) | Frontera, 2021  (High) | 1.03 (1.01 to 1.05) | Very uncertain | Very low^A,b,c^ |
|  |  |  | Liu, 2021  (High) | aOR NR  Univariate OR: 1.02 (1.01 to 1.04) |  |  |
| **Age group** | ≥22 wks | Hospitalized (n = 1) | Shang, 2021  (High) | >65 vs ≤ 65 yrs: aOR NR; adjusted p = 0.148  Univariate OR: 0.68 (0.40 to 1.16) | Very uncertain | Very low^A,b,c^ |
|  |  | Non-hospitalized (n = 1) | Nehme, 2021  (High) | Outcome: Difficulty concentrating (yes/no)  18-39 yrs: ref  40-59 yrs: 1.57 (0.64 to 3.84)  ≥ 60 yrs: 1.11 (0.23 to 5.47)  Outcome: Memory loss (yes/no)  18-39 yrs: ref  40-59 yrs: 0.95 (0.41 to 2.21)  ≥ 60 yrs: 0.18 (0.01 to 3.10) |  |  |
| **Sex (ref: Male)** | ≥22 wks | Hospitalized (n = 2) | Liu, 2021  (High) | aOR NR  Univariate OR: 1.07 (0.86 to 1.32) | Little-to-no association | Low^A^ |
|  |  |  | Shang, 2021  (High) | aOR NR; adjusted p=0.415  Univariate OR: 0. 1.21 (0.74 to 1.99) |  |  |
|  |  |  | **Pooled (n = 2):** 1.09 (0.89 to 1.33); I^2^: 0% | |  |  |
|  |  | Non-hospitalized (n = 1) | Nehme, 2021  (High) | Outcome: Difficulty concentrating (yes/no)  0.67 (0.29 to 1.55)  Outcome: Memory loss (yes/no)  2.43 (0.81 to 7.29) | Very uncertain | Very low ^A,b,c^ |
| **Race/Ethnicity** | ≥22 wks | Hospitalized (n = 1) | Frontera, 2021  (High) | White vs non-White: 0.41 (0.22 to 0.78) | Very uncertain | Very low^A,c^ |
| **Age (continuous)** | **Return to work** | | | | | |
|  | ≥22 wks | Mixed (n = 1) | Westerlind, 2021  (High) | 1.02(1.01 to 1.02) | Very uncertain | Very low^A,b,c^ |
| **Sex (ref: Male)** | ≥22 wks | Mixed (n = 1) | Westerlind, 2021 (High) | 0.94 (0.83 to 1.07) | Very uncertain | Veryl ow^A,c^ |
| **Race/Ethnicity** | ≥22 wks | Hospitalized (n = 1) | Frontera, 2021  (High) | White vs non-White: 5.03 (1.62 to 5.59) | Small-to-moderate association | Low^a,c^ |
| **Immigrant status** | ≥22 wks | Mixed (n = 1) | Westerlind, 2021 (High) | Born in Sweden: ref  Nordic countries: 0.77 (0.53 to 1.12)  European countries: 0.91 (0.76 to 1.10)  Outside Europe: 0.78 (0.67 to 0.90) | Very uncertain | Very low^A,c,d^ |
| **Employment (ref: Employed)** | ≥22 wks | Mixed (n = 1) | Westerlind, 2021 (High) | Self-employed: 1.44 (1.06 to 1.97) | Very uncertain | Very low^A,c,d^ |
|  |  |  |  | Unemployed/other: 1.68 (1.19 to 2.36) | Very uncertain | Very low^A,c^ |
| **Income (ref: Low income)** | ≥22 wks | Mixed (n = 1) | Westerlind, 2021 (High) | Medium: 0.94 (0.81 to 1.08)  High: 0.93 (0.81 to 1.08) | Very uncertain | Very low^A,c^ |

CI: confidence interval; Ref: reference; vs: versus; wks: weeks; NR: not reported; HCW: healthcare worker; aMD: adjusted mean difference; ADL: activities of daily living

**Table S7. Detailed Summary of Findings for Risk Factors Related to Pre-existing Conditions for Post Covid-19 Condition**

| **Risk Factor** | **Outcome & Timing** | **Baseline severity; No Studies** | **Author, year (RoB)** | **Findings, aOR (95% CI)** | **Conclusions** | **GRADE** |
| --- | --- | --- | --- | --- | --- | --- |
| **Number of comorbidities (≥1 vs 0)** | **Non-recovery** | | | | | |
|  | 12-21 wks | Mixed (n=1) | Kayaaslan, 2021  (High) | **1.69 (1.18 to 2.43)** | Small-to-moderate association | Low^a,d^ |
|  | ≥22 wks | Hospitalized (n=1) | Sigfrid 2021  (Some concerns) | **2.28 (0.92 to 5.65)**  Subset: 2.28 (0.92 to 5.65) |  |  |
|  |  | Mixed (n=1) | Menges, 2021  (Some concerns) | **2.08 (1.24 to 3.50)** |  |  |
|  |  | Non-hospitalized (n=1) | Boscolo-Rizzo 2021 (Some concerns) | **1.42 (0.83 to 2.43)** |  |  |
|  |  |  | **Pooled (n=4):** 1.75 (1.36 to 2.24); I^2^=0% | |  |  |
| **Diabetes (any)** | ≥22 wks | Hospitalized (n=1) | Munblit, 2021  (Some concerns) | 0.99 (0.78 to 1.26)  Subset: 1.06 (0.76 to 1.46) | Little-to-no association | Low^a,c^ |
| **Asthma** | ≥22 wks | Hospitalized (n=1) | Munblit, 2021 (Some concerns) | 1.13 (0.77 to 1.65)  Subset: 0.93 (0.56 to 1.51) | Very uncertain | Very low^a,c,d^ |
| **Chronic pulmonary disease** | ≥22 wks | Hospitalized (n=1) | Munblit, 2021 (Some concerns) | 1.47 (1.08 to 1.99)  Subset: 1.32 (0.88 to 1.99) | Very uncertain | Very low^a,c,d^ |
| **Chronic cardiac disease** | ≥22 wks | Hospitalized (n=1) | Munblit, 2021 (Some concerns) | 1.07 (0.84 to 1.37)  Subset: 1.11 (0.80 to 1.53) | Little-to-no association | Low^a,c^ |
| **Hypertension** | ≥22 wks | Hospitalized (n=1) | Munblit, 2021 (Some concerns) | 1.10 (0.91 to 1.33)  Subset: 1.03 (0.79 to 1.35) | Little-to-no association | Low^a,c^ |
| **Rheumatological disorder** | ≥22 wks | Hospitalized (n=1) | Munblit, 2021 (Some concerns) | 1.49 (0.97 to 2.27)  Subset: 1.22 (0.64 to 2.29) | Very uncertain | Very low^a,c,d^ |
| **Active cancer** | ≥22 wks | Hospitalized (n=1) | Munblit, 2021 (Some concerns) | 1.03 (0.66 to 1.60)  Subset: 0.73 (0.41 to 1.28) | Very uncertain | Very low^a,c,D^ |
| **Number of comorbidities (≥1 vs none)** | **Fatigue** | | | | | |
|  | 12-21 wks | Non-hospitalized (n=1) | Stavem, 2021  (Some concerns) | **1.62 (0.94 to 2.77)** | Very uncertain | Very low^a,c,d^ |
|  | ≥22 wks | Hospitalized (n=1) | Huang, 2021  (Some concerns) | **1.08 (0.85 to 1.37)** |  |  |
|  |  | Mixed (n=1) | Menges, 2021 (Some concerns) | **1.27 (0.81 to 2.01)** |  |  |
|  |  |  | **Pooled (n=3):** 1.17 (0.96 to 1.43); I^2^=0% | |  |  |
|  |  | Hospitalized (n=1) | Sigfrid 2021  (Some concerns) | aMD 0.95 (0.35 to 1.55)  Subset: aMD 0.93 (0.32 to 1.54)  (continuous; range: 0-10) | Very uncertain | Very low^a,c, D^ |
| **Number of comorbidities (≥2 vs none)** | 12-21 wks | Non-hospitalized (n=1) | Stavem, 2021 (Some concerns) | 1.52 (0.77 to 3.03) | Very uncertain | Very low^a,c,D^ |
| **Diabetes** | ≥22 wks | Hospitalized (n=1) | Munblit, 2021 (High) | 1.18 (0.91 to 1.52)  Subset: 1.06 (0.74 to 1.50) | Very uncertain | Very low^A,c,d^ |
|  |  | Mixed (n=1) | Blomberg, 2021 (Some concerns) | aRR 1.06 (0.91 to 1.23)  (continuous) | Very uncertain | Very low^a,b,c^ |
| **Asthma** | ≥22 wks | Hospitalized (n=1) | Munblit, 2021 (High) | 1.41 (0.94 to 2.08)  Subset: 1.37 (0.81 to 2.26) | Very uncertain | Very low^A,c,d^ |
| **Chronic pulmonary disease** | ≥22 wks | Hospitalized (n=1) | Munblit, 2021 (High) | **1.68 (1.21 to 2.32)**  Subset: 1.66 (1.08 to 2.54) | Small-to-moderate increase | Low^a,b^ |
|  |  | Mixed (n=1) | Blomberg, 2021 (Some concerns) | Asthma/COPD: **2.17 (0.97 to 4.96)** |  |  |
|  |  |  | **Pooled (n=2):** 1.74 (1.29 to 2.36); I^2^=0% | |  |  |
| **Chronic cardiac disease** | ≥22 wks | Hospitalized (n=1) | Munblit, 2021 (High) | **1.07 (0.84 to 1.37)**  Subset: 1.11 (0.80 to 1.53) | Very uncertain | Very low^a,c,d^ |
|  |  | Mixed (n=1) | Blomberg, 2021 (Some concerns) | **2.23 (0.72 to 7.25)** |  |  |
|  |  |  | **Pooled (n=2):** 1.24 (0.70 to 2.23); I^2^=35% | |  |  |
| **Hypertension** | ≥22 wks | Hospitalized (n=1) | Munblit, 2021 (High) | 1.27 (1.02 to 1.57)  Subset: 1.27 (0.94 to 1.72) | Little-to-no association | Low^a,b^ |
|  |  | Mixed (n=1) | Blomberg, 2021 (Some concerns) | aRR 1.01 (0.90 to 1.13)  (continuous) |  |  |
| **Rheumatological disorder** | ≥22 wks | Hospitalized (n=1) | Munblit, 2021 (High) | 1.21 (0.75 to 1.90)  Subset: 1.04 (0.50 to 2.04) | Little-to-no association | Low^a,b^ |
|  |  | Mixed (n=1) | Blomberg, 2021 (Some concerns) | Rheumatic disease: aRR 1.05 (0.92 to 1.18) (continuous) |  |  |
| **Previous depression** | ≥22 wks | Hospitalized (n=1) | Munblit, 2021 (High) | 1.10 (0.43 to 2.82) | Very uncertain | Very low^A,c,D^ |
| **Active cancer** | ≥22 wks | Hospitalized (n=1) | Munblit, 2021 (High) | 0.99 (0.60 to 1.58)  Subset: 0.62 (0.31 to 1.15) | Very uncertain | Very low^A,c,D^ |
| **Disability** | ≥22 wks | Hospitalized (n=1) | Frontera, 2021 (High) | 1.48 (1.16 to 1.89) | Very uncertain | Very low^A,c,d^ |
| **Number of comorbidities (≥1 vs 0)** | **Dyspnea** | | | | | |
|  | ≥22 wks | Hospitalized (n=1) | Sigfrid 2021  (Some concerns) | 0.74 (0.42 to 1.31)  Subset: 0.76 (0.43 to 1.34)  (change in dyspnea) | Very uncertain | Very low^a,c,d^ |
|  |  | Mixed (n=1) | Menges, 2021 (Some concerns) | 2.71 (1.38 to 5.36) | Very uncertain | Very low^a,C^ |
| **Diabetes** | ≥22 wks | Mixed (n=1) | Munblit, 2021 (Some concerns) | 0.79 (0.57 to 1.08)  Subset: 1.00 (0.65 to 1.50) | Very uncertain | Very low^a,c,d^ |
| **Asthma** | ≥22 wks | Hospitalized (n=1) | Munblit, 2021 (Some concerns) | 1.33 (0.84 to 2.06)  Subset: 1.16 (0.63 to 2.04) | Very uncertain | Very low^a,c,d^ |
| **Chronic pulmonary disease** | ≥22 wks | Hospitalized (n=1) | Munblit, 2021 (Some concerns) | **1.21 (0.82 to 1.76)**  Subset: 1.25 (0.74 to 2.03) | Little-to-no association | Low^a,d^ |
|  |  | Mixed (n=1) | Menges, 2021 (Some concerns) | Respiratory condition: **1.71 (0.70 to 4.01)** |  |  |
|  |  |  | **Pooled (n=2):** 1.28 (0.90 to 1.83); I^2^=0% | |  |  |
| **Chronic cardiac disease** | ≥22 wks | Hospitalized (n=1) | Munblit, 2021 (Some concerns) | 1.20 (0.88 to 1.63)  Subset: 1.20 (0.79 to 1.78) | Very uncertain | Very low^a,c,d^ |
| **Hypertension** | ≥22 wks | Hospitalized (n=1) | Munblit, 2021 (Some concerns) | 1.16 (0.91 to 1.49)  Subset: 1.18 (0.84 to 1.65) | Little-to-no association | Low^a,c^ |
| **Rheumatologic disorder** | ≥22 wks | Hospitalized (n=1) | Munblit, 2021 (Some concerns) | 1.38 (0.81 to 2.26)  Subset: 1.04 (0.43 to 2.22) | Very uncertain | Very low^a,c,d^ |
| **Active cancer** | ≥22 wks | Hospitalized (n=1) | Munblit, 2021 (Some concerns) | 1.08 (0.61 to 1.83)  Subset: 0.74 (0.33 to 1.48) | Very uncertain | Very low^a,c,D^ |
| **Number of comorbidities (≥1 vs none)** | **Quality of Life** | | | | | |
|  | ≥22 wks | Hospitalized (n=1) | Sigfrid 2021  (Some concerns) | aMD -0.02 (-0.06 to 0.02)  Subset: aMD -0.02 (-0.06 to 0.02)  (change in EQ5D-5L) | Very uncertain | Very low^a,b,c^ |
| **Number of comorbidities (≥1 vs none)** | **Depression (depression/anxiety)** | | | | | |
|  | ≥22 wks | Hospitalized (n=1) | Huang, 2021  (Some concerns) | **0.97 (0.74 to 1.27)** | Little-to-no association | Low^a,d^ |
|  |  | Mixed (n=1) | Menges, 2021 (Some concerns) | **1.41 (0.84 to 2.34)** |  |  |
|  |  |  | **Pooled (n=2):** 1.09 (0.78 to 1.54); I^2^=36% | |  |  |
| **Diabetes** | ≥22 wks | Hospitalized (n=1) | Munblit, 2021 (High) | 0.92 (0.63 to 1.33)  Subset: 1.18 (0.72 to 1.89) | Very uncertain | Very low^A,c,d^ |
| **Asthma** | ≥22 wks | Hospitalized (n=1) | Munblit, 2021 (High) | 2.02 (1.24 to 3.18)  Subset: 2.39 (1.31 to 4.2) | Very uncertain | Very low^A,c^ |
| **Chronic pulmonary disease** | ≥22 wks | Hospitalized (n=1) | Munblit, 2021 (High) | 1.19 (0.74 to 1.85)  Subset: 0.79 (0.39 to 1.45) | Very uncertain | Very low^A,c,d^ |
| **Chronic cardiac disease** | ≥22 wks | Hospitalized (n=1) | Munblit, 2021 (High) | 0.95 (0.64 to 1.37)  Subset: 1.11 (0.68 to 1.78) | Very uncertain | Very low^A,c,d^ |
| **Hypertension** | ≥22 wks | Hospitalized (n=1) | Munblit, 2021 (High) | 1.11 (0.82 to 1.51)  Subset: 0.98 (0.65 to 1.47) | Very uncertain | Very low^A,c,d^ |
| **Rheumatologic disorder** | ≥22 wks | Hospitalized (n=1) | Munblit, 2021 (High) | 1.97 (1.12 to 3.33)  Subset: 1.62 (0.69 to 3.45) | Small-to-moderate association | Low ^a,c^ |
| **Active cancer** | ≥22 wks | Hospitalized (n=1) | Munblit, 2021 (High) | 0.61 (0.25 to 1.27)  Subset: 0.48 (0.14 to 1.22) | Very uncertain | Very low^A,c,d^ |
| **Disability** | ≥22 wks | Hospitalized (n=1) | Frontera, 2021 (High) | Depression (worse than average): 1.27 (1.03 to 1.57)  Anxiety (worse than average): 1.25 (1.02 to 1.54) | Very uncertain | Very low^A,c^ |
| **Number of comorbidities (≥1 vs none)** | **Functional capacity** | | | | | |
|  | ≥22 wks | Hospitalized (n=1) | Sigfrid 2021  (Some concerns) | New or worse disability:  2.96 (1.57 to 5.57)  Subset: 3.03 (1.61 to 5.71) | Very uncertain | Very low^a,b,c^ |
| **History of atrial fibrillation** | ≥22 wks | Hospitalized (n=1) | Frontera, 2021 (High) | 4.35 (1.22 to 16.67) | Very uncertain | Very low^A,c^ |
| **Disability** | ≥22 wks | Hospitalized (n=1) | Frontera, 2021 (High) | 1.98 (1.61 to 2.42) | Very uncertain | Very low^A,c^ |
| **COPD** | **Cognitive impairment** | | | | | |
|  | ≥22 wks | Hospitalization (n=1) | Liu, 2021  (Some concerns) | 2.01 (1.40 to 2.88) | Small-to-moderate association | Low^a,c^ |
| **Hypertension** | ≥22 wks | Hospitalization (n=1) | Liu, 2021  (Some concerns) | 1.66 (1.32 to 2.09) | Small-to-moderate association | Low^a,c^ |
| **Dementia** | ≥22 wks | Hospitalization (n=2) | Frontera, 2021 (High) | 4.48 (1.16 to 17.37) | Very uncertain | Very low^A,c^ |
| **Disability** | **Sleep disturbance** | | | | | |
|  | ≥22 wks | Hospitalized (n=1) | Frontera, 2021 (High) | 1.37 (1.09 to 1.72) | Very uncertain | Very low^A,c,d^ |

Certainty (GRADE) was rated down by 0, 1, or 2 levels for risk of bias (a), indirectness in outcome (b), lack of consistency (c), and imprecision/ wide confidence intervals (d); capital letters indicates very serious concern.

**Table S8. Detailed Summary of Findings for Risk Factors Related to Covid-19 Illness Severity**

| **Risk Factor** | **Outcome & Timing** | **Baseline severity (No. of Studies)** | **Author, year (RoB)** | **Findings; OR (95% CI)** | **Conclusions** | **GRADE** |
| --- | --- | --- | --- | --- | --- | --- |
| **Acute Covid-19 severity (critical/ICU vs not)** | **Non-recovery** | | | | | |
|  | ≥ 22 wks | Hospitalized (n = 2) | Sigfrid, 2021  (Some concern) | **Scale 6 or 7 (required IMV or critical**  **care) vs scale 3 (no supplementary O2 or ICU): 1.18 (0.24 to 5.95)**  Subset:  Scale 6 vs scale 3: 1.19 (0.24 to 5.98) | Very uncertain | Very low^a,c,d^ |
|  |  |  | Shang, 2021  (High) | aOR NR; adjusted p = 0.937  **(Univariate OR: 0.94 [0.47 to 1.89])** |  |  |
|  |  | Hospitalized subpopulation in mixed study | Peghin, 2021  (Some concern) | **1.65 (0.61 to 4.64)** |  |  |
|  |  |  | **Pooled (n = 3):** 1.14 (0.66 to 1.94); I^2^: 0%  **Pooled without Shang, 2021:** 1.50 (0.65 to 3.49); I^2^: 0% | |  |  |
|  |  | Mixed (n = 2) | Peghin, 2021  (Some concern) | Admitted to ICU vs outpatient: 3.10 (1.18 to 8.11) | Very uncertain | Very low^a,c,d^ |
|  |  |  | Menges, 2021  (Some concern) | 0.55 (0.10 to 2.49) |  |  |
| **Need for intubation** | ≥ 22 wks | Hospitalized (n = 1) | Shang, 2021  (High) | aOR NR; adjusted p = 0.149  (Univariate OR: 0.43 [0.12 to 1.51]) | Very uncertain | Very low^A,c,e^ |
| **Acute Covid-19 severity (severe/critical vs not)** | 12-21 wks | Mixed (n = 1) | Kayaaslan, 2021 (High) | **3.07 (1.47 to 6.43)** | Small-to-moderate | Moderate^a^ |
|  | ≥ 22 wks | Mixed (n = 1) | Menges, 2021  (Some concern) | **2.05 (1.27 to 3.34)** |  |  |
|  |  |  | **Pooled (n = 2):** 2.31 (1.55 to 3.45): I^2^: 0% | |  |  |
|  |  | Hospitalized (n = 3) | Shang, 2021  (High) | aOR NR; adjusted p = 0.473  (Univariate OR: 0.81 [0.47 to 1.37]) | Little-to-no association | Low^a,c^ |
|  |  |  | Munblit, 2021  (Some concern) | Severe (required NIV, IMV, and/or admission to ICU) vs mild (no supplementary O2 or ICU): 0.84 (0.49 to 1.41)  Subset:  Severe vs mild: 1.07 (0.53 to 2.10) |  |  |
|  |  |  | Sigfrid, 2021  (Some concern) | Scale 5 (required HFNC or NIV) vs scale 3 (no supplementary O2 or ICU): 0.32  (0.07 to 1.46)  Scale 6 or 7 (required IMV or critical  care) vs scale 3 (no supplementary O2 or ICU): 1.18 (0.24 to 5.95)  Subset:  Scale 5 vs scale 3: 0.33 (0.07 to 1.49)  Scale 6 vs scale 3: 1.19 (0.24 to 0.83) |  |  |
| **No. of symptoms (≥1 symptoms vs 0)** | ≥ 22 wks | Mixed (n = 1) | Peghin, 2021  (Some concern) | 1.81 (1.59 to 2.05) | Small-to-moderate association | Low^a,c^ |
| **No. of symptoms (ref: ≤2 symptoms)** | ≥ 22 wks | Non-hospitalized (n = 1) | Boscolo-Rizzo, 2021  (Some concern) | 3 to 7 symptoms: 3.22 (1.01 to 10.24)  ≥8 symptoms: 8.71 (2.73 to 27.76) | Small-to-moderate association | Low^a,c^ |
| **Need for hospitalization** | 12-21 wks | Mixed (n = 1) | Kayaaslan, 2021  (High) | 4.69 (3.23 to 6.83) | Large increase | Low^a,c^ |
|  | ≥ 22 wks | Mixed (n = 2) | Peghin, 2021  (Some concern) | **1.87 (1.19 to 2.94)** | Small-to-moderate association | Low^a,d^ |
|  |  |  | Menges, 2021  (Some concern) | **1.17 (0.63 to 2.16)** |  |  |
|  |  |  | **Pooled (n = 2):** 1.55 (0.99 to 2.44); I^2^: 30% | |  |  |
| **Acute Covid-19 severity (critical/ICU vs not)** | **Fatigue** | | | | | |
|  | ≥ 22 wks | Hospitalized (n = 2) | Sigfrid, 2021  (Some concern) | Fatigue (continuous, 0-10), aMD (95% CI):  Scale 6 or 7 (required IMV or critical  care) vs scale 3 (no supplementary O2 or ICU): -0.18 (-1.90 to 0.74)  **Estimated univariate OR: 0.79 (0.44 to 1.39)**  Subset, aMD (95% CI):  Scale 6 vs scale 3: -0.17 (-1.10 to 0.76)  Estimated univariate OR: 0.38 (0.21 to 0.67) | Very uncertain | Very low ^A,d^ |
|  |  |  | Shang, 2021  (High) | aOR NR; adjusted p = 0.884  (Univariate OR: 0.91 [0.43 to 1.97]) |  |  |
|  |  |  | **Pooled (n =2):** 0.83 (0.53 to 1.32): I^2^: 0% | |  |  |
|  |  | Mixed (n = 1) | Menges, 2021  (Some concern) | 4.63 (1.02 to 32.88) | Very uncertain | Very low^a,c,d^ |
| **Need for intubation** | ≥ 22 wks | Hospitalized (n = 2) | Shang, 2021  (High) | aOR NR; adjusted p = 0.174  (Univariate OR: 0.39 [0.09 to 1.71]) | Very uncertain | Very low^A,c,d^ |
|  |  |  | Frontera, 2021  (High) | 3.83 (1.74 to 8.43) |  |  |
| **Acute Covid-19 severity (severe/critical vs not)** | ≥ 22 wks | Hospitalized (n = 4) | Shang, 2021  (High) | **aOR NR; adjusted p = 0.955**  **(Univariate OR: 0.96 [0.55 to 1.68])** | Very uncertain | Very low^a,c^ |
|  |  |  | Huang, 2021  (Some concern) | **Scale 5 (required HFNC/NIV, or both) to 6 (required ECMO/IMV, or both) vs scale 3 (no supplementary**  **O2): 2.69 (1.46 to 4.96)** |  |  |
|  |  |  | Munblit, 2021  (High) | **Severe (required NIV, IMV, and/or admission to ICU) vs mild (no supplementary O2 or ICU): 1.58 (0.91 to 2.69)**  Subset:  Severe vs mild: 2.03 (0.99 to 4.08) |  |  |
|  |  |  | Sigfrid, 2021  (Some concern) | Fatigue (continuous), aMD (95% CI):  Scale 5 (required HFNC or NIV) vs scale 3 (no supplementary O2 or ICU): -0.20 (-1.22 to 0.83)  Estimated univariate OR: 0.79 (0.40 to 1.52)  Scale 6 or 7 (required IMV or critical  care) vs scale 3 (no supplementary O2 or ICU): -0.18 (-1.90 to 0.74)  Estimated univariate OR: 0.79 (0.44 to 1.39)  Subset, aMD (95% CI):  Scale 5 vs scale 3: -0.17 (-1.21 to 0.86)  Estimated univariate OR, scale 5 vs scale 3: 0.61 (0.31 to 0.83)  Scale 6 vs scale 3: -0.17 (-1.10 to 0.76)  Estimated univariate OR, scale 6 vs scale 3: 0.38 (0.21 to 0.67) |  |  |
|  |  |  | **Pooled (n =3):** 1.58 (0.89 to 2.8); I^2^: 67%  **Pooled without the Shang, 2021:** 2.03 (1.2 to 3.41); I^2^: 38% | |  |  |
|  |  | Mixed (n = 2) | Blomberg, 2021 (Some concern) | **1.99 (1.37 to 2.96)** | Very uncertain | Very low^a,B^ |
|  |  |  | Menges, 2021  (Some concern) | **Severe to very severe vs asymptomatic: 1.38 (0.69 to 2.73)** |  |  |
|  |  |  | **Pooled in mixed (n = 2):** 1.83 (1.32 to 2.55); I^2^: 0% | |  |  |
| **No. of symptoms (ref: 0 to 5 symptoms)** | 12-21 wks | Non-hospitalized (n = 1) | Stavem, 2021  (Some concern) | 6 to 9 symptoms: 1.44 (0.79 to 2.64)  10 to 23 symptoms: 3.66 (1.88 to 7.11) | Small-to-moderate association | Low^a,c^ |
| **Need for hospitalization (ref: No)** | ≥ 22 wks | Mixed (n = 1) | Menges, 2021  (Some concern) | 1.00 (0.59 to 1.71) | Very uncertain | Very low^a,c,D^ |
| **Acute Covid-19 severity (critical/ICU vs not)** | **Dyspnea** | | | | | |
|  | ≥ 22 wks | Hospitalized (n = 1) | Sigfrid, 2021  (Some concern) | Scale 6 or 7 (required IMV or critical care) vs scale 3 (no supplementary O2 or ICU): 1.82 (0.79 to 4.22)  Subset:  Scale 6 vs scale 3: 1.99 (0.85 to 4.64) | Very uncertain | Very low^a,b,c,d^ |
|  |  | Mixed (n = 1) | Menges, 2021  (Some concern) | 1.05 (0.22 to 5.31) | Very uncertain | Very Low^a,c,D^ |
| **Acute Covid-19 severity (severe/critical vs not)** | ≥ 22 wks | Hospitalized (n = 2) | Munblit, 2021  (Some concern) | Severe (required NIV, IMV, and/or admission to ICU) vs mild (no supplementary O2 or ICU): 1.08 (0.54 to 2.01)  Subset:  Severe: 1.09 (0.43 to 2.43) | Little-to-no association | Moderate^a^ |
|  |  |  | Sigfrid, 2021  (Some concern) | Scale 5 (required HFNC or NIV) vs scale 3 (no supplementary O2 or ICU): 0.89 (0.36 to 2.21)  Scale 6 or 7 (required IMV or critical care) vs scale 3 (no supplementary O2 or ICU): 1.82 (0.79 to 4.22)  Subset:  Scale 5 vs scale 3: 0.90 (0.36 to 2.23)  Scale 6 vs scale 3: 1.99 (0.85 to 4.64) |  |  |
|  |  | Mixed (n = 1) | Menges, 2021  (Some concern) | Severe to very severe vs asymptomatic: 1.42 (0.57 to 3.87) | Very uncertain | Very low^a,b,c,D^ |
| **Need for hospitalization (ref: No)** | ≥ 22 wks | Mixed (n = 1) | Menges, 2021  (Some concern) | 4.17 (2.23 to 7.91) | Large increase | Low^a,c^ |
| **Acute Covid-19 severity** | **Quality of Life (overall summary index)** | | | | | |
|  | ≥ 22 wks | Hospitalized (n = 1) | Sigfrid, 2021 (Some concern) | aMD (95% CI):  Scale 3 (no supplementary O2 or ICU): ref  Scale 5 (required HFNC or NIV): 0.01 (-0.06 to 0.08)  Estimated univariate OR, scale 5: 1.00 (0.51 to 1.96)  Scale 6 or 7 (required IMV or critical care): -0.05 (-0.11 to 0.02)  Estimated univariate OR, scale 6 or 7: 1.00 (0.56 to 1.79)  Subset, aMD (95% CI):  Scale 3: ref  Scale 5: 0.01 (-0.06 to 0.08)  Estimated univariate OR, scale 5 vs scale 3: 1.00 (0.51 to 1.96)  Scale 6: -0.04 (-0.10 to 0.02)  Estimated univariate OR, scale 6: 1.00 (0.56 to 1.79) | Very uncertain | Very low^a,c,d^ |
| **Acute Covid-19 severity (critical/ICU vs no)** | **Depression** | | | | | |
|  | ≥ 22 wks | Mixed (n = 1) | Menges, 2021 (Some concern) | 0.47 (0.07 to 2.17) | Very uncertain | Very low^a,c,d^ |
| **Acute Covid-19 severity (severe/critical vs not)** | ≥ 22 wks | Hospitalized (n = 2) | Huang, 2021 (Some concern) | **Scale 5 (required HFNC/NIV, or both) to 6 (required ECMO/IMV, or both) vs scale 3 (no supplementary O2): 1.77 (1.05 to 2.97)** | Small-to-moderate | Low^a,d^ |
|  |  |  | Munblit, 2021 (High) | **Severe (required NIV, IMV, and/or admission to ICU) vs mild (no supplementary O2 or ICU): 1.26 (0.54 to 2.59)**  Subset:  Severe vs mild: 0.93 (0.27 to 2.49) |  |  |
|  |  |  | **Pooled in hospitalized (n = 2):** 1.61 (1.03 to 2.51); I^2^: 0% | |  |  |
|  |  | Mixed (n = 1) | Menges, 2021 (Some concern) | Severe to very severe vs asymptomatic: 2.05 (0.96 to 4.69) | Very uncertain | Very low^a,b,c,d^ |
| **Need for hospitalization (ref: No)** | ≥ 22 wks | Mixed (n = 1) | Menges, 2021 (Some concern) | 1.05 (0.57 to 1.89) | Very uncertain | Very low^a,c,D^ |
| **Acute Covid-19 severity** | **Functional incapacity** | | | | | |
|  | ≥ 22 wks | Hospitalized (n = 1) | Sigfrid, 2021 (Some concern) | Scale 3 (no supplementary O2 or ICU): ref  Scale 5 (required HFNC or NIV): 1.32 (0.49 to 3.51)  Scale 6 or 7 (required IMV or critical care): 1.48 (0.63 to 3.52)  Subset:  Scale 3: ref  Scale 5: 1.26 (0.47 to 3.38)  Scale 6: 1.49 (0.63 to 3.54) | Very uncertain | Very low^a,b,c,d^ |
| **Need for intubation (ref: No)** | ≥ 22 wks | Hospitalized (n = 1) | Frontera, 2021 (High) | 3.57 (1.67 to 7.69) | Small-to-moderate association | Low^a,c^ |
| **Acute Covid-19 severity (critical/ICU vs no)** | **Cognitive impairment** | | | | | |
|  | ≥ 22 wks | Hospitalized (n = 1) | Shang, 2021 (High) | aOR NR; adjusted p = 0.646  (Univariate OR: 2.07 [0.83 to 5.14]) | Very uncertain | Very low^a,c,d^ |
| **Acute Covid-19 severity (severe/critical vs no)** | ≥ 22 wks | Hospitalized (n = 2) | Liu, 2021 (Low) | **Severe vs non-severe: 2.83 (2.06 to 3.89)** | Large | Moderate^a^ |
|  |  |  | Shang, 2021 (High) | **aOR NR; adjusted p = 0.045**  **(Univariate OR: 2.03 [1.01 to 4.07])** |  |  |
|  |  |  | **Pooled (n = 2):** 2.67 (2.00 to 3.57); I^2^: 0% | |  |  |
| **Need for intubation (ref: No)** | ≥ 22 wks | Hospitalized (n = 1) | Shang, 2021 (High) | aOR NR; adjusted p = 0.914  (Univariate OR: 1.42 [0.32 to 6.33]) | Very uncertain | Very low^a,c,D^ |
| **Acute Covid-19 severity (critical/ICU vs no)** | **Sleep disturbance** | | | | | |
|  | ≥ 22 wks | Hospitalized (n = 1) | Shang, 2021 (High) | aOR NR; adjusted p = 0.806  (Univariate OR: 0.87 [0.39 to 1.94]) | Very uncertain | Very low^a,c,D^ |
| **Acute Covid-19 severity (severe/critical vs no)** | ≥ 22 wks | Hospitalized (n = 1) | Shang, 2021 (High) | aOR NR; adjusted p = 0.274  (Univariate OR: 0.69 [0.37 to 1.29]) | Very uncertain | Very low^a,c,D^ |
| **Need for intubation (ref: No)** | ≥ 22 wks | Hospitalized (n = 1) | Shang, 2021 (High) | aOR NR; adjusted p = 0.076  (Univariate OR: 0.20 [0.03 to 1.53]) | Very uncertain | Very low^a,c,d^ |
| **Acute Covid-19 severity** | **Return to work** | | | | | |
|  | ≥ 22 wks | Hospitalized (n = 1) | Frontera, 2021 (High) | 1.38 (1.03 to 1.84) | Very uncertain | Very low^a,c,d^ |
| **Need for hospitalization (ref: No)** | ≥ 22 wks | Mixed (n = 1) | Westerlind, 2021 (High) | 3.75 (3.32 to 4.23) | Large increase | Low^a,c^ |

CI: confidence interval; Ref: reference; vs: versus; wks: weeks; NR: not reported; O2: oxygen; NIV: non- invasive respiratory modalities; IMV: invasive mechanical ventilation; HFNC: high-flow nasal cannula; ECMO: extracorporeal membrane oxygenation

Certainty (GRADE) was rated down by 0, 1, or 2 levels for risk of bias (a), indirectness in outcome (b), lack of consistency (c), and imprecision/ wide confidence intervals (d); capital letters indicates very serious concern.

**Table S9. Study Characteristics for Key Question 2**

| **First Author & Year; Country; Study Design & Timing; Funding Source** | **Total Sample Size at Baseline; Type of Acute Care; Covid-19 Ascertainment; Population Age, Sex, major/common co-morbidities; Covid-19 Illness Severity; Exclusions** | **Intervention/Exposure Group (IG): Description & Timing**  **Control Group (CG)** | **Assessment Timing, Relevant Outcome, Descriptions, and Analysis** | **Results for Outcomes of Interest &**  **Number Analyzed** |
| --- | --- | --- | --- | --- |
| Amini 2021  Iran  Prospective uncontrolled cohort (before-after); 3 month follow-up  Funding: Non-industry funding | 48 (from 86 volunteers)  NR (all discharged from hospital before recruitment)  Ascertainment NR  70±5.4 (range 60-80) yr; 100% male; comorbidities NR  Only patients in stage 1 of COVID disease included (non-severe symptoms)  Exclusions: severe COVID symptoms >stage1, acute psychiatric condition with psychosis, unstable medical condition that would preclude safe participation, a progressive neurological condition (such as Parkinson’s disease, multiple sclerosis, Meniere’s disease), cognitive impairment defined as a Pfeiffer Short Portable Mental Status Questionnaire (SPMSQ) score < 824, or visual or auditory impairment that could not be corrected with assistive devices. | IG: **Cognitive motor therapy (CMT)**; 4-week program done twice per week (n=42); enrolled from the community after recent discharge from hospital (unclear timing from COVID diagnosis to enrollment)  CG: None. | Baseline and 3 month post-intervention  **Quality of life:** General Health Questionnaire and subscales (GHQ-2, higher scores better, <6 on subscale or <22 on overall score indicate pathological symptoms)  **Cognitive impairment:** Mini-Mental State Examination (MMSE, higher scores better, <23 indicates possibility of a disorder) (n=42)  **Depressive symptoms** (from GHQ-2 questionnaire)  **Anxiety symptoms** (from GHQ-2 questionnaire)  **Impairment in functional capacity (**physical symptoms and social performance from GHQ-2 questionnaire)  **Analysis:** crude between time points | IG: 42 (baseline + 3 mo follow-up)  **Quality of Life:**  GHQ-2: IG (baseline) 47.750±1.550 vs IG (3 mo) 46.889±1.246; MD 0.860 (p=0.006)  **Cognitive impairment**  MMSE*: IG (baseline) 17.993±3.311 vs IG (3 mo) 19.714±2.218; MD -1.721 (p=0.001)  **Depressive symptoms:** IG (baseline) 8.589±1.660 vs IG (3 mo) 8.248±1.165  **Anxiety symptoms:** IG (baseline) 13.777±2.448 vs IG (3 mo) 12.622±2.292; MD 1.155 (p=0.001)  **Functional capacity**  Physical symptoms: **I**G (baseline) 12.656±1.359 vs IG (3 mo) 11.628±2.305; MD 1.028 (p=0.001)  Social performance: IG (baseline) 15.050±2.464 vs IG (3 mo) 14.680±2.414; MD 0.370 (p=0.001)  *Several subscales of the MMSE were found significant at 3 month follow-up (attention and calculation, recall & action performance, MD range -0.796 to -0.330), all other domains were found to be non-significant (orientation, information encoding, & lingual skills, MD range -0.149 to -0.033). |
| Anastasio 2021  Italy  Bidirectional controlled cohort; 4 month follow-up after COVID-19 diagnosis  Funding: no external funding | 222 (subgroup of those with pneumonia of 379 in study using random selection from n=1464 eligible)  Hospital standard treatment  Ascertainment PCR  Median 56 (IQR 49-63) yr; 45.9% male; hypertension 29.6%; cardiovascular disease 11.6%; diabetes 6.3%  59% pneumonia; 15% ICU  Exclusions: previous diagnosis of pulmonary disease, excluding asthma | IG: **steroids** **during hospitalization** (n=42)  CG: **no steroids** (n180) | Median 135 (IQR 102-175) d from COVID-19 symptom onset  **Dyspnea:**  Medical Research Council breathlessness scale (MMRc) during the last two weeks of the study  **Analysis:** statistical significance corrected for clinical variables (SpO2/FiO2 ratio, PSI, ARDS, IMV) | IG n=42 vs CG n=180  *Analyzed subgroup of pneumonia patients only (n=222)  **Dyspnea:**  Steroid therapy, also corrected for SpO2/FiO2 ratio, PSI, ARDS development or IMV need, was positively correlated with MMRc (p=0.05) |
| Benzakour 2021  Switzerland  Prospective uncontrolled cohort; 3 month follow-up after hospitalization from Mar 30-Jul 1^st^ 2020  Funding: NR | 109 (61 in follow-up; 364 eligible)  Hospital standard treatment  Ascertainment NR  <40 yr: 17%, 40-64 yr, 50%; ≥65 yr: 34%; 61% male; comorbidities NR  16.5% in ICU (of n=64 with CoviCare follow-up data)  Exclusions: non-French speaking individuals, and the inability to fill the questionnaires due to cognitive or physical impairment | IG: **Screening and treatment for psychiatric symptoms** (psychoeducation about PTSD, stress management, and could benefit from early and short psychotherapeutic interventions of trauma-focused CBT, EMDR or cognitive-therapy sometimes combined with pharmacological treatment) during hospitalization but also after they left hospital if needed | 3 months post-hospitalization  **Psychopathology:**  **PTSD:** Posttraumatic stress disorder checklist for DSM-5 (PCL5, lower scores better; cutoff 31 for PTSD)  **Depressive symptoms:** HADS-Depression questionnaire, lower scores better, cutoff 11 for depression  **Anxiety symptoms:** HADS-Anxiety questionnaire, lower scores better, cutoff 11 for anxiety  **Analysis:** multivariate regression analysis | T0 vs T0**PSTD:**  Positive PCL5 (N, %) T0: 15 (14.6%) vs T1: 7 (10.6%)  **Depressive symptoms:** T0: 20 (18.5%) vs T1; 6 (10.0%) (p>0.05)  **Anxiety symptoms:** T0: 17 (15.7%) vs T1 6 (10.0%) |
| Blomberg 2021  Norway  Bidirectional controlled cohort; 6 month follow-up after diagnosis between Feb 28-May 6 2020  Funding: Industry and non-industry grants | 357 (293 at follow-up for outcome of interest; 381 eligible)  Hospital standard treatment (or home-isolation  Ascertainment RT-PCR (hospitalized + home-isolated); COVID-19 specific antibodies (household seropositive contacts at 2 months follow-up)  Median 46 (IQR 30-58) yr; 49% male; 12% chronic lung disease; 11% hypertension; 7% chronic heart disease  Hospitalized patients: 48% with medical needs; 37% supplemental oxygen; 6% NIV; 9% respirator  Exclusions: address outside of Bergen, Norway | IG: **Antibiotic use** during acute phase (n=31)  CG: **No antibiotic use** (n=262) | 6 months post-diagnosis  **Fatigue:** Chalder fatigue scale (score ≥4 is fatigue)  **Analysis:** multivariate logistic regression | IG=31 vs CG=262  **Fatigue:** IG 17 (55%) vs CG 91 (35%)  **Fatigue, associated with antibiotic use (univariate):** OR 2.28 (1.08-4.84), p<0.05  **Fatigue, associated with antibiotic use (multivariate):** aOR 0.42 (0.10-1.75), p>0.05 |
| Feng 2021  China  Nonrandomized controlled experimental study  Funding: Non-industry research grants | 41 in original study (28 in follow-up) (# eligible NR)  Hospital standard treatment (supplemental oxygen, antivirals [abidor/oseltamivir], antibiotics [moxifloxacin], short-term glucocorticoids)  Ascertainment RT-PCR  51 (IQR 43-64) yr; 46% male; diabetes or hypertension 39%; smokers 29%  Severe illness (0% IV at study baseline)  Exclusions: Exclusion criteria included the following: any kind of cancer, severe liver disease, known allergy or hypersensitivity to hUC-MSCs, and other conditions that the clinician deemed inappropriate for participation. | IG: **Human umbilical cord mesenchymal stem cells (hUC-MSC; clinical-grade)**; 1 intravenous delivery; median 11.5 d (IQR 6-20) from symptom onset; (n=12)  CG: Standard care only; median 14 d (IQR 10-18) from symptom onset (n=29) | 3 months (90-95 days) post-discharge (median LOS 22 d [IQR 19.5-24])  **Quality of life**: St. George’s Respiratory Questionnaire (SGRQ) impact of lung diseases on HRQoL [lower scores better]; range 0-100)  **Fatigue** (from SGRQ questionnaire)  **Wheezing** (from SGRQ questionnaire)  **Lung function**: pulmonary function tests after bronchodialator (vital capacity (VC), forced vital capacity (FVC), forced expiratory volume in 1 s (FEV1), FEV1/ FVC ratio, peak expiratory flow (PEF), maximal mildexpiratory flow (MMF),maximal voluntary ventilation (MVV), and minute ventilation (MV), PaO2, PaCO2, oxygenation index.  **Analysis:** crude between group | IG n=8 vs CG n=20  **Quality of Life**:  IG 15.3 ± 3.7 vs. CG 31.9 ± 8.8; p=0.035  **Fatigue**:  IG 4 (50%) vs. CG 14 (70%); p=0.31  **Wheezing**:  IG 3 (37.5%) vs. 15 (75%); p=0.043  **Lung function**:  FEV1/ FVC ratio <70% IG 1 (12.5%) vs. CG 13 (65%); p=0.033  VC, FVC, PEF, MMF, MVV, MV, PaO2, PaCO2, oxygenation index all favorable for IG but not significant and p>0.15  **AEs**: No patients experienced any adverse reactions such as skin itchiness, dizziness, loss of appetite, or foggy vision after their discharge. All patients who received hUC-MSC intervention maintained normal ranges in terms of blood routine index, renal function, coagulation, electrocardiography patterns, and levels of CRP, PCT, and myocardial injury markers.  **Serious AEs**: 0 |
| Frontera 2021  US  Bidirectional controlled cohort; enrollment of hospitalized patients from Mar 10, 2020 to May 20, 2020.  Funding: NR | 790 (382 at follow-up)  Hospitalized standard treatment (corticosteroids 26.4%, hydroxycholorquine 71.2%, azithromycin 64.4%, therapeutic anticoagulation 35.1%, remdesivir 0.3%, tocilizumab 0.3%, zinc NR).  Ascertainment RT-PCR  Neurologic COVID-19, median age (IQR): 68 (55-77).  COVID-19 control, median age (IQR): 69 (57-78).  65% male, hypertension 43.2%, diabetes 28.8%  Moderate-to-critical (32.2% ICU, 51.3% neurological COVID-19 disorder)  Exclusions: negative or missing SARS-CoV-2 RT-PCR test, or evaluation in an outpatient or emergency department setting only; readmissions were excluded to avoid double  counting. | IG:  Corticosteroids (n=101)  Hydroxychloroquine (n=272)  Azithromycin (n=246)  Therapeutic anticoagulation (n=134)  Zinc (n=NR)  CG:  No corticosteroids (n=281)  No hydroxychloroquine (n=110)  No azithromycin (n=136)  No therapeutic anticoagulation (n=248)  No zinc (n=NR)  During hospitalization | 6 (±1) months post-COVID-19 symptom onset  **Impairment in functional capacity** (Modified Rankin Scale, mRS 3-6 [0=no symptoms, 6=dead]; Barthel Index for activities of daily living <100 [0=completely dependent, 100=independent for all activities]).  **Cognitive impairment** (Telephone Montreal Cognitive Assessment <18 [22=perfect score, ≤18=abnormal cognition]).  **Anxiety** (T-score>50, from Quality of Life in Neurological Disorders [Neuro-QoL], higher scores indicate worse self-reported health)  **Depression** (T-score>50, from Neuro-QoL, higher scores indicate worse self-reported health)  **Fatigue** (T-score>50, from Neuro-QoL, higher scores indicate worse self-reported health)  **Sleep disturbances** (Sleep T-score>50, from Neuro-QoL, higher scores indicate worse self-reported health)  **Return to work** (self-reported)  **Analysis:** univariate logistic regression | **3**46/382 (91%) patients had at least one abnormal outcome at 6-months  All point estimates without 95% CIs or p-values were p>0.100. All with 95% CIs were non-significant at multivariate level (p>0.05)  **Impairment in functional capacity:**  mRS 3-6:  Corticosteroids: OR 0.126  Hydroxychloroquine: OR 0.980  Azithromycin: OR 0.398  Therapeutic anticoagulation: OR 0.178  Zinc: OR 0.671 (0.44-1.03), p=0.066  Barthel Index <100:  Corticosteroids: OR 0.480 (0.29-0.80), p=0.005  Hydroxychloroquine: OR 0.970  Azithromycin: OR 0.078  Therapeutic anticoagulation: OR 0.078  Zinc: OR 0.407  **Cognitive impairment:**  Corticosteroids: OR 0.982  Hydroxychloroquine: OR 0.128  Azithromycin: OR 0.188  Therapeutic anticoagulation: OR 0.995  Zinc: OR 0.523  **Anxiety (T-score>50):**  Corticosteroids: OR 0.822  Hydroxychloroquine: OR 0.945  Azithromycin: OR 0.606  Therapeutic anticoagulation: OR 0.914  Zinc: OR 0.906  **Depression (T-score>50):**  Corticosteroids: OR 0.367  Hydroxychloroquine: OR 0.500  Azithromycin: OR 0.104  Therapeutic anticoagulation: OR 0.982  Zinc: OR 0.621  **Fatigue (T-score>50):**  Corticosteroids: OR 2.2 (1.3-3.9), p=0.004  Hydroxychloroquine: OR 0.221  Azithromycin: OR 0.633  Therapeutic anticoagulation: OR 1.8 (1.1-3.1), p=0.022  Zinc: OR 0.272  **Sleep disturbances (T-score>50):**  Corticosteroids: OR 1.7 (1.0-2.8), p=0.064  Hydroxychloroquine: OR 0.517  Azithromycin: OR 0.819  Therapeutic anticoagulation: OR 1.7 (1.0-2.8), p=0.051  Zinc: OR 0.173  **Return to work:**  Corticosteroids: OR 0.4 (0.2-0.8), p=0.008  Hydroxychloroquine: OR 0.192  Azithromycin: OR 0.458  Therapeutic anticoagulation: OR 0.31 (0.16-0.60), p=0.001  Zinc: OR 2.3 (1.2-4.4), p=0.016 |
| Gherlone 2021  Italy  Bidirectional controlled cohort; July 23, 2020 and Sept 7, 2020  Funding: Research grant from hospital | 122 consecutive patients  Hospitalized (94%) standard treatment NR but 30% steroids, 84% antibiotics, 31% biologics  Ascertainment RT-PCR with clinical/radiologic signs  62.5 (IQR 53.9-74.1) yr; 75% male; hypertension 41%, smokers 39%, diabetes 14%  Moderate-to-critical (25% ICU)  Exclusions NR | IG: **Antibiotics during acute phase** (n=102 [84%])  CG: No antibiotics (n=20 [16%]) | Median 104 d (IQR 95 to 133) post-discharge (LOS NR)  Oral manifestations arising during or after Covid-19 **as potential AE from antibiotics** (e.g. salivary gland ectasia [glands swollen, with a patent duct, and no pus leaking], dry mouth, temporomandibular joint abnormalities, facial pain, and masticatory muscle weakness)  Analysis: multivariate logistic regression for salivary gland ectasia and dry mouth (all variables p<0.05 in univariate i.e. CRP and LDH levels) - *age, sex, Covid-19 severity markers ICU/IV and comorbidities not included* | Oral cavity or facial abnormalities in 101 (83.6%; majority altered taste and smell) patients (vs 0 with pre-Covid-19 disorders)  Salivary gland ectasia present in 46 (38%) patients (more common in severe COVID-19 and were significantly older [64 yrs IQR 59-73] e.g. higher levels of serum CRP and LDH, and lower absolute lymphocyte counts on admission); 93% received antibiotics  **AE from antibiotics**: Association between antibiotic use and salivary gland ectasia (n=122): aOR 8.34 (95% CI, 1.47-158.19); p=0.049 |
| Huang 2021  China  Bidirectional controlled cohort; Jan 7 to Sept 3, 2020  Funding: Non-industry research grants | 1733 (patient-reported outcomes; of 2469 eligible) and 334 (dyspnea via lung function; of 510) outcome with 334/510 eligible at follow-up, 2469 eligible overall)  Hospital standard treatment (23% corticosteroids, 54% antivirals, 77% antibiotics, 17% thymosin, 20% intravenous immunoglobulins)  Ascertainment laboratory-confirmed  Median 57 (IQR 47-65) yr; 52% male; hypertension 29%; diabetes 12%; CVD 7%  68% oxygen therapy; 7% IMV, non-IMV or HFNC; 4% in ICU  Exclusions: died before the follow-up visit, those for whom follow-up would be difficult owing to psychotic disorder, dementia, or re-admission to hospital attributed to underlying diseases, those who were unable to move freely due to concomitant osteoarthropathy or immobile before or after discharge due to diseases such as stroke or pulmonary embolism, those who declined to participate, those unable to be contacted, and those living outside of Wuhan or in nursing or welfare homes. | IG:  Corticosteroids (n=398)  Intravenous immunoglobulins (n=345)  CG:  No corticosteroids (n=1,335)  No intravenous immunoglobulins (n=1,388)  During hospitalization | Median 153 (146-160) d from hospital discharge (186 [175-199] d since symptom onset)  **Dyspnea:** Diffusion impairment (DLCO <80% predicted)  **Anxiety or depression:** (from EQ-5D-5L questionnaire)  **Fatigue or muscle weakness**  **Analysis:** multivariate logistic regression | Corticosteroids: IG=398 vs CG=1,335 (lung function n=NR)  Intravenous immunoglobulins: IG=345 vs CG=1,388 (lung function n=NR)  **Dyspnea**  **Lung function associated with corticosteroids:** aOR 1.18 (0.60-2.34), p=0.63  **Lung function associated with intravenous immunoglobulins:** aOR 0.94 (0.49-1.79), p=0.85  **Anxiety or depression (associated with corticosteroids):** aOR 1.23 (0.88-1.72), p=0.22  **Anxiety or depression (associated with intravenous immunoglobulins):** aOR 0.77 (0.54-1.10), p=0.15  **Fatigue or muscle weakness (associated with corticosteroids):** aOR 1.04 (0.77-1.42), p=0.78  **Fatigue or muscle weakness (associated with intravenous immunoglobulins):** aOR 0.96 (0.70-1.31), p=0.78 |
| Jain 2021  US  Prospective uncontrolled cohort (control group non-Covid not eligible); 90 days after admissions April 9 to September 1, 2020  Funding: NR | 18 (of 64 eligible; 56% nonresponse rate)  Hospital (standard treatment NR)  Ascertainment NR  66 (IQR 58-72) yr; 56% male; BMI <35% obese, 0% stroke/brain injury/cardiac/neurologic diagnosis (compared with 5-6% each for those non-responding)  Severe and requiring inpatient rehabilitation (% ICU NR; 94% required oxygen & 94% ICD-10 code of deficits of cognitive function and self-awareness)  Exclusions: None | IG: Regional **in-patient rehabilitation** following acute hospitalization (details NR)  CG: None | 90 days following discharge from acute inpatient rehabilitation (LOS acute hospitalization 18 d [IQR 11-31] and rehab 10 d [IQR 7-16])  Assessment via brief standardized questionnaire administered following patient discharge via phone  **Hospital re-admissions** (within last 3 weeks of follow-up)  **Post-discharge care sought by patients** (within last 2-3 weeks of follow-up)  **Analysis:** descriptive | **Hospital re-admissions** (within last 2-3 weeks of follow-up): 0 (n=18)  Urgent care/ED visits due to rehab diagnosis (within last 2-3 weeks of follow-up): Data not available for this time period (per author response)  Outpatient visits with cardiology, pulmonary, neurologists, OT/PT/speech (within last 2-3 weeks of follow-up): Data not available for this time period (per author response) |
| Kataria 2021  India  Nonrandomized study (nonconcurrent groups); May-Nov 2020  Funding: none, in-kind by research institutes | 60  Hospital standard treatment (HCQ, antibiotic, antipyretic; 25% no medications)  RT-PCR  IG 38.7±15.4 yr vs CG 29.7±7.3 yr; 52% male; co-morbidities NR  Mild-to-moderate Covid-19 (32% asymptomatic [twice as many in IG due changes to admission criteria]; 65% mild)  Exclusion: severe illness, immunosuppressant disease, known allergies | IG: **Ayurvedic formulation** containing Tinospora cordifolia (Guduchi) and Piper longum (Pippali) twice daily administered by nurses (n=30); acute phase in hospital  CG: Standard care only (n=30) (subsequent to IG) | 3 mo post-discharge (LOS 6 d)  Structured telephone follow-up with questionnaires (“validated” but not cited)  **Quality of Life**: general health (Much better now since discharge)  **Dyspnea:** breathlessness during physical activities since discharge  **Functional limitations**: (23  **Pain**: frequent headaches, chest pain since discharge  **Fatigue:** wording NR  **Sleep disturbances**: problems in sleep since discharge  **Oxygen support at home**  **Analysis:** crude between groups | IG n=28 vs. CG n=29  **Quality of Life:** IG 18 (64.3%) vs 15 (51.7%)  **Dyspnea:** IG 6 (21.4%) vs CG 3 (10.3%)  **Functional limitations**: IG 0 vs CG 0  **Pain (headaches)**: IG 3 (10.7%) vs CG 3 (10.3%)  **Pain (chest pain)**: IG 1 (5%) vs CG 4 (17.4%); p=0.211  **Fatigue**: 1 (5%) vs CG 6 (26.1%); p=0.065  **Sleep disturbances**: IG 3 (10.7%) vs. CG 1 (3.4%)  **Oxygen support at home:** IG 0 vs CG 0 |
| Li 2021  China  Prospective controlled cohort; discharged Jan 21-May 21, 2020  Funding: | 96 (of 105 eligible)  Hospital standard care  “Definitive diagnosis”  49±15 yr; 36% male; co-morbidities NR  All severities (22% severe or critical)  Exclusions: pregnancy, organ failure, complete absorption/recovery of pneumonia on chest CT at discharge | IG: **Chinese medicine** 2 different oral formula twice daily, depending on type of syndrome (pathogen residue or qi and yin deficiency) (n=64); for 28 d after discharge  CG: Western medicine or no medicines (n=32) | 84 d post-discharge (56 d after end of treatment)  Questions on symptoms at follow-up visits  Fatigue  Dyspnea: shortness of breath  Insomnia  Chest tightness | All patients: enrollment nearly 40% of patients had symptoms, such as cough, sputum, dry throat, thirst, insomnia, fatigue, shortness of breath, chest tightness etc. vs. at 84 d insomnia (8.33%), fatigue (9.38%), respiratory symptoms <5%  No significant difference in the improvement rates of symptoms, including fatigue, between the two groups (P>0.05)  Author contact for data by arm but NR as of Aug 26, 2021 |
| Martin 2021  Belgium  Prospective controlled cohort; April-July 2020  No funding | 27 (of 48 eligible)  Hospital standard care  No control & unadjusted RT-PCR  IG 60.8±10.4 yr vs. CG 61.9±10.7 yr; 63% males; co-morbidities NR  Severe or critical illness (NIH definition; 22% ICU)  Exclusions: chronic respiratory disease; pulsed oxygen saturation (SpO2) decreased more than 4% during STST at discharge | IG: **Telerehabilitation program** with real-time clinician-patient interactions, involving a videoconferencing platform (Teams, Microsoft, Redmond, WA, USA), laptop, phone or tablet and a web camera. Supervised by a physiotherapist; home–based individual and group 50-min endurance and strength exercises 2/wk for 6 wk with encouragement to do 2-3/wk unsupervised (timing after discharge NR) (n=14)  CG: Patients refusing program (n=13) | 3 mos post-discharge (LOS 16.1±9.1 d)  **Dyspnea**:  1-min sit-to-stand test (STST) (number of repetitions under 50^th^ and 2.5 %iles with SpO2 and self-reported on visual analogue scale 0-10) after STST  **Any adverse event** | At baseline, the cardio-respiratory parameters were similar between the two groups and in a normal range, either before or after STST.  **Dyspnea**  **STST:** under 50^th^ %ile IG pre 14/14 (100%) vs post 13/14 (93%) vs CG pre 14/14 (100%) vs post 14/14 (100%); under 2.5 %ile pre 77% vs. post 37% (not by group)  **SpO2 after STST: NR**  **Change since baseline in dyspnea after STST** IG 2.5 (-2 to 7) vs CG 2.0 (0-6); p=0.56  **Any adverse event: none reported** |
| Meije 2021  Spain  Prospective uncontrolled cohort; Mar – Sept 2020  Funding: None | 302 (of 461 discharged from hospital)  Hospital standard care (HCQ if mild illness or <65 and no risk factors; HCQ & lopinavir/ritonavir & cextriazone & azithromycin if moderate/severe or >65 with risk factors; others as needed)  RT-PCR or probable/clinical diagnosis (68 [23%] PCR negative but similar baseline and clinical characteristics)  68.8±12.7 yr; 57% males; cardiovascular disease 10.6%, chronic pulmonary disease 13.6%, obesity 13.6%, diabetes mellitus 12.3%, hypertension 45.4%  Variable severity (oxygen supplementation 74.8%; 1% IV; ICU 8.9%)  Exclusions: None | IG: **Outpatient assessment** (standard follow-up protocol checklist of symptoms and adverse events, medical history, physical examination, laboratory testing including chest x-ray) **with telephone follow-up** (same standard checklist); **medical follow-up as required** (50.3% patients; 27.1% to pulmonologist)  One week after enrollment at median 45 (range 43-47) d after hospital discharge  CG: None | Median 7 (range 6-7.4) mos after hospital discharge (LOS 8 [IQR 5-12] d)  **Proportion with persistent symptoms**: 1+ of dyspnoea, cough, chest pain, diarrhoea, migraine, anosmia, dysgeusia, asthenia, myalgias, neurological disorder  **Dyspnea**  **Pain**: migraine and chest pain  **Insomnia**  **Psychopathology:** need for psychological medication; fear of relapse  **Functional capacity:** limitations in their usual life  **Analysis**: not analysis just proportions | N=294  **At post discharge assessment vs. 7 mo follow-up:**  **Proportion with persistent symptoms:** 228 (77.6%) vs 147 (50%)  **Dyspnea**: 88 (29.9%) vs. 28 (9.5%)  **Pain**:  migraine 19 (6.5%) vs. 12 (4.1%)  chest pain 30 (10.2%) vs. 8 (2.7%)  **Insomnia**: 62 (21.1%) vs. 54 (18.4%)  **Psychopathology**:  need for psychological medication 35 (11.9%) vs. 30 (10.2%)  fear of relapse 68 (23.1%) vs. 86 (29.3%)  Functional status: 60 (20.4%) vs. 57 (19.4%) |
| Qin 2021  China  Bidirectional controlled cohort; 3 month follow-up after hospital discharge between Jan-Feb 2020  Funding: Non-industry research grants | 81 at follow-up for lung function tests (647 followed up overall of 749 eligible)  Hospital standard treatment (as per Chinese COVID-19 guideline)  Ascertainment RT-PCR  59 (SD 14) yr; 42% male; hypertension 30%; diabetes 11%; chronic respiratory disease 6%  30% acute respiratory distress syndrome  Exclusions: NR | IG: Corticosteroids (n=17)  CG: No corticosteroids (n=64)  During hospitalization | 3 months (90 days) post-discharge (mean inpatient days 18±8)  **Lung function:** impaired diffusion capacity of the lung for carbon monoxide (DLCO) <80% predicited performed by a professional doctor with 20 years’ experience using the MasterScreen PFT system (Jaegar, Hoechberg, Germany)  **Analysis:** univariate regression analysis | IG n=17 vs CG n=64  **Dyspnea:**  **DLCO <80% predicted:** IG: 10 (59%) vs. CG: 34 (53%)  OR: 1.3 (0.4-3.7), p=0.675 |
| Vetrici 2021  US  Pilot RCT; March-July 2020  Funding: None | N=10  Hospital standard care (oxygen supplementation, fluid and electrolyte balance, standard nursing care), and no additional corticosteroid, antiviral, pharmacological or antibody treatment was provided).  RT-PCR  IG 53.2 ± 16.7 yr vs. CG 53.4 ± 18 yr; 80% males; diabetes 50%; hypertension 50%; asthma 20%  Exclusion: ventilator management, autoimmune disorders or inflammatory conditions not related to COVID-19, and pregnancy | IG: Photobiomodulation  (PBMT) adjunctive anti-inflammatory treatment; daily sessions (each 14 minutes per lung) x 4 d of near-infrared light treatment targeting the lung tissue via a Multiwave Locked System (MLS) laser; during hospitalization.  CG: Usual medical care only (observation for 4 days for attention control) | 5 mo after treatment  **Symptoms (respiratory)**  **AEs from treatment** | **Symptoms (respiratory)**: IG 0 vs. CG: 2 of the 3 living CG patients, one who recovered spontaneously and one who was on a ventilator, continue to experience aggravating pulmonary symptoms  **AEs from treatment: IG 0** |
| Wu 2021  China  Bidirectional controlled cohort; Feb 2020-Feb 2021  Funding: Grants | 83 (of 135 eligible)  Hospitalized standard care (100% received antivirals; 0% corticosteroids)  RT-PCR  60 (IQR 52–66) yr; 57% male; see exclusions for lack of comorbidity.  Severe illness (0% IV)  Exclusions: history of hypertension; diabetes; cardiovascular disease; cancer; and chronic lung disease, including asthma or chronic obstructive pulmonary disease; or a history of smoking at admission; required intubation and mechanical ventilation | IG:  Oseltamivir  Ganciclovir  CG:  No Oseltamivir  No Ganciclovir  During hospitalization | 12 mos (348 [IQR 341–359] d) post-discharge (LOS 29 [IQR 25–35] d)  **Dyspnea/lung function**: deficit in DLCO (diffusing capacity of the lungs for carbon monoxide) <80% predicted  **Analysis**: univariate regression analysis | n=83  27 (33%) patients had impaired DLCO at 12 months  **Dyspnea/lung function:**  **DLCO <80% association with Oseltamivir:** OR 0.75 (95% CI 0.29-1.92)  **DLCO <80% association with Ganciclovir:** OR 1.34 (95% CI 0.53-3.38) |
| Xiong 2021  China  Bidirectional controlled cohort; median 97 d follow-up from hospital discharge before Mar 1 2020  Funding: NR | 538 (of 891 in study)  Hospital standard treatment (corticosteroids 26%, antiviral treatment 91%)  Ascertainment as per WHO interim guidance  Median 52 (IQR 41-62); 55.5% male; hypertension 15.2%; diabetes 7.4%  33.5% severe, 5% critical COVID-19  Exclusions: severe and complex underlying diseases or receiving invasive treatment, as well as women who were pregnant or breastfeeding | IG: Corticosteroids (n=138)  CG: No corticosteroids (n=400)  During hospitalization | Median 97 (IQR 95-102) d post-discharge from hospital (range 91-116d)  **Fatigue** (physical decline/fatigue)  **Dyspnea** (post-activity polypnoea)  **Analysis:** crude between groups | IG: n=138 vs. CG: n=400  **Fatique:** IG: 39 (28%) vs. CG: 113 (28%)  **Dsypnea:** IG: 34 (25%) vs. CG: 81 (20%) |
| Zhao 2020  China  Bidirectional controlled cohort; enrollment Jan 20, 2020 to Feb 24, 2020 with 3 month follow-up post-discharge  Funding: Grant | 55 (of 77 COVID-19 cases)  Hospital standard treatment (traditional Chinese medicine 38%, low-dose corticosteroids 13%)  Ascertainment RT-PCR  47.7 (SD 15.5); 58.2% male; hypertension 10.9%; diabetes 3.64%; cardiovascular disease 3.6%  7% severe, 85% moderate cases  Exclusions: critical cases were excluded from the study. | IG: Low-dose corticosteroids (n-=7), in hospital  CG: No low-dose corticosteroids (n=48) | 3 month post-discharge from hospital  **Dyspnea** (impaired DLCO <80% predicted) | IG: n=7 vs. CG: n=48  **Dyspnea:**  IG: 1 (14.3%) vs. CG: 8 (16.7%) |

**Abbreviations**: IV: invasive ventilation, d: day(s); yr: year(s); mo: month(s); IQR: interquartile range; SD: standard deviation; LOS: length of study (hospitalization unless otherwise stated); HCQ: hydroxychloroquine; NR: not reported; CRP: C-reactive protein; PCT: procalcitonin; AE: adverse events; ICU: intensive care unit; OR: odds ratio; aOR: adjusted odds ratio; LDH: lactate dehydrogenase; BMI: body mass index; ED: emergency department; OT: occupational therapist; PT: physiotherapist; NIH: National Institute of Health; STST: sit-to-stand test; SpO2: pulsed oxygen saturation

**Table S10. Risk of Bias for Key Question 2 Cohort Studies (N=15)**

| **Author, year** | **Were the two groups similar and recruited from the same population?** | **Were the exposures measured similarly to assign people to both exposed and unexposed groups?** | **Was the exposure measured in a valid and reliable way?** | **Were confounding factors identified?** | **Were strategies to deal with confounding factors stated?** | **Were the groups/ participants free of the outcome at the start of the study (or at the moment of exposure)?** | **Were the outcomes measured in a valid and reliable way?** |
| --- | --- | --- | --- | --- | --- | --- | --- |
| Amini 2021 | NA | NA | + | ? | ? | + | + |
| Anastasio 2021 (subjective outcomes) | + | + | ? | + | + | ? | + |
| Anastasio 2021 (objective outcomes) | + | + | ? | + | + | ? | + |
| Benzakour 2021 | NA | NA | + | ? | + | ? | + |
| Blomberg 2021 | + | + | - | + | + | - | ? |
| Frontera 2021 (subjective outcomes) | + | + | + | + | + | ? | + |
| Frontera 2021 (objective outcomes) | + | + | + | + | + | + | + |
| Gherlone 2021 | + | + | + | + | + | ? | + |
| Huang 2021 (subjective outcomes) | + | + | + | + | + | ? | + |
| Huang 2021 (objective outcomes) | + | + | + | + | + | ? | + |
| Jain 2021 | + | + | + | + | + | NA | ? |
| Li 2021 | + | + | + | + | - | ? | ? |
| Martin 2021 | + | + | + | - | + | NA | + |
| Meije 2021 | + | NA | + | + | + | ? | ? |
| Qin 2021 | + | ? | ? | + | + | ? | + |
| Wu 2021 | + | ? | ? | + | + | ? | + |
| Xiong 2021 | + | + | + | + | + | - | + |
| Zhao 2020 | + | + | + | + | + | - | + |

+: Yes/low risk of bias; -: No/high risk of bias; ?: Unclear risk of bias; NA: Not applicable

**Table S10 Continued**

| **Author, year** | **Was the follow up time reported and sufficient to be long enough for outcomes to occur?** | **Was follow up complete, and if not, were the reasons to loss to follow up described and explored?** | **Were strategies to address incomplete follow up utilized?** | **Was appropriate statistical analysis used (i.e. age, sex, comorbidities, illness severity)?** | **No evidence of selective reporting of results and/or analyses?** | **No evidence of missing outcome data?** | **Was a large proportion (e.g., >50%) of eligible participants enrolled in the study?** | **Overall rating** |
| --- | --- | --- | --- | --- | --- | --- | --- | --- |
| Amini 2021 | + | + | NA | ? | + | + | - | Some concerns |
| Anastasio 2021 (subjective outcomes) | + | - | - | ? | ? | + | ? | High |
| Anastasio 2021 (objective outcomes) | + | - | - | ? | ? | + | ? | High |
| Benzakour 2021 | + | - | ? | + | + | + | - | High |
| Blomberg 2021 | + | ? | NA | + | + | + | + | High |
| Frontera 2021 (subjective outcomes) | + | ? | - | - | + | + | - | High |
| Frontera 2021 (objective outcomes) | + | ? | - | - | + | + | - | High |
| Gherlone 2021 | + | ? | ? | + | + | + | - | Some concerns |
| Huang 2021 (subjective outcomes) | + | + | NA | + | + | + | + | Some concerns |
| Huang 2021 (objective outcomes) | + | - | - | + | + | + | - | High |
| Jain 2021 | Y | ? | ? | - | + | ? | - | High |
| Li 2021 | + | + | NA | - | ? | + | + | High |
| Martin 2021 | + | + | NA | - | + | + | + | High |
| Meije 2021 | + | + | NA | - | + | + | + | Some concerns |
| Qin 2021 | + | - | - | - | + | + | + | High |
| Wu 2021 | + | + | NA | - | + | + | + | Some concerns |
| Xiong 2021 | + | - | - | - | + | ? | + | High |
| Zhao 2020 | + | ? | - | - | + | + | + | High |

**Table S11. Risk of Bias for Key Question 2 Quasi-experimental Studies (N=2)**

| **Author, year** | **Is it clear in the study what is the ‘cause’ and what is the ‘effect’ (i.e. there is no confusion about which variable comes first)?** | **Were the participants included in any comparisons similar?** | **Were the participants included in any comparisons receiving similar treatment/care, other than the exposure or intervention of interest?** | **Was there a control group?** | **Were there multiple measurements of the outcome both pre and post the intervention/**  **exposure?** | **Was follow up complete and if not, were differences between groups in terms of their follow up adequately described and analyzed?** | **Were the outcomes of participants included in any comparisons measured in the same way?** | **Were outcomes measured in a reliable way?** | **Was appropriate statistical analysis used?** | **No evidence of selective reporting of results and/or analyses?** | **No evidence of missing outcome data?** | **Was a large proportion (e.g., >50%) of eligible participants enrolled in the study?** | **Overall rating** |
| --- | --- | --- | --- | --- | --- | --- | --- | --- | --- | --- | --- | --- | --- |
| Feng 2021 (subjective outcomes) | + | + | + | + | - | + | + | + | + | + | + | + | Low |
| Feng 2021 (objective outcomes, lung function) | + | + | + | + | - | + | + | + | + | + | + | + | Low |
| Feng 2021 (objective outcomes, AEs) | + | NA | NA | - | NA | + | NA | ? | NA | ? | + | + | Some concerns |
| Katarina 2021 | + | ? | ? | + | - | + | + | ? | + | + | + | ? | Some concerns |

+: Yes/low risk of bias; -: No/high risk of bias; ?: Unclear risk of bias; NA: Not applicable

**Table S12: Risk of Bias for Key Question 2 Randomized Trials (N=1)**

| **Author, year** | **Domain 1. Randomization process** | **Domain 2. Deviations from the intended interventions (effect of assignment to intervention)** | **Domain 3. Missing outcome data** | **Domain 4. Measurement of the outcome** | **Domain 5. Selection of reported results** | **Overall rating** |
| --- | --- | --- | --- | --- | --- | --- |
| Vetrici 2021 | Some concerns | High | Some concerns | High | Some reasons | High |
